# The impact of poverty on mental illness: Emerging evidence of a causal relationship

**DOI:** 10.1101/2023.05.19.23290215

**Authors:** Mattia Marchi, Anne Alkema, Charley Xia, Chris H. L. Thio, Li-Yu Chen, Winni Schalkwijk, Gian M. Galeazzi, Silvia Ferrari, Luca Pingani, Hyeokmoon Kweon, Sara Evans-Lacko, W. David Hill, Marco P. Boks

## Abstract

The link between poverty and mental illness has sparked discussions concerning the poverty role as a risk factor for poor mental health. If poverty has as a causal role in mental illness, it would have profound implications for our comprehension of mental well-being and guide efforts to address the increasing incidence of mental health disorders.

Building on the recent breakthrough discovery of heritability of poverty traits and utilizing large-scale genome-wide association studies of mental illness, we used Genomic Structural Equation Modeling (GSEM) and Mendelian randomization (MR) to examine the evidence of a causal relationship between poverty and mental illness. A common factor of poverty was derived from household income (HI), occupational income (OI), and social deprivation (SD). The causal effect of poverty was examined on 9 mental illnesses: attention deficit and hyperactivity disorder (ADHD), anorexia nervosa (AN), anxiety disorders (ANX), autism spectrum disorders (ASD), bipolar disorder (BD), major depressive disorder (MDD), obsessive-compulsive disorder (OCD), post-traumatic stress disorder (PTSD), and schizophrenia (SZ), while accounting for the influence of cognitive ability (CA).

Our analysis highlights HI as the measure of poverty with the strongest correlation with the common factor, when compared to OI and SD. Using the common factor of poverty, bidirectional MR provided evidence that mental illness leads to poverty, consistent with the existing paradigm. What is new is evidence that higher levels of poverty likely pose a causal factor in developing ADHD (Inverse Variance Weighted Odds Ratio per Standard Deviation change [IVW OR]=3.43[95%CI:2.95-3.99]), MDD (IVW OR=1.49[95%CI:1.29-1.72]), and SZ (IVW OR=1.53[95%CI:1.35-1.73]), but exerts a protective effect against AN (IVW OR=0.50[95%CI:0.40-0.62]). The direct effect of poverty on mental illness remained following adjustment for CA, albeit with reduced effect sizes.

Our research indicates that higher poverty levels are likely causal risk factors for MDD and SZ, but protective against AN. Notably, CA explains a significant portion of the impact of poverty, aligning with prior reports that highlight the contribution of impaired cognitive function to severe mental illnesses. Although individuals’ skills and abilities tied to earning capacity may be the variables with the actual causal effect of poverty on mental illness, our findings warrant further investigations into interventions targeting poverty and cognitive abilities to advance mental health.

## Introduction

The association between mental illness and social class was first demonstrated in a 1958 study by Hollingshead and Redlich, who found that individuals from lower socioeconomic backgrounds had a higher incidence of severe and persistent mental illness and received less adequate treatment ^1^. More than fifty years later, the same social conditions persist, and affect mental health worldwide. Epidemiological studies throughout the world have demonstrated an association between mental health and socio-economic status (SES) ^2–4^, with mental illness being more common among people from lower social classes ^5, 6^. Also, studies on income fluctuations found consistent changes in mental health ^7, 8^.

Although the association between poverty and mental illness is strong across these studies, there is limited evidence that supports a causal relationship. Several factors, such as reverse causation and residual confounding, make it difficult to determine whether poverty causes mental illness or if it is the other way around and mental illness leads to poverty. However, understanding the causality of the relationship between poverty and mental illness may be crucial for public health policies as they may target essential aspects of poverty and improve public mental health ^9^.

To date, uncertainty remains with respect to the direction of the association between poverty and mental illness, and two explanatory hypotheses compete: social causation and social selection ^10^. According to the social causation theory, the socio-economic adversity faced by lower socio-economic groups precipitates mental illness in vulnerable individuals possibly mediated by factors such as housing insecurity, substance use, and stress. Conversely, the social selection theory suggests that the overrepresentation of low socio-economic status among people with mental illness is mainly attributable to downward social mobility as a consequence of the impairment associated with poor mental health^11^.

Conducting a randomized controlled trials to determine causality of the role of poverty is not feasible nor ethical. An alternative method to investigate is Mendelian Randomization (MR), which uses genetic data from genome-wide association studies (GWAS) to examine whether a risk factor fits in a causal model for an outcome^12^. MR method takes advantage of the fact that genetic variants are fixed at conception and are less susceptible to the confounding effects that make results from observational studies difficult to interpret ^13^.

Such a study on poverty come with a conceptional challenge. Studies on poverty often make use of a single measure, usually based on income recorded at the household level or as individual income or operationalized employment status of an individual, by occupational income ^7, 11^. However, no single indicator can capture the multiple dimensions of poverty, such as lack of material goods, limited access to education and healthcare services, inadequate living standards, disempowerment, poor quality of work, the threat of violence, and living in areas that are environmentally hazardous, among others ^14–17^. For that reason, the UK health authorities commonly use a composite measure known as the Townsend Deprivation Index to assess material deprivation within a population ^18^.

In this study, we applied MR to examine the causal effects of poverty on nine mental illnesses. First, to maximize the power required to examine the effects of poverty and its multidimensional aspects, we derived a poverty factor using household income (HI), occupational income (OI), and social deprivation (SD. These three measures capture poverty at the level of the individual, the household, and of the area in which one lives and so facilitate a more informed understanding of which aspects of poverty are those that present the greatest risk to individual level mental health. Second, we investigated the causal relationship of the common factor of poverty and each of the three indicators of poverty with nine mental illnesses: attention deficit and hyperactivity disorder (ADHD), anorexia nervosa (AN), anxiety disorder (ANX), autism spectrum disorder (ASD), bipolar disorder (BD), major depressive disorder (MDD), obsessive-compulsive disorder (OCD), post-traumatic stress disorder (PTSD), and schizophrenia (SZ). To account for potential confounding effects, we also extended the MR analyses to include cognitive abilities (CA).

## Methods

### Study design and data sources

We conducted a two-sample MR study using summary-level GWAS data that for the most part were publicly available. Ethical approval was obtained in all original studies.

MR relies on three main assumptions on the validity of the genetic instrument variable, which are: relevance, independence, and exclusion restriction ^19^. These require that the genetic variant 1) must be related to the exposure (i.e., only SNPs that reach genome-wide significance in the association with the exposure, that is 5e-8, are used), 2) it is not correlated with confounders in the exposure–outcome relationship, and 3) affects the outcome only through the risk factor (i.e., “no horizontal pleiotropy” rule).

The schematic overview of the study is represented in Figure 1.

**Figure 1:**
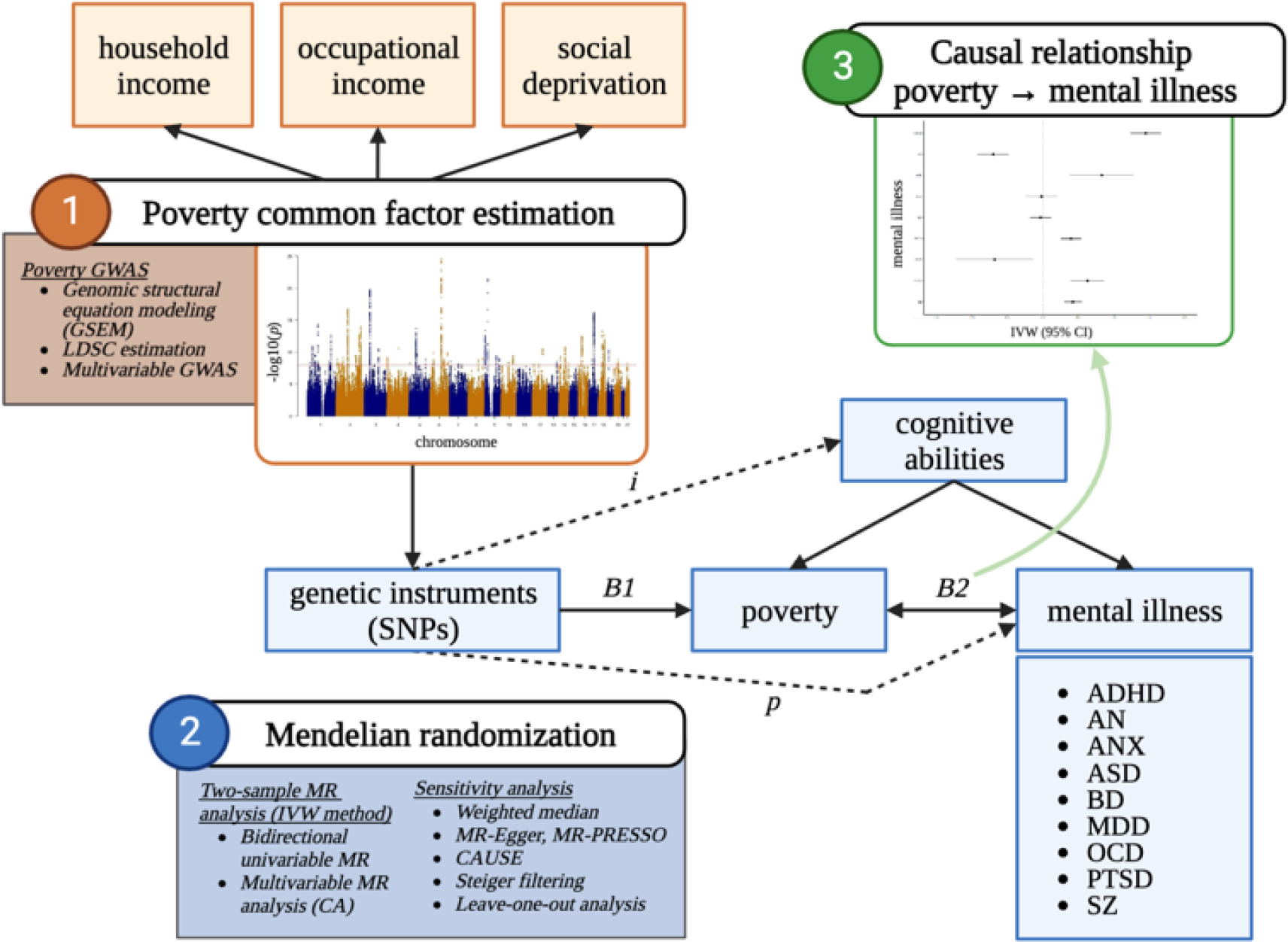
Schematic overview of the research process. Abbreviations: ADHD: attention deficit hyperactivity disorder; AN: anorexia nervosa; ANX: anxiety disorder; ASD: autism spectrum disorders; BD: bipolar disorder; CA: cognitive abilities; CAUSE: causal analysis using summary effect estimates; IVW: inverse variance weighted; LDSC: multivariable linkage disequilibrium score regression; MDD: major depressive disorder; MR: mendelian randomization; MR-PRESSO: MR pleiotropy residual sum and outlier; OCD: obsessive-compulsive disorder; PTSD: post-traumatic stress disorder; SZ: schizophrenia; SNP: single nucleotide polymorphism. Legend: B1 is the association between the genetic variants and the exposure of interest (MR relevance assumption); i: potential violation of the MR independence assumption; p: potential pleiotropic effect of the genetic variants on the outcome (i.e., violation of the MR exclusion restriction assumption); B2 is the causal association of interest. The figure was created with BioRender.com.

### Poverty instruments

The source of the summary data on HI, SD, and CA can be found in Table 1. A total of 379,598 participants of European ancestry provided genotype data and data on their level of yearly HI before tax. HI was primarily analyzed as a continuous variable. To investigate whether the relation between HI and mental illness is particularly strong at any specific level of income, we further categorized HI as a dichotomous variable, defining the cases in the following way: (1) low HI: being less than £18,000; (2) low-mid HI: being less than £29,999; (3) mid-high HI: being more than £52,000; (4) high HI: being more than £100,000. One GWAS for each HI category versus all the other categories was performed. A total of 440,350 individuals of European ancestry had genotype data and data on their level of SD, measured with Townsend Deprivation index, and it was analyzed as a continuous variable. Finally, GWAS summary data on cognitive ability of 248,482 individuals was included ^20^. The CA measure was derived from multiple cohorts that each had administered a battery of cognitive tests to their participants. Correlations between tests of CA are high with estimates between r = 0.8 and r = 1.0 being reported ^21, 22^. CA was analyzed as a continuous variable.

**Table 1:**
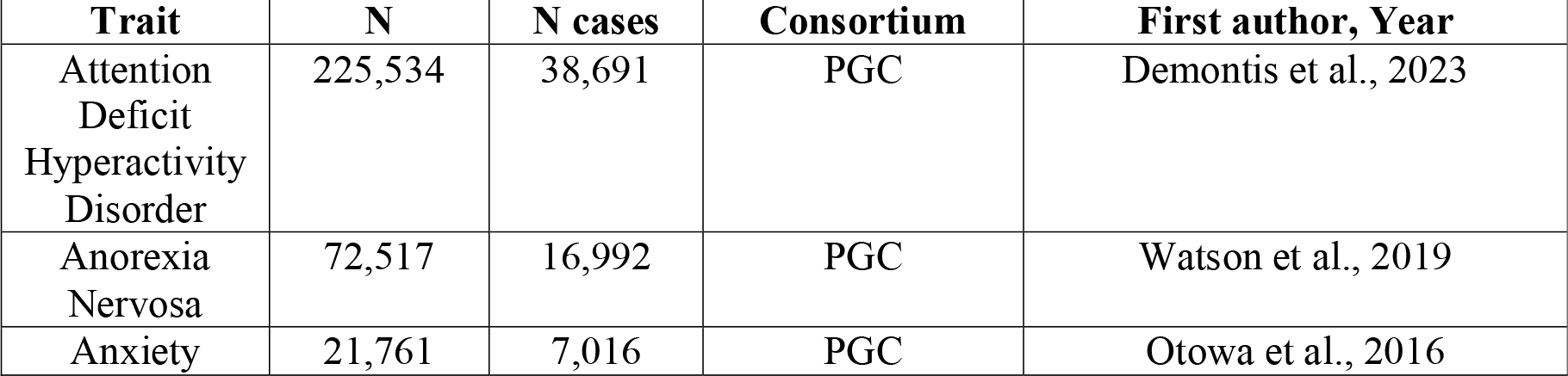
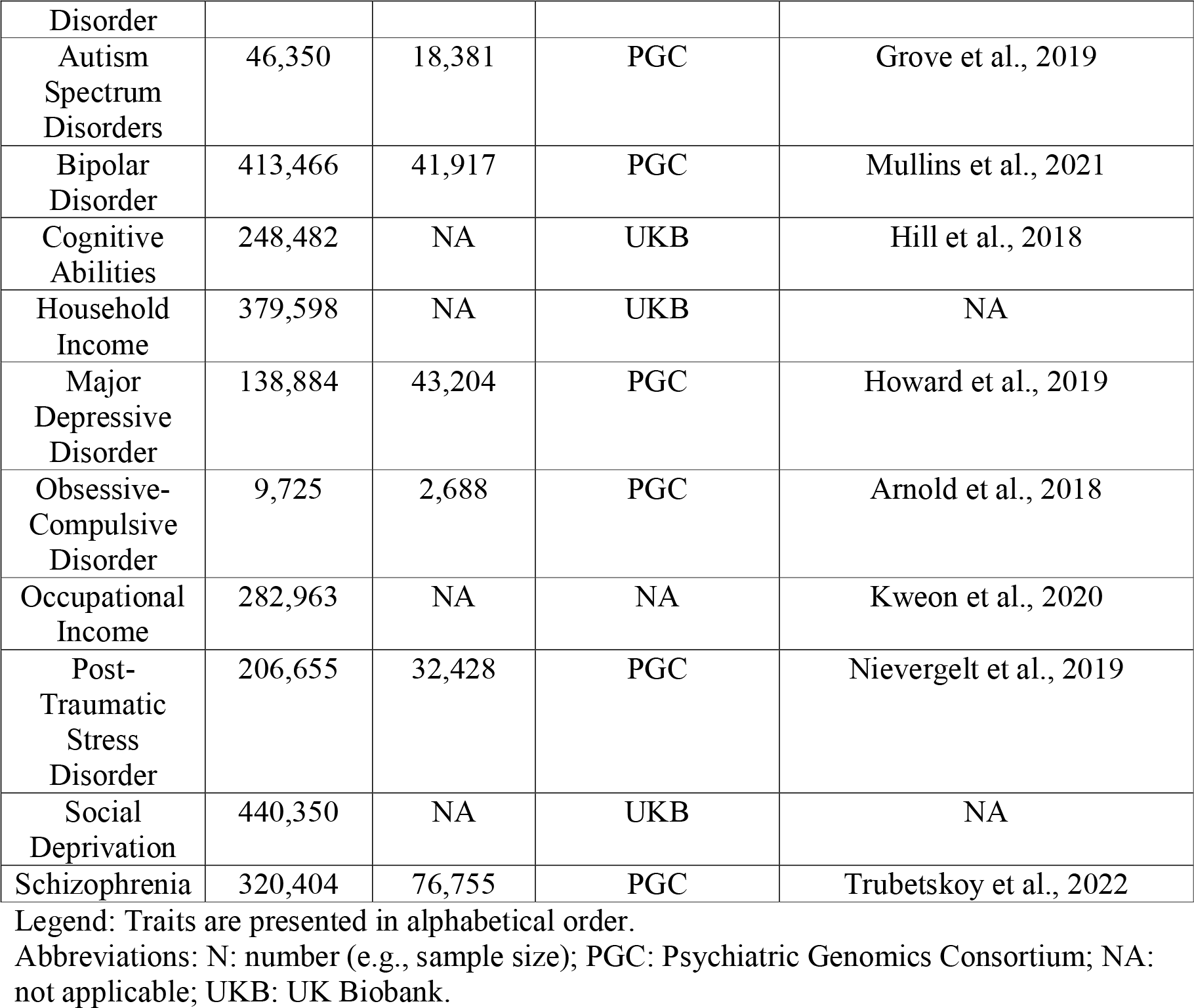
GWAS information for each phenotype

Results from a meta-analysis of two OI GWAS ^23^, including UK Biobank and the Health and Retirement Study (the latter involving participants from White American ancestry), were used to generate the instruments for OI which were analyzed as continuous variable.

Finally, we estimated a latent factor GWAS of poverty (P) by jointly modelling cross-trait liability of continuous HI, OI, and SD, using Genomic structural equation modeling (GSEM). First, we estimated the Multivariable Linkage Disequilibrium Score Regression (LDSC) ^24^ across HI, OI, and SD. Then, we combined the LDSC output with the HI, OI, and SD summary statistics to run the multivariable common factor GWAS. This common factor poverty GWAS was built using as unit of identification HI. To allow estimation of poverty - instead of income – in the MR analyses on mental illness, the regression coefficients have been reversed.

### Mental illness instruments

Results from the most updated GWAS of ADHD ^25^, AN ^26^, ANX ^27^, ASD ^28^, BD ^29^, MDD ^30^, OCD ^31^, PTSD ^32^, and SZ ^33^ were obtained. Two-sample MR method requires minimal sample overlap between exposure and outcome GWAS ^34^; therefore, mental illness summary-level data were obtained mainly from the Psychiatric Genomic Consortium (PGC) excluding UK Biobank participants. The GWAS of the mental illness traits considered were provided and analyzed as case/control. The effective sample size across the cohorts contributing to the GWAS meta-analysis was calculated for each trait and ranged from 320k (for SZ) to 10k (for OCD). Table 1 summarizes the GWAS information.

### Mendelian Randomization

The full set of GWAS summary statistics for each exposure was first restricted to the variants reaching the genome-wide significance threshold (i.e., p-value<5e-8). Then, to ensure independence between instruments, we applied a strict clumping procedure (LD r^2^< 0.001 within 10 Mb, using the 1000G EUR as the reference panel). Following that, SNP alleles were harmonized between exposure GWAS and outcome GWAS before running MR.

We conducted bidirectional univariable MR between each poverty measure and mental illness. The inverse variance weighted (IVW) method was used to estimate effects in the primary analyses ^35^ and the results were presented using forest plots. The weighted median (WM) ^36^, MR Egger regression (MR-Egger) ^37^, and MR pleiotropy residual sum and outlier (MR-PRESSO) ^38^ methods were used as sensitivity analyses, because each method makes different assumptions regarding instrument validity. Specifically, MR-Egger and MR-PRESSO are better than IVW in case of horizontal pleiotropy (i.e., violation of the exclusion restriction assumption). In addition, we used the Causal Analysis Using Summary Effect Estimates (CAUSE) ^39^ to account for the presence of correlated or uncorrelated pleiotropy (i.e., violation of the independence and exclusion restriction assumptions). Further information on MR-PRESSO and CAUSE is available in the Supplementary File 1.

We presented the results of MR analyses as Beta (B) corresponding to the log-odds for binary traits (e.g., mental illness) or to the unstandardized linear regression coefficients for continuous traits (e.g., poverty measures) with their respective 95% confidence intervals (95%CI). When the outcome of the analysis was binary (i.e., mental illness and HI levels) we also provided a conversion to Odds Ratio (OR) with corresponding 95%CI.

We also conducted leave-one-out analyses to investigate if the effects are driven by one or a subset of the SNPs and investigated whether the instruments represent the correct causal direction using Steiger analyses, including Steiger test and Steiger filtering ^40^. Steiger filtering is particularly useful to avoid false positive findings due to reverse causation (i.e., violation of the exclusion restriction assumption). The *mr_wrapper()* function of the *TwosampleMR* package performs Steiger tests on each SNP, evaluating if the R^2^ of the exposure was greater than the R^2^ of the outcome, indicating the correct direction of the association. Subsequent Steiger filtering excluded SNPs with false results, allowing MR analyses to be performed on the subset of SNPs with verified associations.

Finally, we hypothesized that CA is a likely confounder in the poverty and mental illness relationship, and our instruments for poverty are possibly related to CA (i.e., violation of the independence assumption). Therefore, we used multivariable MR (MVMR) ^19^ to estimate the direct effect of poverty on mental illness, independent of CA. For this purpose, we clumped the full list of SNPs from each poverty indicator GWAS (to ensure only independent SNPs are included) and restricted it to those SNPs found in the outcome GWAS, and then ran the analyses on each instruments-mental illness set.

Instrument strength was quantified using the mean F statistic within the univariable IVW analyses, considering a value of F <20 as indicative of weak instruments ^41^.

All the analyses were conducted in R ^42^, using the packages *TwosampleMR* ^43^, *CAUSE* ^39^, and *GenomicSEM* ^44^. The significance threshold was p<0.05. Since there is only one relationship that is tested - that between poverty and mental illness – we did not apply a multiple testing p-value correction.

## Results

### Latent poverty-factor estimation

To analyze the joint genetic architecture of poverty we ran a multivariable GWAS for which a common factor defined by genetic indicators is regressed on a SNP. This allows for estimation of a set of summary statistics for the common factor that represent the SNP effects on the common factor. The multivariable GWAS of the latent poverty factor (P) was modelled from three indicators: household income, social deprivation, and occupational income. Specifically, indicators were ordered such that the phenotype with the largest, unstandardized loading on the common factor is listed first, that led to set household income as the unit load identification (see Supplementary Table 1 showing the factor loading of each indicator). The heritability of the common factor GWAS was estimated with the h^2^ and resulted around 8.4%. Mean chi^2^, LDSC intercept, and h^2^ for the common factor GWAS and GWAS of HI, SD, and OI are presented in Supplementary Table 2.

The GWAS of the latent poverty factor identified 99 significantly associated independent loci at GWAS threshold (p-value≤5e-8), as displayed in Figure 2 (see also Supplementary File 2 for the lead SNPs list). The full GWAS summary statistics are available upon reasonable request to the lead author (MM).

**Figure 2:**
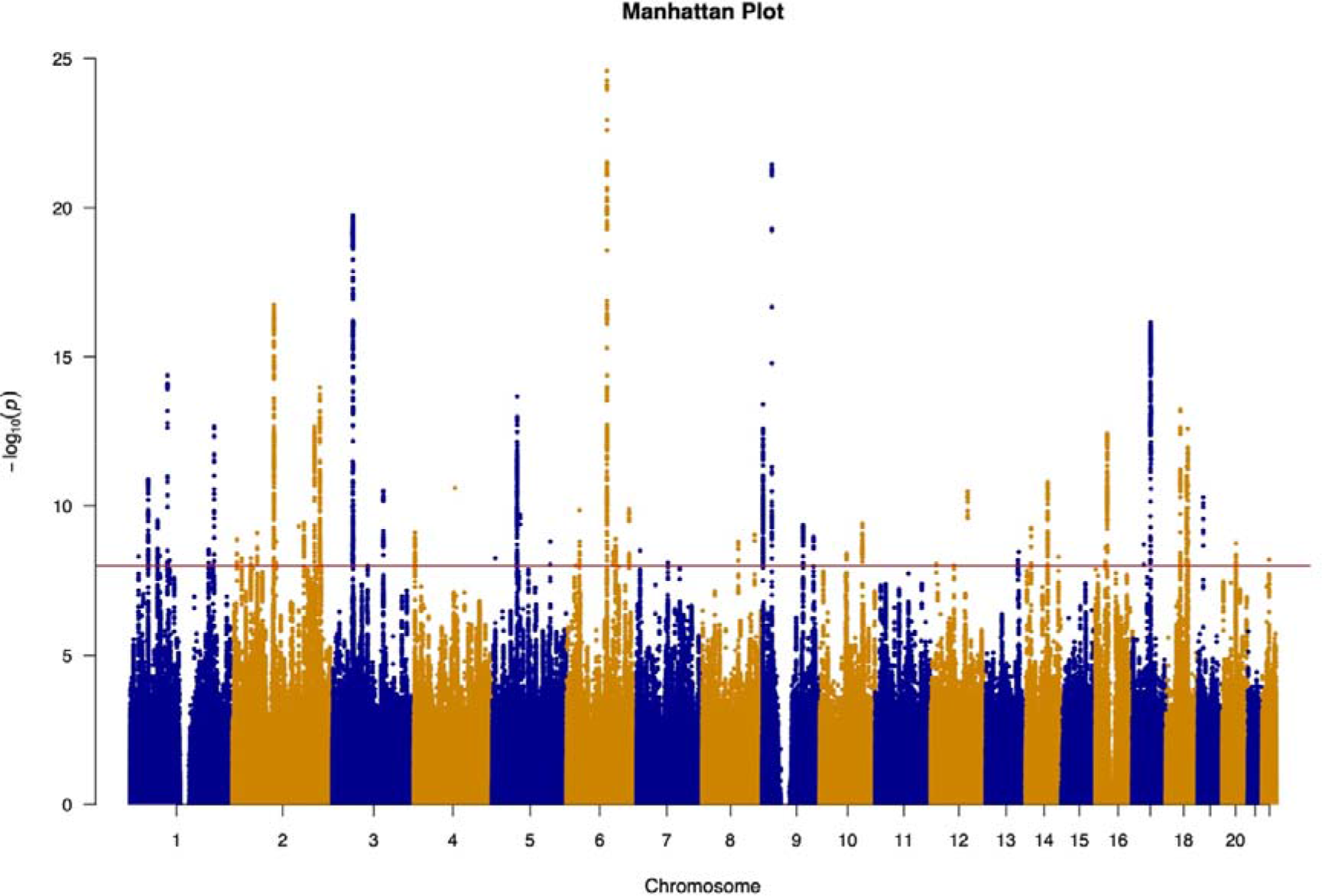
Manhattan plot of the latent poverty factor. Legend: The x-axis indicates chromosomal position and the y-axis is the significance of association (−log10(p-value)). The red line represents genome-wide significance level (5×10−8).

The genetic correlation between P and CA was strong (r_g_ was 0.74; see Supplementary Table 3). Running bidirectional MR of CA against P, we found stronger evidence supporting the causal effect of CA on P (IVW B_CA→P_=-0.390 [95%CI: -0.408; -0.372]) rather than vice versa (IVW B_P→CA_=-0.987 [95%CI: -1.03; -0.939]), as suggested by the results of Steger’s test (p<0.001; p=0.811, respectively). The full results are presented in Supplementary Table 4 and Supplementary Figures 1-4.

### Bidirectional univariable MR of common factor poverty against mental illness

For the univariable analyses, the mean F statistic ranged from 35.3 to 45.6, indicating that the estimates were not likely subject to weak instrument bias. Forward IVW analysis of P against the considered mental illnesses showed significant causal effect of P on ADHD (B=1.23 [95%CI: 1.08; 1.38]), ANX (B=0.831 [95%CI: 0.377; 1.28]), MDD (B=0.398 [95%CI: 0.254; 0.542]), PTSD (B=0.629 [95%CI: 0.395; 0.863]), SZ (B=0.425 [95%CI: 0.300; 0.550]), and with opposite direction on AN (B=-0.697 [95%CI:-0.918; -0.477]) and OCD (B=-0.676 [95%CI: -1.22; -0.137]). The estimate of the causal effect using WM method, was consistent in the magnitude for ADHD, AN, MDD, PTSD, and SZ, but not for ANX and OCD. We did not identify any significant causal effect of P on BD and ASD.

Backward IVW analysis of mental illness against P showed significant causal effect of ADHD (B=0.111 [95%CI: 0.096; 0.126]), BD (B=-0.026 [95%CI: -0.037; -0.014]), and SZ (B=0.023 [95%CI: 0.017; 0.029]) on P. The estimate of the causal effect using WM method, was confirmed for both ADHD and SZ, but not for BD.

The results are displayed in Figure 3 (see also scatterplots displayed in Supplementary Figures 5-13 and Supplementary Figures 23-26), and in the Supplementary Table 5 (see also Supplementary Table 6 for conversion to OR).

**Figure 3:**
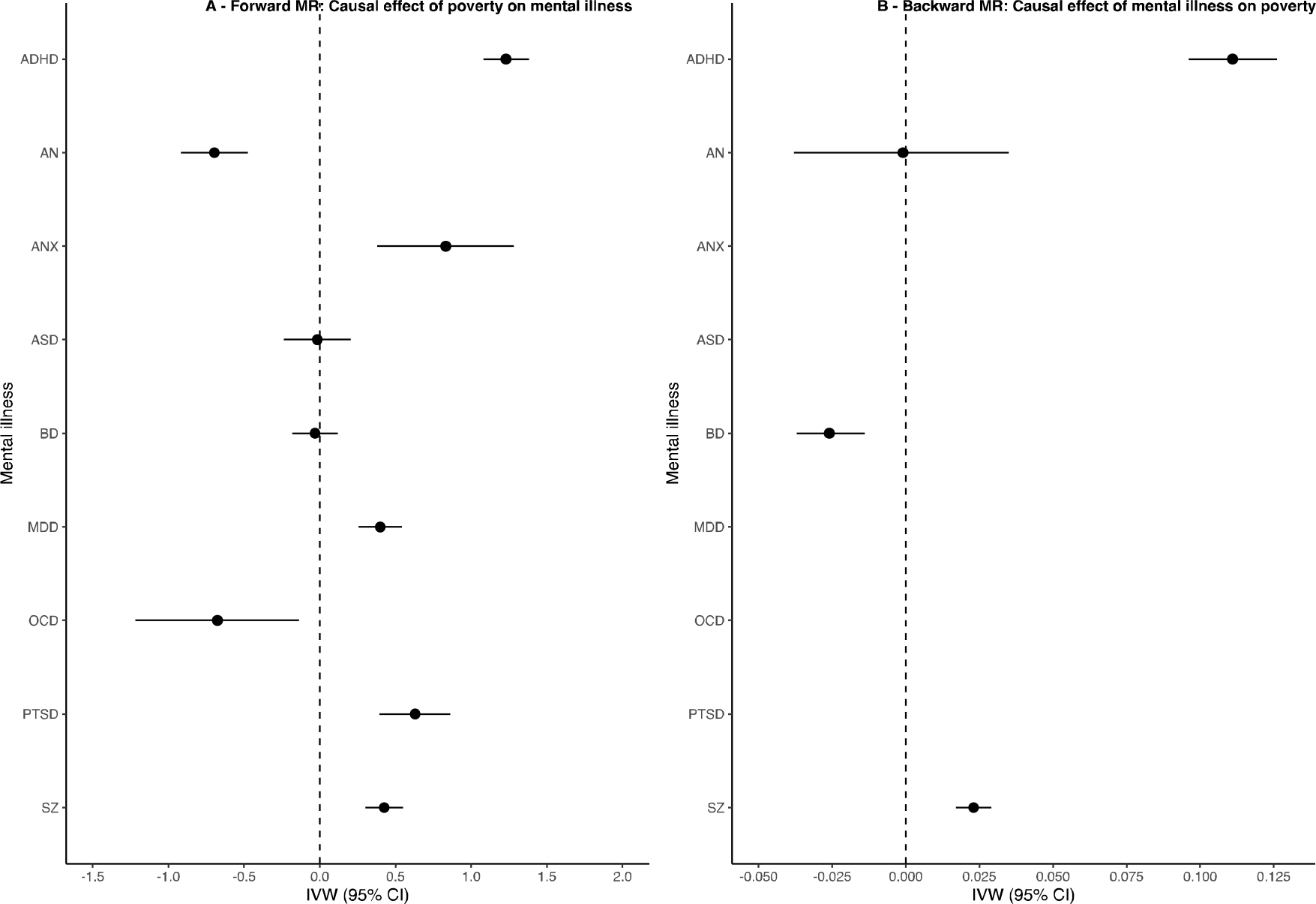
Forest plot of the results of univariable bidirectional Mendelian randomization analysis of poverty against mental illness (Panel A: forward analysis; Panel B: backward analysis) Abbreviations: Fw: forward MR analysis; Bw: backward MR analysis; ADHD: attention deficit hyperactivity disorder; AN: anorexia nervosa; ANX: anxiety disorder; ASD: autism spectrum disorders; BD: bipolar disorder; MDD: major depressive disorder; OCD: obsessive-compulsive disorder; PTSD: post-traumatic stress disorder; SZ: schizophrenia; MR: mendelian randomization; SNP: single nucleotide polymorphism; IVW: inverse variance weighted (fixed effect); 95% CI: 95% confidence interval. Legend: Poverty is a latent variable built using household income as unit identification, therefore an increase in the indicator’s load stands for increased income, therefore the regression coefficients have been reversed to facilitate interpretation of the effect of poverty. The effect estimates on the x-axis are log-odds for binary traits (i.e., for mental illnesses) and unstandardized linear regression coefficient for continuous traits (i.e., for poverty).

Steiger test indicated that the causal direction between the exposure and outcome was correct in all the analyses. Cochran’s Q heterogeneity statistics was significant in all the analyses (except for the effect of P on ANX), which is suggestive of pleiotropy. Furthermore, we looked at MR-Egger intercept, which was significantly deviating from 0 in the analysis of P against ADHD (p<0.001) and of P against ASD (p=0.015), suggesting that pleiotropy was unbalanced in these relationships. The MR-Egger causal effect, which provides better estimate in case of unbalanced pleiotropy than IVW MR, yielded opposite results for the effect of P on ADHD (B=-1.12 [95%CI: -2.21; -0.019]) and evidence of inverse relationship between P and ASD (B=-1.79 [95%CI: -3.23; -0.348], Egger’s intercept p=0.015). MR-PRESSO did not detect bias in the estimates due to horizontal pleiotropy in all the estimates. CAUSE analysis confirmed the causal effect of P on ADHD bidirectionally, and unidirectionally on AN, MDD, and PTSD. No significant difference between the sharing and the causal model was found for the effect of P on SZ bidirectionally. The results of CAUSE are presented in Supplementary Table 7.

Leave-one-out analyses, both in forward and backward directions, showed that the direction of the effect of each SNP is consistent with the direction of the overall causal estimate. In addition, for most of the analyses, a change in the estimations of less than 10% was obtained by leaving out each SNP, suggesting that none of the genetic variants were overly influential. The results of leave-one-out analyses are available as Supplementary Figures 14-22 and Supplementary Figures 27-29. Finally, to test if our results were robust to bias due to reverse causality, we repeated the MR analyses on a subset of SNPs selected through Steiger filtering. This sensitivity analysis confirmed the results of the main analysis, except for the effect of P on OCD that was no longer significant after Steiger filtering (B=-0.037 [95%CI: -0.702; 0.628]) and for the MR-Egger analysis of P on ADHD (B=-0.976 [95%CI: -1.94; 0.009]), suggesting that the relationship between P and ADHD is likely biased from reverse causation and unbalanced pleiotropy (as suggested by the Egger’s intercept test, p<0.001). The results are presented in Supplementary Table 8.

To further explore whether the effect of poverty on mental illness is driven by specific indicators, we performed bidirectional univariable MR using each of the poverty indicators (i.e., HI, OI, and SD) as the exposure. The results are presented in Figure 4, Supplementary Tables 9-10 (HI), 13-14 (OI), 17-18 (SD), and Supplementary Figures 30-107. The HI GWAS had a higher level of power than OI and SD GWASs, as indicated by the number of SNPs selected for the analyses (∼50, ∼30, and ∼10, respectively).

**Figure 4:**
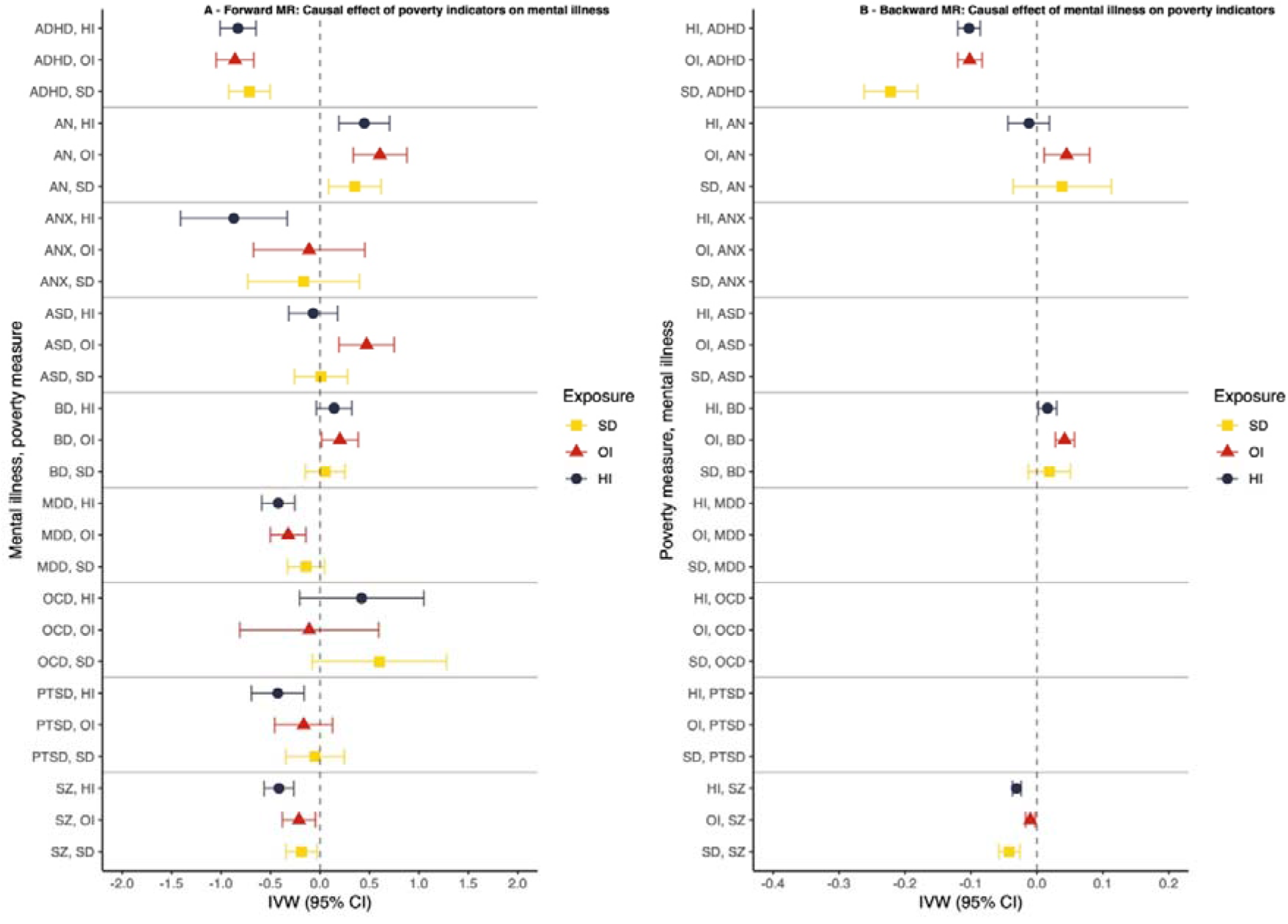
Forest plot of the results of univariable bidirectional Mendelian randomization analysis for each poverty measure against mental illness (Panel A: forward analysis; Panel B: backward analysis) Abbreviations: Fw: forward MR analysis; Bw: backward MR analysis; ADHD: attention deficit hyperactivity disorder; AN: anorexia nervosa; ANX: anxiety disorder; ASD: autism spectrum disorders; BD: bipolar disorder; MDD: major depressive disorder; OCD: obsessive-compulsive disorder; PTSD: post-traumatic stress disorder; SZ: schizophrenia; HI: household income; OI: occupational income; SD: social deprivation; MR: mendelian randomization; SNP: single nucleotide polymorphism; IVW: inverse variance weighted (fixed effect); 95% CI: 95% confidence interval. Legend: To have a consistent direction of the effect, social deprivation effect has been reversed. The effect estimates on the x-axis are log-odds for binary traits (i.e., for mental illnesses) and unstandardized linear regression coefficient for continuous traits (i.e., for the poverty indicators).

Overall, the direction of the associations obtained using the poverty common factor was consistent with those obtained for each indicator, where bidirectional causal effects of poverty on some mental illnesses was observed. Specifically, using HI as a poverty measure, we found evidence of bidirectional effect on ADHD (B=-0.830 [95%CI: -1.01; -0.647]; B=-0.103 [95%CI: -0.120; -0.086], IVW forward and backward respectively) and SZ (B=-0.415 [95%CI: -0.565; -0.265]; B=-0.031 [95%CI: -0.037; -0.024], IVW forward and backward respectively), and unidirectional causal effect to MDD (B=-0.422 [95%CI: -0.589; -0.255]) and AN (B=0.448 [95%CI: 0.191; 0.704]). However, the effect of HI on ADHD was biased by high heterogeneity and unbalanced pleiotropy (Egger’s intercept p<0.001), with MR-Egger estimate yielding inconsistent results (B=0.606 [95%CI: -0.684; 1.90]). The bidirectional effect of OI on ADHD (B=-0.859 [95%CI: -1.05; -0.669]; B=-0.102 [95%CI: -0.120; -0.083], IVW forward and backward respectively) and unidirectionally to AN (B=-0.187 [95%CI: - 0.344; -0.030]) and MDD (B=-0.322 [95%CI: -0.502; -0.143]) were confirmed also using OI as a measure of poverty. The effect of OI on SZ was also confirmed bidirectionally (B=-0.187 [95%CI: -0.344; -0.030]; B=-0.010 [95%CI: -0.017; -0.003]), but sensitivity analyses using CAUSE and Steiger filtering selection detected potential bias due to pleiotropy in the forward analysis. Notably, also the effect of OI and ADHD changed after Steiger filtering, with evidence of pleitropy (Egger’s intercept, p=0.033) and not significant MR-Egger estimate (B=0.094 [95%CI: -1.36; 1.55]). Backward analyses using OI as the outcome, confirmed the effect of BD on poverty using IVW but not WM methods (B=0.042 [0.028; 0.057]; B=0.020 [95%CI: -0.005; 0.046], IVW and WM respectively) as found using the common factor poverty; but novel evidence was found to the causal effect of AN to OI (B=0.045 [95%CI: 0.011; 0.080]), not replicated using the poverty common factor or other measures of poverty. Finally, using SD as a measure of poverty, the bidirectional effect on SZ (B=0.213 [95%CI: 0.046; 0.380]; B=0.042 [95%CI: 0.026; 0.058], IVW forward and backward respectively) and the unidirectional effect to AN (B=-0.352 [95%CI: -0.618; -0.087]) were confirmed. The bidirectional effect of SD on ADHD was also replicated (B=0.713 [95%CI: 0.504; 0.922]; B=0.222 [95%CI: 0.181; 0.262], IVW forward and backward respectively), with stronger evidence for the reverse effect as indicated by Steiger test (forward, p=0.079; backward, p<0.001). However, by applying the Steiger filtering selection, the effect of SD on ADHD held strong (B=0.557 [95%CI: 0.331; 0.784]). None of the other effects of SD remained significant after Steiger filtering, which is consistent with potential bias due to reverse causation in the relationship between SD and mental illness (for the CAUSE and Steiger filtering MR results see Supplementary Tables 11-12 [HI], 15-16 [OI], 19-20 [SD]).

### Bidirectional univariable MR of household income categories against mental illness

To explore the shape of the relationship between HI and mental health, and specifically to answer the question whether there is a particular HI threshold at which the effect of HI on mental health kicks-in, we used GWAS data for the HI categories and investigated the effect of each category on the considered mental illnesses. That led to the creation of 4 dichotomous dummy variables, consisting of: (1) low HI (LHI): cases were those <£18k, controls were those ≥£18k; (2) low-mid HI (LMHI): cases were those <£29.99k, controls were those ≥£29.99k; (3) mid-high HI (MHHI): cases were those >£52k, controls were those ≤£52k; (4) high HI (HHI): cases were those >£100k, controls were those ≤£100k. For each of these bidirectional univariable MR was performed.

As can be seen in Figure 5 and Supplementary Tables 21-24 (see also Supplementary Table 25 for conversion to OR), we found a significant causal effect of being in the lowest HI class on ANX (B=0.628 [95%CI: 0.095; 1.16]), BD (B=0.306 [95%CI: 0.125; 0.487]), MDD (B=0.351 [95%CI: 0.189; 0.513]), PTSD (B=0.506 [95%CI: 0.253; 0.759]), and bidirectionally on ADHD (forward, B=0.610; [95%CI: 0.415; 0.805]; backward, B=0.210 [95%CI: 0.170; 0.250]) and SZ (forward, B=0.648 [95%CI: 0.488; 0.808]; backward, B=0.082 [95%CI: 0.066; 0.098]). In the LMHI class, the direction of the associations observed in the LHI was maintained, and interestingly, the effect size decreased, consistent with our leading hypothesis of more deleterious effect of poverty on mental health at very low levels of income. Exceptions were AN, ASD, and OCD for which being in the LMHI class resulted to be protective (B=-0.309 [95CI: -0.526; -0.091], B=-0.268 [95%CI: -0.488; -0.047], B=-0.774 [95%CI: -1.34; -0.212], respectively). The analyses in the higher classes (i.e., MHHI and HHI) showed that with each incremental increase or decrease of income there is a corresponding effect on mental illness. For instance, AN and ASD - but not OCD – resulted causally associated with higher incomes, with stronger evidence for AN (B_MHHI→AN_=0.371 [95%CI: 0.193; 0.550]; B_MHHI→ASD_=0.480 [95%CI: 0.303; 0.658]; B_HHI→AN_=0.333 [95%CI: 0.065; 0.600]; B_HHI→ASD_=0.094 [95%CI: -0.121; 0.308]), whereas the association with ADHD, MDD, PTSD, and SZ resulted with opposite sign, confirming the causal relationship with lower income. Notably, ADHD and SZ showed a consistent bidirectional pattern across income levels, where a lower income was causally related to, and resulted also causal factors for lower incomes. This evidence is supporting a vicious circle between poverty and severe mental illness that reiterates.

**Figure 5:**
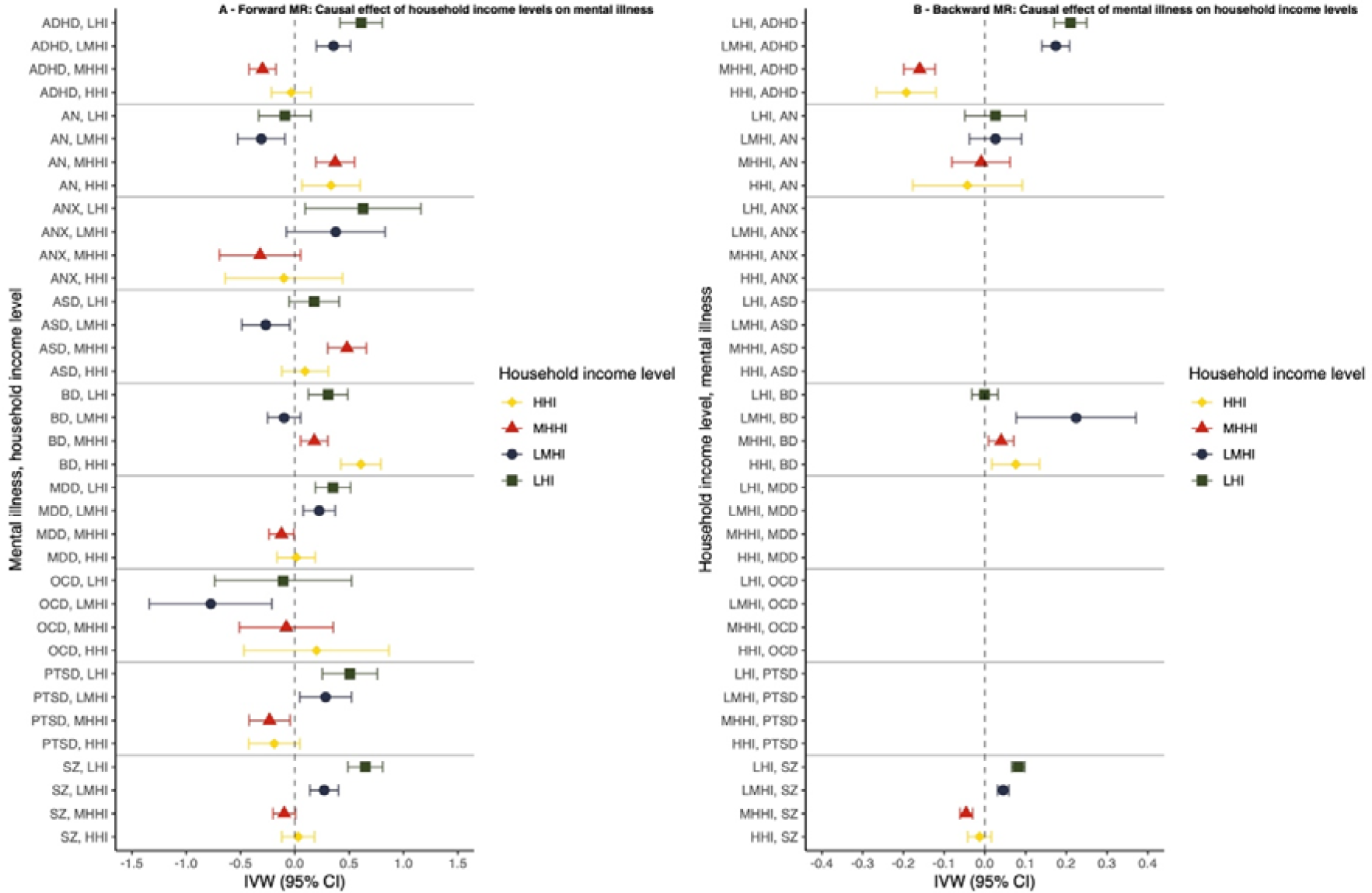
Forest plot of the results of univariable bidirectional Mendelian randomization analysis of household income levels against mental illnesses (Panel A: forward analysis; Panel B: backward analysis) Abbreviations: Fw: forward MR analysis; Bw: backward MR analysis; LHI: low household income; LMHI: low-mid household income; MHHI: mid-high household income; HHI: high household income; ADHD: attention deficit hyperactivity disorder; AN: anorexia nervosa; ANX: anxiety disorder; ASD: autism spectrum disorders; BD: bipolar disorder; MDD: major depressive disorder; OCD: obsessive-compulsive disorder; PTSD: post-traumatic stress disorder; SZ: schizophrenia; MR: mendelian randomization; SNP: single nucleotide polymorphism; IVW: inverse variance weighted (fixed effect); 95% CI: 95% confidence interval. Legend: LHI: low household income, cases <£18,000 – controls ≥£18,000; LMHI: low-mid HI, cases <£29,999 – controls ≥£30,000; MHHI: mid-high HI, cases >£52,000 – controls ≤£52,000; HHI: high HI, cases >£100,000 – controls ≤£100,000. The effect estimates on the x-axis are log-odds for both the forward and backward analyses given that all traits are binary.

The effect of the HI levels on BD showed a U-shape distribution, resulting causally associated with both low and high HI (B_LHI→BD_=0.306 [95%CI:0.125; 0.487]; B_MHHI→BD_=0.179 [95%CI: 0.053; 0.304]; B_HHI→BD_=0.608 [95%CI: 0.423; 0.792]). However, these associations were not replicated using the WM method, and MR-PRESSO detected significant distortion in the estimate due to pleiotropy in the relationship between BD and MHHI, with outlier adjusted causal estimates (OACE) resulting not significant (OACE=0.002 [95%CI: -0.039; 0.042]).

The estimate of the causal effect using WM method, was consistent in the magnitude for LHI to ADHD and SZ forward and backward, unidirectionally on ANX, MDD, PTSD, and for MHHI and HHI on ASD.

Q statistics were suggestive of high levels of heterogeneity in most of the analyses, however, MR-Egger showed significant intercept and causal effect only for the relationship between MHHI and BD (B=1.69 [95%CI: 0.376; 3.00]; Egger’s intercept p=0.033). Steiger test indicated that the causal direction between the exposure and outcome was correct in all the analyses except those on ASD (p=0.994 and p=0.057 respectively for the association with MLHI and MHHI).

Finally, to test if our results were robust to bias due to reverse causality, we repeated the MR analyses on a subset of SNPs selected through Steiger filtering. This sensitivity analysis confirmed a stronger causal effect played by the lowest HI level on ADHD, MDD, PTSD, and SZ. The protective effect of MHHI on ASD also was confirmed. Interestingly, the effect observed in the main analysis were not confirmed for BD and AN. The results can be found in Supplementary Table 26.

### MVMR of poverty factor and CA against mental illness

Like other measures of socioeconomic status, genetic effects are unlikely to influence poverty directly. Rather, genetic effects are likely to influence poverty through intermediary traits (such as health, personality, intelligence, and other characteristics) that are themselves heritable ^45^. Previous studies have shown that cognitive ability (referred to as general cognitive function, performance, or as intelligence) is one likely causal factor in both income differences ^45^ as well as being genetically associated with mental illness ^20^. MVMR was used to estimate the effect of poverty on each mental illness, while controlling for cognitive ability. First, bidirectional univariable MR of CA on mental illness was performed, finding evidence supporting a bidirectional inverse causal relationship between CA and ADHD (B=-0.638 [95%CI:-0.720; -0.556]; B=-0.151 [95%CI:-0.172; -0.131], forward and backward direction, respectively) and SZ (B=-0.297 [95%CI:-0.365; -0.228]; B=-0.055 [95%CI:-0.063; -0.047], forward and backward direction, respectively), unidirectional negative causal effect of CA on MDD and PTSD (B=-0.140 [95%CI:-0.215; -0.065] and B=-0.139 [95%CI:-0.264; -0.013], respectively) and unidirectional causal effect of CA on AN and ASD (B=0.306 [95%CI:0.190; 0.422] and B=0.310 [95%CI:0.193; 0.427], respectively). The results are displayed in Supplementary Tables 27-30, and Supplementary Figures 108-133. MVMR analysis yielded results supporting a causal effect of P on ADHD, AN, ANX, MDD, OCD, PTSD, and SZ, beyond CA. Still, the effect of P on mental illness decreased when including CA in the model. These results suggest that the univariable MR results of P on mental illness are slightly biased by CA, a direct effect of P on ADHD, AN, and ANX remains, nonetheless. The results of MVMR are shown in Table 2.

**Table 2:** Multivariable Mendelian Randomization results of poverty and cognitive abilities on mental illness

| **MVMR** | **N SNP** | **IVW B (95% CI)** | **p-value** |
| --- | --- | --- | --- |
| *Outcome: ADHD* | 211 |  |  |
| Exposure 1: P |  | 0.834 (0.606; 1.06) | <0.001 |
| Exposure 2: CA |  | -0.499 (-0.363; -0.634) | <0.001 |
| *Outcome: AN* | 213 |  |  |
| Exposure 1: P |  | -0.406 (-0.668; -0.144) | 0.002 |
| Exposure 2: CA |  | 0.158 (0.003; 0.313) | 0.046 |
| *Outcome: ANX*<br>Exposure 1: P<br>Exposure 2: CA | 213 | 0.411 (0.024; 0.798)<br>-0.290 (-0.518; -0.062) | 0.038<br>0.013 |
| *Outcome: ASD*<br>Exposure 1: P<br>Exposure 2: CA | 216 | 0.127 (-0.147; 0.401)<br>0.208 (0.045; 0.370) | 0.363<br>0.012 |
| *Outcome: BD*<br>Exposure 1: P<br>Exposure 2: CA | 216 | 0.057 (-0.165; 0.280)<br>-0.005 (-0.137; 0.128) | 0.614<br>0.947 |
| *Outcome: MDD*<br>Exposure 1: P<br>Exposure 2: CA | 216 | 0.266 (0.102; 0.429)<br>-0.093 (-0.190; 0.004) | 0.001<br>0.060 |
| *Outcome: OCD*<br>Exposure 1: P<br>Exposure 2: CA | 216 | -0.682 (-1.17; -0.196)<br>0.304 (0.015; 0.594) | 0.006<br>0.040 |
| *Outcome: PTSD*<br>Exposure 1: P<br>Exposure 2: CA | 216 | 0.477 (0.253; 0.701)<br>-0.159 (-0.293; -0.025) | <0.001<br>0.020 |
| *Outcome: SZ*<br>Exposure 1: P<br>Exposure 2: CA | 216 | 0.345 (0.082; 0.609)<br>-0.306 (-0.464; -0.148) | 0.010<br><0.001 |

Abbreviations: MVMR: multivariable Mendelian randomization; SNP: single nucleotide polymorphism; IVW: multivariable mendelian randomization via inverse variance weighted method (random effects); B: effect estimates are log-odds; 95% CI: 95% confidence intervals; ADHD: attention deficit hyperactivity disorder; P: poverty; CA: cognitive abilities; AN: anorexia nervosa; ANX: anxiety disorder; ASD: autism spectrum disorder; BD: bipolar disorder; MDD: major depressive disorder; OCD: obsessive-compulsive disorder; PTSD: post-traumatic stress disorder; SZ: schizophrenia
Legend: Poverty is a latent variable built using household income as unit identification, therefore an increase in the indicator's load stands for increased income, therefore the regression coefficients have been reversed to facilitate interpretation of the effect of poverty.

Importantly, and unlike the univariable models, the inclusion of CA in a MVMR model produced highly dissimilar results comparing between the poverty factor and each indicator of poverty. First, the inclusion of CA in a MVMR model removed the effect of HI on mental illness. Second, for both OI and SD some significant effects remained where OI demonstrated a direct effect on ASD and SD showed a direct effect on AN. The results of these additional MVMR models are shown in Supplementary Tables 30-33. Taken together these results support the idea that the majority of the genetic effects that link poverty to mental illness do act on CA. However, by utilizing a general factor of poverty the resulting increase in statistical power facilitated the discovery of genetic effects acting to link poverty to mental health independent of the effects of CA.

## Discussion

Building on data of 14 GWAS this study provides support of a causal relationship between poverty and mental illness. Jointly modeling different indicators of poverty using Genomic structural equation modeling, and subsequent MR, provided converging evidence supporting the bidirectional causal effect of poverty on ADHD and SZ, a unidirectional causal effect of poverty on MDD and an inverse causal relationship between poverty and AN. Notably, the evidence of causal relationship between poverty and ADHD was stronger for reverse causation (ADHD leading to poverty) rather than direct causality. These findings complement previous evidence of social inequalities in mental health across different countries, sample sizes, and study designs ^2,7,11, 46, 47^, but for the first time adds evidence that this relation is not merely an association caused by reversed causality (mental illness leading to poverty), but also of a causal role of poverty leading to mental illness. While this is not completely contra-intuitive, this study presents a first but important piece of evidence.

This study builds on genetic evidence but whilst poverty and other measures of socioeconomic status are heritable traits, it is very unlikely that there are direct genetic effects. Genetic associations with socioeconomic traits are likely the result of vertical pleiotropy. Vertical pleiotropy describes instances where one trait is causally associated with a second trait. In these cases, the genetic effects that act on the first trait will also be detected as being associated on the second ^48^. In that context, MR can best be viewed as a way to approximately randomly assigned heritable traits that give rise to income differences. Two key candidates of such underlying traits are intelligence and personality. For example, intelligence may lead to both educational advantage and socioeconomic success as well as more healthy behaviors and lead to good mental health ^49–53^. In this study we adjusted for cognitive ability and made two additional discoveries. First, when examining each of the indicators of poverty separately, controlling for CA removed the causal effect of HI on mental illness. For both OI and SD most of the evidence of a direct causal effect of poverty was absent after adjusting for CA however, lower levels of OI remained causally associated with increases in ADHD and higher SD remained in an inverse association with AN. Second, whilst controlling for CA the causal effect of the general factor of poverty reduced by around 30% compared with univariate estimates indicating the CA was a major contributor to the causal association between poverty and mental illness. However, the high power obtained by using the common factor of poverty facilitated the influence of the other heritable poverty traits above and beyond the role of CA and enabled detection of the causal role of poverty on mental health. Of note is the vast literature showing that health characteristics such as obesity, physical appearance, and mental state can influence economic outcomes including employment opportunities or wages ^54–56^. Therefore, our results of a causal effect of poverty on mental illness should not be taken as proof supporting genetic determinism, but rather as epidemiological evidence of the detrimental effects of poverty and socio-economic inequalities on mental health, regardless of the mechanisms (see also the Frequently Asked Questions [FAQ] in the Supplementary File 3).

The causal relationship between poverty and mental illness is likely to involve material, psychological, behavioral, and biological pathways. For example, the level of development of the welfare system may be a material mediator to the health-damaging effects of income losses ^57, 58^. Psychosocial mechanisms are a result of the interaction between people’s social environment and their feelings: living on low income is stressful, and at the same time people in disadvantaged situations may have fewer resources to cope with difficult circumstances.

Increasingly, biological research is providing evidence showing how experiences such as social defeat can “get under the skin” causing biochemical changes in the body and brain and increasing the risk of developing mental health problems ^59, 60^. Other contributing factors to the relation between poverty and mental illness are the negative health behaviors that are more prevalent in socially disadvantaged groups ^61^, likely as a consequence of the higher cost of healthy behaviors. For example, a healthy diet is more expensive than processed foods, and joining a gym or sporting clubs can be costly. Moreover, unhealthy behaviors such as smoking or drinking alcohol may be used as coping strategies for stressful situations ^62^. This aligns with the findings of our analysis on income categories, indicating that poverty does not have a continuous impact. Instead, experiencing extreme poverty is associated with an increased risk of mental illness.

Previous quasi-experimental research using natural experiments, such as on the lottery winner population, provided supportive evidence for causal relations between SES and mental health, although with small effect sizes ^63^. The effect sizes in this study type were moderate for the effect of poverty on ADHD, and small for the other relations.

### ADHD

Our finding of a causal association between poverty and ADHD is consistent with previous evidence that poverty has a greater impact on the development of mental illness in children and adolescents ^64^. It is important to note that the causal association between ADHD and poverty is subject to some uncertainty regarding the potential influence of reverse causation and pleiotropic effects. Since ADHD is generally recognized as a neurodevelopmental disorder, the observed association with lower CA assumes particular significance as a potential mechanism linking ADHD to socioeconomic disadvantage. Furthermore, the evidence of bidirectional causality between poverty and ADHD could also result from the intergenerational deleterious effects of poverty. For example, parental income may influence children’s health and children’s poor health may, in turn, impact their educational outcomes^65^.

### Anorexia nervosa

A limited number of studies have previously indicated that AN is more prevalent among affluent Western cultures ^66^. A recent cohort study in Denmark found that high parental SES was associated with an increased risk of AN among offspring ^67^, in line with our results. The same study also found that parental SES had a weaker association with Bulimia Nervosa and unspecified eating disorders in offspring, suggesting that parental SES has different effects on AN than on other eating disorders ^67^ or eating disorders as a whole^68, 69^. A recent review of the literature found that industrialization and urbanization were contributing factors to the rise of eating disorders in Asia ^70^. Altogether the current evidence is consistent with a model whereby higher income plays a role in the etiology of AN.

### Major depressive disorder

The literature on the relationship between poverty and depression is relatively extensive. Most studies suggest low income or income inequalities as risk factors for depression ^71–74^, in line with the causal effect of poverty on MDD here reported. Other poverty aspects like food insecurity, no former education, unemployment, and social-economic deprivation are also associated with depression ^75, 76^. However, analysis of SD on MDD risk in this study yielded no significant results. That may be due to the composition of the Townsend deprivation index used to measure deprivation in the UK Biobank. Interestingly, quasi-experimental studies on cash transfer programs and antipoverty programs effectively reduced symptoms of depression, providing more support for a causal effect in addition to prevalence or observational studies ^77, 78^.

### Schizophrenia

The existing literature suggests the association between SZ and lower income ^79–82^, lower SES ^83^, lower employment rate ^79, 83, 84^, and lower educational level ^79, 83^. According to a longitudinal cohort study in Denmark, the amount of time spent in low-income conditions during childhood is associated with an increased risk of SZ ^80^. The same study also revealed an increased risk of SZ for those who experienced a decrease in the parental income during childhood, whereas lower SZ risk was observed for those with upward parental income mobility, regardless of the baseline parental income at birth ^80^. All this evidence provides further support of causal effects of poverty, deprivation, and low HI and OI on SZ. The causal effect of CA on SZ also echoes previous evidence of inequalities in educational attainment and cognitive performance among people with schizophrenia ^85^. In addition, consistent with a reverse causal effect of SZ on poverty found in the current study, previous research detected a low employment rate among individuals suffering from SZ, particularly after being diagnosed with the disorder, for these people income transfer payments are usually the main source of income ^84^. Low income and deprivation were also acknowledged as severe psychological and physiological stressors able to worsen SZ symptoms ^81^, which sustain a negative circle of bidirectional causality.

### Other disorders

For the other disorders, the level of consistency across the analyses and previous literature is much lower. The literature on the relationship between poverty and PTSD is very scarce, overall suggesting that low income and material hardship are associated with the development of PTSD among those exposed to traumatic events ^86–88^. Moreover, in low- and middle-income countries, most people with PTSD do not access evidence-based treatment ^89^. An interesting finding concerns the risk of BD to the level of income. This exhibited a U-shape pattern, with an increased risk of BD for both lower and mid-high levels of income, which is similar to that found for the relationship between BD and intelligence in a previous study ^90^. The diagnosis of BD and MDD can be sometimes challenging due to the presence of overlapping symptoms. Therefore, the differential effect of income on these mental illnesses may be a useful insight for future research on their gene-environment underpinnings. The evidence for causal effects of poverty on ANX, ASD, BD, and OCD showed low consistency across sensitivity analyses, partly due to the lower power of the GWAS of ANX and OCD and to possible competing mechanisms and pleiotropic confounders for BD and ASD.

### Limitations

This study has limitations that should be considered when interpreting the results. First, for ANX, ASD, MDD, OCD, and PTSD, GWAS sample sizes were relatively small, precluding meaningful MR analysis. Future, more powerful GWAS on these conditions may yield instruments that are suitable for MR. Second, most genetic studies were conducted in populations of European ancestry from high income countries. Generalizability to populations from other ancestries, and to low- and medium-income countries, is therefore uncertain ^91^.

Particularly, this raises questions about the specificity and universality of the effects of poverty on mental health in different cultures and contexts with different political and social systems. Third, it is well acknowledged that psychiatric phenotypes are complex and heterogeneous, which generally translates to low power. This is reflected by the relatively modest effects of each poverty indicator on mental illness. Fourth, it is essential to acknowledge the potential influence of the dynastic effect, wherein characteristics transmitted across generations may contribute to the observed MR causal estimates. Detecting the exact magnitude of bias resulting from this effect is challenging. Therefore, we advise triangulation of our findings using complementary research methods in future studies. Finally, other limitations of MR concern temporality and linearity: MR is thought to provides estimates of lifetime risk and assumes linear effects. Although the use of MR on HI levels and the inclusion of mental illness with onsets across the whole person’s lifespan (e.g., ranging from ADHD and ASD to MDD and ANX) may have mitigated these limitations, future studies should particularly focus on assessing if there are critical windows or acute reactions to poverty exposure.

### Implications for research and practice

To the best of our knowledge, this is the first study to provide evidence of a causal effect of poverty on mental illness, using MR. Although individuals’ skills and abilities tied to earning capacity may be the variables with the actual causal effect on mental illness, this is a highly relevant message for public health and policy because our findings warrant further investigations into interventions targeting poverty and cognitive abilities to advance mental health. The choice of which action to take to address the problem is a political matter, but attention is warranted considering increasing income inequalities worldwide ^92^, as well as increasing incidence of mental illness ^93, 94^.

Previous studies examining antipoverty programs reported positive and sustained effects on mental health ^95^, including reduction of depression and suicide rates ^11, 96, 97^. For example, a recent RCT found that cash incentives provided to low-income individuals led to meaningful improvements in depression ^98^. Additionally, specific economic interventions such as family assets, employment support, and rental assistance may be effective in enhancing the mental health of program beneficiaries ^99, 100^. However, it is important to note that certain aspects of these interventions, such as the perception of receiving money as a debt or burdensome responsibility ^101, 102^, may contribute to increased stress ^103, 104^. Therefore, further research is necessary to identify the factors within economic support interventions (e.g., microcredit, loans, personal health budgets) that influence mental health outcomes. Early studies that combined antipoverty interventions, such as financial incentives and financial mentoring programs, appeared more effective in improving mental health compared to providing financial incentives alone ^97^. Given these complexities, policymakers could consider policies that can be beneficial both for individuals living in poverty and those living with severe mental illness. Policymakers should also consider a key finding of this study about the effect of cognitive ability in the causal pathway linking poverty to mental illness. CA are closely associated with educational attainment and occupational status, which are often regarded as socio-economic status variables relevant to health ^49, 51, 105^. Although the genetic relationship between CA and mental illness differs from the genetic relationship between education and mental illness, education and occupation may still serve as potential targets for interventions. Future research should explore strategies aimed at facilitating individuals’ participation in education and employment, which may lead to better mental health ^8,106^.

## Conclusions

This study presents evidence supporting causal effects of poverty on mental illness particularly for SZ and MDD. Conversely, AN was found in an inverse causal relationship with poverty, suggesting increased risk of AN among less poor people. Taken together with previous literature this research points to new possibilities for public mental health improvement, a possibility that should be welcomed in an era with increasing health inequalities.

## Data Availability

The lead SNPs list and summary statistics of the common factor poverty GWAS is available as Supplementary File 2. The full GWAS summary statistics is available upon reasonable request to the lead author (MM).
The codes for replicating the analyses can be accessed here: https://github.com/MattiaMarchi/Common-factor-GWAS---MR

## Acknowledgement

Thanks to Prof. Maria Cristina Murari from University of Modena and Reggio Emilia, for the help with HPC.

## Data availability statement

The lead SNPs list and summary statistics of the common factor poverty GWAS is available as Supplementary File 2. The full GWAS summary statistics is available upon reasonable request to the lead author (MM).

The codes for replicating the analyses can be accessed here: https://github.com/MattiaMarchi/Common-factor-GWAS---MR

