## Supplementary File 1 for "The impact of poverty on mental illness: Emerging evidence of a causal relationship"

Summary

### **Methods details**

#### **Household income and social deprivation GWAS**

*Samples*

Among 502,408 UK biobank participants, 440,964 samples of recent European ancestry that have genetic information, have identical reported sex as genotype inferred sex, not have sex chromosome aneuploidy, not have been detected as extreme outliers of heterozygosity and missingness, and have a genotyping rate greater than 0.9 were retained in our analysis. European ancestry was identified as self-reported whites of which each of their first six principal components (PC) was within its corresponding 99.7% confidence intervals (i.e., mean ± 3 standard deviations).

*Genotype and imputation data*

A total number of 784,256 and 93,095,624 autosomal variants are available in UK biobank genotype and imputation data, respectively. For genotype data, 581,097 variants with MAF > 0.01, call rate > 0.9, and HWE-p value > 10^-15^ in the European subset were kept. For imputation data, 25,590,374 variants with MAF >= 0.0005 and INFO >= 0.3 in the whole population were retained for the GWAS.

*Phenotype*

Six phenotypes were extracted for study samples. Social deprivation is a continuous trait with N = 440,350. Household income is a five-level category trait with N = 379,598 (‘Do not known’ and ‘Prefer not to answer’ removed). Another four binary traits were derived from household income. Low income (coded 1 for level 1 and 0 for the rest), LowMid income (coded 1 for levels 1 and 2 and 0 for the rest), MidHigh income (coded 1 for levels 4 and 5 and 0 for the rest), and High income (coded 1 for level 5 and 0 for the rest).

*GWAS using Regenie*

GWAS was conducted in Regenie v3.1.3, a two-step GWAS software that accounts for sample relatedness and population structure. In the first step, a whole genome regression model was fit to each trait using 581,097 post-QC genotype variants. In the second step, association test was performed for each of the 25,590,374 post-QC imputed variant using a LOCO (leave-one-chromosome out) scheme. The per-chromosome LOCO genomic predictions produced in the first step were fitted in the second step to account for sample relatedness and population structure. In addition, sex, age at assessment, assessment centers, genotyping array, genotyping bathes, and the first 40 PCs were fitted as covariates in both steps. For binary phenotypes, firth logistic regression test was performed in the second step to account for unbalanced case-control ratio. Afterwards, variants with MAF < 0.0005 and INFO < 0.3 in each subset were removed, resulting in 20,408,331 final variants for household income related phenotypes and 20,413,590 for Townsend score.

#### **MR-PRESSO**

The Mendelian randomization pleiotropy residual sum and outlier (MR-PRESSO) test identifies possible bias from horizontal pleiotropy. The test consists of three parts. (1) the MR-PRESSO global test which detects horizontal pleiotropy. (2) the outlier corrected causal estimate which corrects for the detected horizontal pleiotropy and (3) the MR-PRESSO distortion test which estimates if the causal estimate is significantly different (at p<0.05) after adjustment for outliers. We conduct all three stages (with the argument NbDistribution=1000. namely using1000 simulation form the null distribution to compute empirical p-values) and present the outlier adjusted causal estimates (OACE) when both global and distortion tests are significant.

#### **CAUSE**

In addition to MR-Egger and MR-PRESSO. we accounted for false positive due to horizontal pleiotropy using Causal Analysis Using Summary Effect Estimates (CAUSE).

CAUSE grounds on estimating two models. One under the assumption that the relation between the instrument and the outcome is due to a pleiotropic effect (i.e., the shared model). and the second assuming that the relation is due to causal effect (i.e., the causal model). CAUSE also provides a test that the posteriors estimated under the causal model fit the data significantly better than posteriors estimated under the sharing model. If this is the case, it is possible to conclude that the data are consistent with a causal effect.

CAUSE consists in 4 steps: (1) format the data for use with CAUSE; (2) calculate nuisance parameters; (3) LD pruning; (4) fit CAUSE. We estimated nuisance parameters setting a random subset of 1.000.000 variants and performed LD pruning setting R^2^ threshold to 0.01 and p-value threshold to 0.001.

### **Multivariable GWAS of the latent poverty factor**

#### Supplementary Table 1: factor loading of each poverty indicator used for the estimation of the latent poverty factor

| **Regression** | **Unstandardized B (SE)** | **Standardized B (SE)** | **p-value** |
| --- | --- | --- | --- |
| F~HI | 0.280 (0.007) | 1,00 (0.028) | <0.001 |
| F~SD | -0.127 (0.004) | -0.733 (0.025) | <0.001 |
| F~OI | 0.261 (0.008) | 0.862 (0.025) | <0.001 |

Abbreviations: F: common factor; HI: household income; SD: social deprivation; OI: occupational income; B: linear regression coefficient; SE: standard error.

#### Supplementary Table 2: Summary statistics of the common poverty factor, household income, social deprivation, and occupational income

|  | **Mean Chi^2^** | **LDSC intercept (SE)**  **[N SNPs]** | **Heritability**  **h^2^ % (SE)** |
| --- | --- | --- | --- |
| HI | 1.5669 | 1.0426 (0.0099)  [1165506] | 7.08% (0.0031)  p<5e-8 |
| SD | 1.3379 | 1.0423 (0.0081)  [1165534] | 3.01% (0.0015)  p<5e-8 |
| OI | 1.5145 | 1.0015 (0.0092)  [1177612] | 9.14% (0.004)  p<5e-8 |
| Common factor: P | 1.7301 | 0.9883 (0.0108)  [1158117] | 8.38% (0.0031)  p<5e-8 |

Abbreviations: LDSC: Linkage Disequilibrium Score Regression; SE: standard error; P: poverty; HI: household income; SD: social deprivation; OI: occupational income.

Legend: Mean Chi^2^ measures the overall strength of association between genetic variants and the phenotype of interest; a high Mean Chi^2^ value indicates that there are many genetic variants that are strongly associated with the trait. The Linkage Disequilibrium Score Regression (LDSC) intercept captures the contribution of factors other than polygenicity (such as population stratification) to inflation in association test statistic. Narrow sense heritability (h²) is a measure of the proportion of phenotypic variation that is attributable to genetic variation.

#### Supplementary Table 3: genetic correlation (rg) between the common factor poverty, household income, social deprivation, occupational income, and cognitive abilities

|  | **P** |  |  |  |  |
| --- | --- | --- | --- | --- | --- |
| **P** | 1.00 | **HI** |  |  |  |
| **HI** | 0.9826  (0.0398) | 1.00 | **SD** |  |  |
| **SD** | -0.8007  (0.0344) | -0.7712  (0.0345) | 1.00 | **OI** |  |
| **OI** | 0.9509  (0.0372) | 0.9067  (0.0391) | -0.6318  (0.0325) | 1.00 | **CA** |
| **CA** | 0.7396  (0.0289) | 0.7019  (0.0315) | -0.4092  (0.0276) | 0.8147  (0.0334) | 1.00 |

Abbreviations: P: poverty; HI: household income; SD: social deprivation; OI: occupational income; CA: cognitive abilities.

Legend: genetic correlations are presented as rg(standard error)

#### Supplementary Table 4: Results of bidirectional Mendelian Randomization of Cognitive Abilities against Poverty

| **MR** | **N SNP** | **IVW, B (95% CI)** | **p-value** | **WM, B (95% CI)** | **p-value** | **MR-Egger, B (95% CI)** | **p-value** | **Egger intercept p-value** | **Steiger Test p-value** | **MR-PRESSO** | **Mean F** |
| --- | --- | --- | --- | --- | --- | --- | --- | --- | --- | --- | --- |
| Fw: CA on P | 133 | -0.390 (-0.408; -0.372) | <0.001 | -0.352 (-0.387; -0.317) | <0.001 | -0.458 (-0.591; -0.326) | <0.001 | 0.303 | <0.001 | DT; p=0.851 | 44.0 |
| Bw: P on CA | 86 | -0.987 (-1.03; -0.939) | <0.001 | -0.865 (-0.961; -0.770) | <0.001 | -1.12 (-1.51; -0.728) | <0.001 | 0.499 | 0.811 | DT; p=0.358 | 40.0 |

Abbreviations: Fw: forward analysis; Bw: backward analysis; CA: cognitive abilities; P: common factor poverty; MR: mendelian randomization; SNP: single nucleotide polymorphism; IVW: inverse variance weighted (fixed effect); B: effect estimates are unstandardized regression coefficient; 95% CI: 95% confidence interval; WM: weighted median; DT: distortion test; GT: global test.

Legend:

Legend: Poverty is a latent variable built using household income as unit identification, therefore an increase in the indicator’s load stands for increased income, therefore the regression coefficients have been flipped to facilitate interpretation of the effect of poverty.

P-value threshold <5e-8

^a^ The Mendelian randomization pleiotropy residual sum and outlier (MR-PRESSO) test identifies possible bias from horizontal pleiotropy. The test consists of three parts, (1) the MR-PRESSO global test which detects horizontal pleiotropy, (2) the outlier corrected causal estimate which corrects for the detected horizontal pleiotropy and (3) the MR-PRESSO distortion test which estimates if the causal estimate is significantly different (at p<0.05) after adjustment for outliers. We conduct all three stages (with the argument NbDistribution=1000, namely using1000 simulation form the null distribution to compute empirical p-values) and present the outlier adjusted causal estimates (OACE) when both global and distortion tests are significant.

^b^ We did not run Steiger Test if none of the MR analysis resulted significant (NR: not reported in the cell).

^c^ Not enough SNP to perform MR (NR: not reported in the cell).

#### Plots - Forward analyses

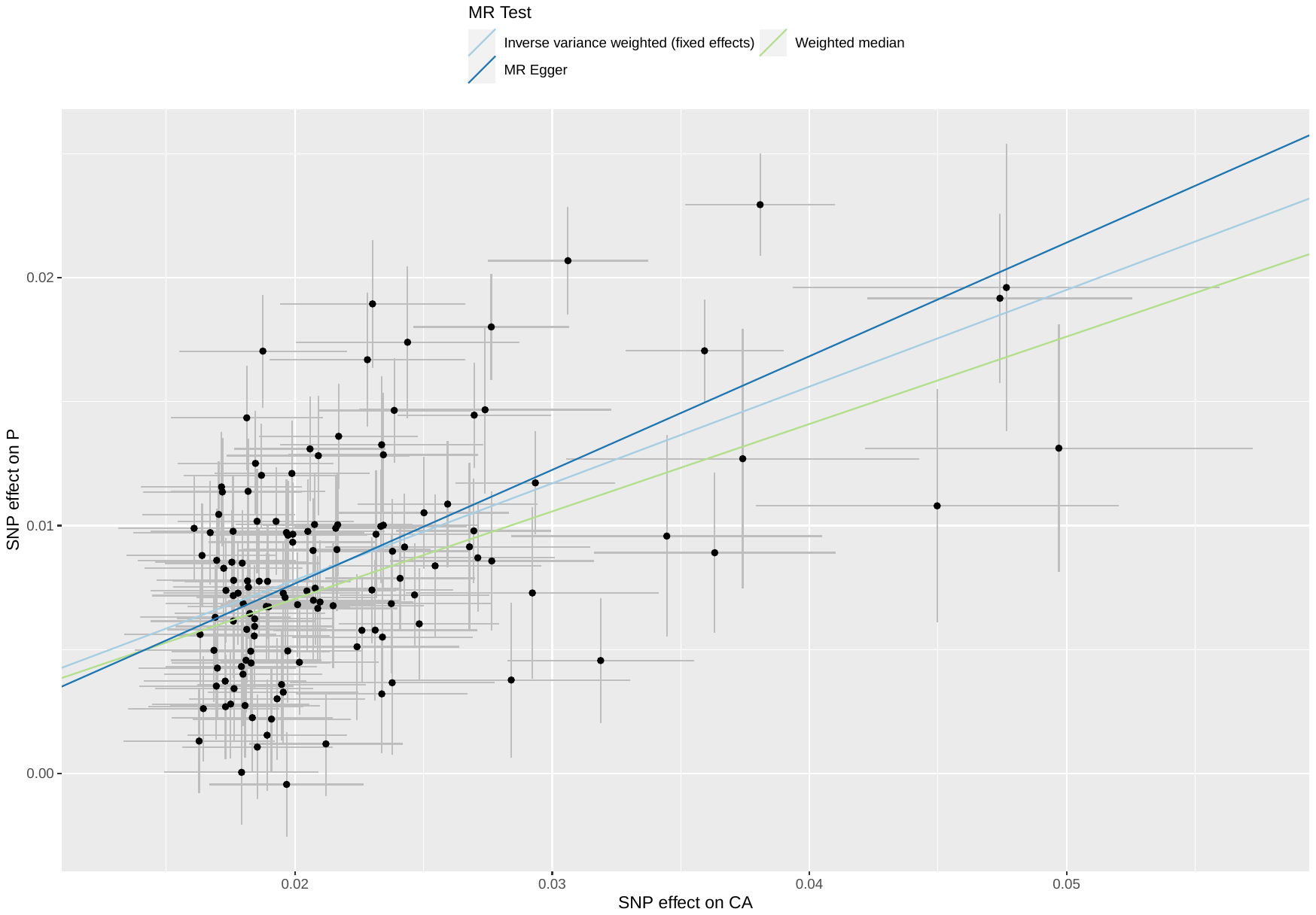

##### Supplementary Figure 1: scatterplot of cognitive abilities against common factor poverty

Abbreviations: MR: Mendelian randomization; SNP: single nucleotide polymorphism; CA: cognitive abilities; P: common factor poverty

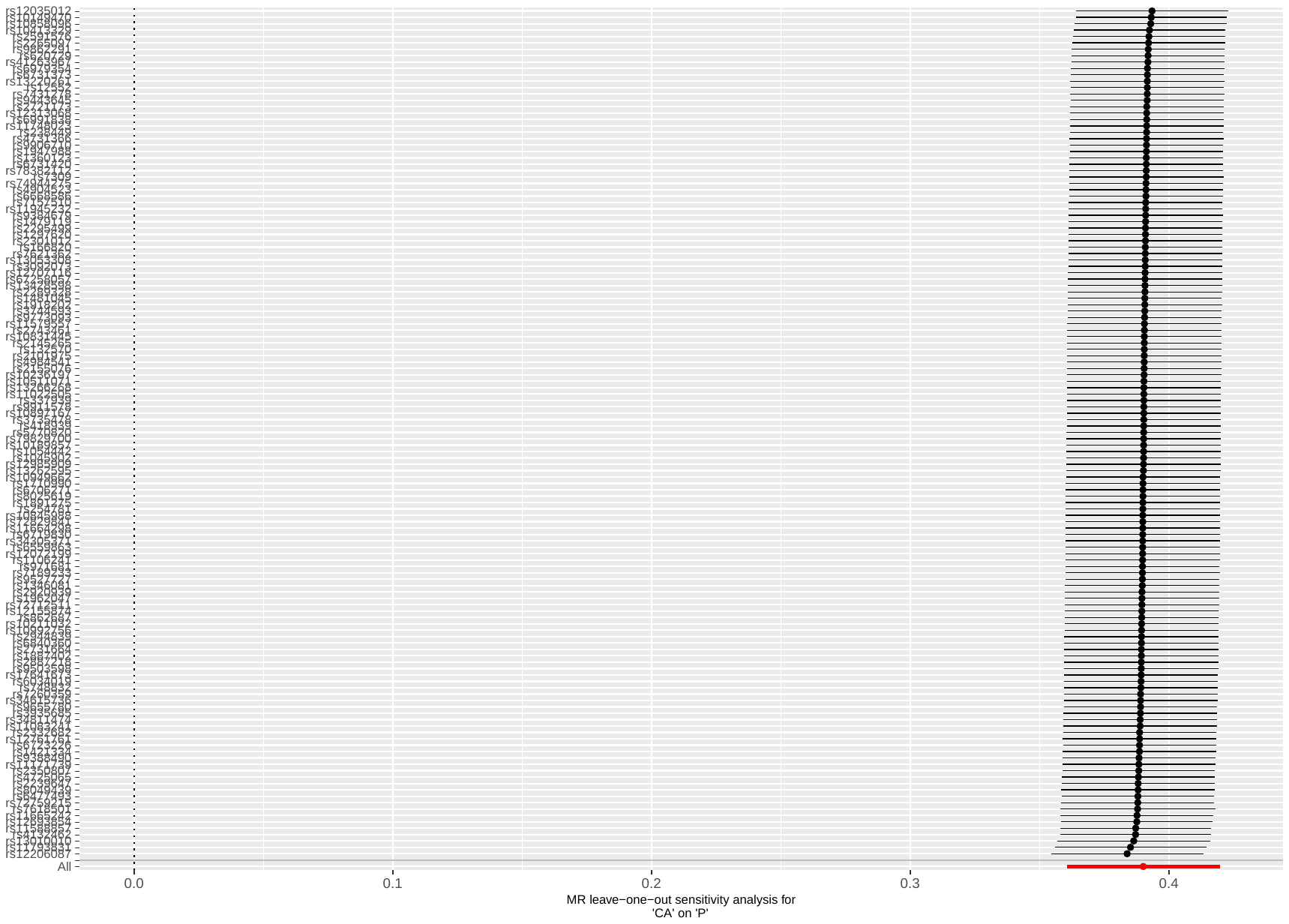

##### Supplementary Figure 2: leave-one out analysis of cognitive abilities against common factor poverty

Abbreviations: MR: Mendelian randomization; CA: cognitive abilities; P: common factor poverty

#### Plots - Backward analyses

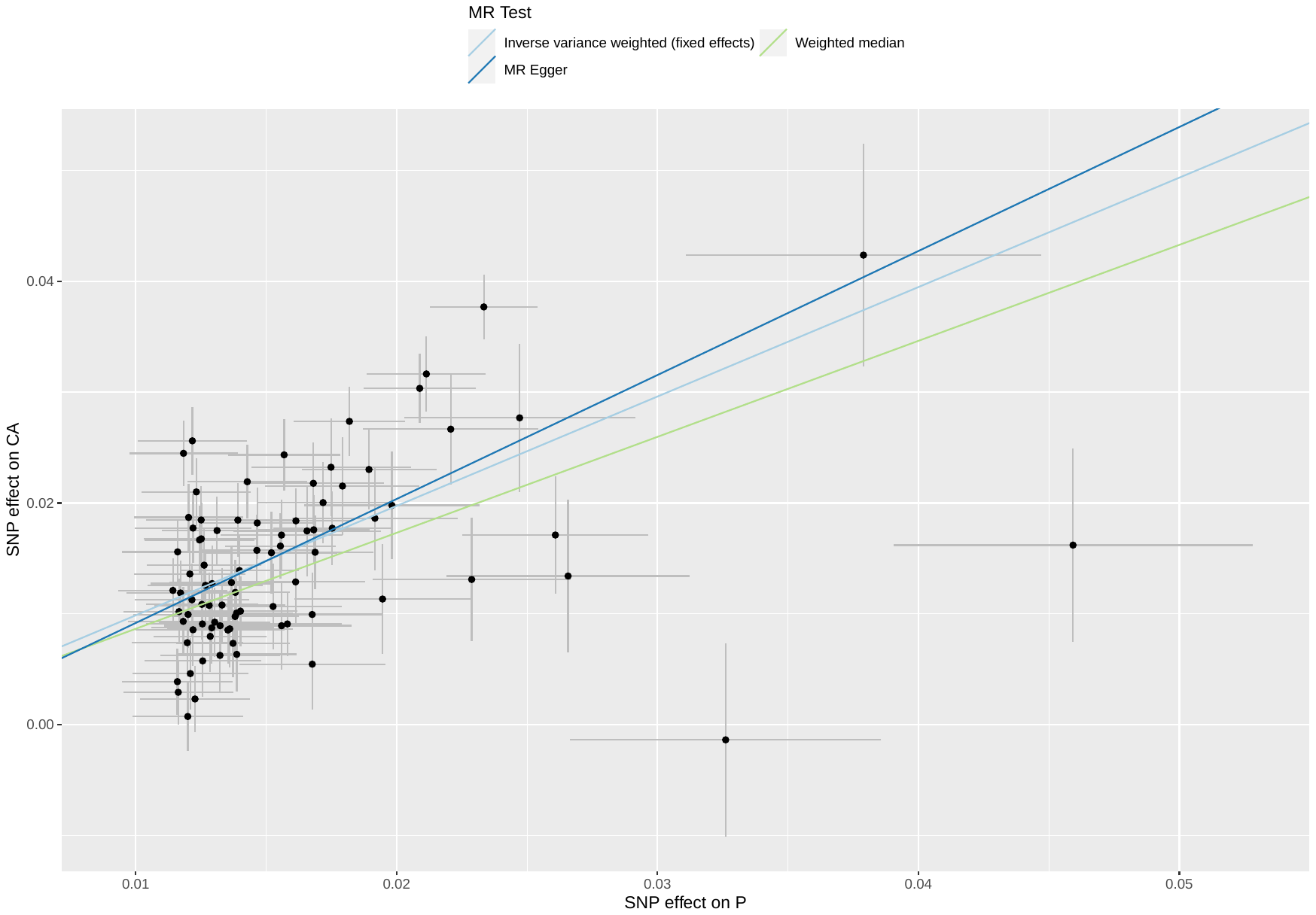

##### Supplementary Figure 3: scatterplot of common factor poverty against cognitive abilities

Abbreviations: MR: Mendelian randomization; SNP: single nucleotide polymorphism; CA: cognitive abilities; P: common factor poverty

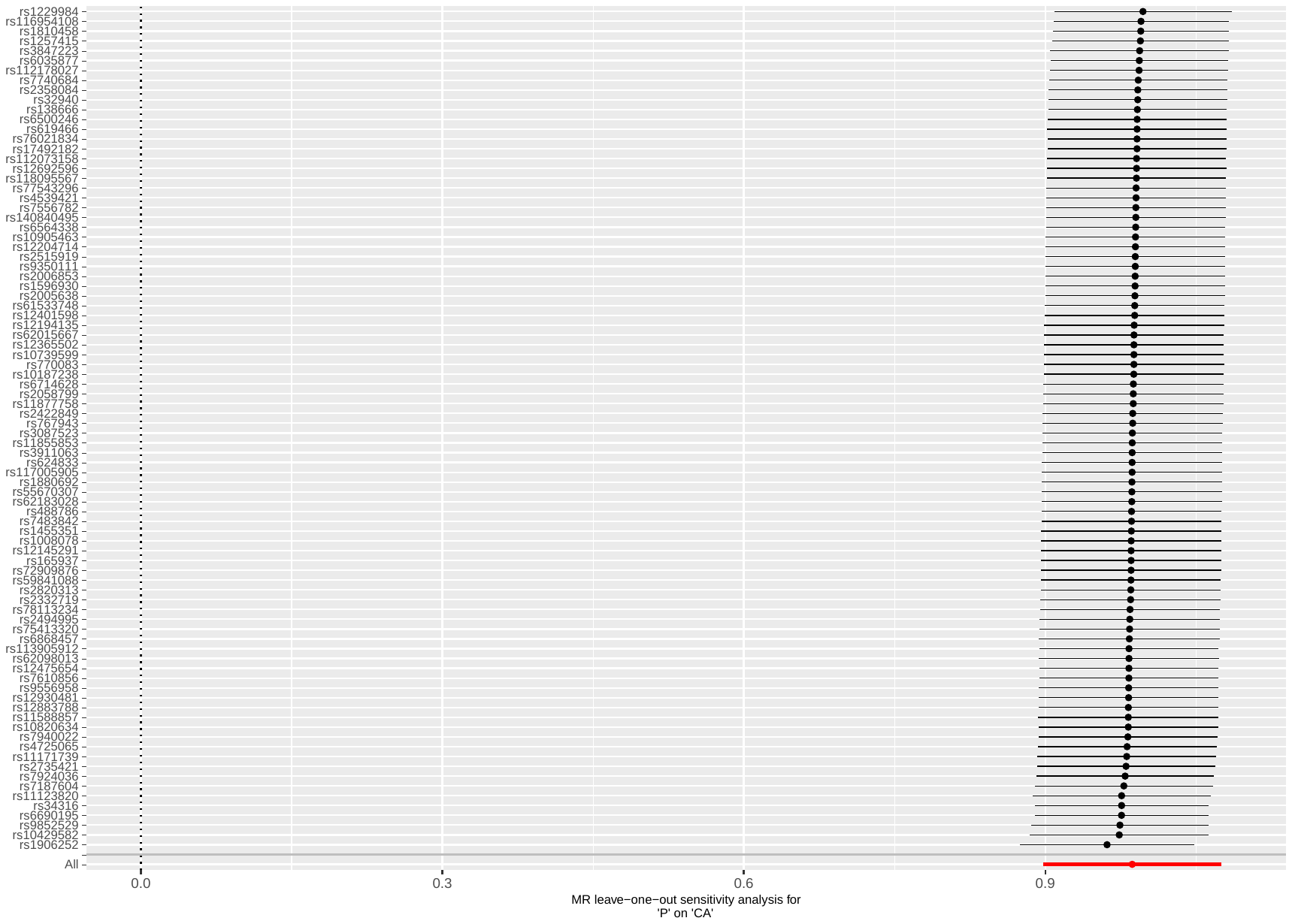

##### Supplementary Figure 4: leave-one out analysis of common factor poverty against cognitive abilities

Abbreviations: MR: Mendelian randomization; CA: cognitive abilities; P: common factor poverty

### **Univariable Mendelian randomization of poverty and mental illnesses**

#### Supplementary Table 5: results of bidirectional MR of poverty against mental illness

| **MR** | **N SNP** | **IVW, B (95% CI)** | **p-value** | **IVW Q(df)** | **Q p-value** | **WM, B (95% CI)** | **p-value** | **MR-Egger, B (95% CI)** | **p-value** | **Egger intercept p-value** | **Steiger Test p-value** | **MR-PRESSO** | **Mean F** |
| --- | --- | --- | --- | --- | --- | --- | --- | --- | --- | --- | --- | --- | --- |
| Fw: P on ADHD | 85 | 1.23 (1.08; 1.38) | <0.001 | 243 (84) | <0.001 | 0.808 (0.530; 1.09) | <0.001 | -1.12 (-2.21; -0.019) | 0.049 | <0.001 | <0.001 | DT; p=0.633 | 40.5 |
| Bw: ADHD on P | 22 | 0.111 (0.096; 0.126) | <0.001 | 102 (21) | <0.001 | 0.097 (0.069; 0.125) | <0.001 | 0.036 (-0.159; 0.230) | 0.723 | 0.448 | <0.001 | GT; p=0.853 | 38.3 |
| Fw: P on AN | 85 | -0.697 (-0.918; -0.477) | <0.001 | 160 (84) | <0.001 | -0.691 (-1.05; -0.337) | <0.001 | -1.29 (2.74; 0.160) | 0.085 | 0.415 | <0.001 | DT; p=0.786 | 40.4 |
| Bw: AN on P | 2 | -0.001 (-0.038; 0.035) | 0.947 | 7 (1) | 0.008 | NR ^c^ | NR ^c^ | NR ^c^ | NR ^c^ | NR ^c^ | NR ^b^ | NR ^c^ | NR ^c^ |
| Fw: P on ANX | 85 | 0.831 (0.377; 1.28) | <0.001 | 92 (84) | 0.257 | 0.590 (0.095; 1.27) | 0.091 | -0.054 (-2.41; 2.30) | 0.964 | 0.455 | <0.001 | GT; p=0.270 | 40.6 |
| Bw: ANX on P | 0 | NR ^c^ | NR ^c^ | NR ^c^ | NR ^c^ | NR ^c^ | NR ^c^ | NR ^c^ | NR ^c^ | NR ^c^ | NR ^c^ | NR ^c^ | NR ^c^ |
| Fw: P on ASD | 87 | -0.018 (-0.239; 0.203) | 0.873 | 199 (86) | <0.001 | -0.074 (-0.426; 0.279) | 0.682 | -1.79 (-3.23; -0.348) | 0.017 | 0.015 | <0.001 | DT; p=0.981 | 40.3 |
| Bw: ASD on P | 0 | NR ^c^ | NR ^c^ | NR ^c^ | NR ^c^ | NR ^c^ | NR ^c^ | NR ^c^ | NR ^c^ | NR ^c^ | NR ^c^ | NR ^c^ | NR ^c^ |
| Fw: P on BD | 87 | -0.032 (-0.181; 0.118) | 0.678 | 264 (86) | <0.001 | -0.048 (-0.302; 0.206) | 0.713 | -0.727 (1.90; 0.449) | 0.229 | 0.238 | NR ^b^ | DT; p=0.163 | 40.3 |
| Bw: BD on P | 36 | -0.026 (-0.037; -0.014) | <0.001 | 233 (35) | <0.001 | -0.004 (-0.025; 0.017) | 0.732 | -0.016 (-0.179; 0.147) | 0.847 | 0.909 | <0.001 | DT; p=0.110 | 39.2 |
| Fw: P on MDD | 86 | 0.398 (0.254; 0.542) | <0.001 | 150 (85) | <0.001 | 0.300 (0.071; 0.528) | 0.010 | -0.126 (-0.964; 0.713) | 0.770 | 0.212 | <0.001 | DT; p=0.824 | 40.0 |
| Bw: MDD on P | 0 | NR ^c^ | NR ^c^ | NR ^c^ | NR ^c^ | NR ^c^ | NR ^c^ | NR ^c^ | NR ^c^ | NR ^c^ | NR ^c^ | NR ^c^ | NR ^c^ |
| Fw: P on OCD | 86 | -0.676 (-1.22; -0.137) | 0.014 | 100 (85) | 0.132 | -0.484 (-1.29; 0.327) | 0.242 | 0.516 (-1.91; 2.95) | 0.678 | 0.325 | 0.090 | GT; p=0.141 | 40.0 |
| Bw: OCD on P | 0 | NR ^c^ | NR ^c^ | NR ^c^ | NR ^c^ | NR ^c^ | NR ^c^ | NR ^c^ | NR ^c^ | NR ^c^ | NR ^c^ | NR ^c^ | NR ^c^ |
| Fw: P on PTSD | 87 | 0.629 (0.395; 0.863) | <0.001 | 118 (86) | 0.013 | 0.460 (0.100; 0.820) | 0.012 | -0.526 (-1.72; 0.667) | 0.390 | 0.055 | <0.001 | DT; p=0.952 | 40.3 |
| Bw: PTSD on P | 0 | NR ^c^ | NR ^c^ | NR ^c^ | NR ^c^ | NR ^c^ | NR ^c^ | NR ^c^ | NR ^c^ | NR ^c^ | NR ^c^ | NR ^c^ | NR ^c^ |
| Fw: P on SZ | 87 | 0.425 (0.300; 0.550) | <0.001 | 547 (86) | <0.001 | 0.457 (0.193; 0.720) | 0.001 | -0.160 (-1.44; 1.12) | 0.808 | 0.360 | <0.001 | DT; p=0.333 | 40.3 |
| Bw: SZ on P | 176 | 0.023 (0.017; 0.029) | <0.001 | 706 (175) | <0.001 | 0.014 (0.004; 0.025) | 0.006 | 0.002 (-0.043; 0.047) | 0.922 | 0.352 | <0.001 | DT; p=0.351 | 45.6 |

Abbreviations: Fw: forward analysis; Bw: backward analysis; P: poverty; ADHD: attention deficit hyperactivity disorder; AN: anorexia nervosa; ANX: anxiety disorder; ASD: autism spectrum disorder; BD: bipolar disorder; MDD: major depressive disorder; OCD: obsessive-compulsive disorder; PTSD: post-traumatic stress disorder; SZ: schizophrenia; MR: mendelian randomization; SNP: single nucleotide polymorphism; IVW: inverse variance weighted (fixed effect); B: effect estimates are log-odds for binary traits (i.e., for mental illnesses) and unstandardized regression coefficient for continuous traits (i.e., for poverty); 95% CI: 95% confidence interval; Q: Cochran’s Q measure of heterogeneity; df: degree of freedom; WM: weighted median; DT: distortion test; GT: global test.

Legend:

P-value threshold <5e-8

Poverty is a latent variable built using household income as unit identification, therefore an increase in the indicator’s load stands for increased income, therefore the regression coefficients have been reversed to facilitate interpretation of the effect of poverty.

^a^ The Mendelian randomization pleiotropy residual sum and outlier (MR-PRESSO) test identifies possible bias from horizontal pleiotropy. The test consists of three parts, (1) the MR-PRESSO global test which detects horizontal pleiotropy, (2) the outlier corrected causal estimate which corrects for the detected horizontal pleiotropy and (3) the MR-PRESSO distortion test which estimates if the causal estimate is significantly different (at p<0.05) after adjustment for outliers. We conduct all three stages (with the argument NbDistribution=1000, namely using1000 simulation form the null distribution to compute empirical p-values) and present the outlier adjusted causal estimates (OACE) when both global and distortion tests are significant.

^b^ We did not run Steiger Test if none of the MR analysis resulted significant (NR: not reported in the cell).

^c^ Not enough SNP to perform MR (NR: not reported in the cell)

#### Supplementary Table 6: Odds Ratio of univariable forward Mendelian randomization analysis of poverty against mental illnesses

| **MR: method** | **OR (95% CI)** | **p-value** |
| --- | --- | --- |
| P → ADHD:  IVW  WM  MR-Egger | 3.43 (2.95; 3.99)  2.24 (1.70; 2.96)  0.327 (0.109; 0.981) | <0.001  <0.001  0.049 |
| P → AN:  IVW  WM  MR-Egger | 0.498 (0.400; 0.621)  0.500 (0.355; 0.709)  0.275 (0.065; 1.17) | <0.001  <0.001  0.085 |
| P → ANX:  IVW  WM  MR-Egger | 2.29 (1.46; 3.61)  1.81 (0.926; 3.52)  0.943 (0.090; 100) | <0.001  0.084  0.964 |
| P → ASD:  IVW  WM  MR-Egger | 0.980 (0.787; 1.23)  0.926 (0.641; 1.34)  0.167 (0.040; 0.704) | 0.873  0.695  0.017 |
| P → BD:  IVW  WM  MR-Egger | 0.971 (0.833; 1.12)  0.952 (0.730; 1.24)  0.483 (0.149; 1.56) | 0.678  0.724  0.229 |
| P → MDD:  IVW  WM  MR-Egger | 1.49 (1.29; 1.72)  1.35 (1.09; 1.67)  0.885 (0.382; 2.04) | <0.001  0.006  0.770 |
| P → OCD:  IVW  WM  MR-Egger | 0.508 (0.297; 0.870)  0.617 (0.277; 1.37)  1.68 (0.147; 18.9) | 0.014  0.236  0.678 |
| P → PTSD:  IVW  WM  MR-Egger | 1.88 (1.48; 2.37)  1.58 (1.10; 2.28)  0.592 (0.179; 1.95) | <0.001  0.013  0.390 |
| P → SZ:  IVW  WM  MR-Egger | 1.53 (1.35; 1.73)  1.58 (1.23; 2.03)  0.855 (0.236; 3.08) | <0.001  <0.001  0.808 |

Abbreviations: MR: Mendelian randomization; OR: Odds Ratio; 95% CI: 95% confidence intervals; P: poverty; ADHD: attention deficit hyperactivity disorder; AN: anorexia nervosa; ANX: anxiety disorder; ASD: autism spectrum disorders; BD: bipolar disorder; MDD: major depressive disorder; OCD: obsessive-compulsive disorder; PTSD: post-traumatic stress disorder; SZ: schizophrenia; IVW: inverse variance weighted (fixed effect); WM: weighted median.

Legend: Poverty is a latent variable built using household income as unit identification, therefore an increase in the indicator’s load stands for increased income, therefore the ORs have been reversed to facilitate interpretation of the effect of poverty.

#### Plots - Forward analyses

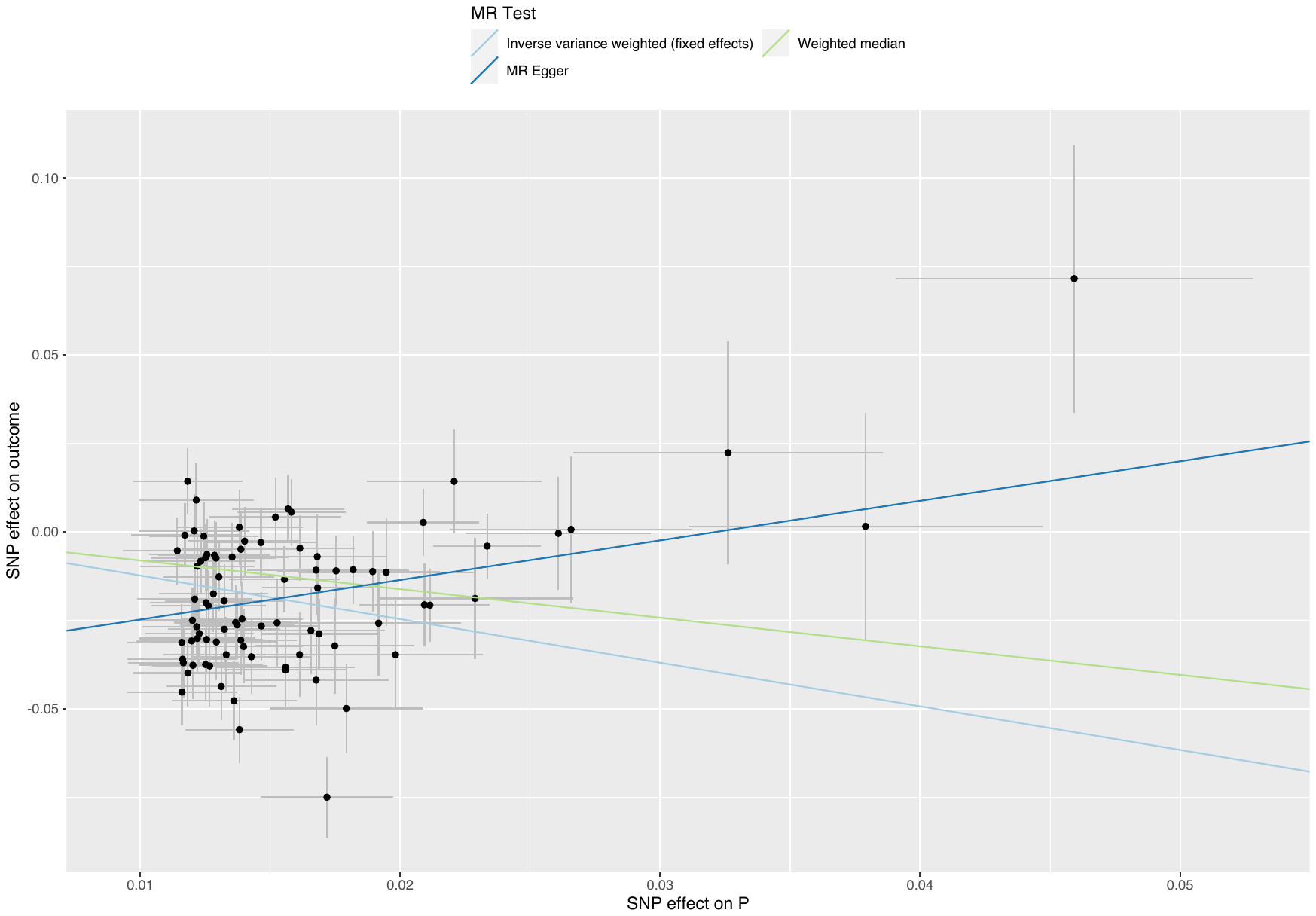

##### Supplementary Figure 5: scatterplot of poverty against ADHD

Abbreviations: MR: Mendelian randomization; SNP: single nucleotide polymorphism; P: poverty; ADHD: attention deficit hyperactivity disorder.

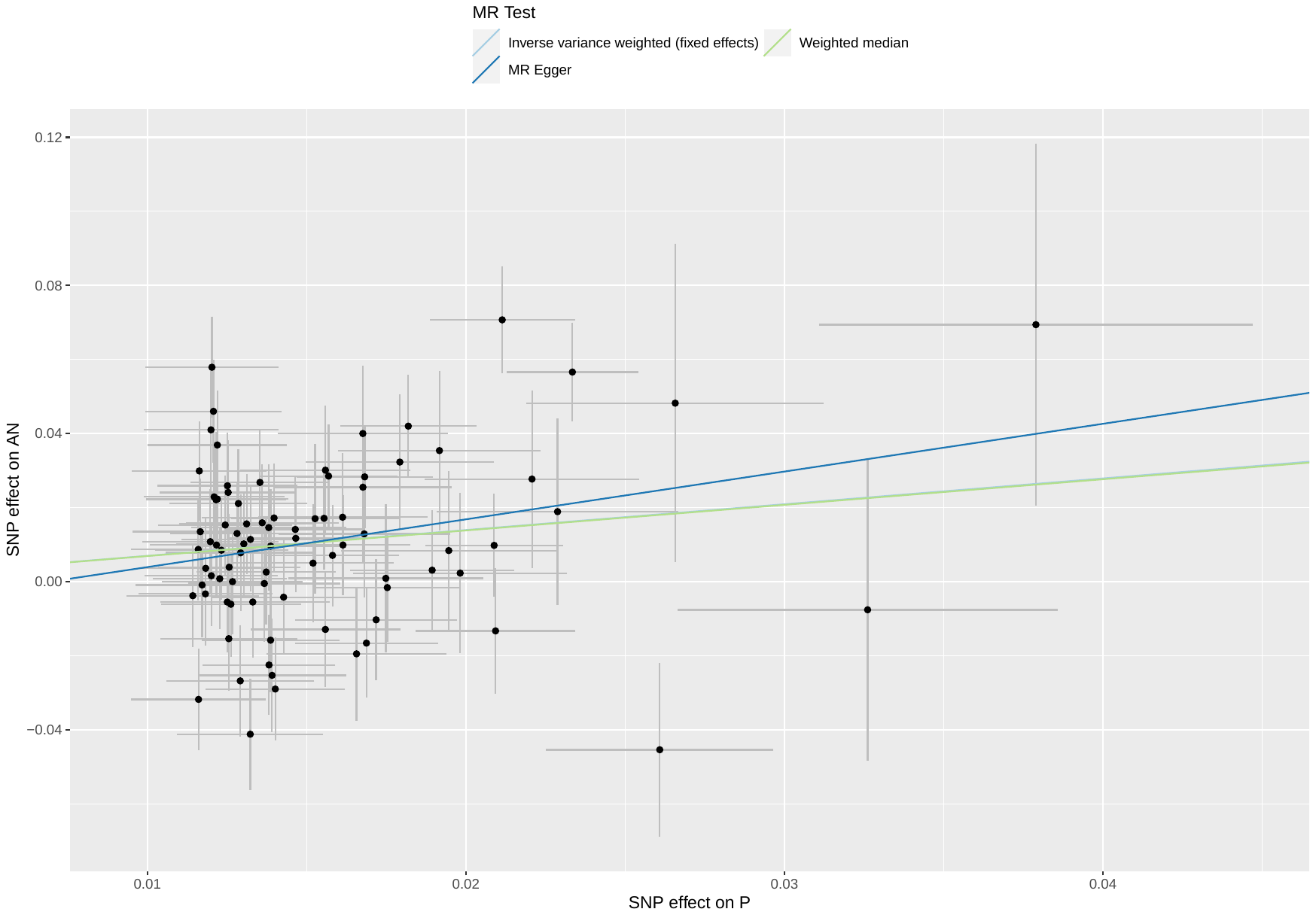

##### Supplementary Figure 6: scatterplot of poverty against AN

Abbreviations: MR: Mendelian randomization; SNP: single nucleotide polymorphism; P: poverty; AN: anorexia nervosa.

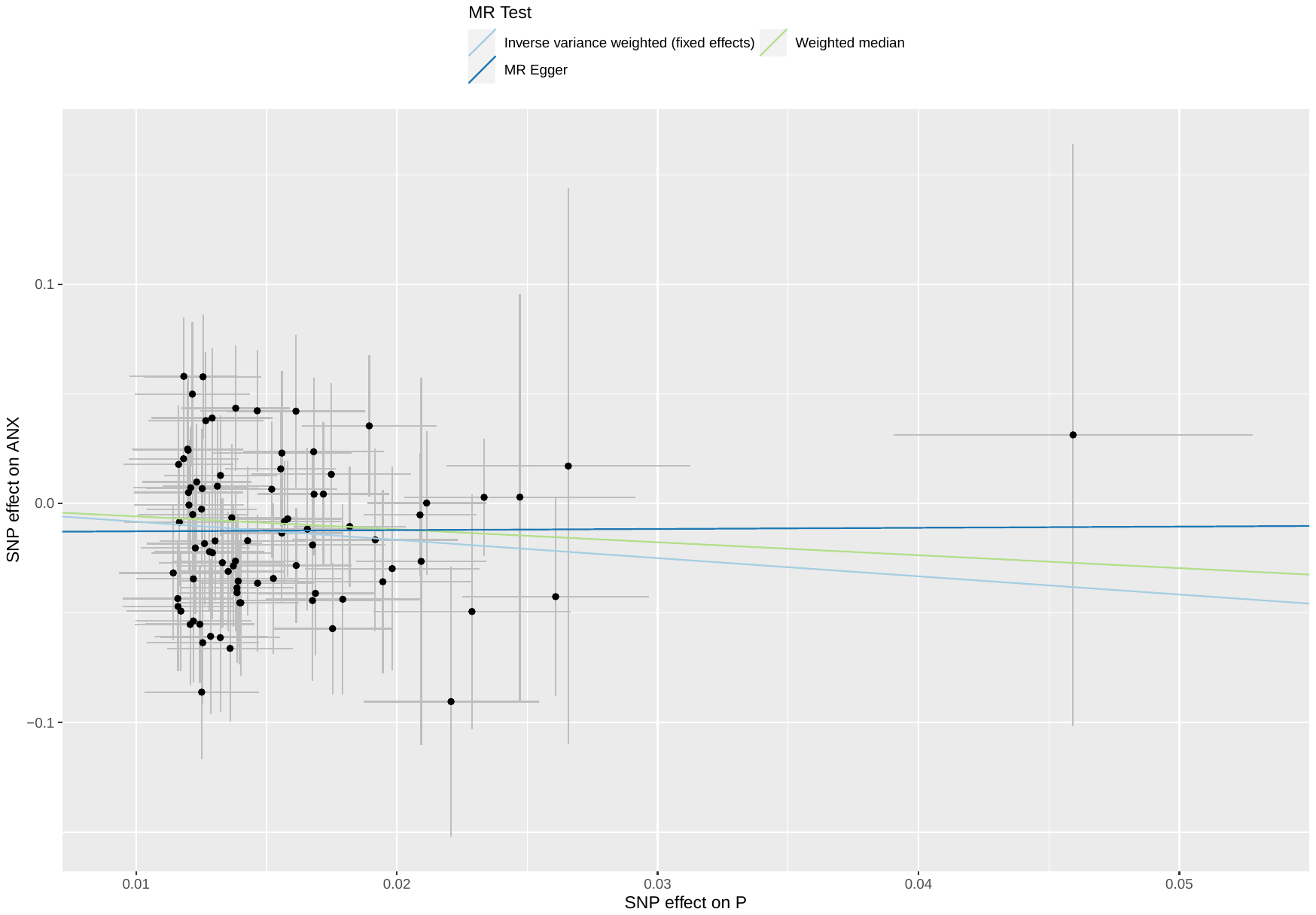

##### Supplementary Figure 7: scatterplot of poverty against ANX

Abbreviations: MR: Mendelian randomization; SNP: single nucleotide polymorphism; P: poverty; ANX: anxiety disorders.

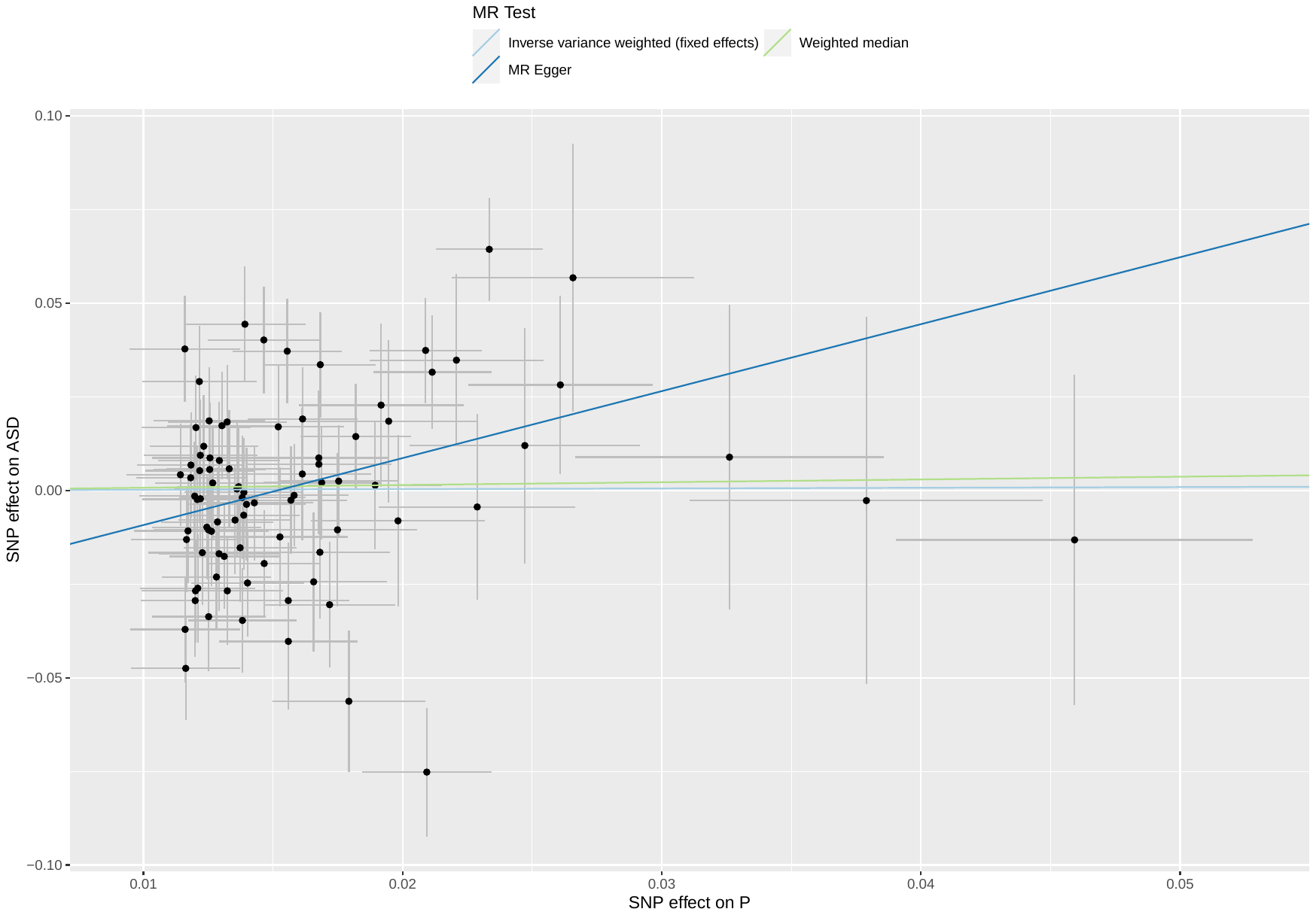

##### Supplementary Figure 8: scatterplot of poverty against ASD

Abbreviations: MR: Mendelian randomization; SNP: single nucleotide polymorphism; P: poverty; ASD: autism spectrum disorders.

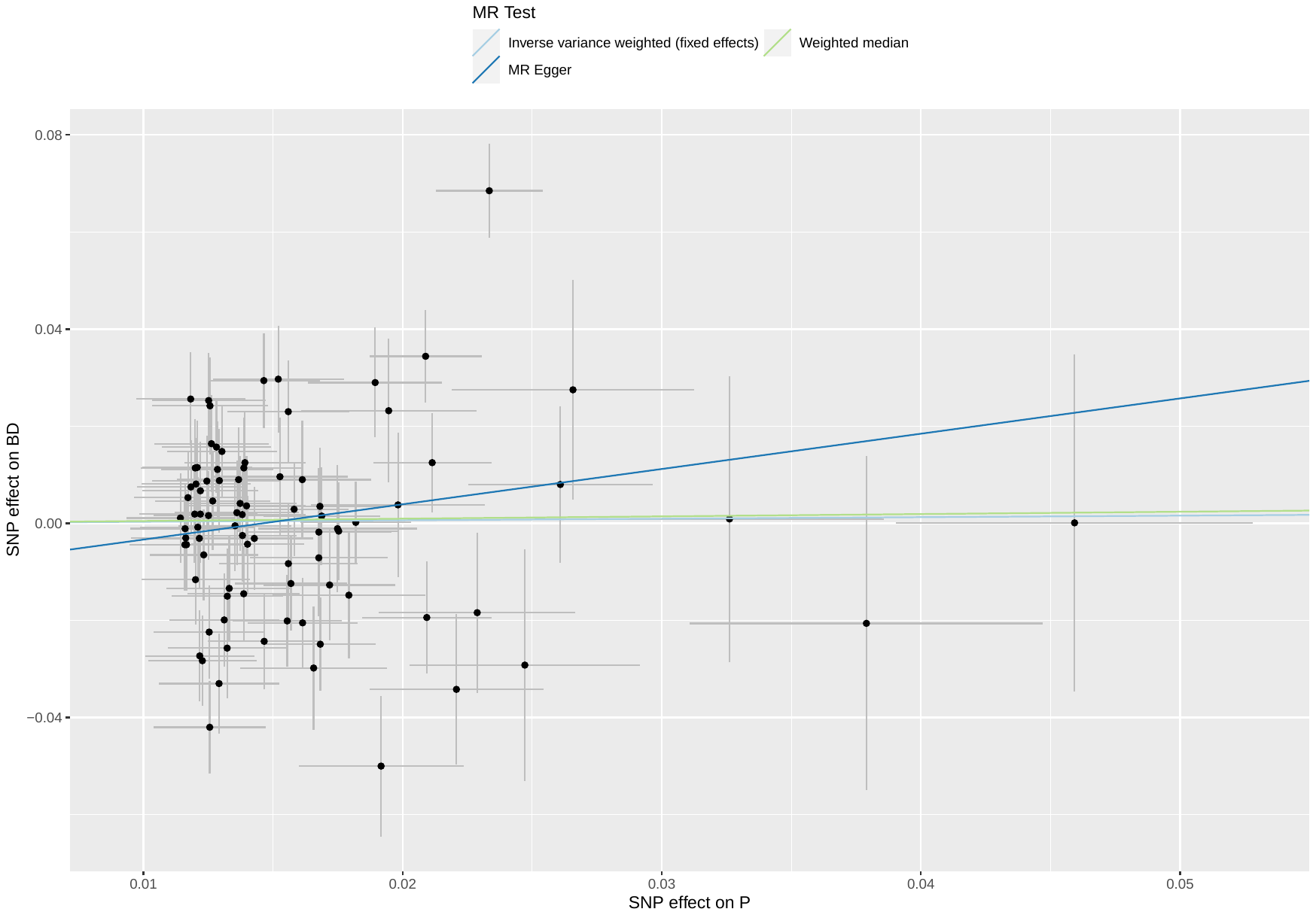

##### Supplementary Figure 9: scatterplot of poverty against BD

Abbreviations: MR: Mendelian randomization; SNP: single nucleotide polymorphism; P: poverty; BD: bipolar disorder.

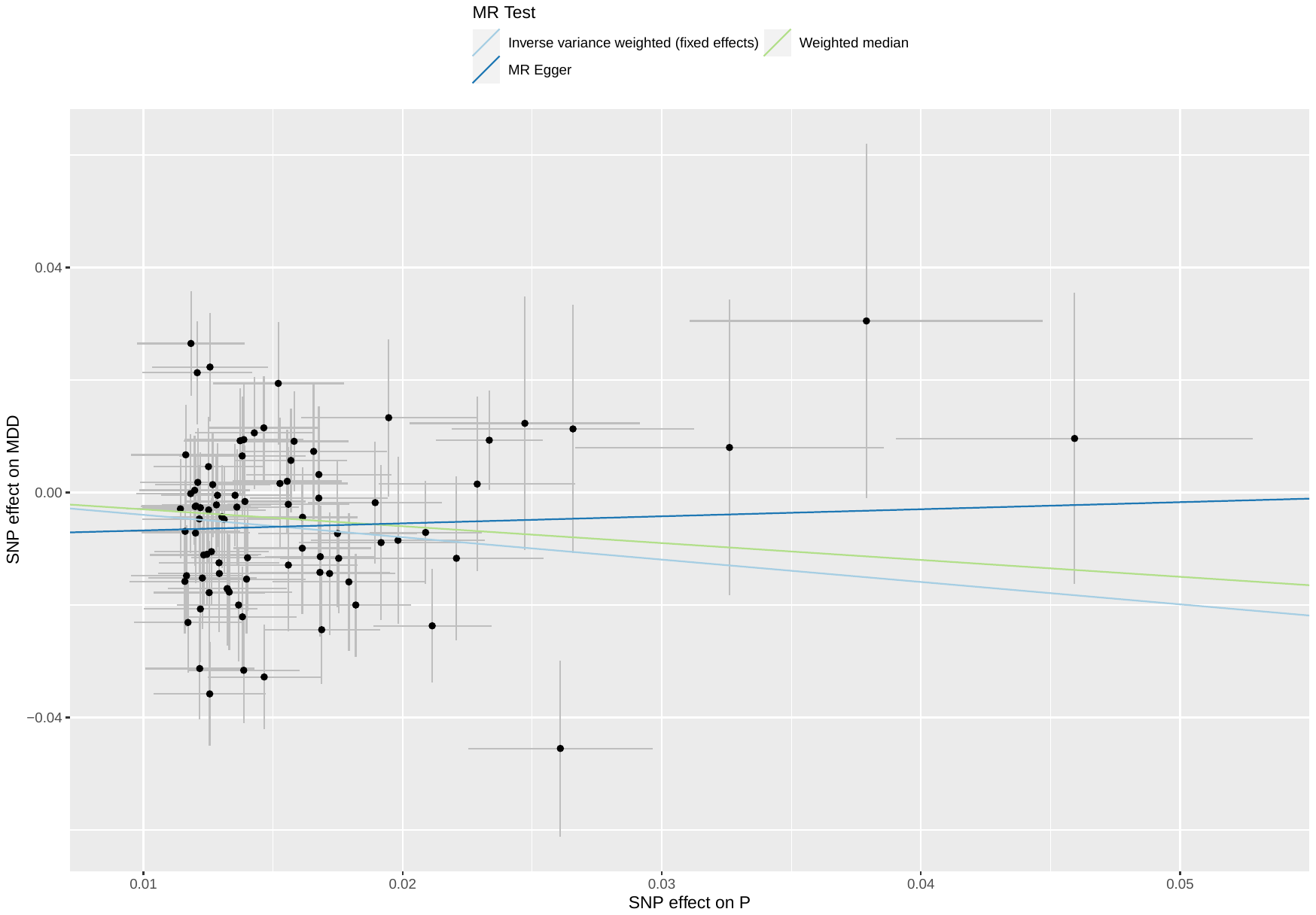

##### Supplementary Figure 10: scatterplot of poverty against MDD

Abbreviations: MR: Mendelian randomization; SNP: single nucleotide polymorphism; P: poverty; MDD: major depressive disorder.

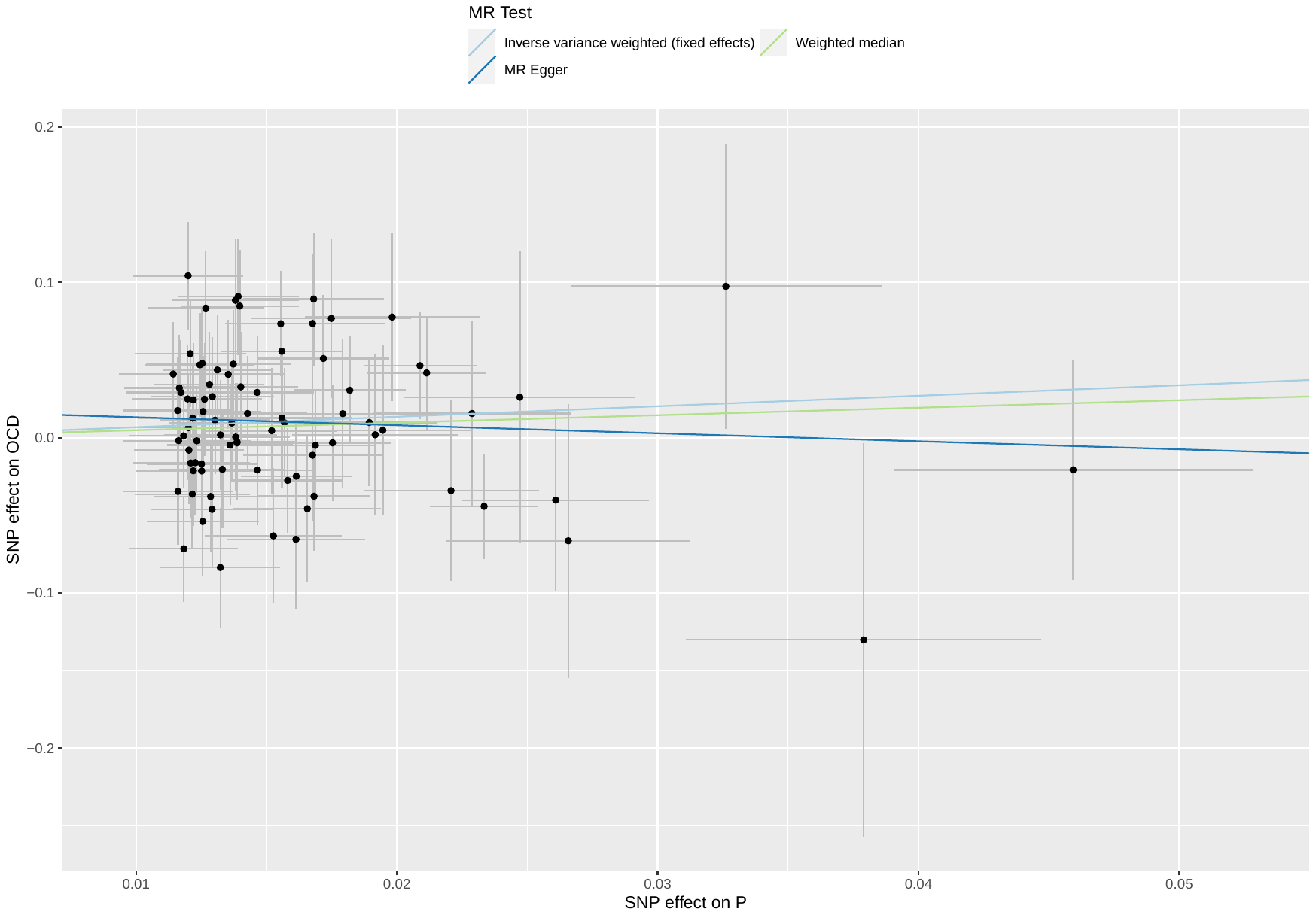

##### Supplementary Figure 11: scatterplot of poverty against OCD

Abbreviations: MR: Mendelian randomization; SNP: single nucleotide polymorphism; P: poverty; OCD: obsessive-compulsive disorder.

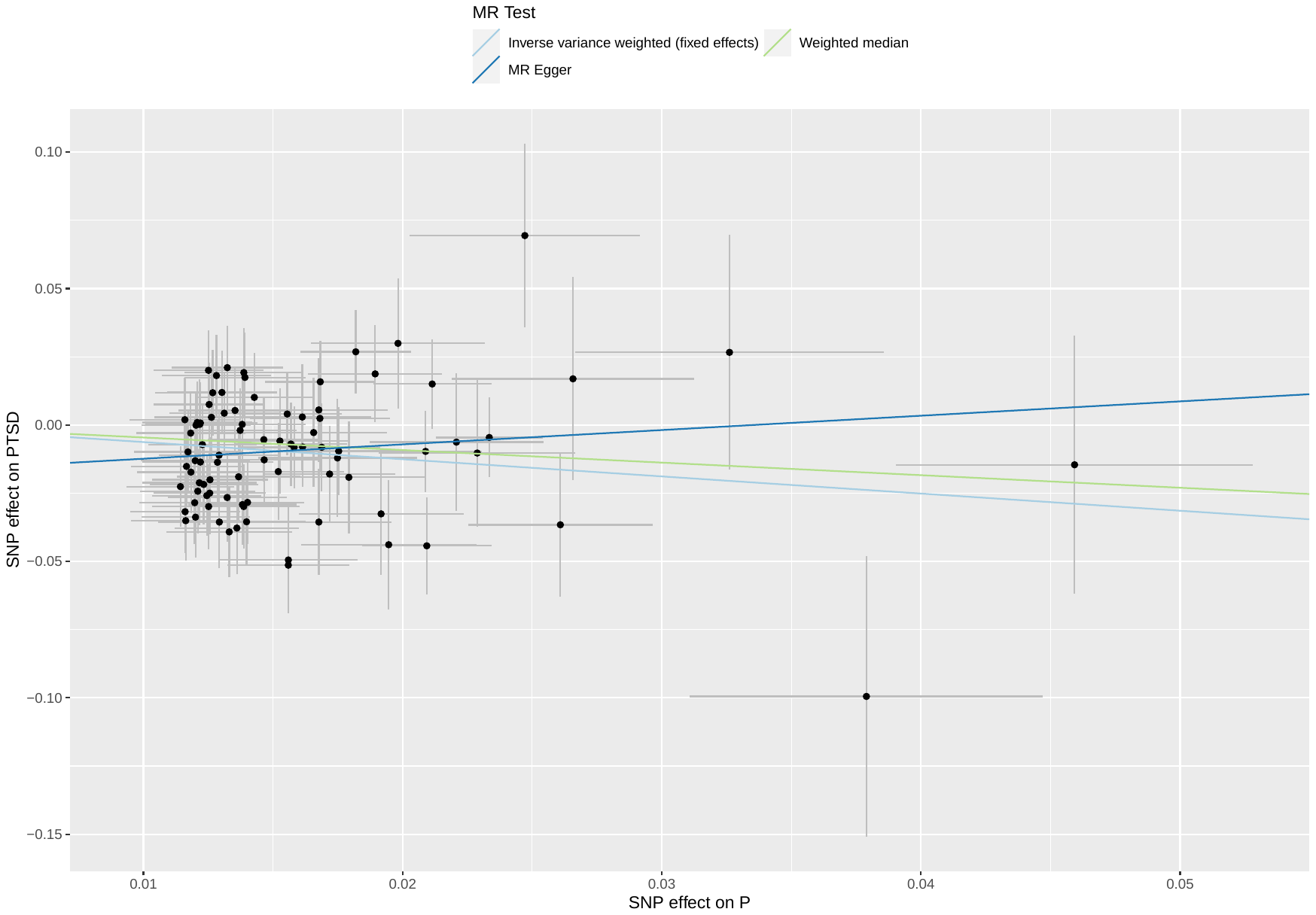

##### Supplementary Figure 12: scatterplot of poverty against PTSD

Abbreviations: MR: Mendelian randomization; SNP: single nucleotide polymorphism; P: poverty; PTSD: post-traumatic stress disorder.

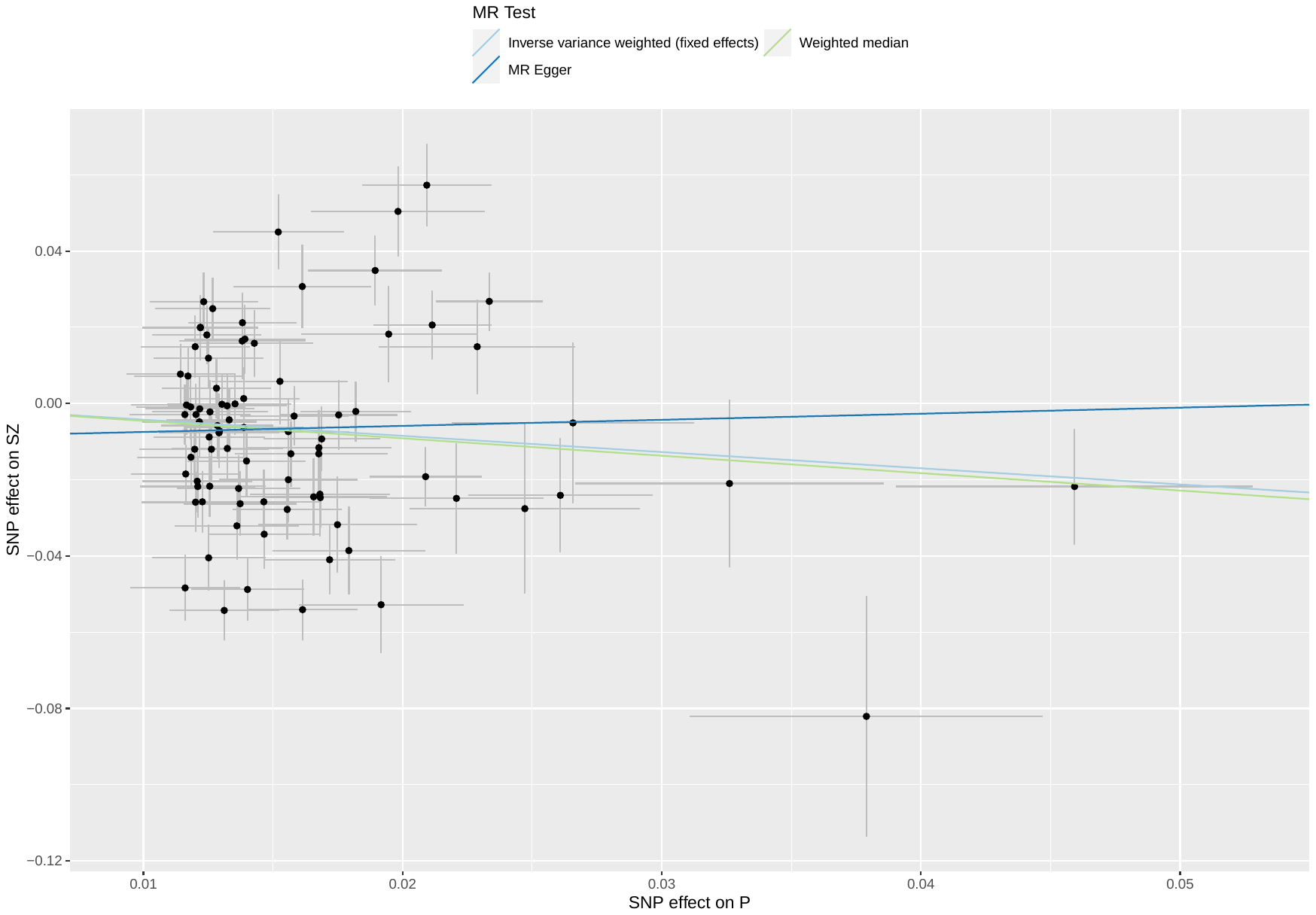

##### Supplementary Figure 13: scatterplot of poverty against SZ

Abbreviations: MR: Mendelian randomization; SNP: single nucleotide polymorphism; P: poverty; SZ: schizophrenia.

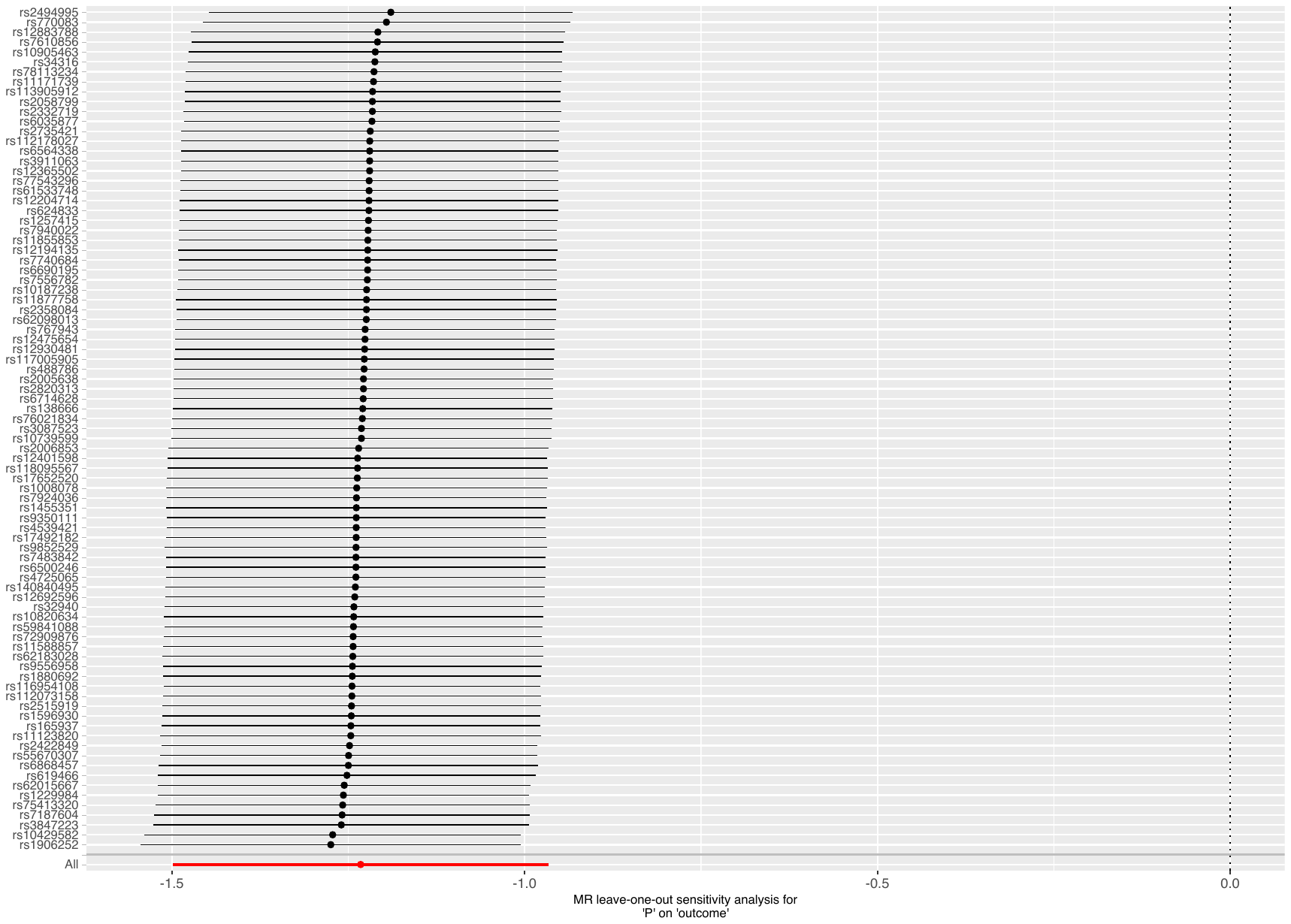

Supplementary Figure 14: leave-one-out analysis of poverty against ADHD

Abbreviations: MR: Mendelian randomization; P: poverty; ADHD: attention deficit hyperactivity disorder.

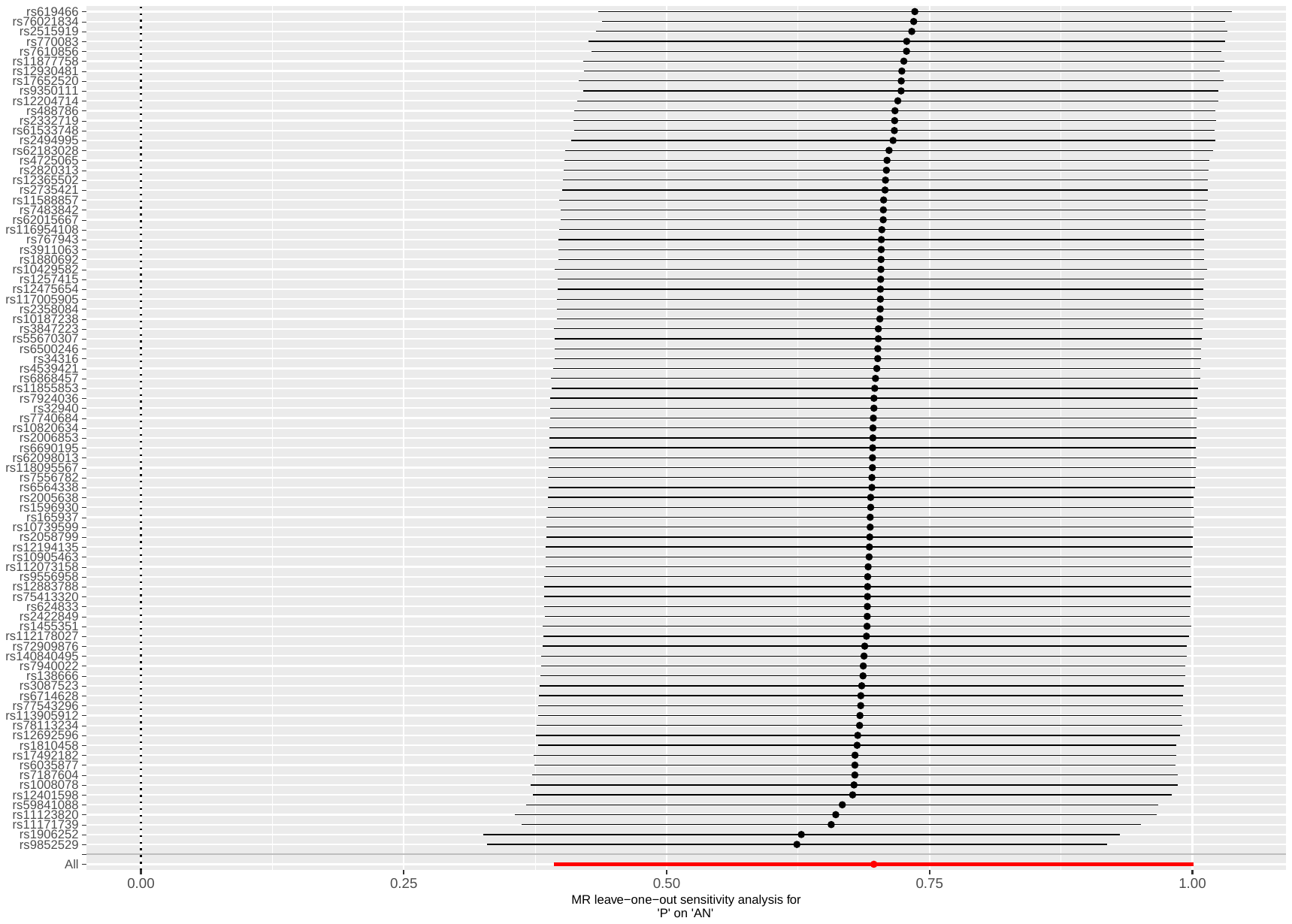

Supplementary Figure 15: leave-one-out analysis of poverty against AN

Abbreviations: MR: Mendelian randomization; P: poverty; AN: anorexia nervosa.

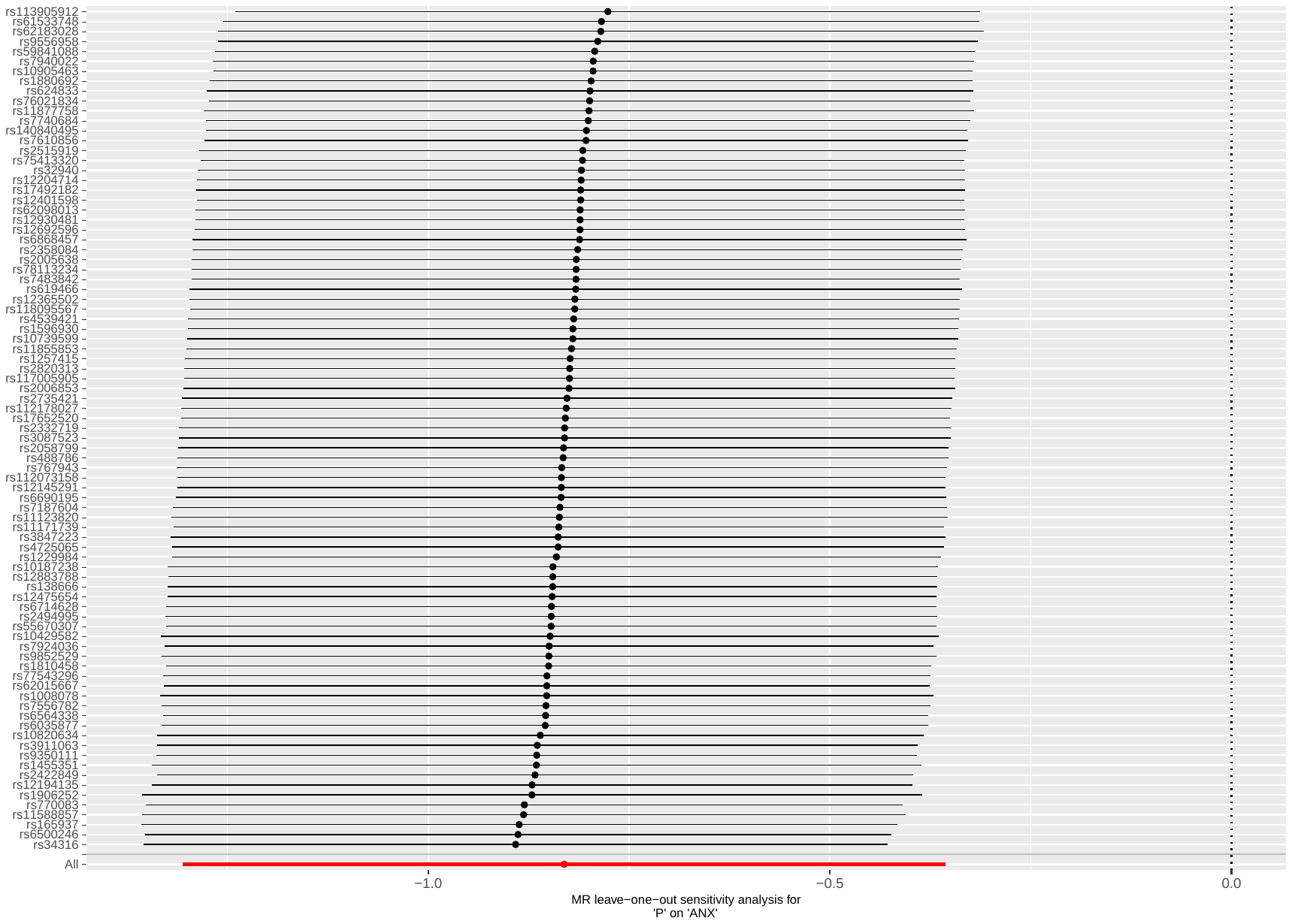

Supplementary Figure 16: leave-one-out analysis of poverty against ANX

Abbreviations: MR: Mendelian randomization; P: poverty; ANX: anxiety disorders.

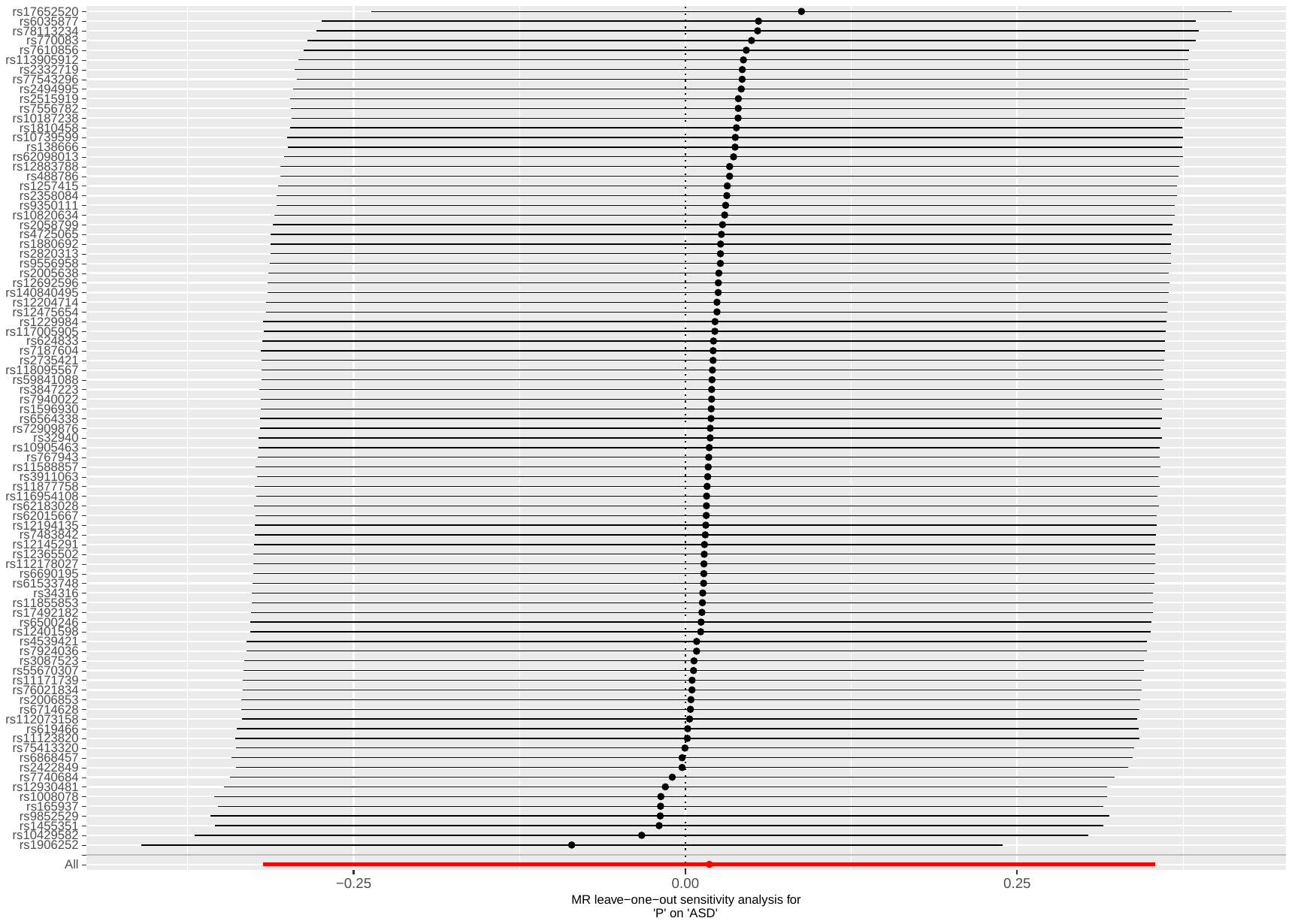

Supplementary Figure 17: leave-one-out analysis of poverty against ASD

Abbreviations: MR: Mendelian randomization; P: poverty; ASD: autism spectrum disorder.

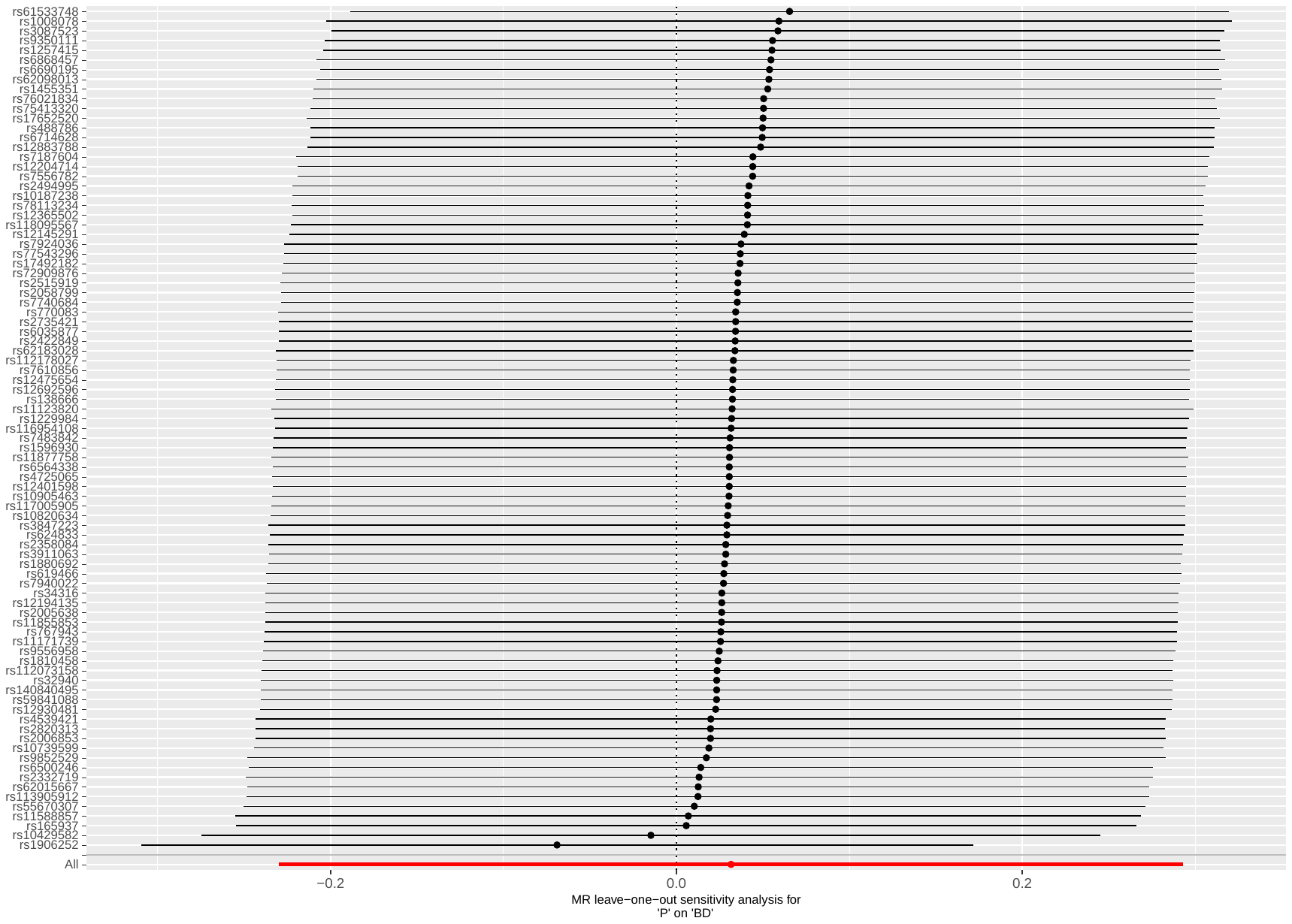

Supplementary Figure 18: leave-one-out analysis of poverty against BD

Abbreviations: MR: Mendelian randomization; P: poverty; BD: bipolar disorder.

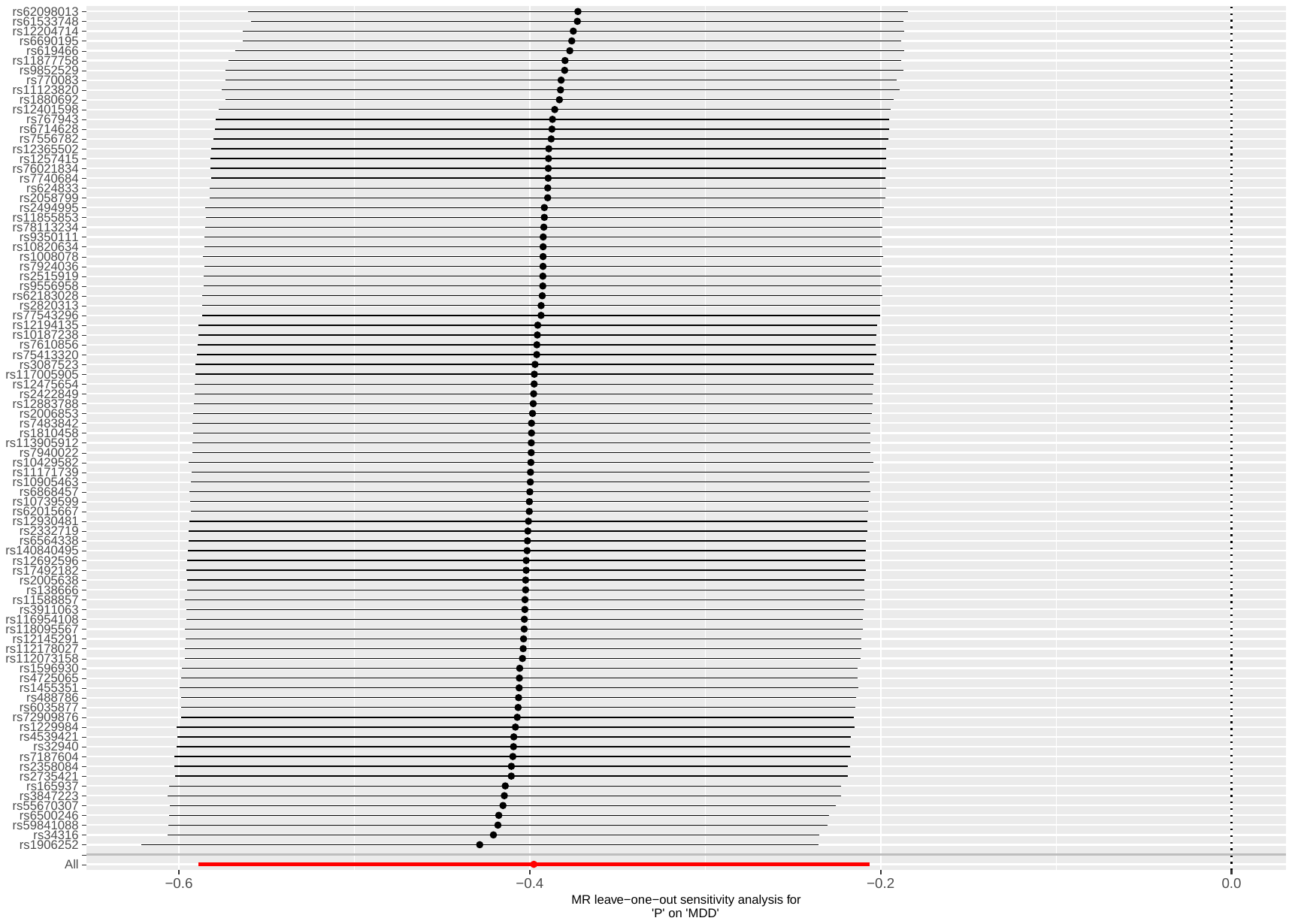

Supplementary Figure 19: leave-one-out analysis of poverty against MDD

Abbreviations: MR: Mendelian randomization; P: poverty; MDD: major depressive disorder.

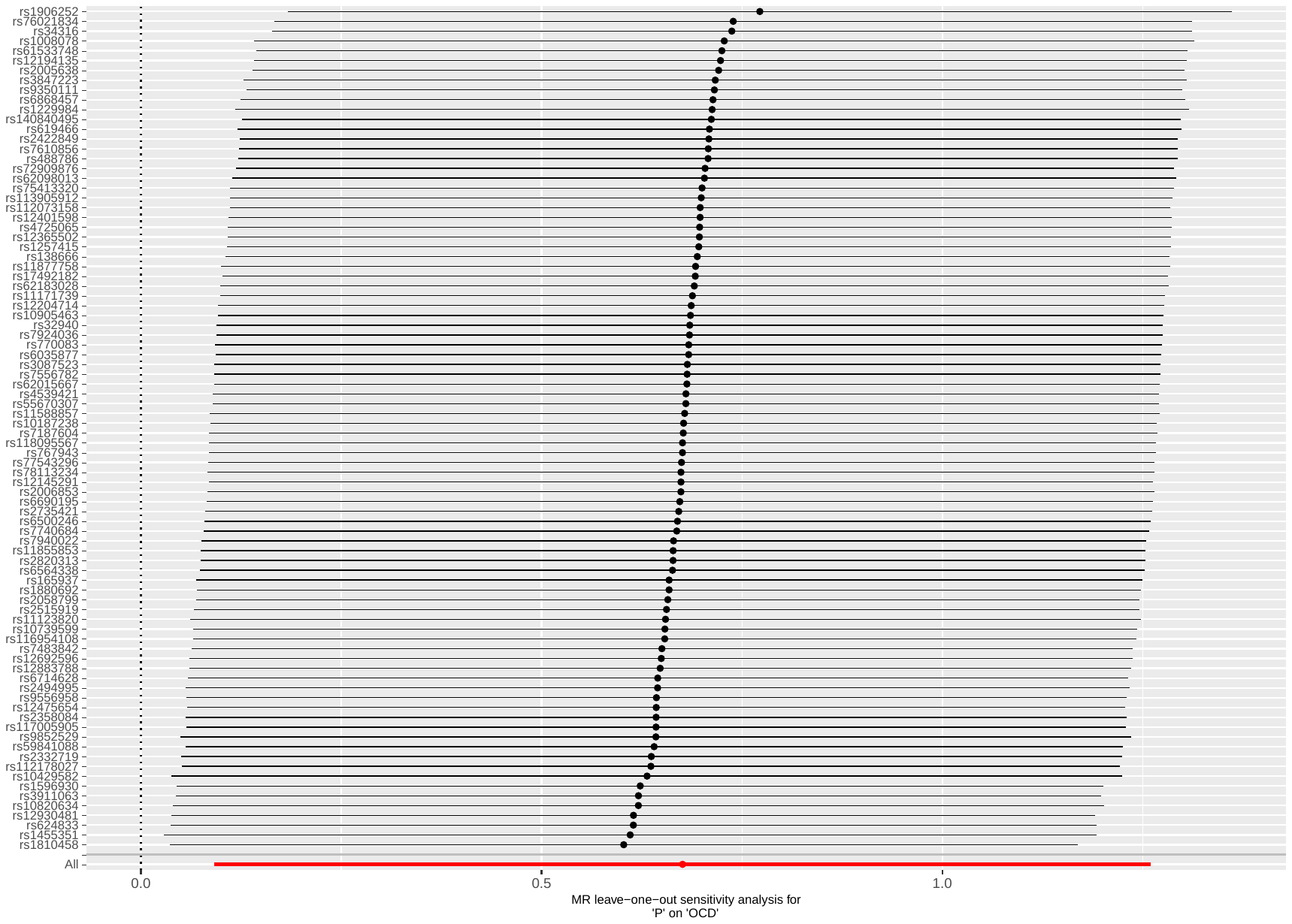

Supplementary Figure 20: leave-one-out analysis of poverty against OCD

Abbreviations: MR: Mendelian randomization; P: poverty; OCD: obsessive-compulsive disorder.

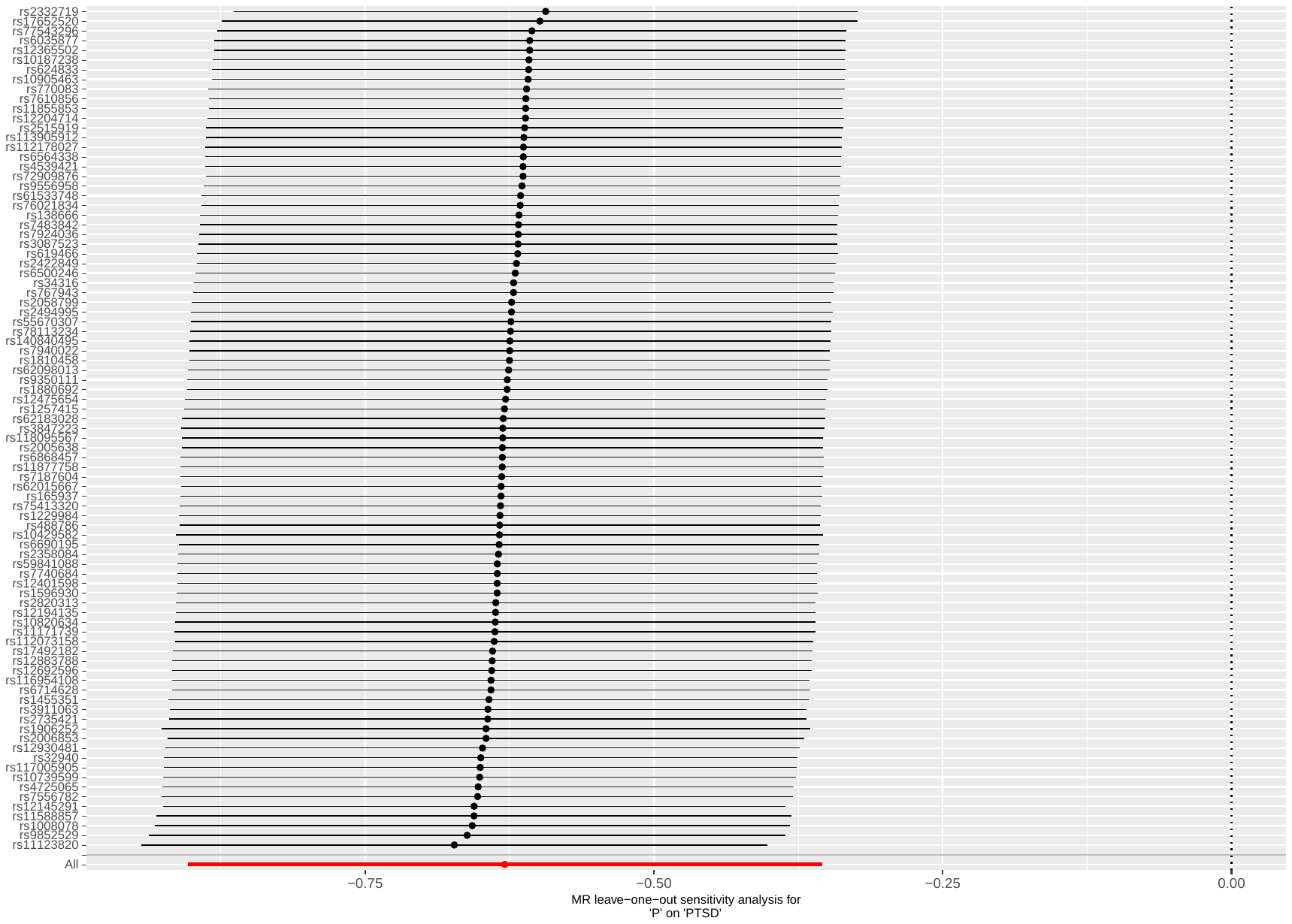

Supplementary Figure 21: leave-one-out analysis of poverty against PTSD

Abbreviations: MR: Mendelian randomization; P: poverty; PTSD: post-traumatic stress disorder.

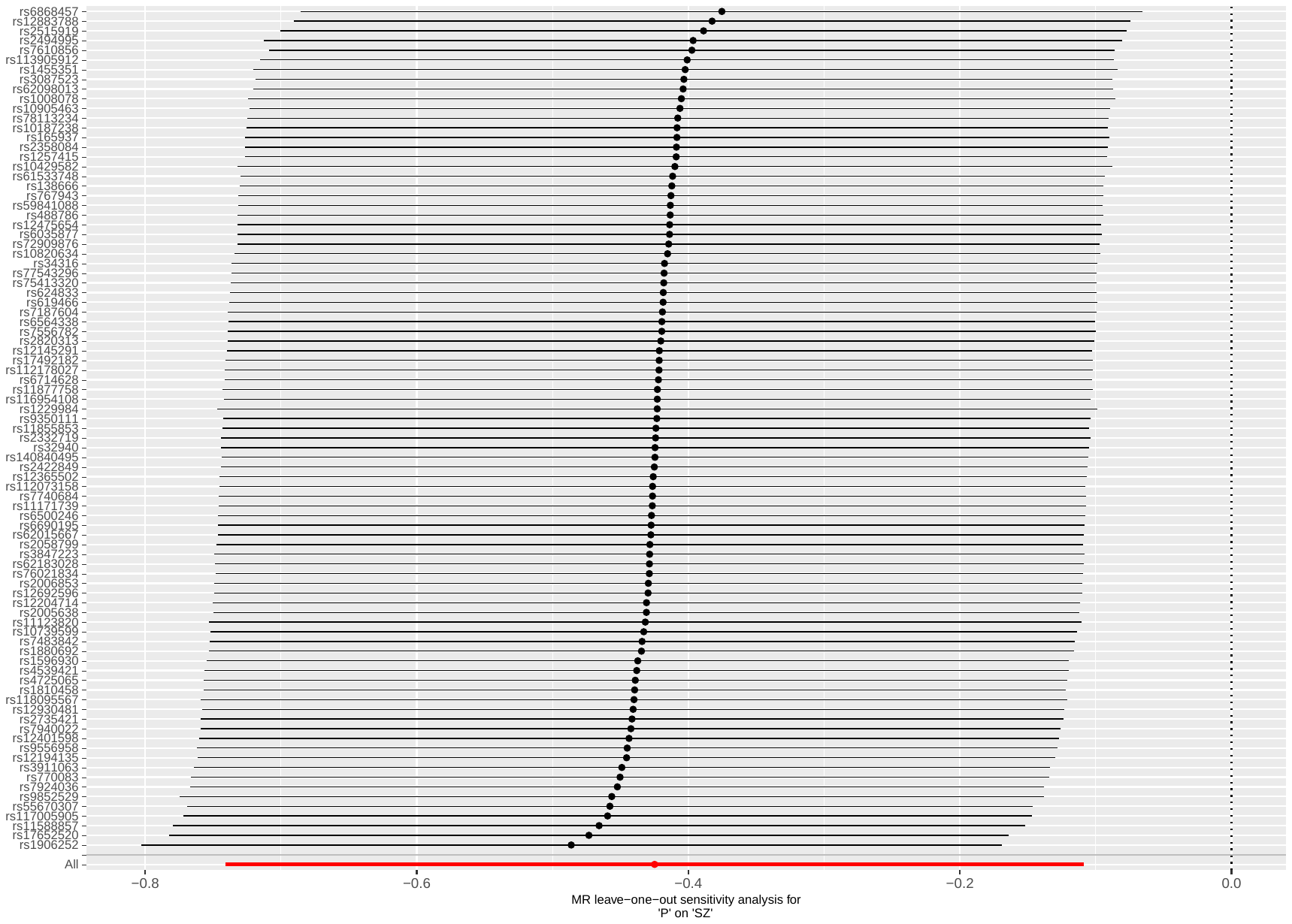

Supplementary Figure 22: leave-one-out analysis of poverty against SZ

Abbreviations: MR: Mendelian randomization; P: poverty; SZ: schizophrenia.

#### Plots - Backward analyses

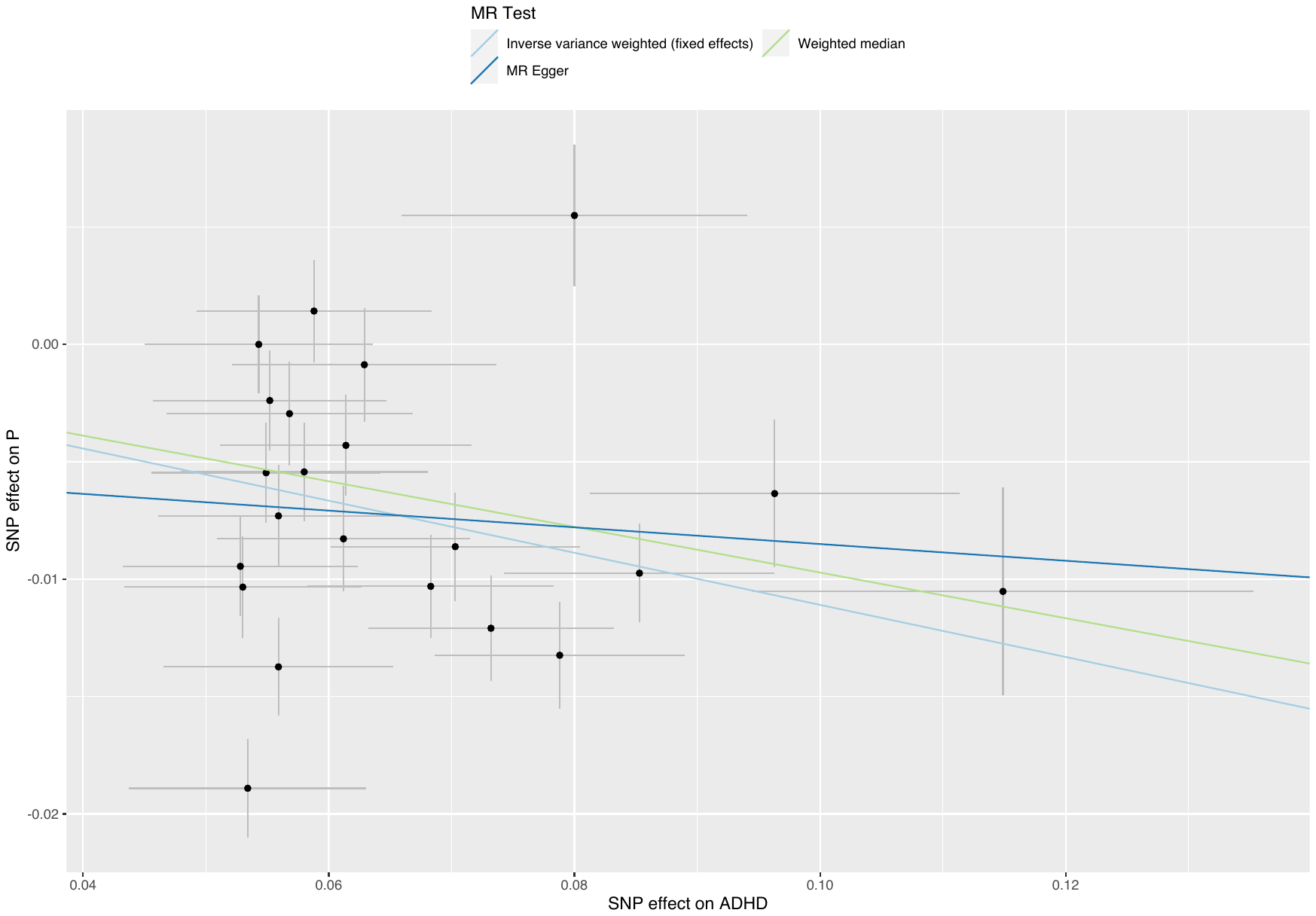

##### Supplementary Figure 23: scatterplot of ADHD against poverty

Abbreviations: MR: Mendelian randomization; SNP: single nucleotide polymorphism; P: poverty; ADHD: attention deficit hyperactivity disorder.

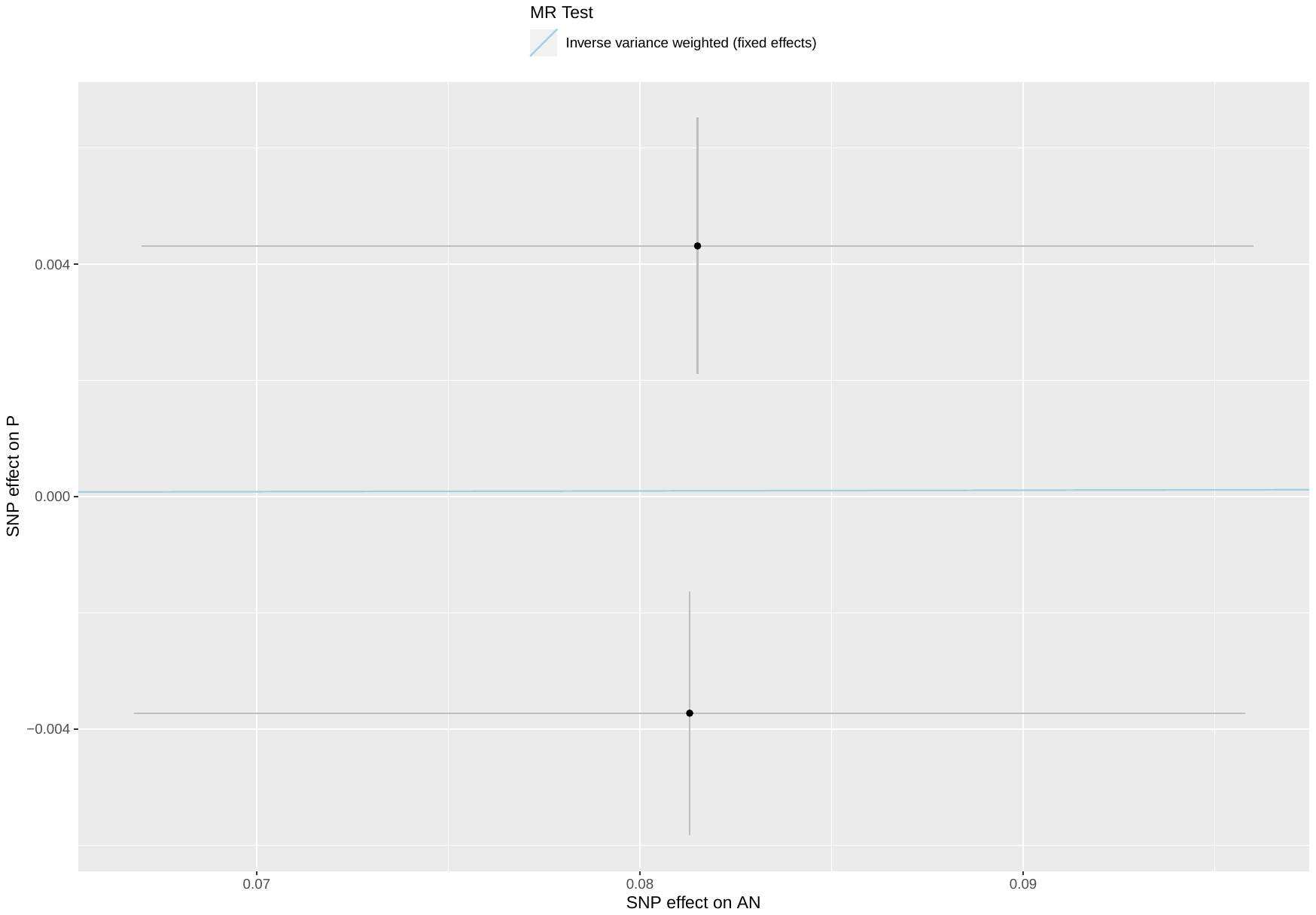

##### Supplementary Figure 24: scatterplot of AN against poverty

Abbreviations: MR: Mendelian randomization; SNP: single nucleotide polymorphism; P: poverty; AN: anorexia nervosa.

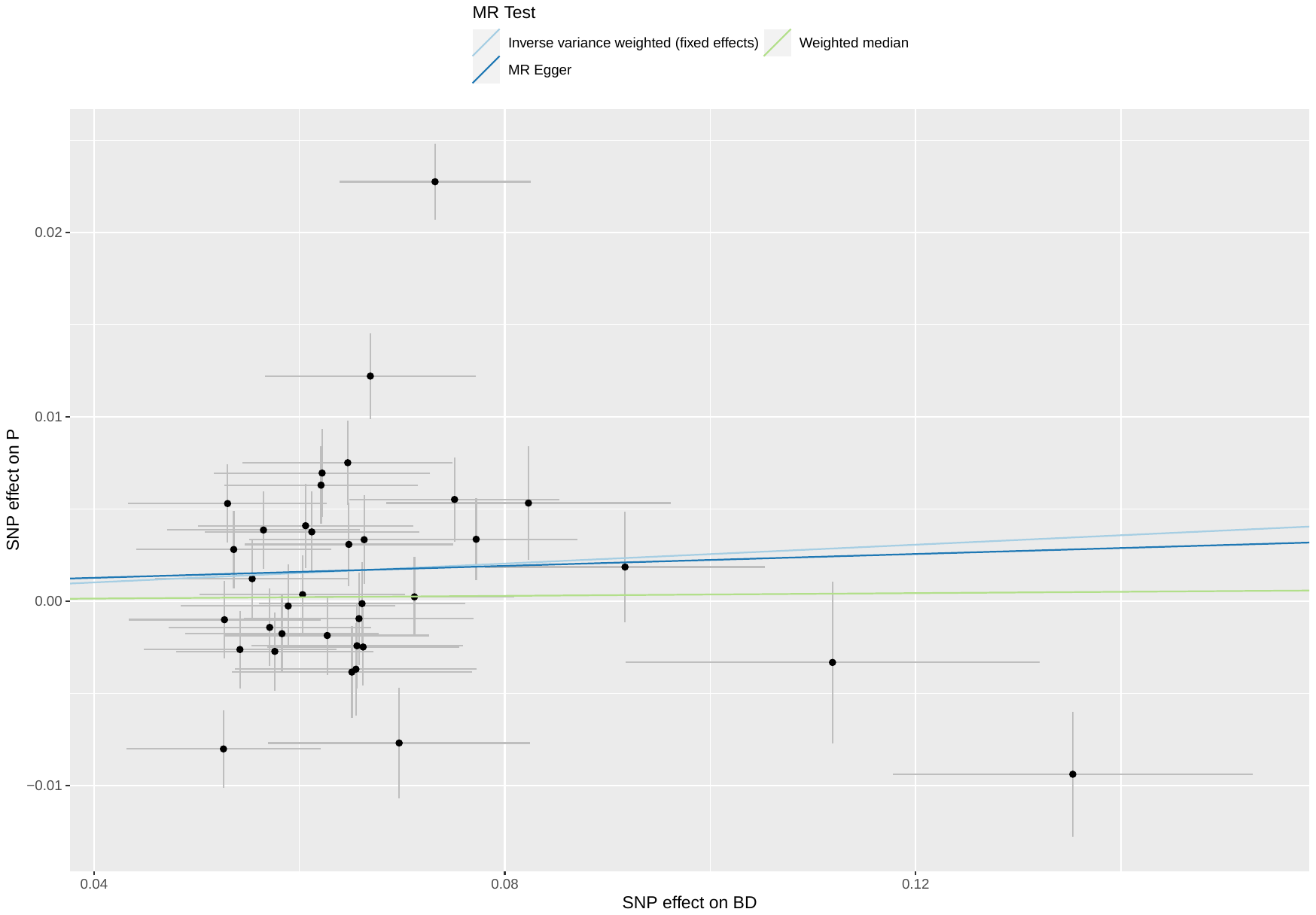

##### Supplementary Figure 25: scatterplot of BD against poverty

Abbreviations: MR: Mendelian randomization; SNP: single nucleotide polymorphism; P: poverty; BD: bipolar disorder.

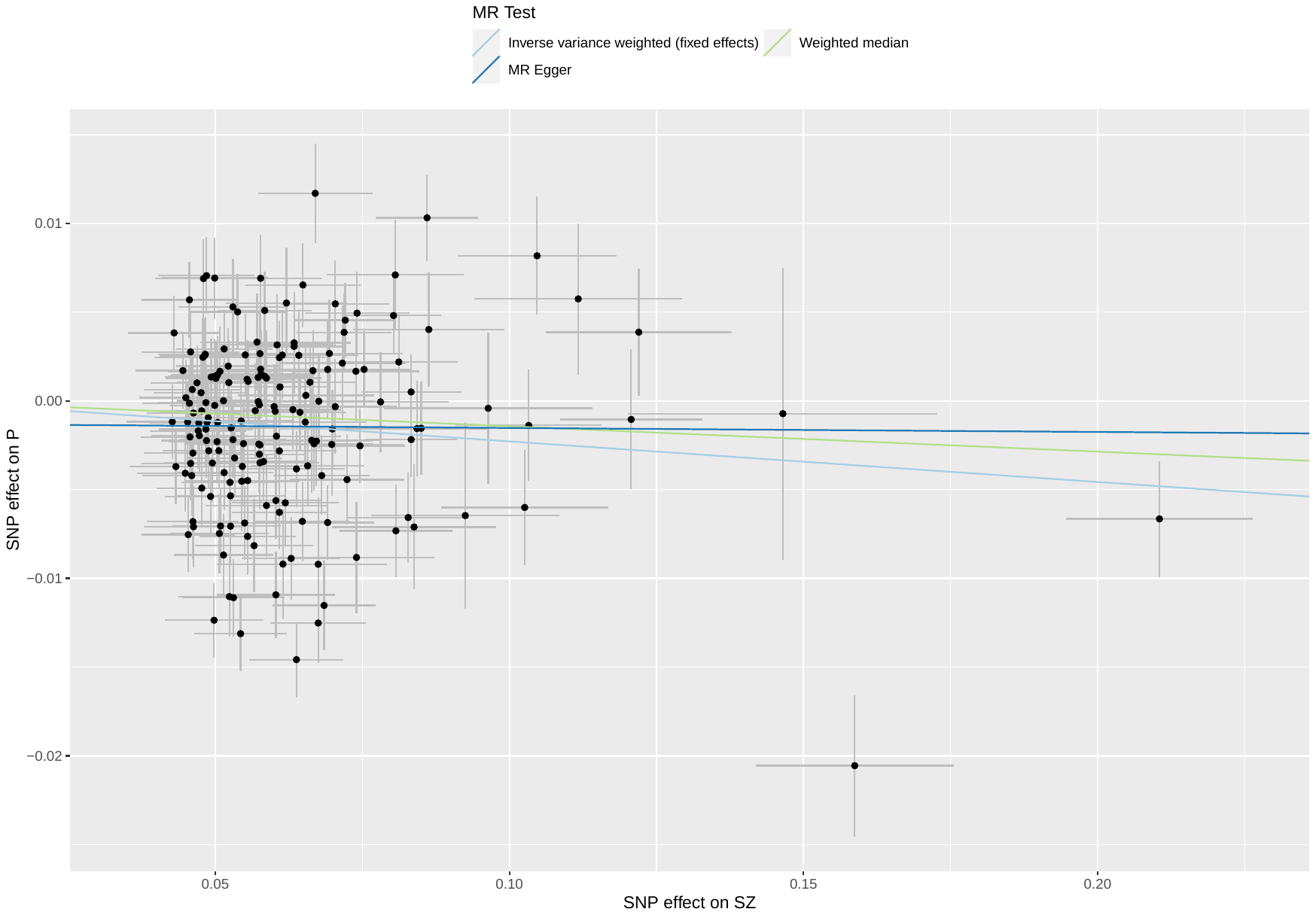

##### Supplementary Figure 26: scatterplot of SZ against poverty

Abbreviations: MR: Mendelian randomization; SNP: single nucleotide polymorphism; P: poverty; SZ: schizophrenia.

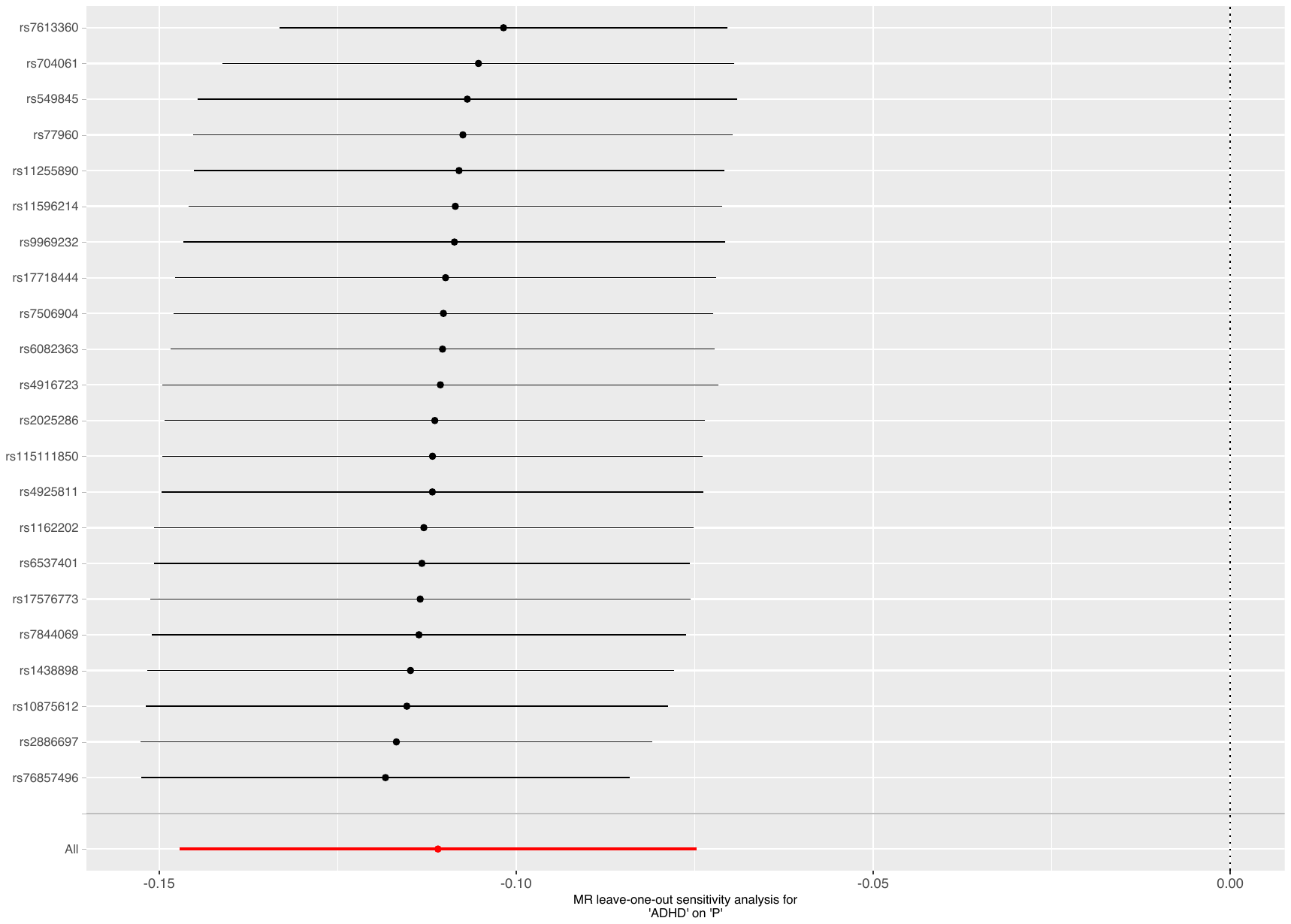

Supplementary Figure 27: leave-one-out analysis of ADHD against poverty

Abbreviations: MR: Mendelian randomization; P: poverty; ADHD: attention deficit hyperactivity disorder.

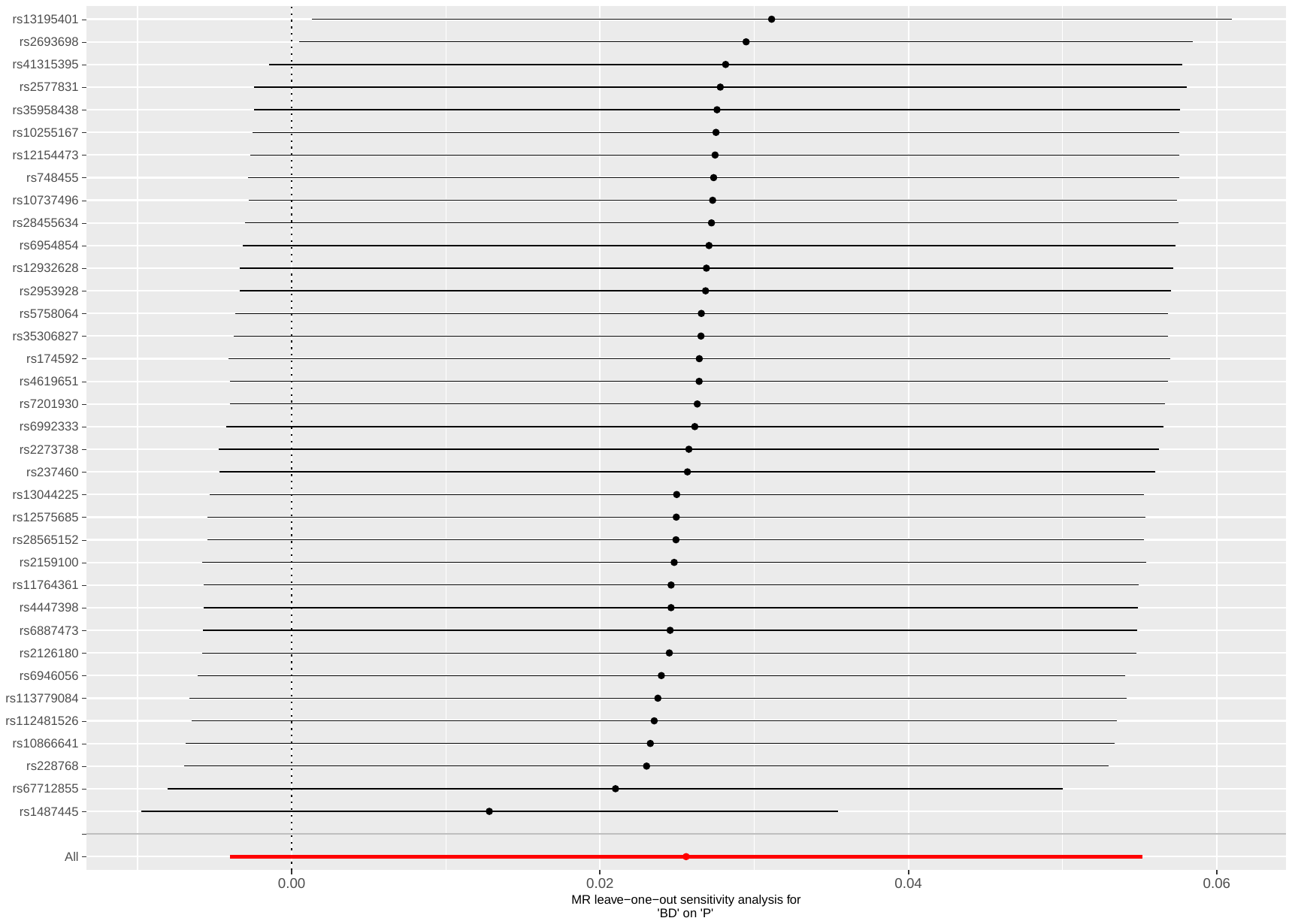

Supplementary Figure 28: leave-one-out analysis of BD against poverty

Abbreviations: MR: Mendelian randomization; P: poverty; BD: bipolar disorder.

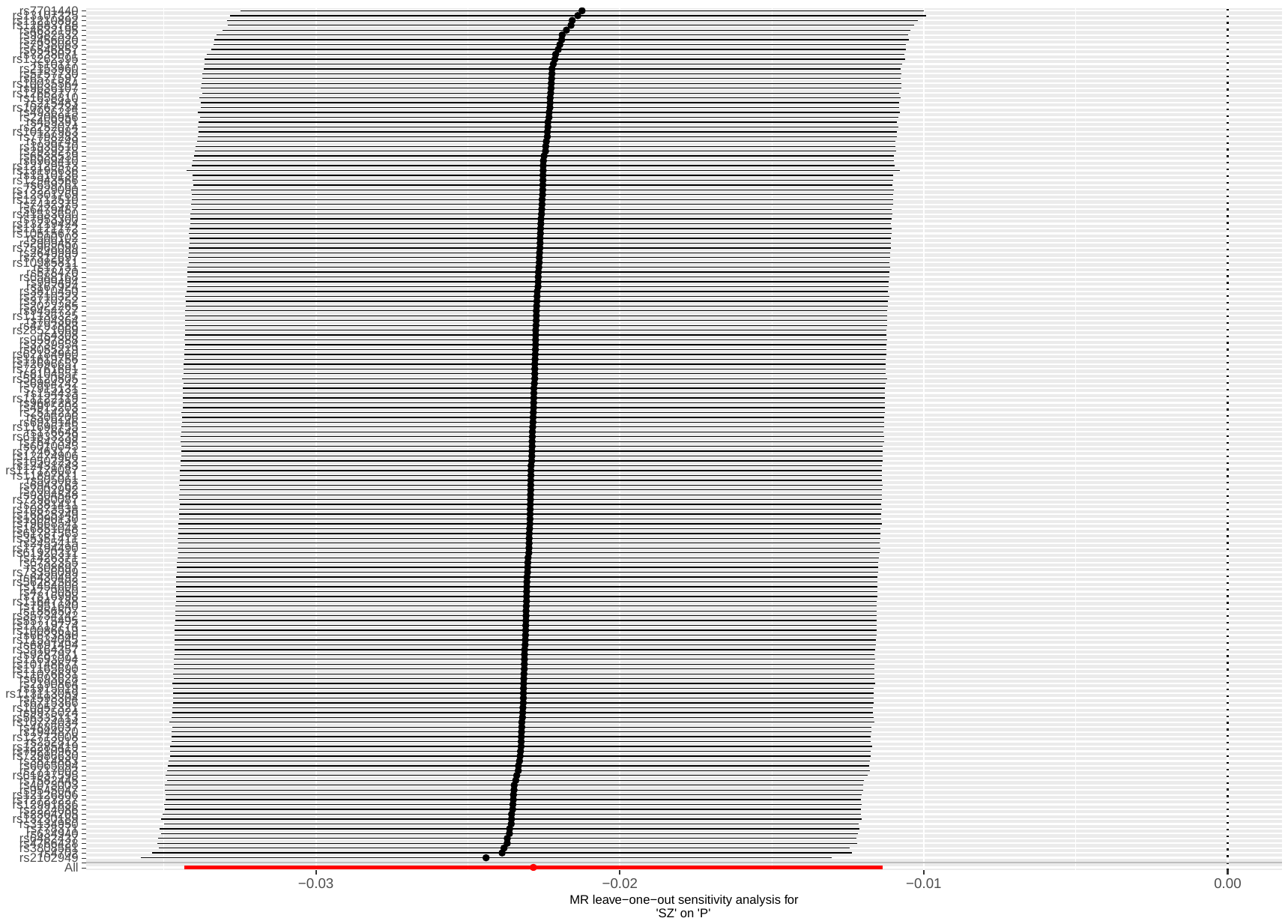

Supplementary Figure 29: leave-one-out analysis of SZ against poverty

Abbreviations: MR: Mendelian randomization; P: poverty; SZ: schizophrenia.

#### Supplementary Table 7: CAUSE results of the relations between poverty and mental illness

| **Model 1** | **Model 2** | **∆ ELPD** | **SE ∆ ELPD** | **z-score** | **p-value** |
| --- | --- | --- | --- | --- | --- |
| *Fw: P on ADHD* | | | | | |
| Null  Null  Sharing | Sharing  Causal  Causal | -67.54  -74.39  -6.85 | 9.89  10.94  1.34 | -6.83  -6.80  -5.12 | <0.001  <0.001  <0.001 |
| *Bw: ADHD on P* | | | | | |
| Null  Null  Sharing | Sharing  Causal  Causal | -4.26  -9.60  -5.34 | 1.78  3.77  2.01 | -2.40  -2.55  -2.65 | 0.016  0.011  0.008 |
| *Fw: P on AN* | | | | | |
| Null  Null  Sharing | Sharing  Causal  Causal | -9.37  -13.61  -4.24 | 3.49  5.00  1.67 | -2.69  -2.72  -2.54 | 0.007  0.006  0.011 |
| *Bw: AN on P* | | | | | |
| Null  Null  Sharing | Sharing  Causal  Causal | 0.39  0.60  0.21 | 0.15  0.98  0.84 | 2.62  0.61  0.25 | 0.009  0.540  0.803 |
| *Fw: P on ANX* | | | | | |
| Null  Null  Sharing | Sharing  Causal  Causal | -0.41  -0.64  -0.24 | 1.11  2.02  1.03 | -0.36  -0.32  -0.23 | 0.715  0.750  0.818 |
| *Bw: ANX on P* | | | | | |
| Null  Null  Sharing | Sharing  Causal  Causal | 0.17  0.81  0.65 | 0.04  0.22  0.19 | 4.09  3.70  3.41 | <0.001  <0.001  0.001 |
| *Fw: P on ASD* | | | | | |
| Null  Null  Sharing | Sharing  Causal  Causal | 0.52  1.38  0.86 | 0.07  0.11  0.03 | 7.04  12.99  25.23 | <0.001  <0.001  <0.001 |
| *Bw: ASD on P* | | | | | |
| Null  Null  Sharing | Sharing  Causal  Causal | 0.48  1.26  0.78 | 0.04  0.06  0.04 | 10.65  21.68  21.17 | <0.001  <0.001  <0.001 |
| *Fw: P on BD* | | | | | |
| Null  Null  Sharing | Sharing  Causal  Causal | 0.47  1.27  0.81 | 0.09  0.50  0.43 | 5.20  2.53  1.89 | <0.001  0.011  0.059 |
| *Bw: BD on P* | | | | | |
| Null  Null  Sharing | Sharing  Causal  Causal | 0.44  1.28  0.84 | 0.04  0.05  0.02 | 12.51  26.16  41.95 | <0.001  <0.001  <0.001 |
| *Fw: P on MDD* | | | | | |
| Null  Null  Sharing | Sharing  Causal  Causal | -9.90  -13.68  -3.78 | 3.72  5.09  1.51 | -2.66  -2.69  -2.50 | 0.008  0.007  0.013 |
| *Bw: MDD on P* | | | | | |
| Null  Null  Sharing | Sharing  Causal  Causal | 0.01  -1.16  -1.18 | 0.36  1.81  1.45 | 0.04  -0.64  -0.81 | 0.968  0.521  0.418 |
| *Fw: P on OCD* | | | | | |
| Null  Null  Sharing | Sharing  Causal  Causal | -0.36  -1.11  -0.75 | 0.95  2.19  1.29 | -0.38  -0.51  -0.59 | 0.707  0.612  0.558 |
| *Bw: OCD on P* | | | | | |
| Null  Null  Sharing | Sharing  Causal  Causal | 0.29  0.97  0.67 | 0.05  0.36  0.32 | 5.83  2.70  2.08 | <0.001  0.007  0.037 |
| *Fw: P on PTSD* | | | | | |
| Null  Null  Sharing | Sharing  Causal  Causal | -12.72  -16.75  -4.03 | 4.22  5.58  1.51 | -3.01  -3.00  -2.68 | 0.003  0.003  0.007 |
| *Bw: PTSD on P* | | | | | |
| Null  Null  Sharing | Sharing  Causal  Causal | 0.19  0.65  0.46 | 0.06  0.58  0.52 | 3.39  1.12  0.87 | 0.001  0.264  0.384 |
| *Fw: P on SZ* | | | | | |
| Null  Null  Sharing | Sharing  Causal  Causal | -1.08  -4.24  -3.16 | 0.96  2.87  1.92 | -1.12  -1.48  -1.65 | 0.261  0.140  0.100 |
| *Bw: SZ on P* | | | | | |
| Null  Null  Sharing | Sharing  Causal  Causal | -1.78  -5.03  -3.25 | 1.30  3.24  1.95 | -1.36  -1.55  -1.67 | 0.173  0.121  0.095 |

Abbreviations: MR: mendelian randomization; ELPD: expected log pointwise posterior density; 95%CI: 95% confidence interval; SE: standard error; Fwd: forward MR; Bwd: backward MR; P: latent factor poverty; ADHD: attention deficit hyperactivity disorder; AN: anorexia nervosa; ANX: anxiety disorder; ASD: autism spectrum disorder; BD: bipolar disorder; MDD: major depressive disorder; OCD: obsessive-compulsive disorder; PTSD: post-traumatic stress disorder; SZ: schizophrenia.

#### Supplementary Table 8: Results of univariable bidirectional Mendelian Randomization of poverty against mental illnesses, after Steiger filtering

| **MR** | **N SNP** | **IVW, B (95% CI)** | **p-value** | **WM, B (95% CI)** | **p-value** | **MR-Egger, B (95% CI)** | **p-value** | **Egger intercept p-value** | **Mean F** |
| --- | --- | --- | --- | --- | --- | --- | --- | --- | --- |
| P on ADHD | 79 | 1.05 (0.898; 1.21) | <0.001 | 0.661 (0.386; 0.937) | <0.001 | -0.976 (-1.94; 0.009) | 0.052 | <0.001 | 40.8 |
| P on AN | 77 | -0.558 (-0.789; -0.327) | <0.001 | -0.633 (-0.980; -0.285) | <0.001 | -1.20 (-2.44; 0.045) | 0.063 | 0.340 | 40.6 |
| P on ANX | 72 | 0.509 (0.025; 0.994) | 0.039 | 0.470 (-0.245; 1.19) | 0.198 | 1.25 (-1.14; 3.63) | 0.309 | 0.997 | 42.1 |
| P on ASD | 71 | -0.088 (-0.334; 0.159) | 0.485 | -0.075 (-0.432; 0.282) | 0.680 | -1.45 (-2.54; -0.350) | 0.012 | 0.010 | 39.6 |
| P on BD | 69 | 0.026 (-0.144; 0.197) | 0.761 | -0.020 (-0.283; 0.242) | 0.879 | 0.501 (-0.300; 1.30) | 0.224 | 0.074 | 39.1 |
| P on MDD | 84 | 0.351 (0.205; 0.496) | <0.001 | 0.272 (0.041; 0.504) | 0.021 | 0.024 (-0.774; 0.822) | 0.952 | 0.549 | 40.1 |
| P on OCD | 52 | -0.037 (-0.702; 0.628) | 0.913 | 0.033 (-0.891; 0.957) | 0.944 | 0.643 (-1.93; 3.22) | 0.627 | 0.581 | 43.2 |
| P on PTSD | 87 | 0.629 (0.395; 0.863) | <0.001 | 0.460 (0.088; 0.833) | 0.015 | -0.526 (-1.72; 0.667) | 0.390 | 0.020 | 40.3 |
| P on SZ | 83 | 0.264 (0.136; 0.392) | <0.001 | 0.446 (0.182; 0.710) | 0.001 | -0.023 (-1.18; 1.14) | 0.970 | 0.003 | 40.2 |

Abbreviations: P: poverty; ADHD: attention deficit hyperactivity disorder; AN: anorexia nervosa; ANX: anxiety disorder; ASD: autism spectrum disorders; BD: bipolar disorder; MDD: major depressive disorder; OCD: obsessive-compulsive disorder; PTSD: post-traumatic stress disorder; SZ: schizophrenia; MR: mendelian randomization; SNP: single nucleotide polymorphism; IVW: inverse variance weighted (fixed effect); B: effect estimates are log-odds; 95% CI: 95% confidence interval; WM: weighted median; NR: not reported because not enough SNP to perform MR.

Legend: Poverty is a latent variable built using household income as unit identification, therefore an increase in the indicator’s load stands for increased income, therefore the regression coefficients have been reversed to facilitate interpretation of the effect of poverty.

### **Univariable Mendelian randomization of household income and mental illnesses**

#### Supplementary Table 9: Odds Ratio of univariable forward Mendelian randomization analysis of household income against mental illnesses

| **MR: method** | **OR (95% CI)** | **p-value** |
| --- | --- | --- |
| HI → ADHD:  IVW  WM  MR-Egger | 0.436 (0.363; 0.524)  0.571 (0.422; 0.774)  1.83 (0.504; 6.67) | <0.001  <0.001  0.362 |
| HI → AN:  IVW  WM  MR-Egger | 1.56 (1.21; 2.02)  1.45 (0.978; 2.14)  2.69 (0.492; 14.7) | 0.001  0.078  0.259 |
| HI → ANX:  IVW  WM  MR-Egger | 0.418 (0.243; 0.718)  0.608 (0.271; 1.37)  0.913 (0.050; 16.7) | 0.002  0.229  0.951 |
| HI → ASD:  IVW  WM  MR-Egger | 0.933 (0.729; 1.19)  0.910 (0.600; 1.38)  2.23 (0.428; 11.6) | 0.583  0.654  0.346 |
| HI → BD:  IVW  WM  MR-Egger | 1.15 (0.964; 1.38)  1.07 (0.790; 1.44)  3.71 (0.707; 19.5) | 0.120  0.685  0.128 |
| HI → MDD:  IVW  WM  MR-Egger | 0.656 (0.555; 0.775)  0.743 (0.572; 0.966)  0.647 (0.274; 1.53) | <0.001  0.027  0.325 |
| HI → OCD:  IVW  WM  MR-Egger | 1.52 (0.814; 2.85)  1.35 (0.553; 3.30)  1.08 (0.089; 13.2) | 0.189  0.528  0.951 |
| HI → PTSD:  IVW  WM  MR-Egger | 0.652 (0.500; 0.851)  0.736 (0.497; 1.09)  0.728 (0.205; 2.59) | 0.002  0.135  0.626 |
| HI → SZ:  IVW  WM  MR-Egger | 0.660 (0.569; 0.767)  0.649 (0.487; 0.865)  0.864 (0.171; 4.38) | <0.001  0.005  0.861 |

Abbreviations: MR: Mendelian randomization; OR: Odds Ratio; 95% CI: 95% confidence intervals; HI: household income; ADHD: attention deficit hyperactivity disorder; AN: anorexia nervosa; ANX: anxiety disorder; ASD: autism spectrum disorders; BD: bipolar disorder; MDD: major depressive disorder; OCD: obsessive-compulsive disorder; PTSD: post-traumatic stress disorder; SZ: schizophrenia; IVW: inverse variance weighted (fixed effect); WM: weighted median; NR: not reported because not enough SNP to perform MR.

#### Supplementary Table 10: Results of univariable bidirectional Mendelian Randomization of household income against mental illnesses

| **MR** | **N SNP** | **IVW, B (95% CI)** | **p-value** | **IVW Q(df)** | **Q p-value** | **WM, B (95% CI)** | **p-value** | **MR-Egger, B (95% CI)** | **p-value** | **Egger intercept p-value** | **Steiger Test p-value** | **MR-PRESSO** | **Mean F** |
| --- | --- | --- | --- | --- | --- | --- | --- | --- | --- | --- | --- | --- | --- |
| Fw: HI on ADHD | 46 | -0.830 (-1.01; -0.647) | <0.001 | 125 (45) | <0.001 | -0.560 (-0.863; -0.256) | <0.001 | 0.606 (-0.684; 1.90) | 0.362 | 0.030 | <0.001 | DT; p=0.215 | 36.7 |
| Bw: ADHD on HI | 23 | -0.103 (-0.120; -0.086) | <0.001 | 75.2 (22) | <0.001 | -0.079 (-0.108; -0.049) | <0.001 | 0.019 (-0.159; 0.197) | 0.836 | 0.185 | <0.001 | DT; p=0.572 | 39.2 |
| Fw: HI on AN | 50 | 0.448 (0.191; 0.704) | 0.001 | 100 (49) | <0.001 | 0.370 (-0.041; 0.781) | 0.078 | 0.990 (-0.710; 2.69) | 0.259 | 0.524 | <0.001 | DT; p=0.149 | 37.2 |
| Bw: AN on HI | 4 | -0.012 (-0.044; 0.019) | 0.444 | 9 (3) | 0.027 | -0.005 (-0.043; 0.034) | 0.817 | 0.268 (-0.187; 0.722) | 0.367 | 0.348 | NR ^b^ | GT; p=0.093 | 31.9 |
| Fw: HI on ANX | 48 | -0.872 (-1.41; -0.331) | 0.002 | 48 (47) | 0.434 | -0.497 (-1.31; 0.312) | 0.229 | -0.091 (-3.00; 2.82) | 0.951 | 0.594 | <0.001 | GT; p=0.427 | 37.3 |
| Bw: ANX on HI | 0 | NR ^c^ | NR ^c^ | NR ^c^ | NR ^c^ | NR ^c^ | NR ^c^ | NR ^c^ | NR ^c^ | NR ^c^ | NR ^c^ | NR ^c^ | NR ^c^ |
| Fw: HI on ASD | 54 | -0.069 (-0.316; 0.178) | 0.583 | 134 (53) | <0.001 | -0.095 (-0.508; 0.319) | 0.654 | 0.801 (-0.850; 2.45) | 0.346 | 0.292 | NR ^b^ | DT; p=0.909 | 37.5 |
| Bw: ASD on HI | 0 | NR ^c^ | NR ^c^ | NR ^c^ | NR ^c^ | NR ^c^ | NR ^c^ | NR ^c^ | NR ^c^ | NR ^c^ | NR ^c^ | NR ^c^ | NR ^c^ |
| Fw: HI on BD | 47 | 0.143 (-0.037; 0.322) | 0.120 | 207 (46) | <0.001 | 0.063 (-0.242; 0.368) | 0.685 | 1.31 (-0.347; 2.97) | 0.128 | 0.163 | NR ^b^ | DT; p=0.093 | 37.2 |
| Bw: BD on HI | 36 | 0.016 (0.002; 0.030) | 0.020 | 189 (35) | <0.001 | 0.018 (-0.005; 0.042) | 0.122 | -0.015 (-0.188; 0.158) | 0.869 | 0.726 | <0.001 | DT; p=0.098 | 39.2 |
| Fw: HI on MDD | 50 | -0.422 (-0.589; -0.255) | <0.001 | 72 (49) | 0.017 | -0.297 (-0.561; -0.032) | 0.027 | -0.435 (-1.29; 0.423) | 0.325 | 0.975 | <0.001 | DT; p=0.725 | 36.9 |
| Bw: MDD on HI | 0 | NR ^c^ | NR ^c^ | NR ^c^ | NR ^c^ | NR ^c^ | NR ^c^ | NR ^c^ | NR ^c^ | NR ^c^ | NR ^c^ | NR ^c^ | NR ^c^ |
| Fw: HI on OCD | 50 | 0.420 (-0.206; 1.05) | 0.189 | 48 (49) | 0.499 | 0.301 (-0.633; 1.23) | 0.528 | 0.079 (-2.42; 2.58) | 0.951 | 0.784 | NR ^b^ | GT; p=0.514 | 36.9 |
| Bw: OCD on HI | 0 | NR ^c^ | NR ^c^ | NR ^c^ | NR ^c^ | NR ^c^ | NR ^c^ | NR ^c^ | NR ^c^ | NR ^c^ | NR ^c^ | NR ^c^ | NR ^c^ |
| Fw: HI on PTSD | 54 | -0.427 (-0.693; -0.161) | 0.002 | 66 (53) | 0.113 | -0.306 (-0.708; 0.096) | 0.135 | -0.317 (-1.59; 0.952) | 0.626 | 0.861 | <0.001 | GT; p=0.112 | 37.5 |
| Bw: PTSD on HI | 0 | NR ^c^ | NR ^c^ | NR ^c^ | NR ^c^ | NR ^c^ | NR ^c^ | NR ^c^ | NR ^c^ | NR ^c^ | NR ^c^ | NR ^c^ | NR ^c^ |
| Fw: HI on SZ | 47 | -0.415 (-0.565; -0.265) | <0.001 | 338 (46) | <0.001 | -0.432 (-0.733; -0.130) | 0.005 | -0.146 (-1.77; 1.48) | 0.861 | 0.739 | <0.001 | DT; p=0.431 | 37.2 |
| Bw: SZ on HI | 176 | -0.031 (-0.037; -0.024) | <0.001 | 603 (175) | <0.001 | -0.025 (-0.037; -0.012) | 0.001 | -0.018 (-0.067; 0.031) | 0.477 | 0.597 | <0.001 | DT; p=0.240 | 45.6 |

Abbreviations: Fw: forward analysis; Bw: backward analysis; HI: household income; ADHD: attention deficit hyperactivity disorder; AN: anorexia nervosa; ANX: anxiety disorder; ASD: autism spectrum disorders; BD: bipolar disorder; MDD: major depressive disorder; OCD: obsessive-compulsive disorder; PTSD: post-traumatic stress disorder; SZ: schizophrenia; MR: mendelian randomization; SNP: single nucleotide polymorphism; IVW: inverse variance weighted (fixed effect); B: effect estimates are log-odds for binary traits (i.e., for mental illnesses) and unstandardized regression coefficient for continuous traits (i.e., for household income); 95% CI: 95% confidence interval; Q: Cochran’s Q measure of heterogeneity; df: degree of freedom; WM: weighted median; DT: distortion test; GT: global test.

Legend:

P-value threshold <5e-8

^a^ The Mendelian randomization pleiotropy residual sum and outlier (MR-PRESSO) test identifies possible bias from horizontal pleiotropy. The test consists of three parts. (1) the MR-PRESSO global test which detects horizontal pleiotropy. (2) the outlier corrected causal estimate which corrects for the detected horizontal pleiotropy and (3) the MR-PRESSO distortion test which estimates if the causal estimate is significantly different (at p<0.05) after adjustment for outliers. We conduct all three stages (with the argument NbDistribution=1000. namely using1000 simulation form the null distribution to compute empirical p-values) and present the outlier adjusted causal estimates (OACE) when both global and distortion tests are significant.

^b^ We did not run Steiger Test if none of the MR analysis resulted significant (NR: not reported in the cell).

^c^ Not enough SNP to perform MR (NR: not reported in the cell).

#### Plots - Forward analyses

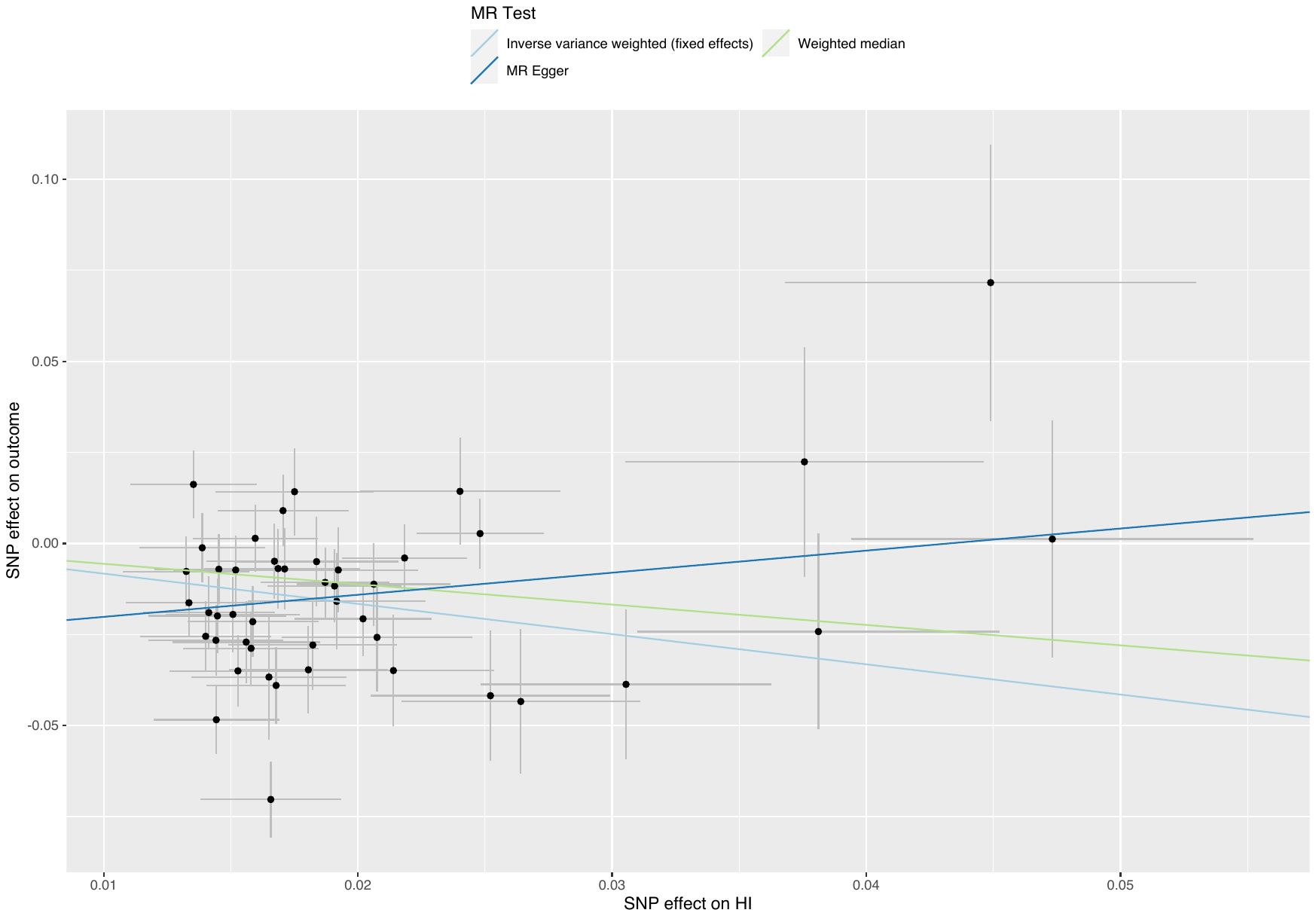

##### Supplementary Figure 30: scatterplot of household income against ADHD

Abbreviations: MR: Mendelian randomization; SNP: single nucleotide polymorphism; HI: household income; ADHD: attention deficit hyperactivity disorder.

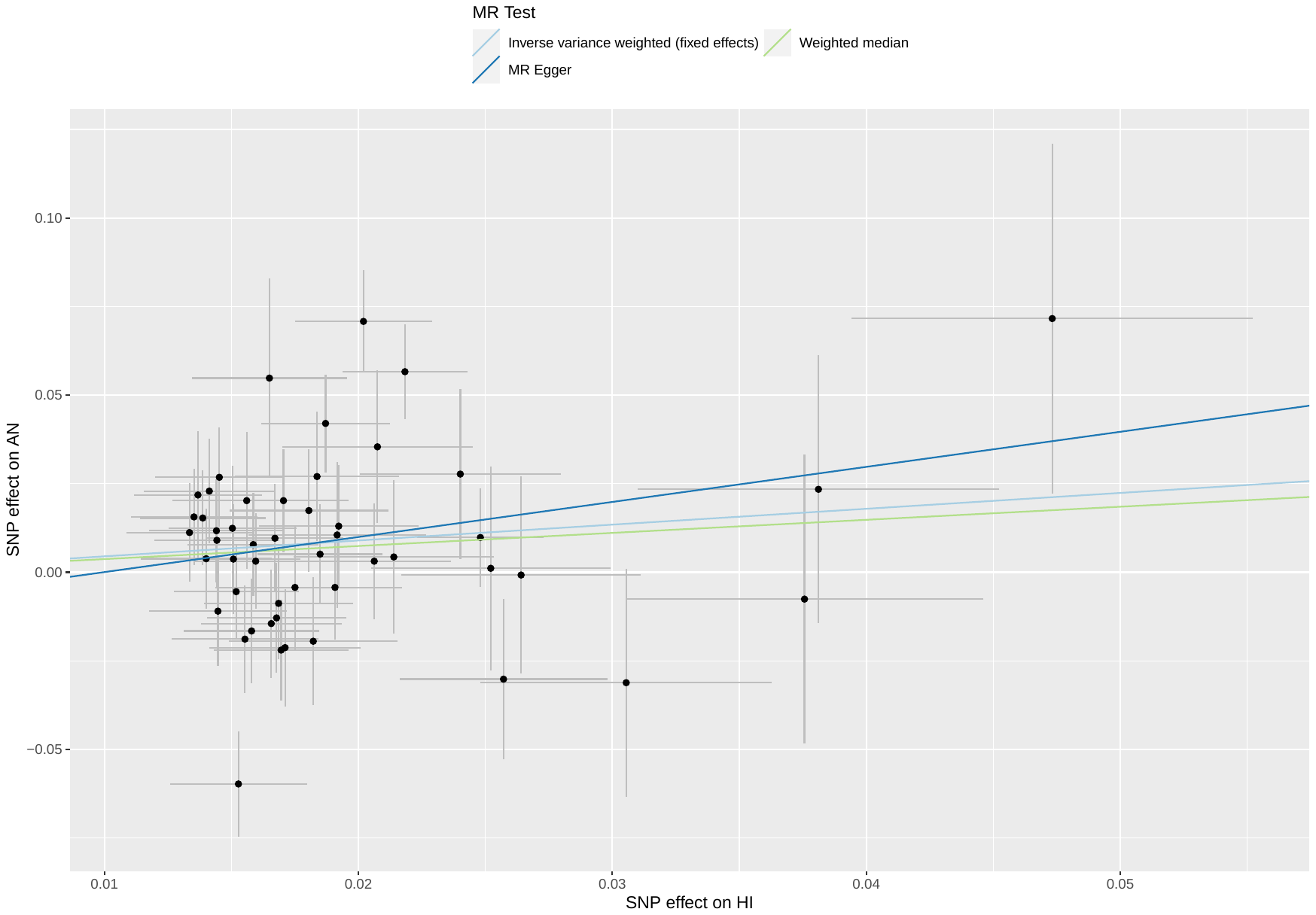

##### Supplementary Figure 31: scatterplot of household income against AN

Abbreviations: MR: Mendelian randomization; SNP: single nucleotide polymorphism; HI: household income; AN: anorexia nervosa.

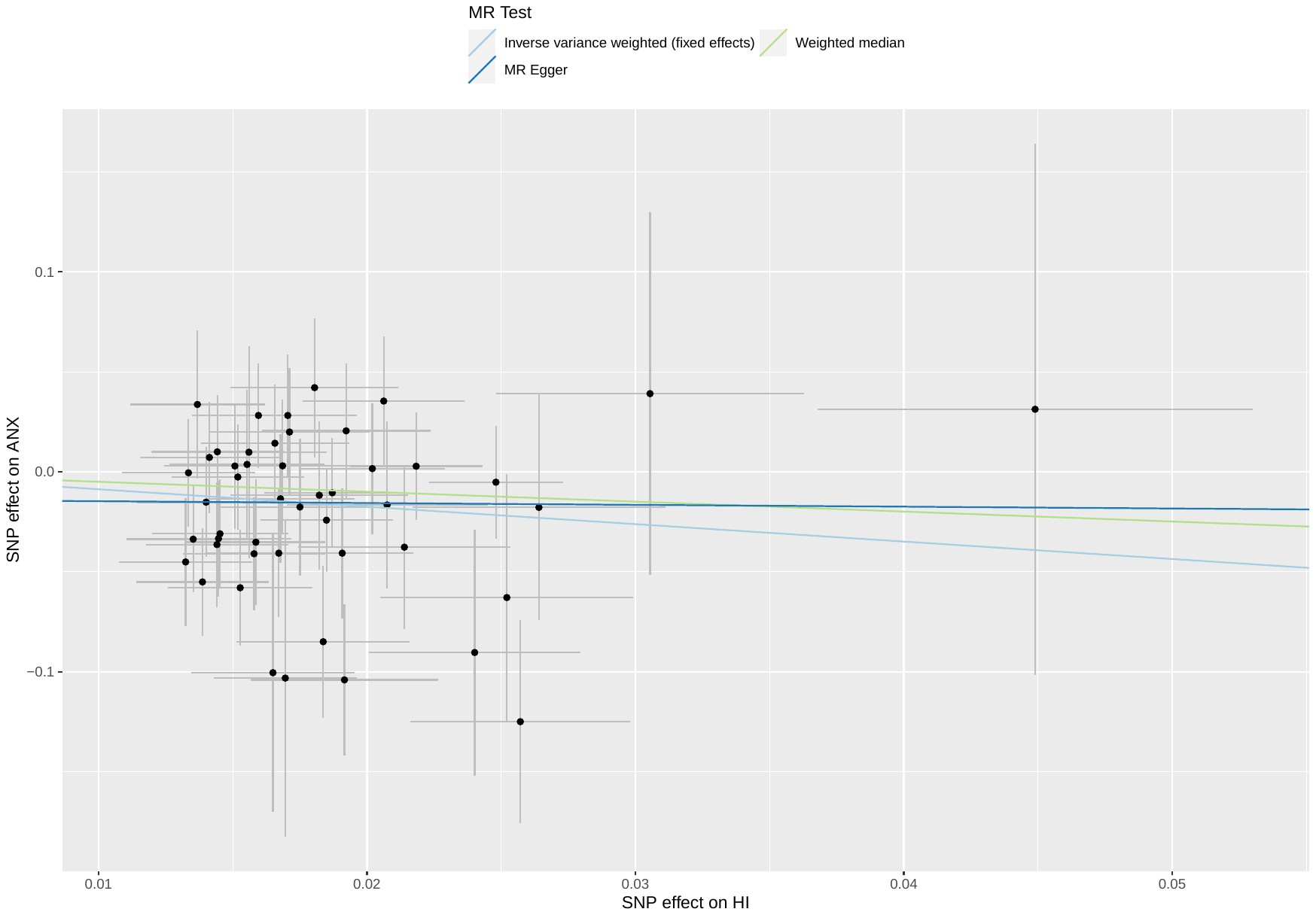

##### Supplementary Figure 32: scatterplot of household income against ANX

Abbreviations: MR: Mendelian randomization; SNP: single nucleotide polymorphism; HI: household income; ANX: anxiety disorder.

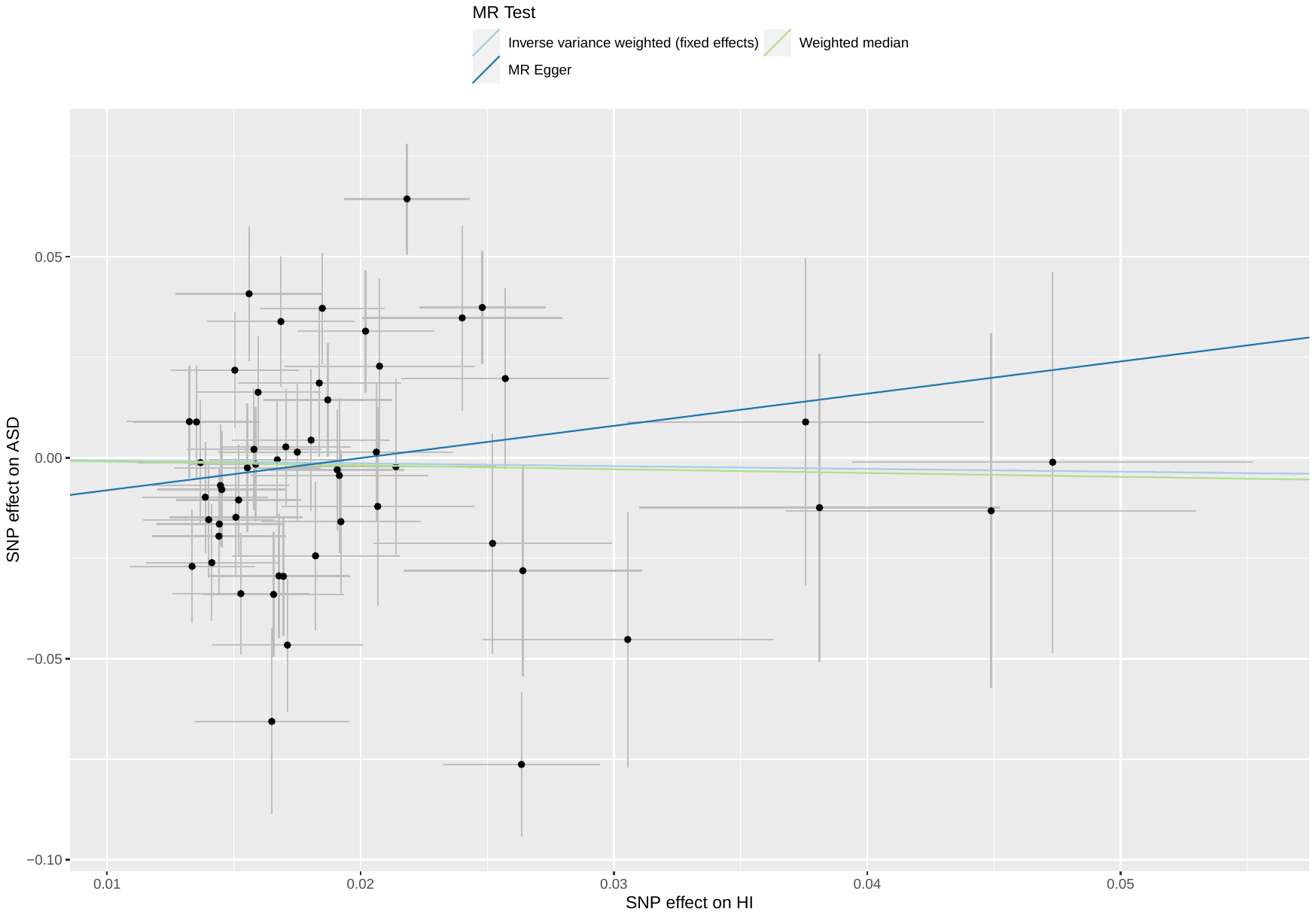

##### Supplementary Figure 33: scatterplot of household income against ASD

Abbreviations: MR: Mendelian randomization; SNP: single nucleotide polymorphism; HI: household income; ASD: autism spectrum disorder.

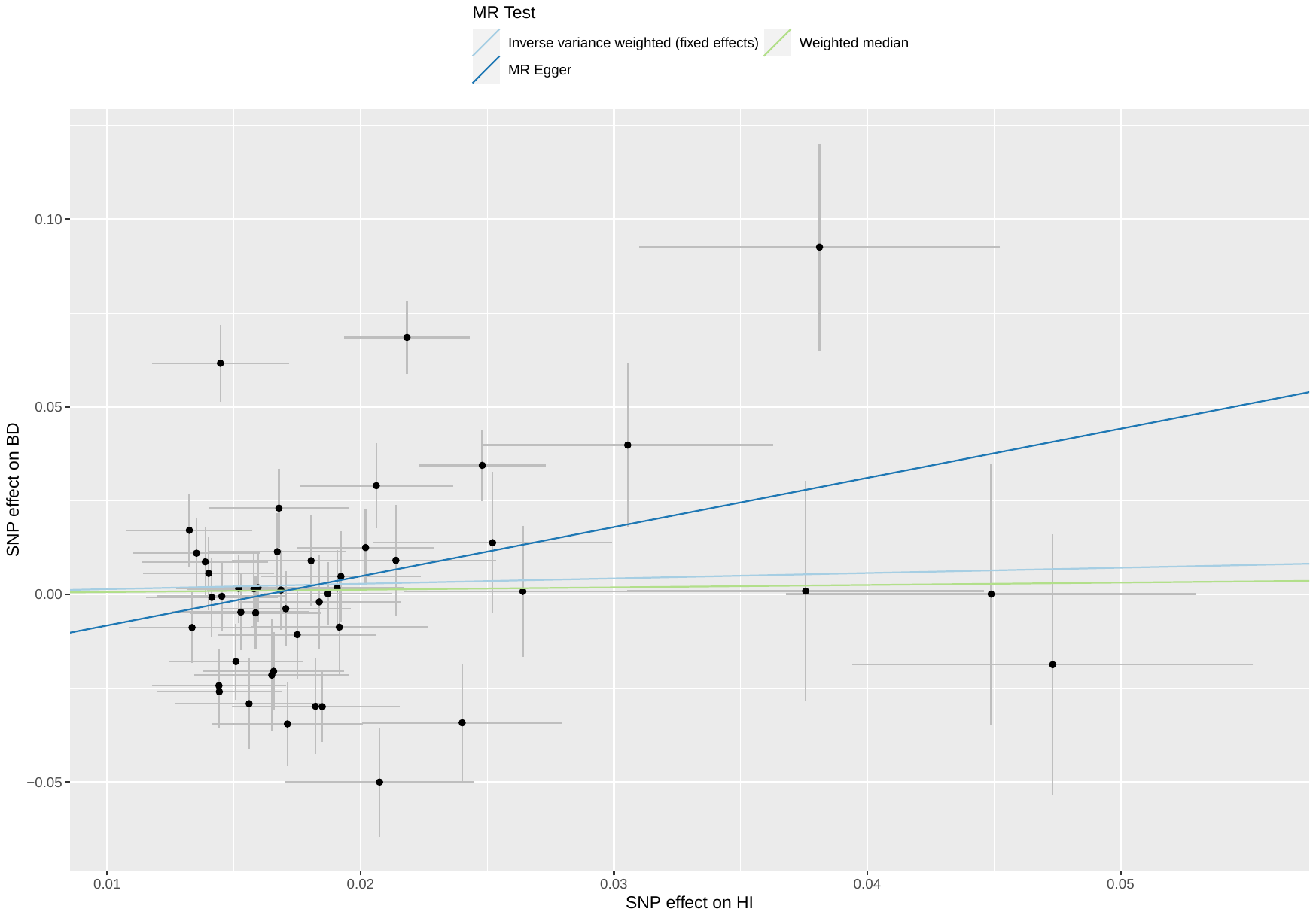

##### Supplementary Figure 34: scatterplot of household income against BD

Abbreviations: MR: Mendelian randomization; SNP: single nucleotide polymorphism; HI: household income; BD: bipolar disorder.

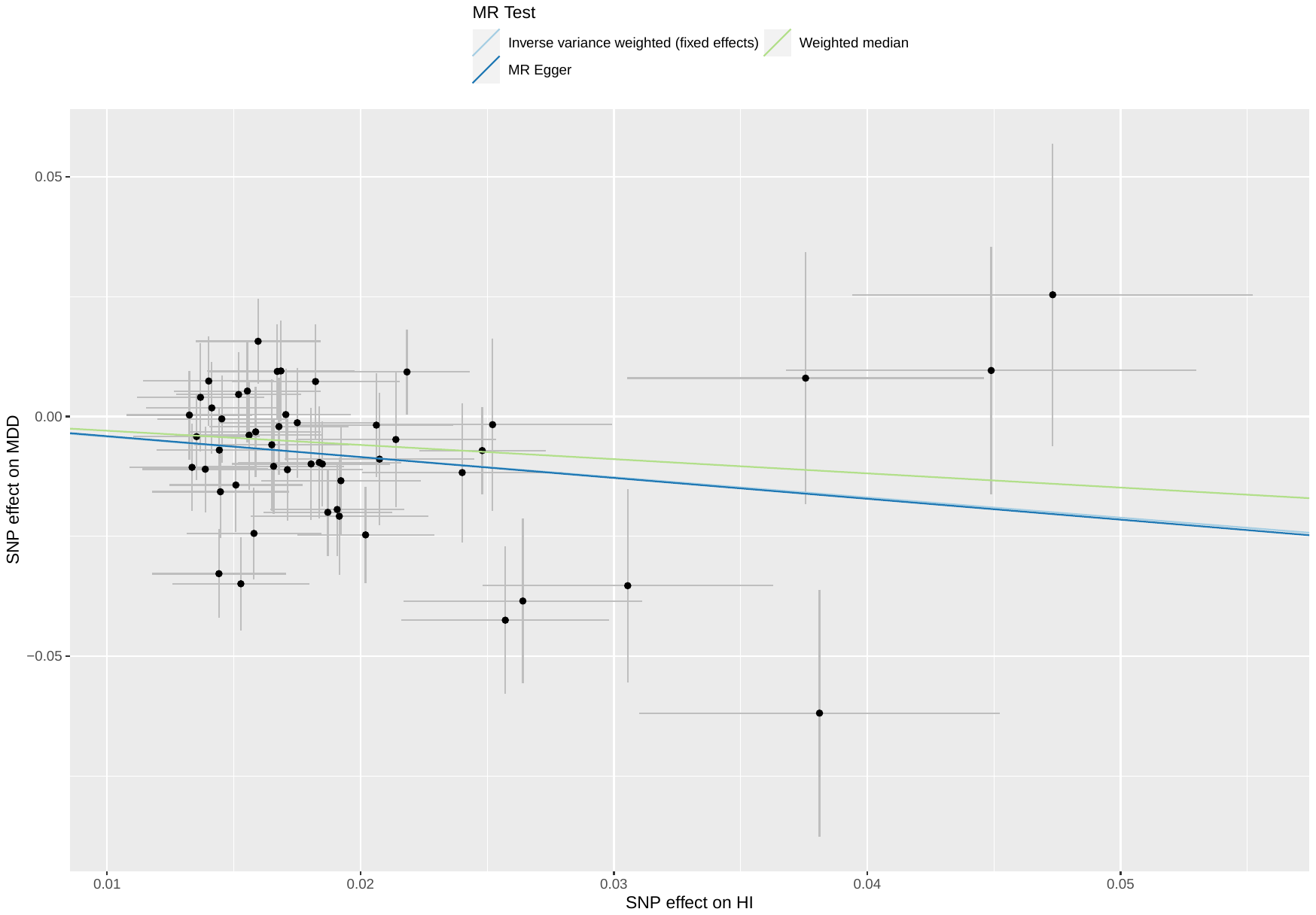

##### Supplementary Figure 35: scatterplot of household income against MDD

Abbreviations: MR: Mendelian randomization; SNP: single nucleotide polymorphism; HI: household income; MDD: major depressive disorder.

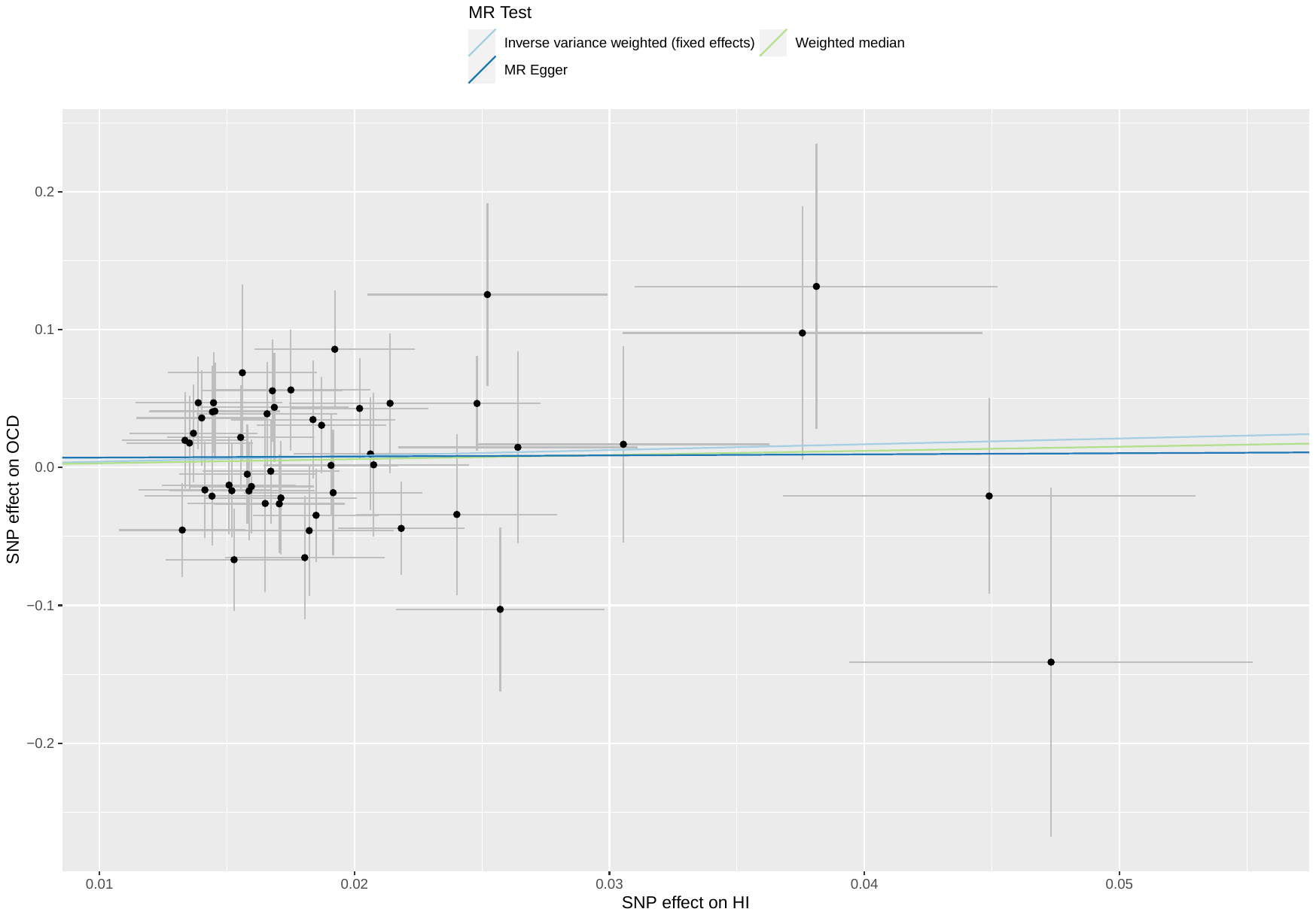

##### Supplementary Figure 36: scatterplot of household income against OCD

Abbreviations: MR: Mendelian randomization; SNP: single nucleotide polymorphism; HI: household income; OCD: obsessive compulsive disorder.

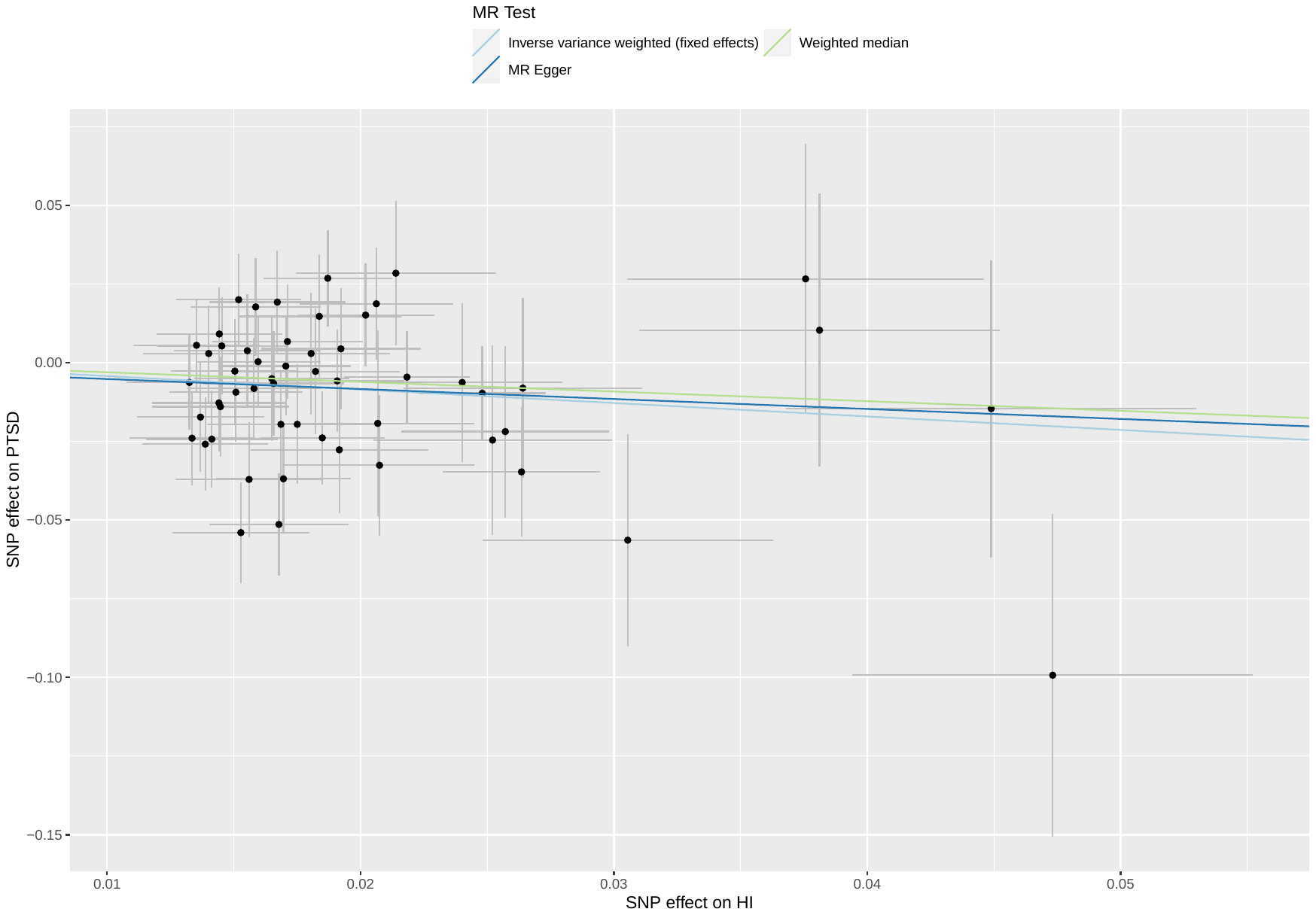

##### Supplementary Figure 37: scatterplot of household income against PTSD

Abbreviations: MR: Mendelian randomization; SNP: single nucleotide polymorphism; HI: household income; PTSD: post-traumatic stress disorder.

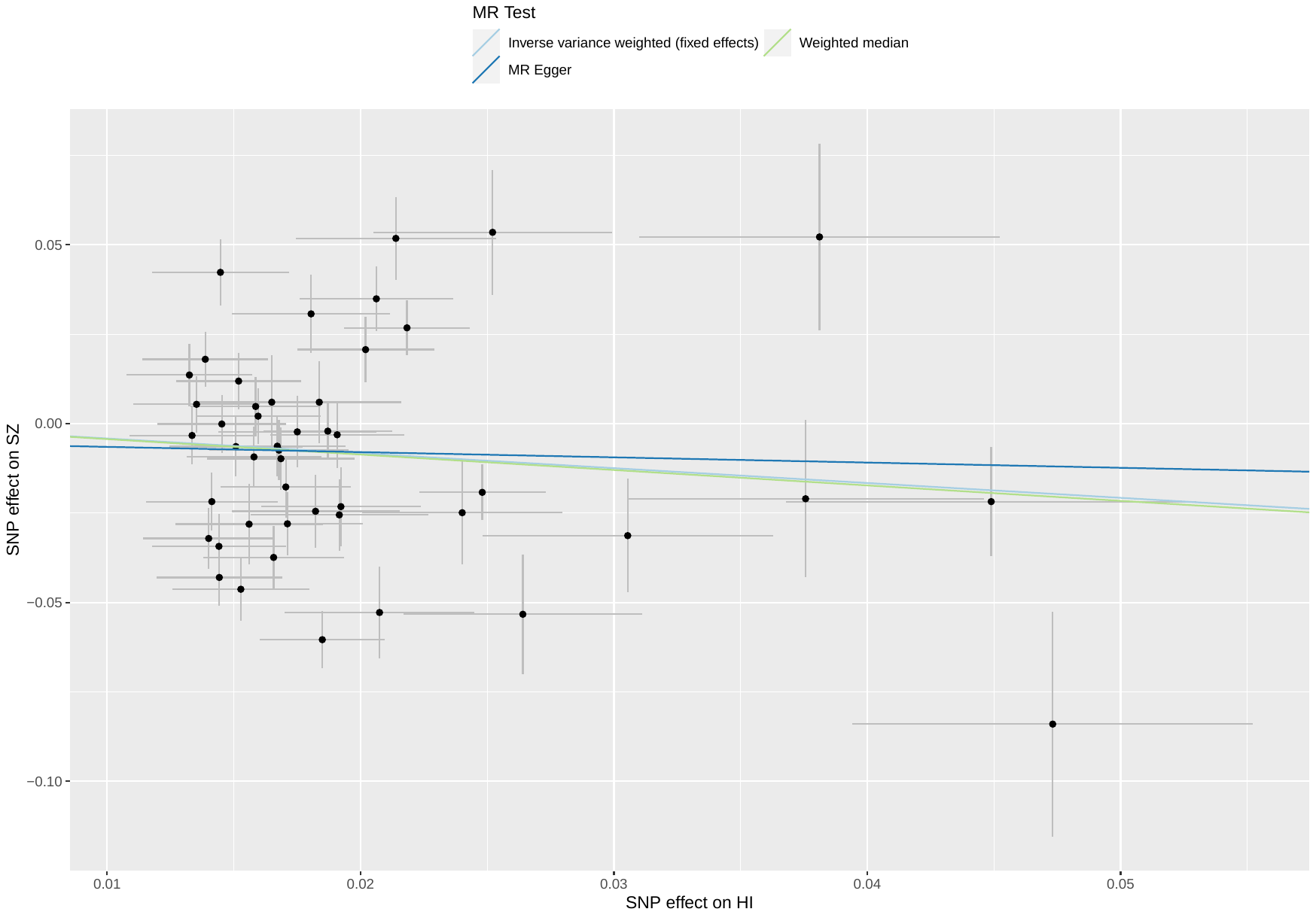

##### Supplementary Figure 38: scatterplot of household income against SZ

Abbreviations: MR: Mendelian randomization; SNP: single nucleotide polymorphism; HI: household income; SZ: schizophrenia.

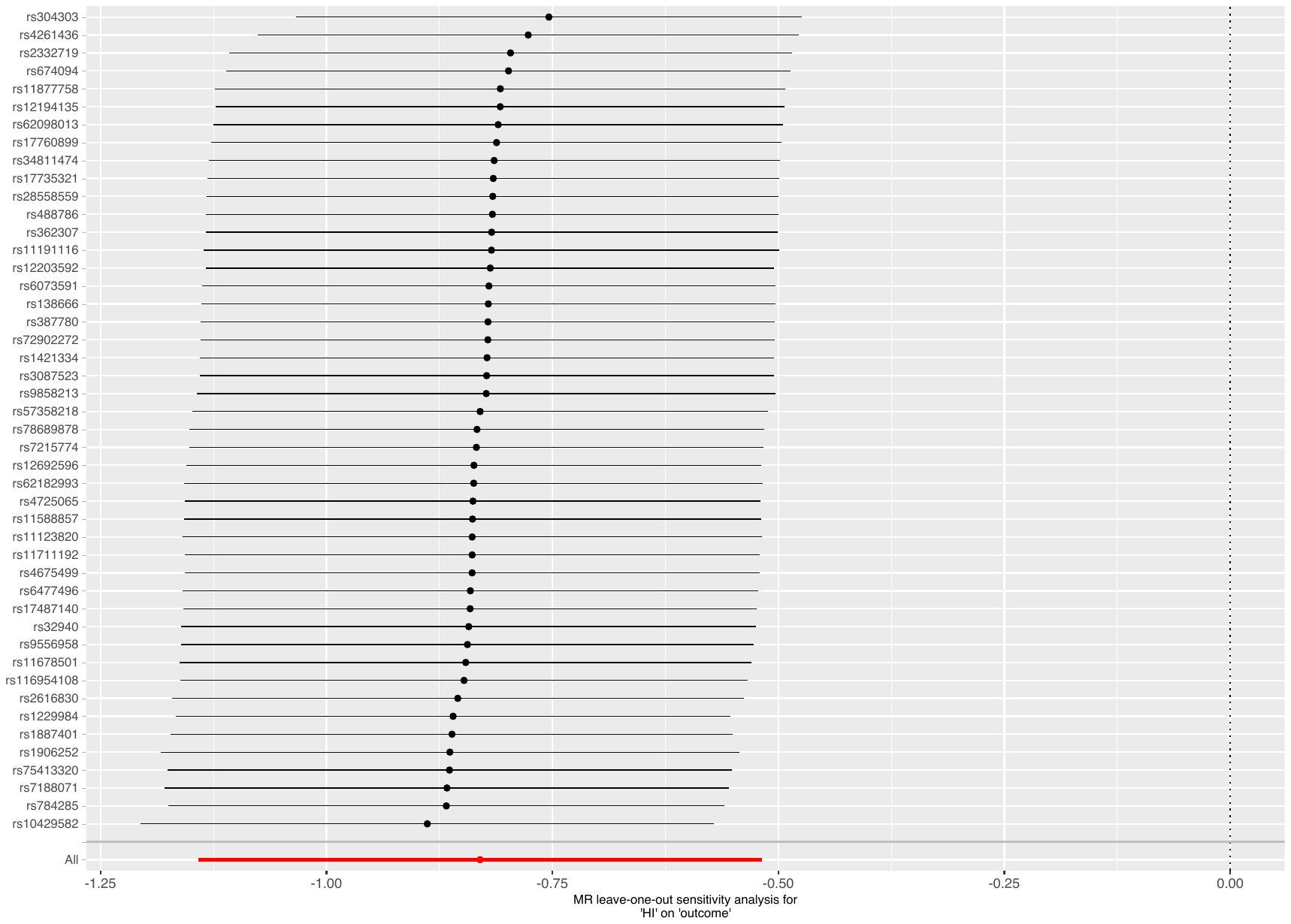

##### Supplementary Figure 39: leave-one-out analysis of household income against ADHD

Abbreviations: MR: Mendelian randomization; HI: household income; ADHD: attention deficit hyperactivity disorder.

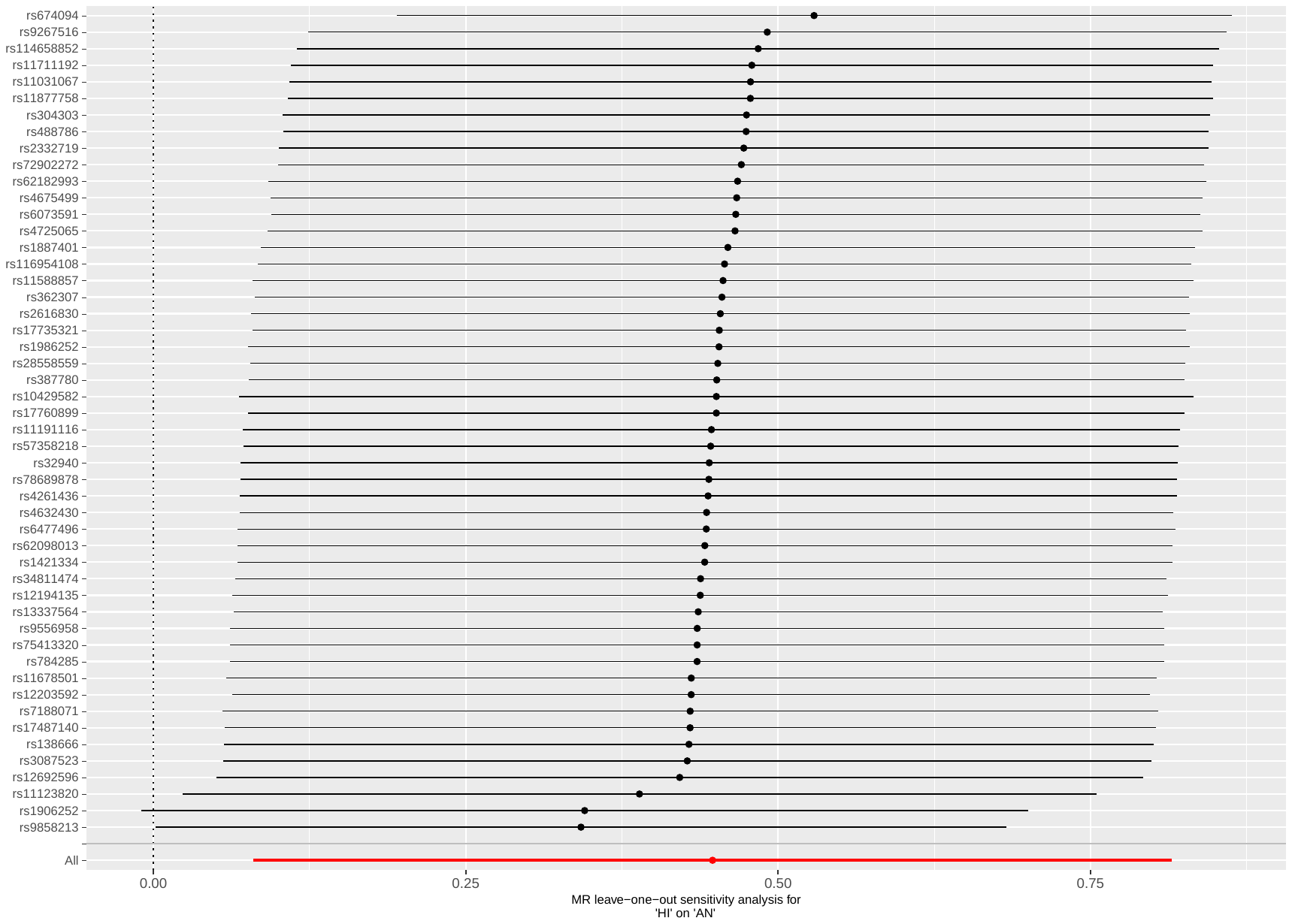

##### Supplementary Figure 40: leave-one-out analysis of household income against AN

Abbreviations: MR: Mendelian randomization; HI: household income; AN: anorexia nervosa.

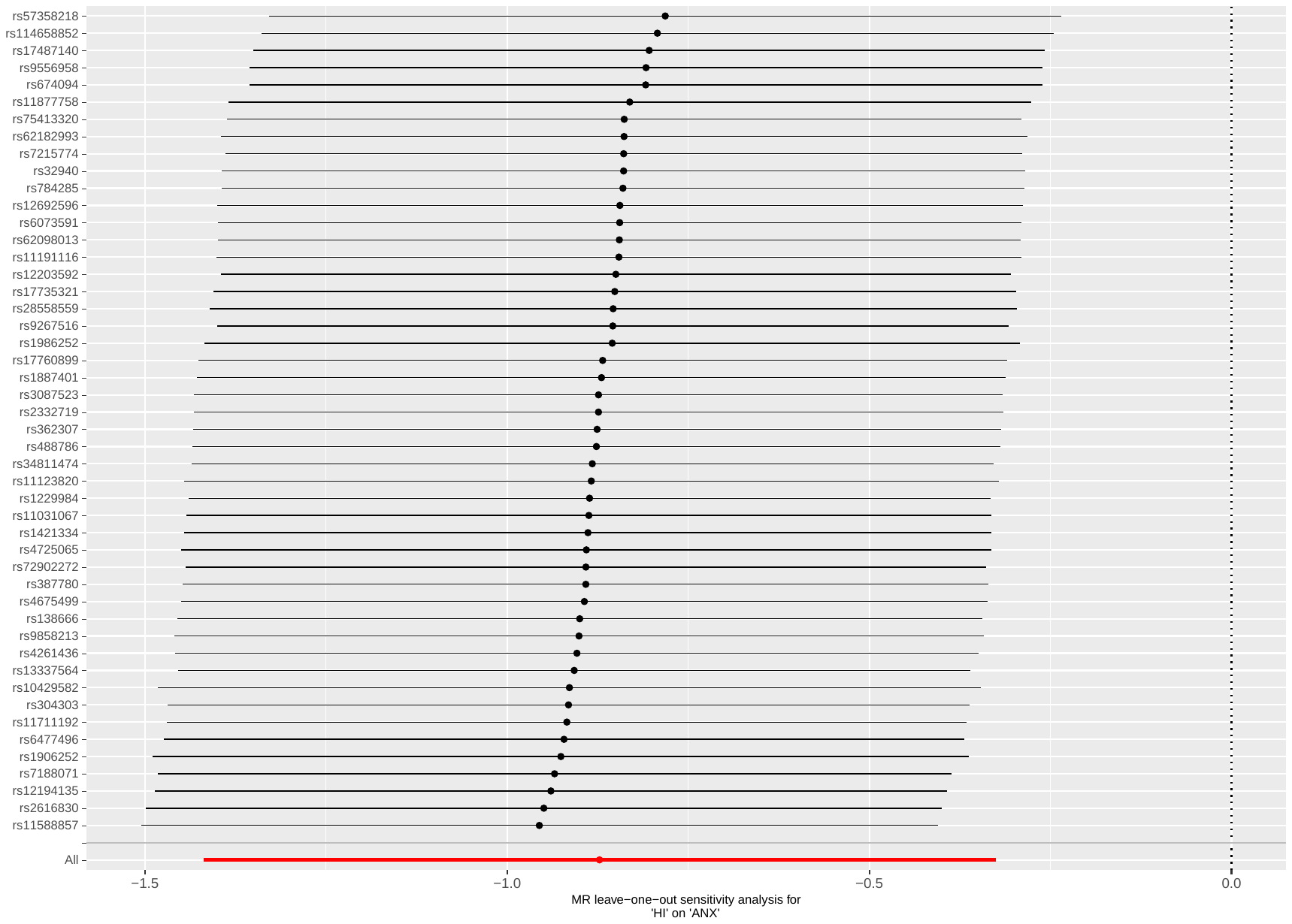

##### Supplementary Figure 41: leave-one-out analysis of household income against ANX

Abbreviations: MR: Mendelian randomization; HI: household income; ANX: anxiety disorder.

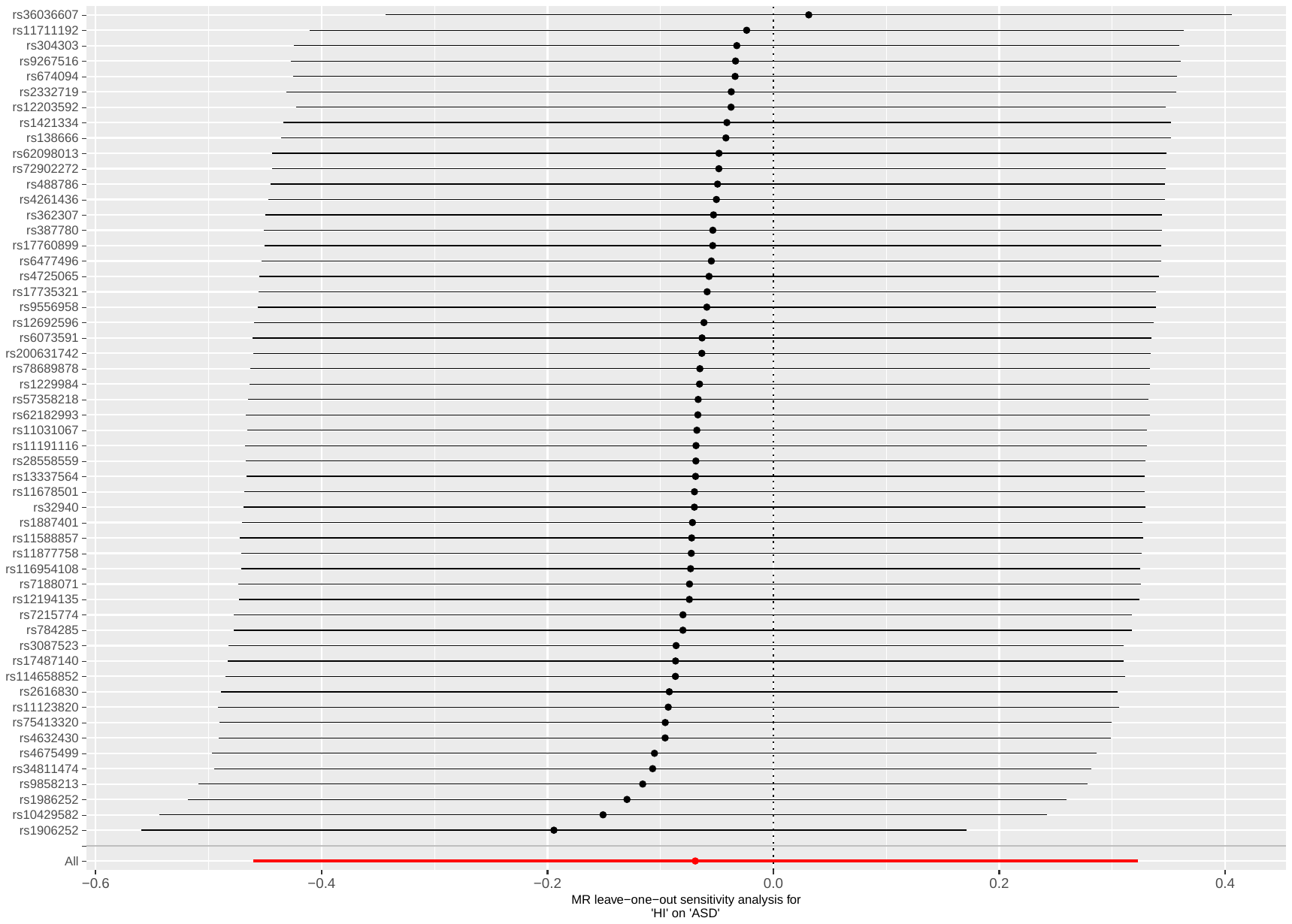

##### Supplementary Figure 42: leave-one-out analysis of household income against ASD

Abbreviations: MR: Mendelian randomization; HI: household income; ASD: autism spectrum disorders.

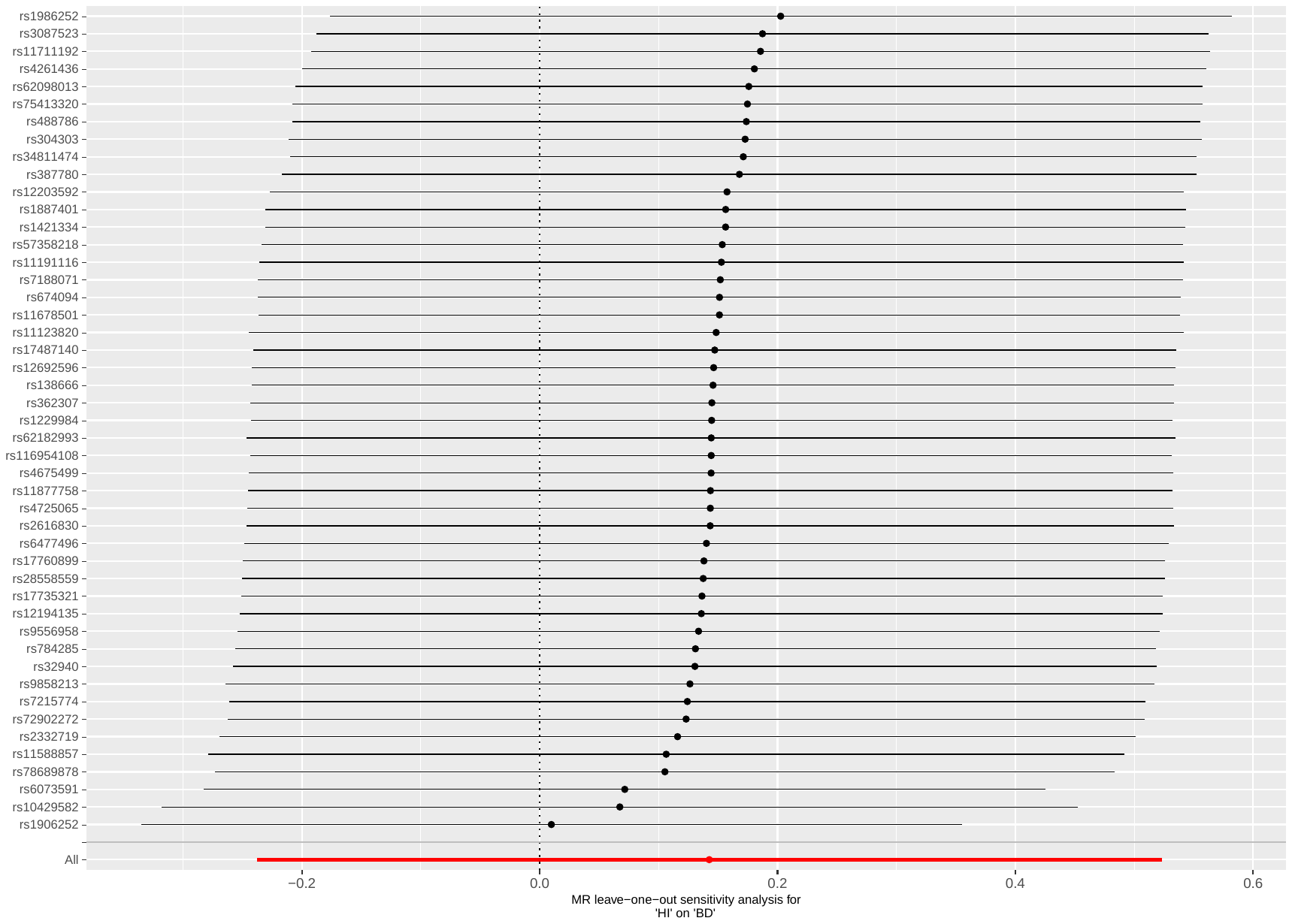

##### Supplementary Figure 43: leave-one-out analysis of household income against BD

Abbreviations: MR: Mendelian randomization; HI: household income; BD: bipolar disorder.

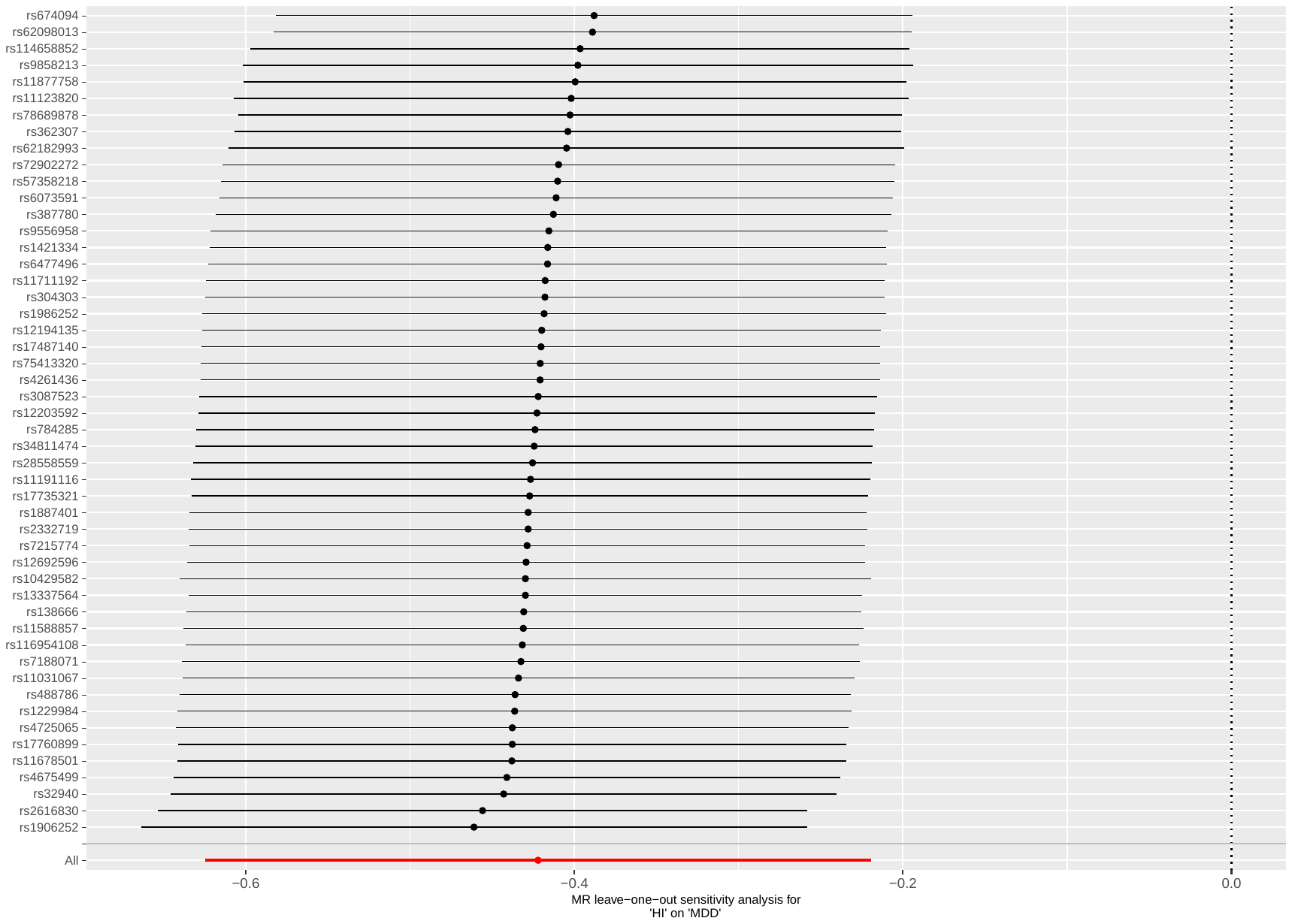

##### Supplementary Figure 44: leave-one-out analysis of household income against MDD

Abbreviations: MR: Mendelian randomization; HI: household income; MDD: major depressive disorder.

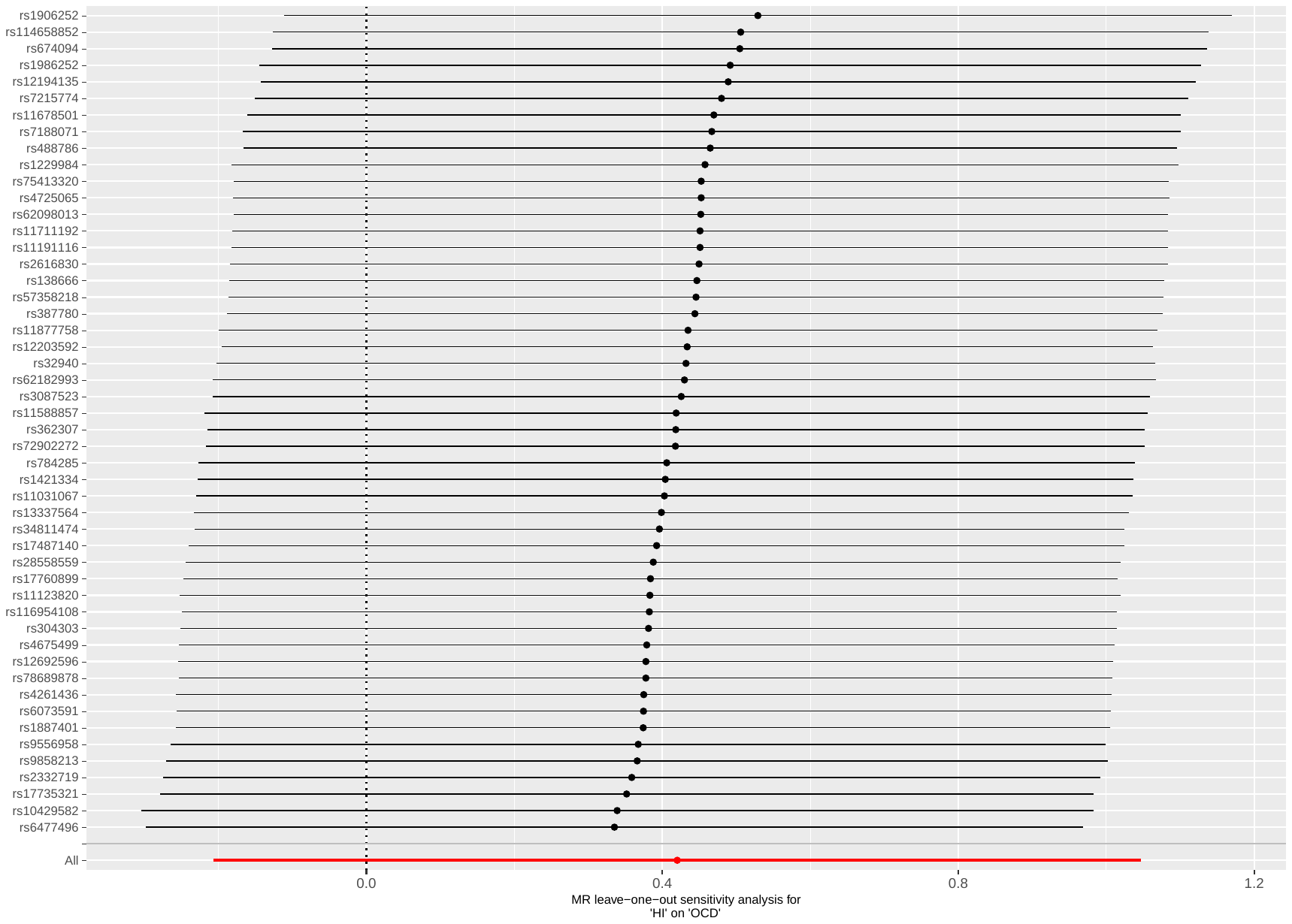

##### Supplementary Figure 45: leave-one-out analysis of household income against OCD

Abbreviations: MR: Mendelian randomization; HI: household income; OCD: obsessive compulsive disorder.

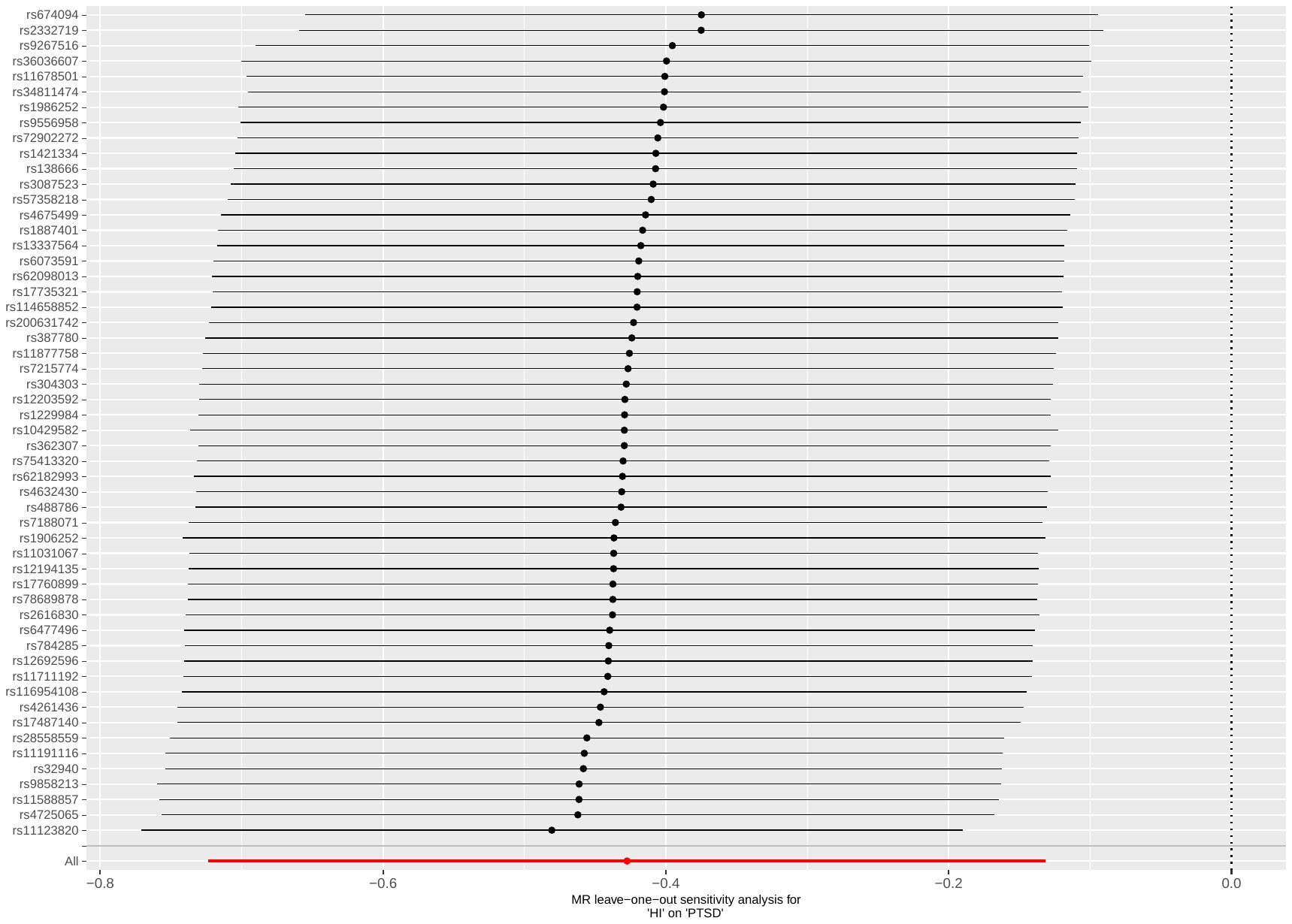

##### Supplementary Figure 46: leave-one-out analysis of household income against PTSD

Abbreviations: MR: Mendelian randomization; HI: household income; PTSD: post-traumatic stress disorder.

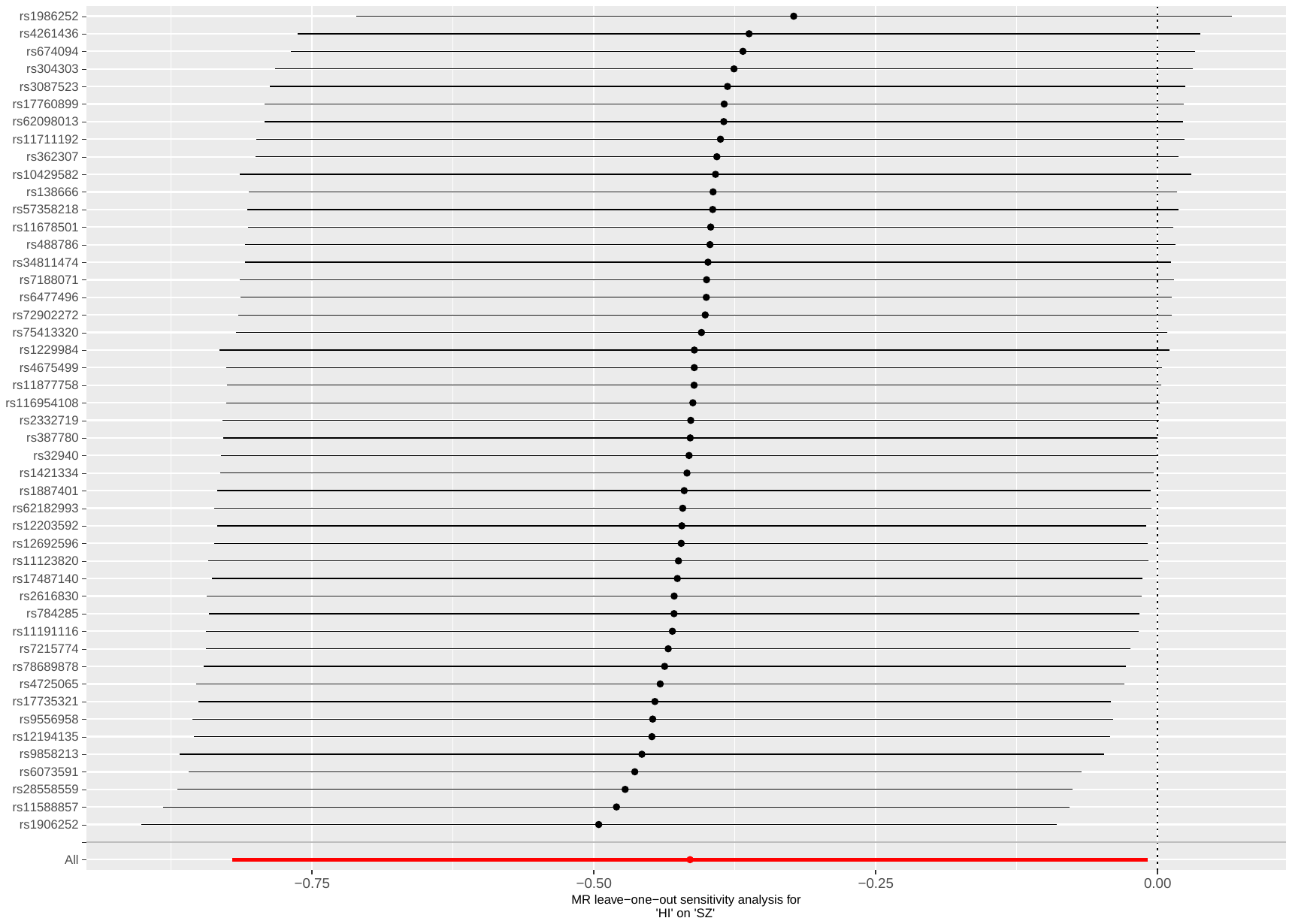

##### Supplementary Figure 47: leave-one-out analysis of household income against SZ

Abbreviations: MR: Mendelian randomization; HI: household income; SZ: schizophrenia.

#### Plots - Backward analyses

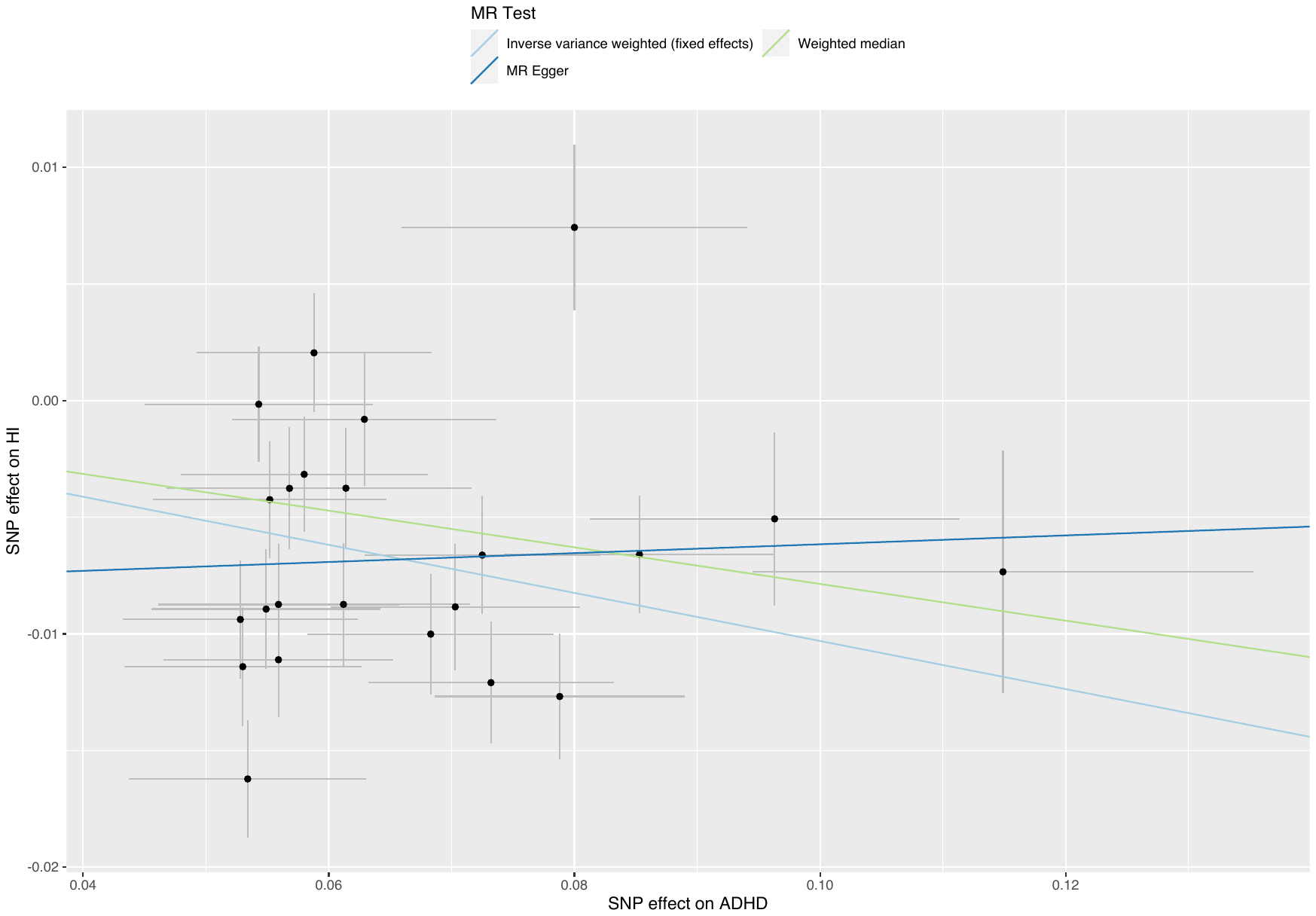

##### Supplementary Figure 48: scatterplot of ADHD against household income

Abbreviations: MR: Mendelian randomization; SNP: single nucleotide polymorphism; HI: household income; ADHD: attention deficit hyperactivity disorder.

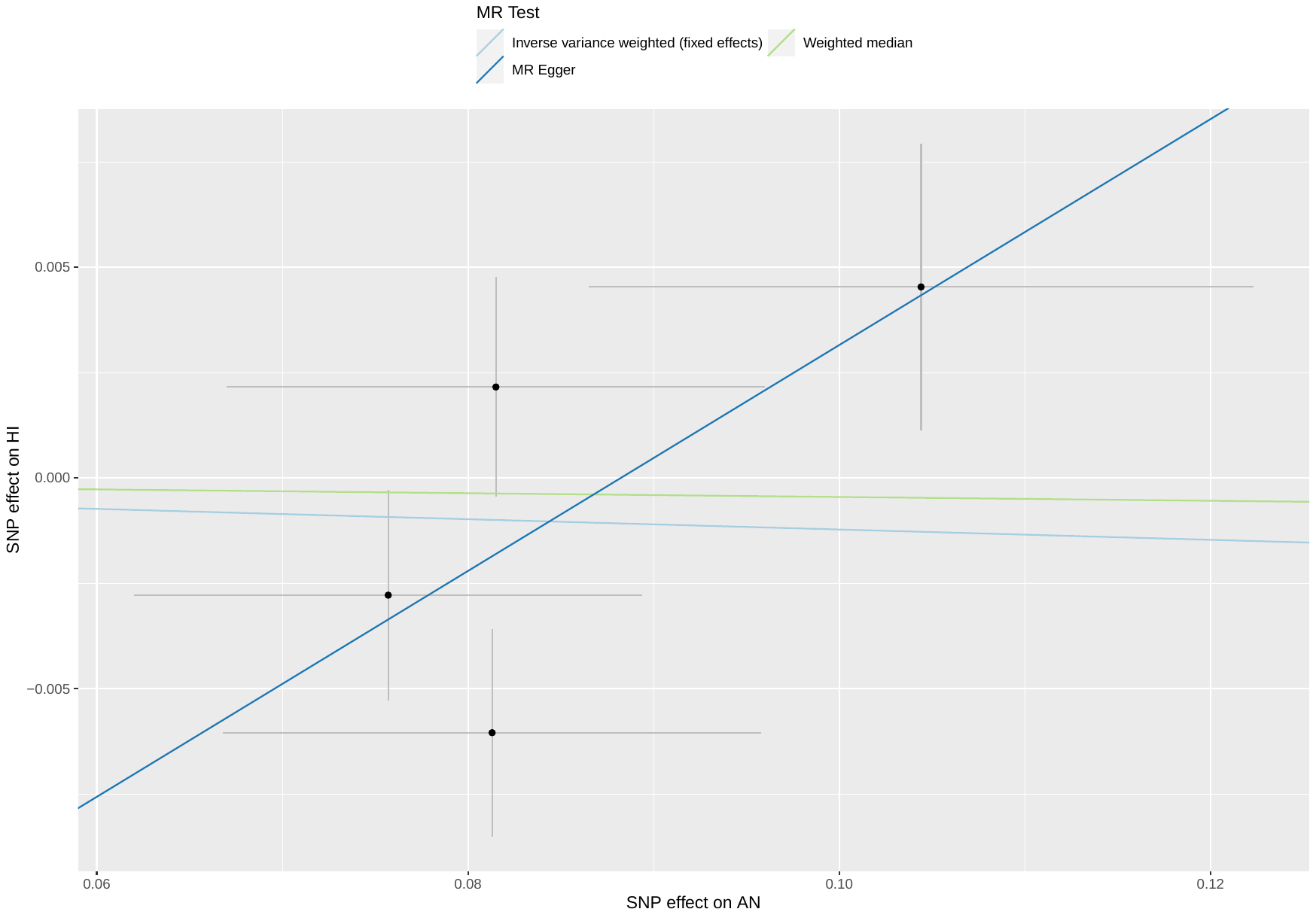

##### Supplementary Figure 49: scatterplot of AN against household income

Abbreviations: MR: Mendelian randomization; SNP: single nucleotide polymorphism; HI: household income; AN: anorexia nervosa.

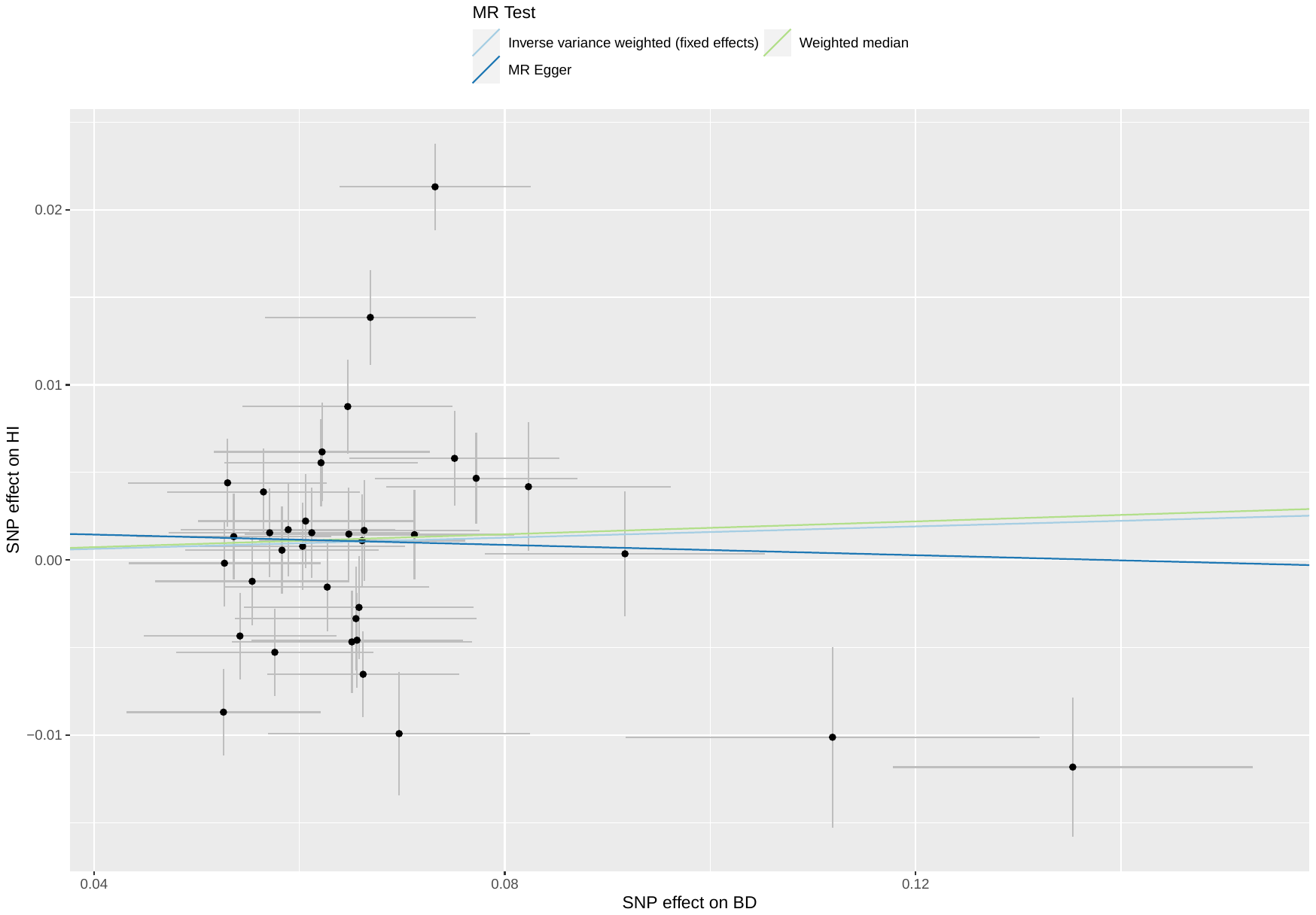

##### Supplementary Figure 50: scatterplot of BD against household income

Abbreviations: MR: Mendelian randomization; SNP: single nucleotide polymorphism; HI: household income; BD: bipolar disorder.

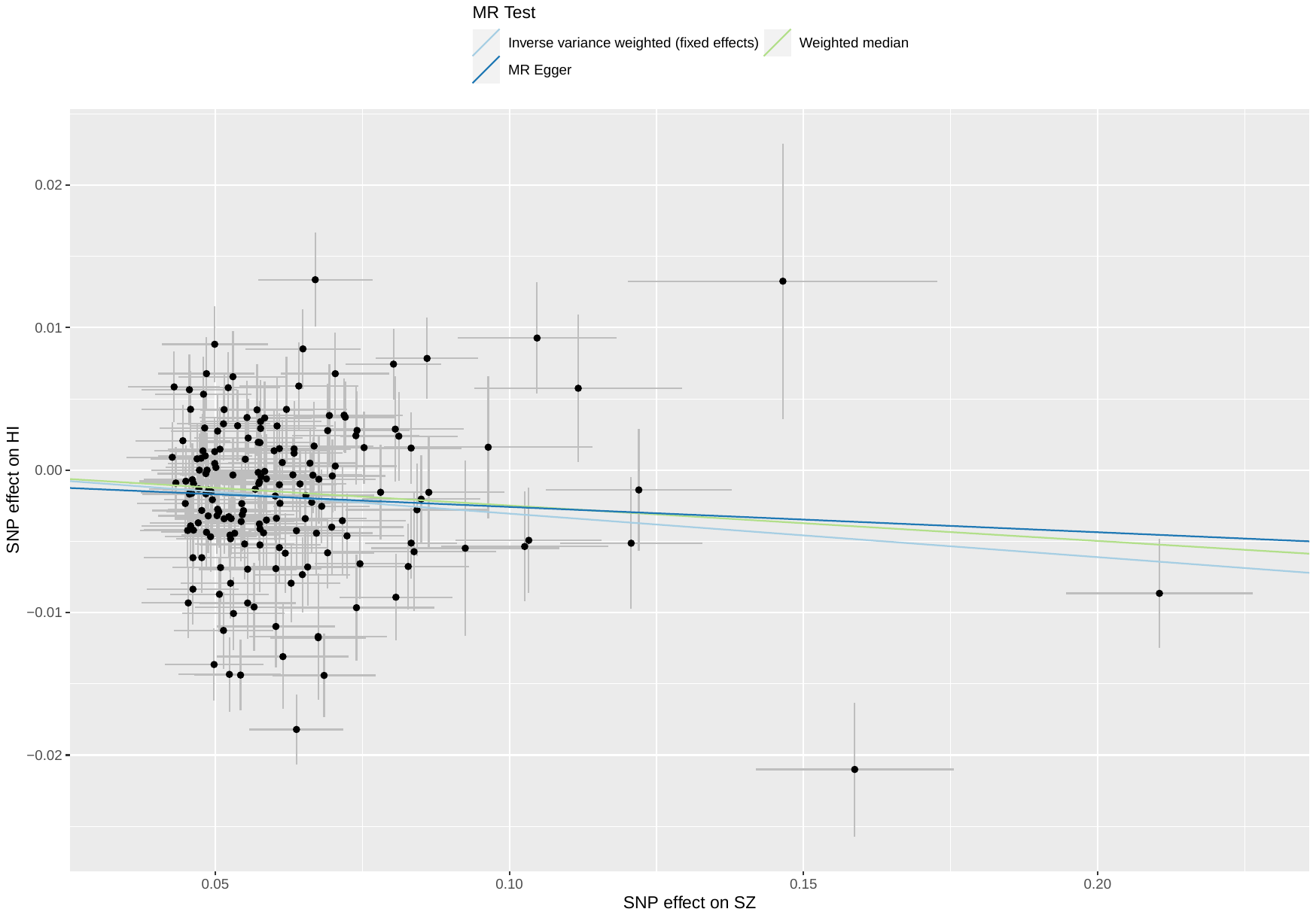

##### Supplementary Figure 51: scatterplot of SZ against household income

Abbreviations: MR: Mendelian randomization; SNP: single nucleotide polymorphism; HI: household income; SZ: schizophrenia.

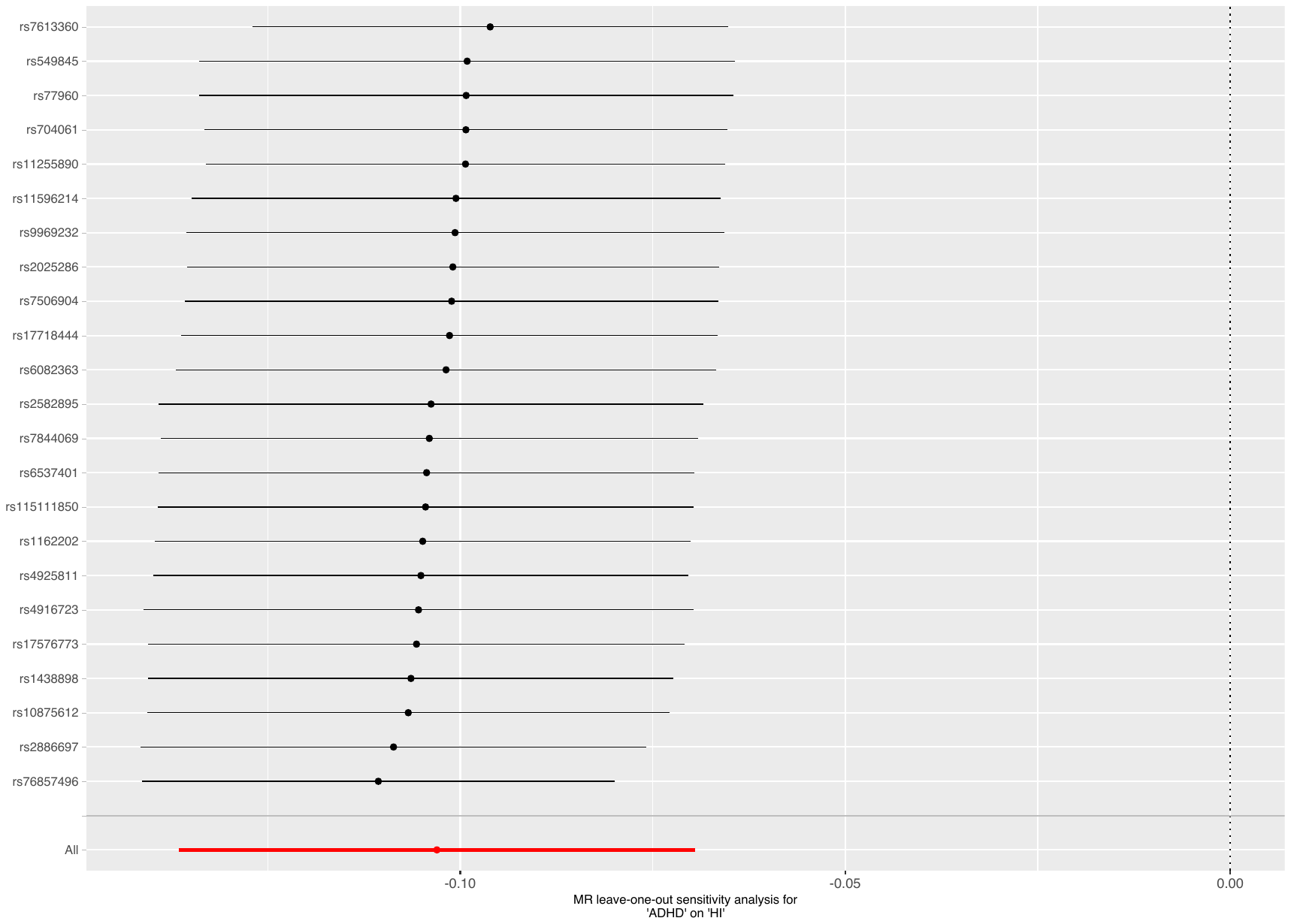

##### Supplementary Figure 52: leave-one-out analysis of ADHD against household income

Abbreviations: MR: Mendelian randomization; HI: household income; ADHD: attention deficit hyperactivity disorder.

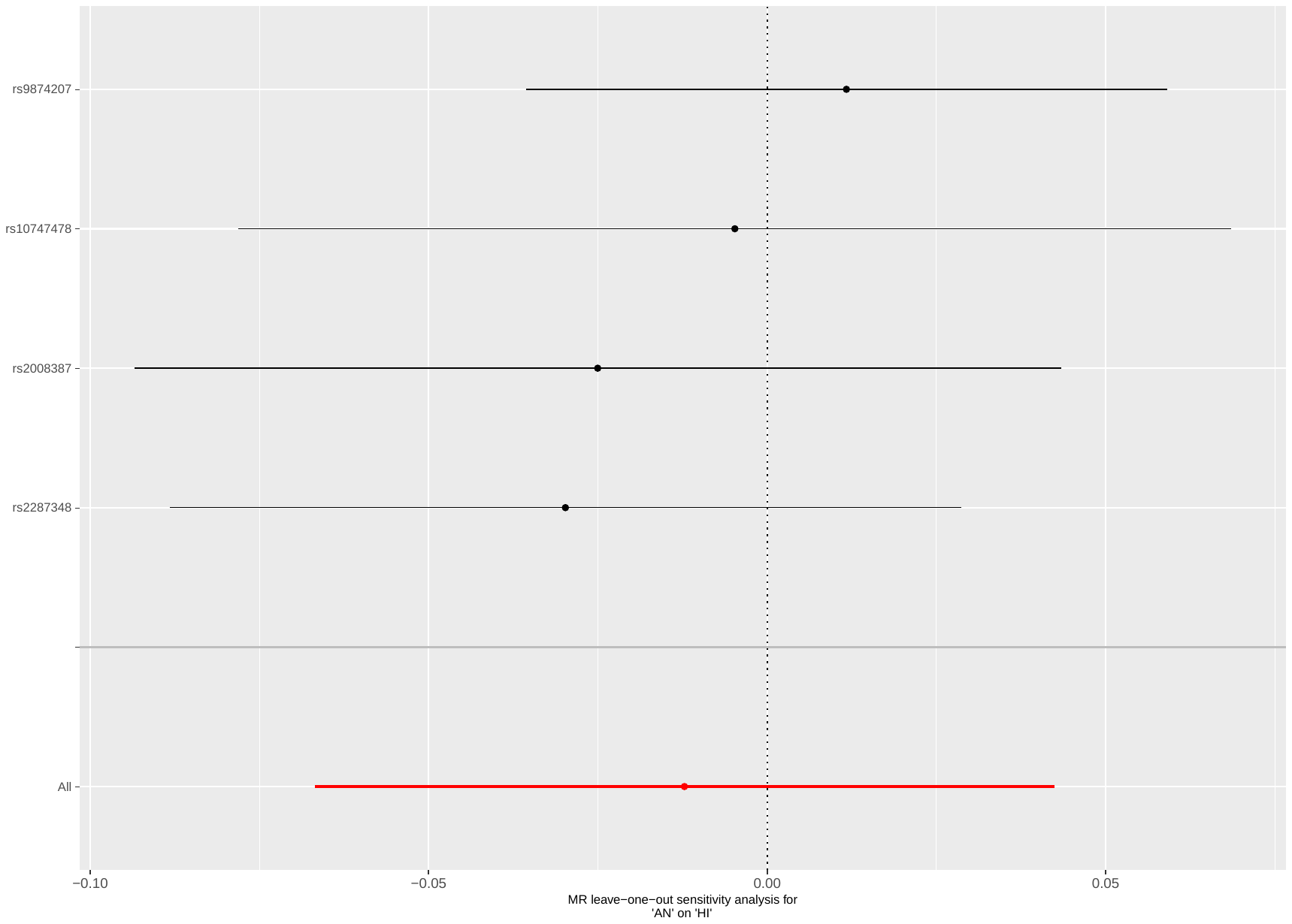

##### Supplementary Figure 53: leave-one-out analysis of AN against household income

Abbreviations: MR: Mendelian randomization; HI: household income; AN: anorexia nervosa.

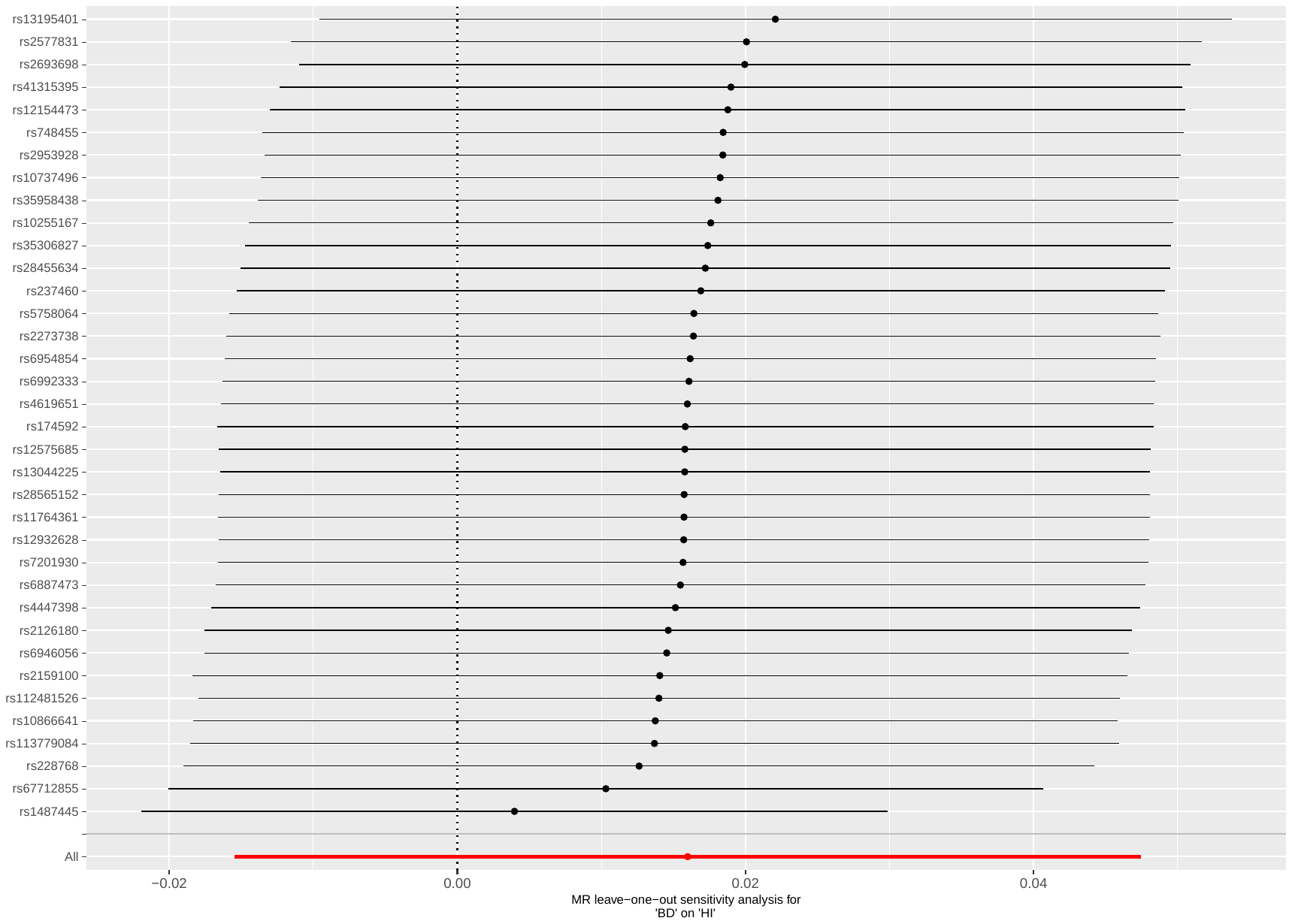

##### Supplementary Figure 54: leave-one-out analysis of BD against household income

Abbreviations: MR: Mendelian randomization; HI: household income; BD: bipolar disorder.

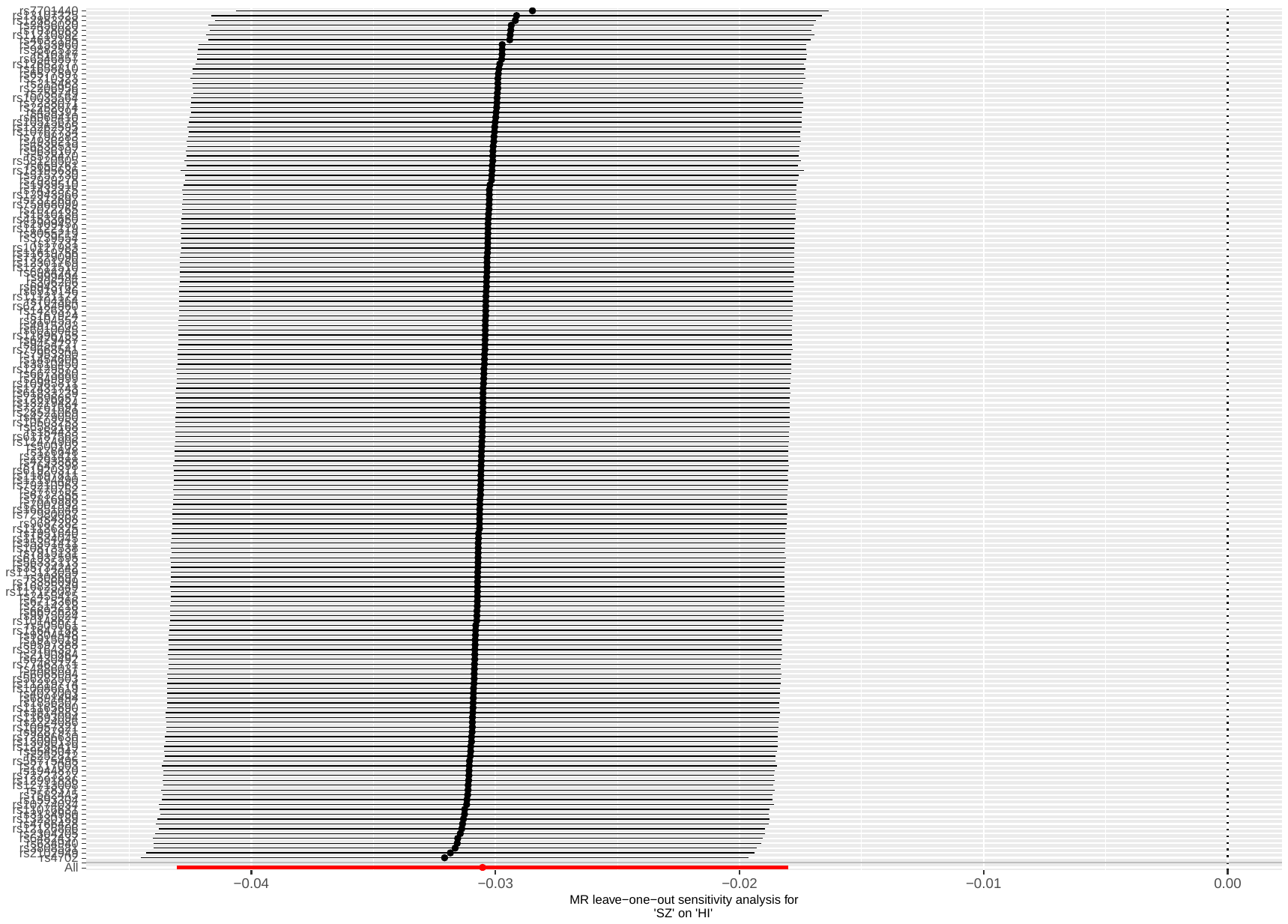

##### Supplementary Figure 55: leave-one-out analysis of SZ against household income

Abbreviations: MR: Mendelian randomization; HI: household income; SZ: schizophrenia.

#### Supplementary Table 11: CAUSE results of the relations between household income and mental illnesses

| **Model 1** | **Model 2** | **∆ ELPD** | **SE ∆ ELPD** | **z-score** | **p-value** |
| --- | --- | --- | --- | --- | --- |
| *Fw: HI on ADHD* | | | | | |
| Null  Null  Sharing | Sharing  Causal  Causal | -26.00  -31.83  -5.83 | 5.93  7.26  1.53 | -4.39  -4.38  -3.80 | <0.001  <0.001  <0.001 |
| *Bw: ADHD on HI* | | | | | |
| Null  Null  Sharing | Sharing  Causal  Causal | -17.77  -23.53  -5.76 | 4.47  6.00  1.60 | -3.98  -3.92  -3.61 | <0.001  <0.001  <0.001 |
| *Fw: HI on AN* | | | | | |
| Null  Null  Sharing | Sharing  Causal  Causal | -4.18  -8.41  -4.24 | 2.04  3.89  1.90 | -2.05  -2.17  -2.24 | 0.040  0.030  0.025 |
| *Bw: AN on HI* | | | | | |
| Null  Null  Sharing | Sharing  Causal  Causal | 0.45  1.15  0.70 | 0.06  0.54  0.48 | 7.08  2.15  1.44 | <0.001  0.032  0.150 |
| *Fw: HI on ANX* | | | | | |
| Null  Null  Sharing | Sharing  Causal  Causal | -1.54  -2.04  -0.50 | 1.86  2.70  1.21 | -0.83  -0.75  -0.41 | 0.407  0.453  0.682 |
| *Bw: ANX on HI* | | | | | |
| Null  Null  Sharing | Sharing  Causal  Causal | 0.09  0.62  0.53 | 0.04  0.36  0.33 | 2.12  1.70  1.57 | 0.034  0.089  0.116 |
| *Fw: HI on ASD* | | | | | |
| Null  Null  Sharing | Sharing  Causal  Causal | 0.49  1.39  0.90 | 0.07  0.26  0.21 | 7.16  5.32  4.20 | <0.001  <0.001  <0.001 |
| *Bw: ASD on HI* | | | | | |
| Null  Null  Sharing | Sharing  Causal  Causal | -0.51  -3.19  -2.68 | 0.62  2.44  1.84 | -0.83  -1.31  -1.46 | 0.407  0.190  0.144 |
| *Fw: HI on BD* | | | | | |
| Null  Null  Sharing | Sharing  Causal  Causal | 0.44  1.03  0.59 | 0.11  0.70  0.61 | 4.12  1.48  0.98 | 0.001  0.069  0.327 |
| *Bw: BD on HI* | | | | | |
| Null  Null  Sharing | Sharing  Causal  Causal | 0.43  1.35  0.92 | 0.03  0.11  0.11 | 13.18  11.76  8.19 | <0.001  <0.001  <0.001 |
| *Fw: HI on MDD* | | | | | |
| Null  Null  Sharing | Sharing  Causal  Causal | -11.26  -15.12  -3.86 | 4.00  5.31  1.54 | -2.82  -2.85  -2.50 | 0.005  0.004  0.012 |
| *Bw: MDD on HI* | | | | | |
| Null  Null  Sharing | Sharing  Causal  Causal | -0.51  -3.19  -2.68 | 0.62  2.44  1.84 | -0.83  -1.31  -1.46 | 0.407  0.190  0.144 |
| *Fw: HI on OCD* | | | | | |
| Null  Null  Sharing | Sharing  Causal  Causal | -0.10  -0.55  -0.45 | 0.72  1.94  1.25 | -0.14  -0.28  -0.36 | 0.889  0.779  0.719 |
| *Bw: OCD on HI* | | | | | |
| Null  Null  Sharing | Sharing  Causal  Causal | 0.27  0.92  0.65 | 0.08  0.52  0.45 | 3.30  1.77  1.44 | 0.001  0.077  0.150 |
| *Fw: HI on PTSD* | | | | | |
| Null  Null  Sharing | Sharing  Causal  Causal | -6.47  -8.97  -2.50 | 3.09  4.47  1.53 | -2.09  -2.00  -1.64 | 0.037  0.046  0.101 |
| *Bw: PTSD on HI* | | | | | |
| Null  Null  Sharing | Sharing  Causal  Causal | 0.18  0.24  0.06 | 0.10  0.91  0.81 | 1.74  0.26  0.07 | 0.082  0.795  0.944 |
| *Fw: HI on SZ* | | | | | |
| Null  Null  Sharing | Sharing  Causal  Causal | -1.55  -6.27  -4.73 | 0.91  3.00  2.10 | -1.70  -2.09  -2.25 | 0.089  0.037  0.024 |
| *Bw: SZ on HI* | | | | | |
| Null  Null  Sharing | Sharing  Causal  Causal | -2.78  -6.76  -3.98 | 1.61  3.53  1.94 | -1.73  -1.91  -2.05 | 0.084  0.056  0.040 |

Abbreviations: MR: mendelian randomization; ELPD: expected log pointwise posterior density; 95%CI: 95% confidence interval; SE: standard error; Fwd: forward MR; Bwd: backward MR; HI: household income; ADHD: attention deficit hyperactivity disorder; AN: anorexia nervosa; ANX: anxiety disorder; ASD: autism spectrum disorder; BD: bipolar disorder; MDD: major depressive disorder; OCD: obsessive-compulsive disorder; PTSD: post-traumatic stress disorder; SZ: schizophrenia.

#### Supplementary Table 12: Results of univariable bidirectional Mendelian Randomization of household income against mental illnesses, after Steiger filtering

| **MR** | **N SNP** | **IVW, B (95% CI)** | **p-value** | **WM, B (95% CI)** | **p-value** | **MR-Egger, B (95% CI)** | **p-value** | **Egger intercept p-value** | **Mean F** |
| --- | --- | --- | --- | --- | --- | --- | --- | --- | --- |
| HI on ADHD | 44 | -0.698 (-0.885; -0.511) | <0.001 | -0.549 (-0.856; 0.243) | <0.001 | 0.366 (-0.716; 1.45) | 0.511 | 0.022 | 36.8 |
| HI on AN | 47 | 0.315 (0.045; 0.585) | 0.022 | 0.301 (-0.101; 0.702) | 0.142 | -0.018 (-1.29; 1.25) | 0.978 | 0.315 | 36.1 |
| HI on ANX | 43 | -0.473 (-1.04; 0.097) | 0.104 | -0.206 (-1.01; 0.596) | 0.615 | 0.619 (-2.41; 3.65) | 0.690 | 0.075 | 37.8 |
| HI on ASD | 44 | -0.044 (-0.321; 0.233) | 0.754 | -0.092 (-0.500; 0.317) | 0.660 | 0.791 (-0.362; 1.94) | 0.186 | <0.001 | 36.2 |
| HI on BD | 36 | 0.148 (-0.058; 0.354) | 0.160 | 0.083 (-0.219; 0.386) | 0.589 | 0.547 (-0.476; 1.57) | 0.302 | 0.021 | 36.7 |
| HI on MDD | 48 | -0.354 (-0.524; -0.183) | <0.001 | -0.288 (-0.549; -0.027) | 0.031 | -0.650 (-1.42; 0.116) | 0.103 | 0.930 | 37.1 |
| HI on OCD | 29 | 0.043 (-0.820; 0.734) | 0.913 | -0.284 (-1.35; 0.785) | 0.602 | 0.322 (-2.91; 3.56) | 0.847 | 0.578 | 40.3 |
| HI on PTSD | 54 | -0.427 (-0.693; -0.161) | 0.002 | -0.306 (-0.708; 0.096) | 0.135 | -0.317 (-1.59; 0.952) | 0.626 | 0.861 | 37.5 |
| HI on SZ | 44 | -0.214 (-0.370; -0.059) | 0.007 | -0.405 (-0.695; -0.114) | 0.006 | -0.374 (-1.82; 1.07) | 0.615 | 0.207 | 36.9 |

Abbreviations: HI: household income; ADHD: attention deficit hyperactivity disorder; AN: anorexia nervosa; ANX: anxiety disorder; ASD: autism spectrum disorders; BD: bipolar disorder; MDD: major depressive disorder; OCD: obsessive-compulsive disorder; PTSD: post-traumatic stress disorder; SZ: schizophrenia; MR: mendelian randomization; SNP: single nucleotide polymorphism; IVW: inverse variance weighted (fixed effect); B: effect estimates are log-odds; 95% CI: 95% confidence interval; WM: weighted median; NR: not reported because not enough SNP to perform MR.

### **Univariable Mendelian randomization of occupational income and mental illnesses**

#### Supplementary Table 13: Odds Ratio of univariable forward Mendelian randomization analysis of occupational income against mental illnesses

| **MR: method** | **OR (95% CI)** | **p-value** |
| --- | --- | --- |
| OI → ADHD:  IVW  WM  MR-Egger | 0.424 (0.350; 0.512)  0.564 (0.410; 0.776)  1.14 (0.215; 6.05) | <0.001  <0.001  0.878 |
| OI→ AN:  IVW  WM  MR-Egger | 1.84 (1.40; 2.41)  1.55 (0.980; 2.45)  12.3 (1.89; 80.3) | <0.001  0.054  0.013 |
| OI → ANX:  IVW  WM  MR-Egger | 0.896 (0.510; 1.57)  0.879 (0.385; 2.00)  2.15 (0.095; 49.0) | 0.702  0.749  0.634 |
| OI → ASD:  IVW  WM  MR-Egger | 1.60 (1.21; 2.12)  1.40 (0.872; 2.26)  10.6 (1.32; 85.1) | 0.001  0.167  0.034 |
| OI → BD:  IVW  WM  MR-Egger | 1.22 (1.02; 1.47)  1.15 (0.828; 1.59)  6.58 (1.14; 37.9) | 0.033  0.407  0.043 |
| OI → MDD:  IVW  WM  MR-Egger | 0.724 (0.605; 0.867)  0.883 (0.655; 1.19)  1.36 (0.333; 5.57) | <0.001  0.408  0.671 |
| OI → OCD:  IVW  WM  MR-Egger | 1.82 (0.925; 3.59)  1.70 (0.620; 4.66)  0.116 (0.004; 3.09) | 0.083  0.290  0.208 |
| OI → PTSD:  IVW  WM  MR-Egger | 0.950 (0.707; 1.28)  0.763 (0.497; 1.17)  0.665 (0.105; 4.23) | 0.736  0.235  0.669 |
| OI → SZ:  IVW  WM  MR-Egger | 0.830 (0.709; 0.971)  0.839 (0.607; 1.16)  1.22 (0.126; 11.9) | 0.020  0.272  0.863 |

Abbreviations: MR: Mendelian randomization; OR: Odds Ratio; 95% CI: 95% confidence intervals; OI: occupational income; ADHD: attention deficit hyperactivity disorder; AN: anorexia nervosa; ANX: anxiety disorder; ASD: autism spectrum disorders; BD: bipolar disorder; MDD: major depressive disorder; OCD: obsessive-compulsive disorder; PTSD: post-traumatic stress disorder; SZ: schizophrenia; IVW: inverse variance weighted (fixed effect); WM: weighted median; NR: not reported because not enough SNP to perform MR.

#### Supplementary Table 14: results of bidirectional MR of Occupational Income (OI) against mental health traits

| **MR** | **N SNP** | **IVW, B (95% CI)** | **p-value** | **IVW Q(df)** | **Q p-value** | **WM, B (95% CI)** | **p-value** | **MR-Egger, B (95% CI)** | **p-value** | **Egger intercept p-value** | **Steiger Test p-value** | **MR-PRESSO** | **Mean F** |
| --- | --- | --- | --- | --- | --- | --- | --- | --- | --- | --- | --- | --- | --- |
| Fw: OI on ADHD | 33 | -0.859 (-1.05; -0.669) | <0.001 | 100 (32) | <0.001 | -0.572 (-0.891; -0.253) | <0.001 | 0.132 (-1.54; 1.80) | 0.878 | 0.243 | <0.001 | DT; p=0.560 | 40.3 |
| Bw: ADHD on OI | 23 | -0.102 (-0.120; -0.083) | <0.001 | 113 (22) | <0.001 | -0.102 (-0.135; -0.069) | <0.001 | -0.052 (-0.297; 0.192) | 0.679 | 0.692 | <0.001 | DT; p=0.598 | 39.2 |
| Fw: OI on AN | 34 | 0.606 (0.337; 0.879) | <0.001 | 72 (33) | <0.001 | 0.437 (-0.007; 0.882) | 0.054 | 2.51 (0.635; 4.39) | 0.013 | 0.051 | <0.001 | DT; p=0.392 | 40.4 |
| Bw: AN on OI | 4 | 0.045 (0.011; 0.080) | 0.009 | 5 (3) | 0.204 | 0.048 (0.002; 0.094) | 0.042 | -0.056 (-0.500; 0.388) | 0.827 | 0.696 | <0.001 | GT; p=0.299 | 31.9 |
| Fw: OI on ANX | 32 | -0.110 (-0.672; 0.453) | 0.702 | 39 (31) | 0.163 | -0.129 (-0.920; 0.661) | 0.749 | 0.767 (-2.36; 3.89) | 0.634 | 0.578 | NR ^b^ | GT; p=0.164 | 40.6 |
| Bw: ANX on OI | 0 | NR ^c^ | NR ^c^ | NR ^c^ | NR ^c^ | NR ^c^ | NR ^c^ | NR ^c^ | NR ^c^ | NR ^c^ | NR ^c^ | NR ^c^ | NR ^c^ |
| Fw: OI on ASD | 33 | 0.471 (0.192; 0.750) | 0.001 | 82 (32) | <0.001 | 0.340 (-0.142; 0.822) | 0.167 | 2.36 (0.281; 4.44) | 0.034 | 0.078 | <0.001 | DT; p=0.200 | 40.3 |
| Bw: ASD on OI | 0 | NR ^c^ | NR ^c^ | NR ^c^ | NR ^c^ | NR ^c^ | NR ^c^ | NR ^c^ | NR ^c^ | NR ^c^ | NR ^c^ | NR ^c^ | NR ^c^ |
| Fw: OI on BD | 34 | 0.201 (0.016; 0.386) | 0.033 | 136 (33) | <0.001 | 0.137 (-0.187; 0.461) | 0.407 | 1.88 (0.135; 3.63) | 0.043 | 0.063 | <0.001 | DT; p=0.096 | 40.4 |
| Bw: BD on OI | 36 | 0.042 (0.028; 0.057) | <0.001 | 147 (35) | <0.001 | 0.020 (-0.005; 0.046) | 0.116 | 0.068 (-0.099; 0.235) | 0.431 | 0.763 | <0.001 | DT; p=0.174 | 39.2 |
| Fw: OI on MDD | 33 | -0.322 (-0.502; -0.143) | <0.001 | 83 (32) | <0.001 | -0.125 (-0.420; 0.171) | 0.408 | 0.309 (-1.10; 1.72) | 0.671 | 0.376 | <0.001 | DT; p=0.849 | 40.3 |
| Bw: MDD on OI | 0 | NR ^c^ | NR ^c^ | NR ^c^ | NR ^c^ | NR ^c^ | NR ^c^ | NR ^c^ | NR ^c^ | NR ^c^ | NR ^c^ | NR ^c^ | NR ^c^ |
| Fw: OI on OCD | 33 | 0.600 (-0.078; 1.28) | 0.083 | 34 (32) | 0.362 | 0.530 (-0.453; 1.51) | 0.290 | -2.15 (-5.44; 1.13) | 0.208 | 0.103 | NR ^b^ | GT; p=0.361 | 40.3 |
| Bw: OCD on OI | 0 | NR ^c^ | NR ^c^ | NR ^c^ | NR ^c^ | NR ^c^ | NR ^c^ | NR ^c^ | NR ^c^ | NR ^c^ | NR ^c^ | NR ^c^ | NR ^c^ |
| Fw: OI on PTSD | 33 | -0.051 (-0.347; 0.245) | 0.736 | 53 (32) | 0.012 | -0.271 (-0.717; 0.176) | 0.235 | -0.407 (-2.26; 1.44) | 0.669 | 0.702 | NR ^b^ | DT; p=0.934 | 40.3 |
| Bw: PTSD on OI | 0 | NR ^c^ | NR ^c^ | NR ^c^ | NR ^c^ | NR ^c^ | NR ^c^ | NR ^c^ | NR ^c^ | NR ^c^ | NR ^c^ | NR ^c^ | NR ^c^ |
| Fw: OI on SZ | 34 | -0.187 (-0.344; -0.030) | 0.020 | 278 (33) | <0.001 | -0.176 (-0.489; 0.138) | 0.272 | 0.202 (-2.07; 2.48) | 0.863 | 0.734 | <0.001 | DT; p=0.818 | 40.4 |
| Bw: SZ on OI | 176 | -0.010 (-0.017; -0.003) | 0.008 | 512 (175) | <0.001 | -0.008 (-0.021; 0.005) | 0.228 | 0.005 (-0.044; 0.055) | 0.830 | 0.526 | <0.001 | DT; p=0.065 | 45.6 |

Abbreviations: Fw: forward analysis; Bw: backward analysis; OI: occupational income; ADHD: attention deficit hyperactivity disorder; AN: anorexia nervosa; ANX: anxiety disorder; ASD: autism spectrum disorder; BD: bipolar disorder; MDD: major depressive disorder; OCD: obsessive-compulsive disorder; PTSD: post-traumatic stress disorder; SZ: schizophrenia; MR: mendelian randomization; SNP: single nucleotide polymorphism; IVW: inverse variance weighted (fixed effect); B: effect estimates are log-odds for binary traits (i.e., for mental illnesses) and unstandardized regression coefficient for continuous traits (i.e., for occupational income); 95% CI: 95% confidence interval; Q: Cochran’s Q measure of heterogeneity; df: degree of freedom; WM: weighted median; DT: distortion test; GT: global test.

Legend:

P-value threshold <5e-8

^a^ The Mendelian randomization pleiotropy residual sum and outlier (MR-PRESSO) test identifies possible bias from horizontal pleiotropy. The test consists of three parts, (1) the MR-PRESSO global test which detects horizontal pleiotropy, (2) the outlier corrected causal estimate which corrects for the detected horizontal pleiotropy and (3) the MR-PRESSO distortion test which estimates if the causal estimate is significantly different (at p<0.05) after adjustment for outliers. We conduct all three stages (with the argument NbDistribution=1000, namely using1000 simulation form the null distribution to compute empirical p-values) and present the outlier adjusted causal estimates (OACE) when both global and distortion tests are significant.

^b^ We did not run Steiger Test if none of the MR analysis resulted significant (NR: not reported in the cell).

^c^ Not enough SNP to perform MR (NR: not reported in the cell).

#### Plots - Forward analyses

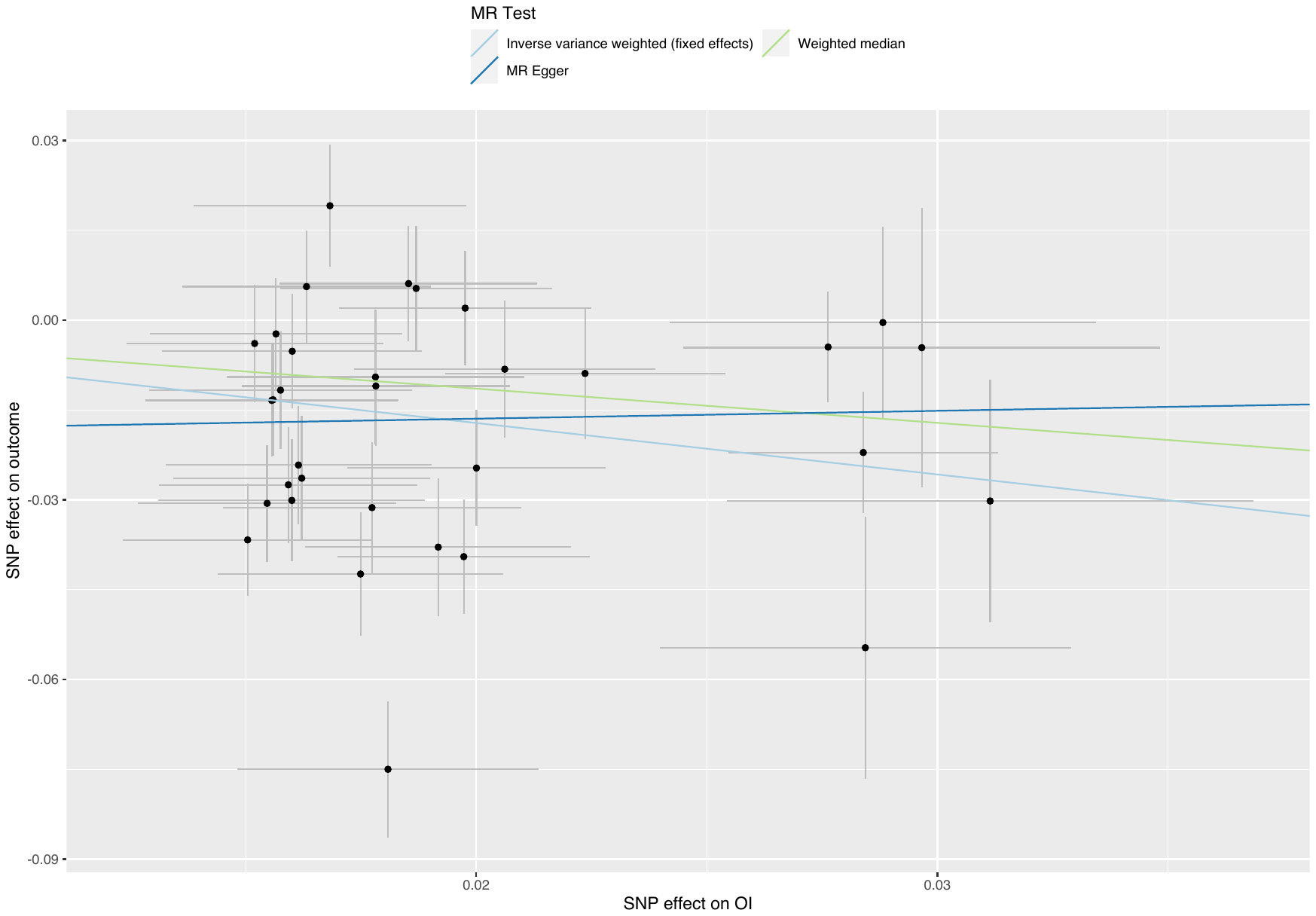

##### Supplementary Figure 56: scatterplot of occupational income against ADHD

Abbreviations: MR: Mendelian randomization; SNP: single nucleotide polymorphism; OI: occupational income; ADHD: attention deficit hyperactivity disorder.

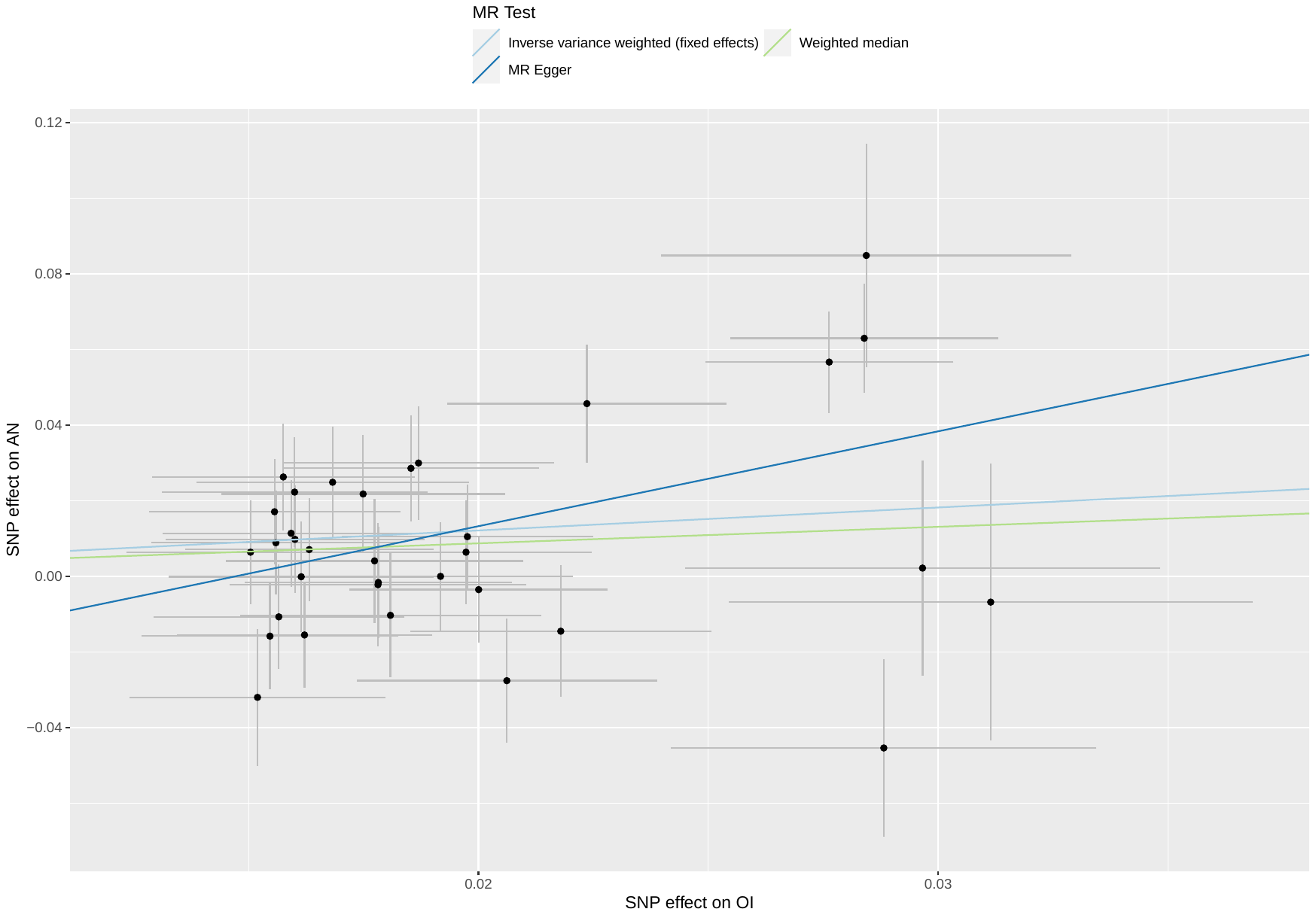

##### Supplementary Figure 57: scatterplot of occupational income against AN

Abbreviations: MR: Mendelian randomization; SNP: single nucleotide polymorphism; OI: occupational income; AN: anorexia nervosa.

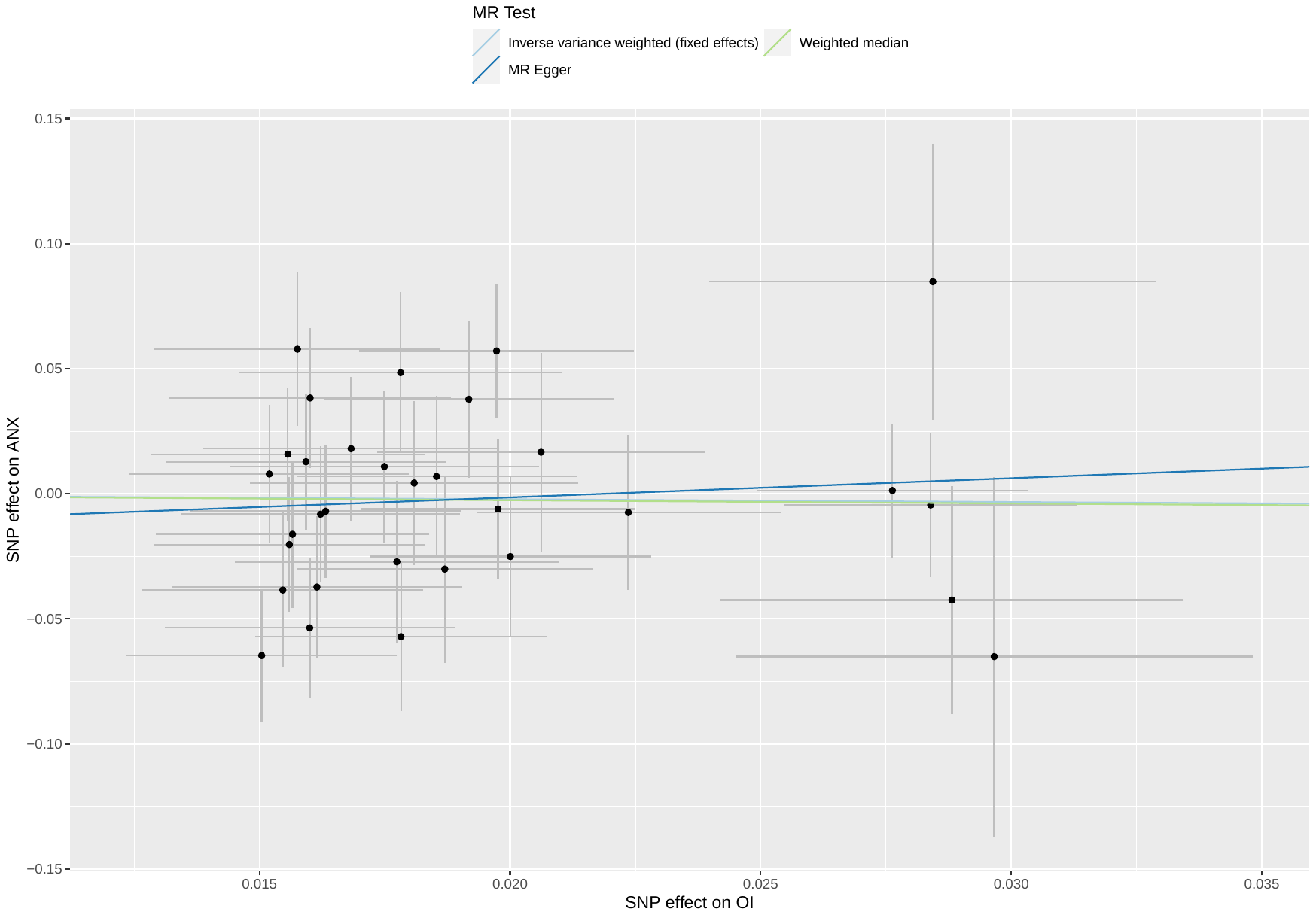

##### Supplementary Figure 58: scatterplot of occupational income against ANX

Abbreviations: MR: Mendelian randomization; SNP: single nucleotide polymorphism; OI: occupational income; ANX: anxiety disorders.

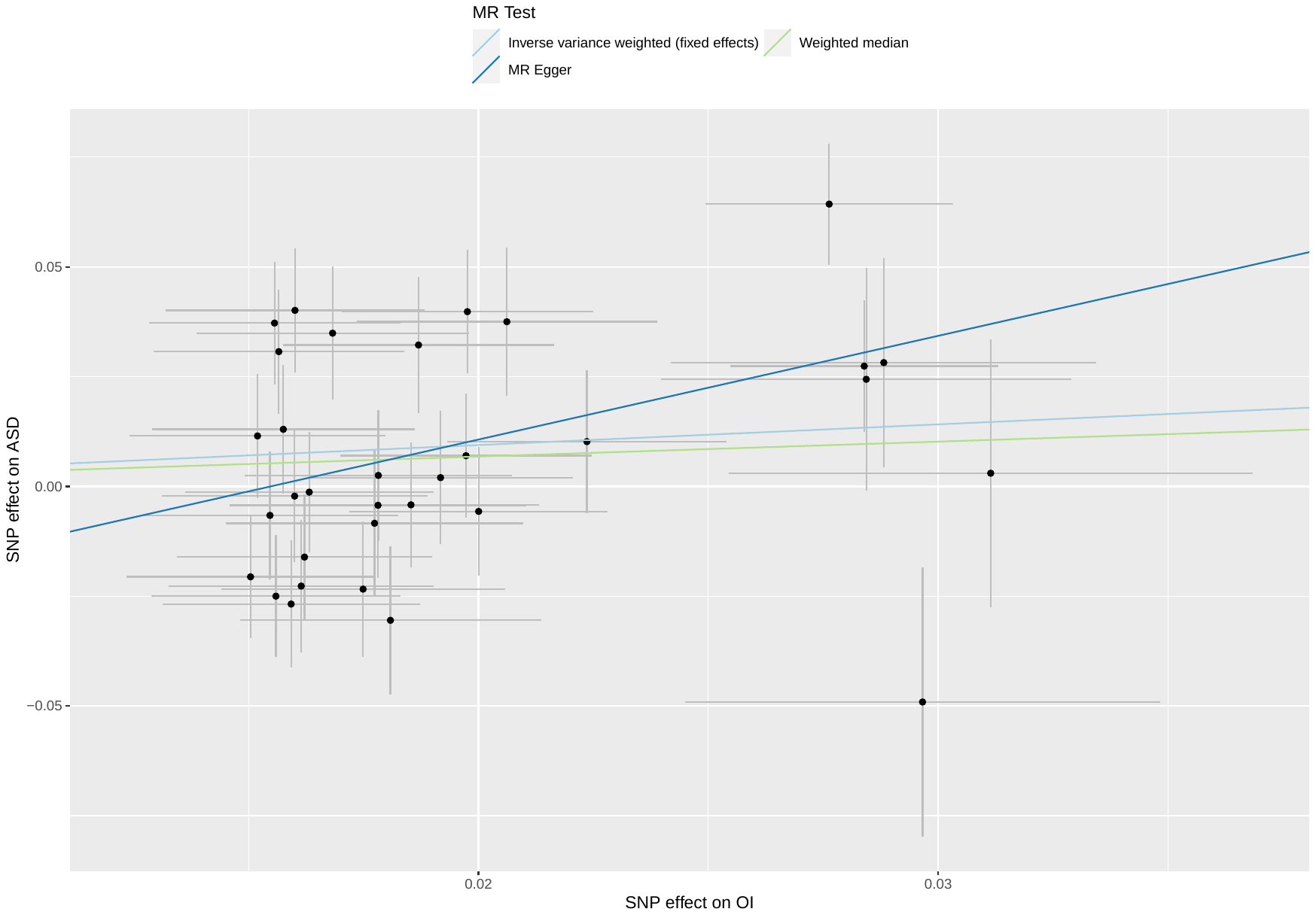

##### Supplementary Figure 59: scatterplot of occupational income against ASD

Abbreviations: MR: Mendelian randomization; SNP: single nucleotide polymorphism; OI: occupational income; ASD: autism spectrum disorders.

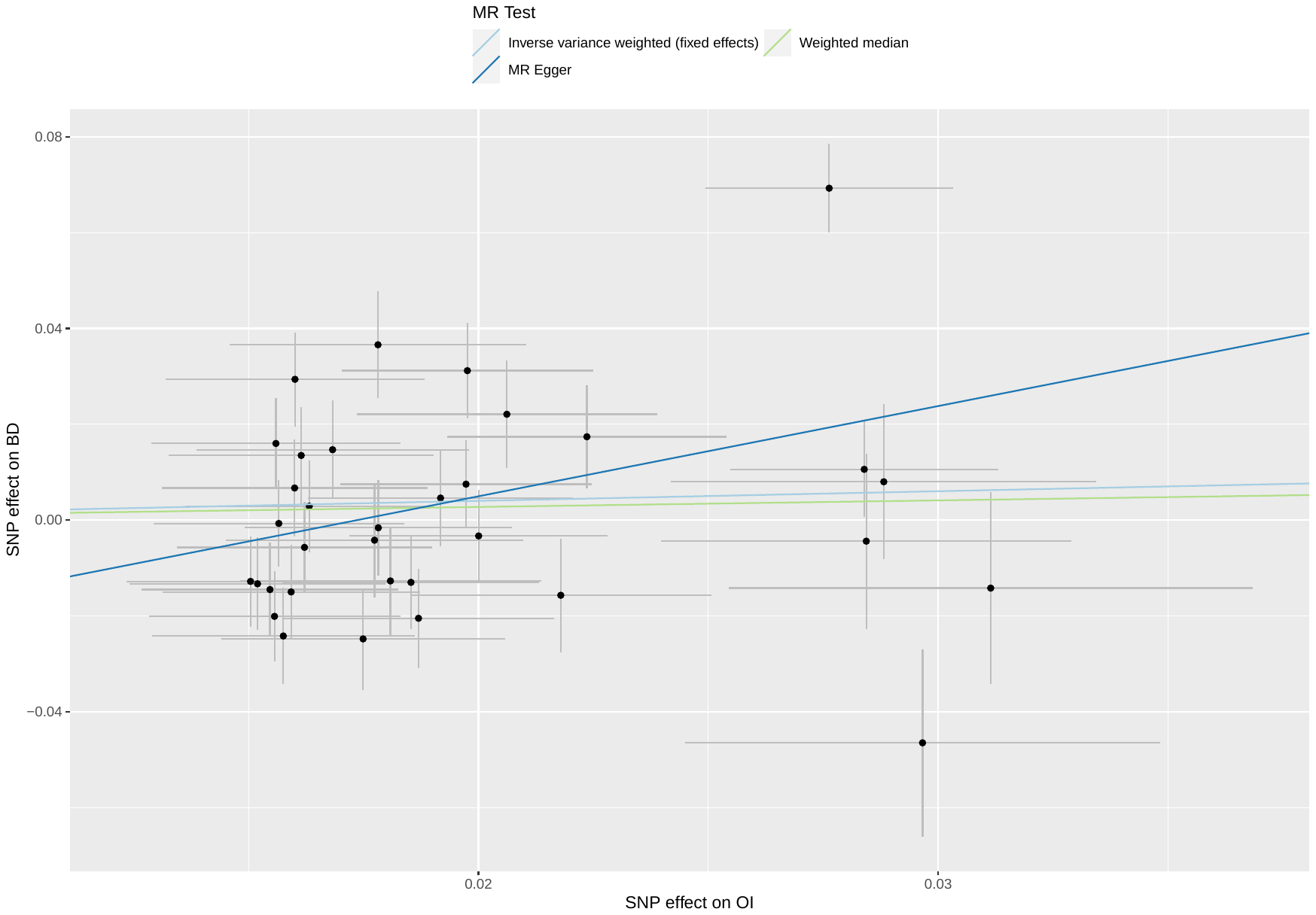

##### Supplementary Figure 60: scatterplot of occupational income against BD

Abbreviations: MR: Mendelian randomization; SNP: single nucleotide polymorphism; OI: occupational income; BD: bipolar disorder.

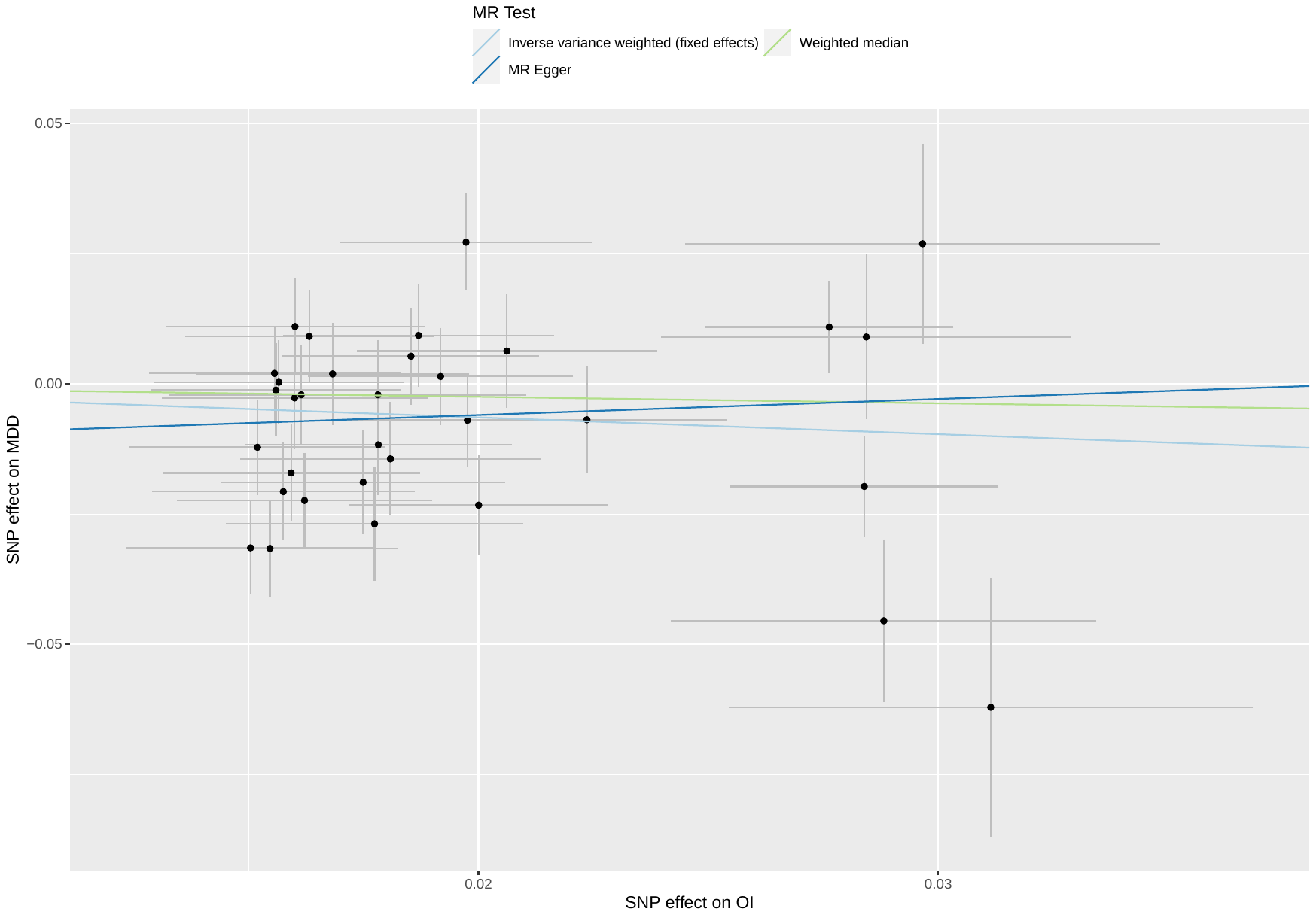

##### Supplementary Figure 61: scatterplot of occupational income against MDD

Abbreviations: MR: Mendelian randomization; SNP: single nucleotide polymorphism; OI: occupational income; MDD: major depressive disorder.

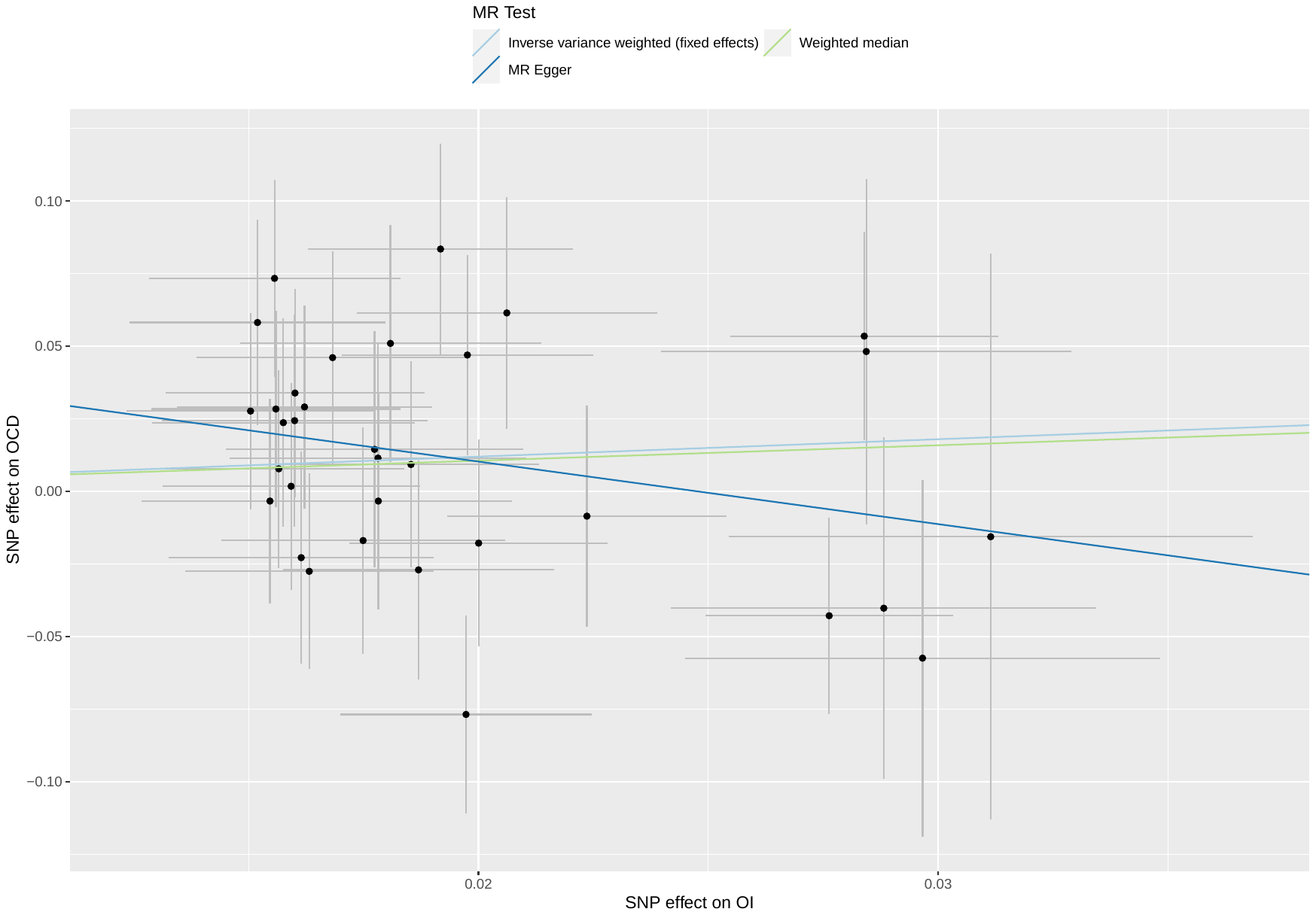

##### Supplementary Figure 62: scatterplot of occupational income against OCD

Abbreviations: MR: Mendelian randomization; SNP: single nucleotide polymorphism; OI: occupational income; OCD: obsessive-compulsive disorder.

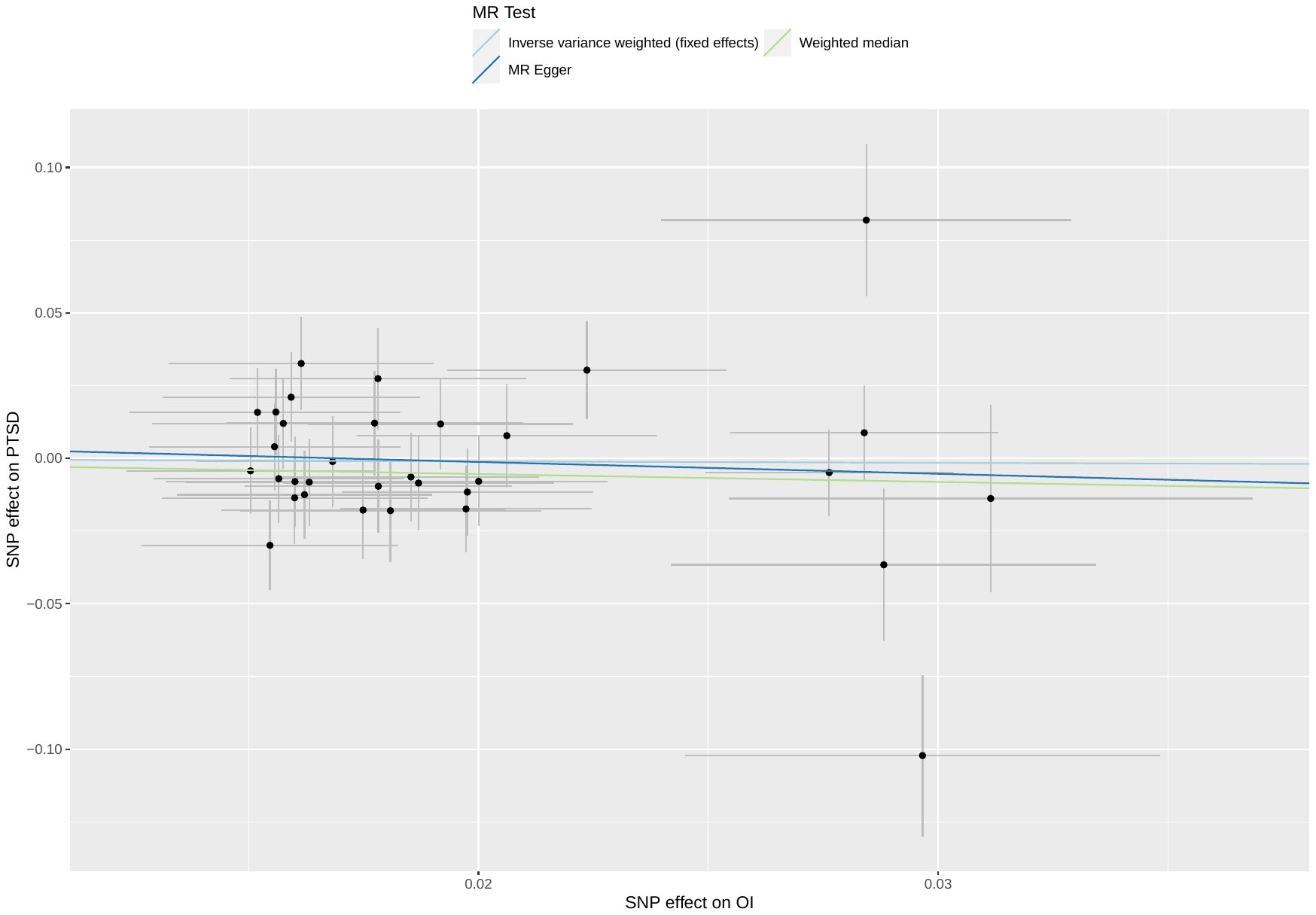

##### Supplementary Figure 63: scatterplot of occupational income against PTSD

Abbreviations: MR: Mendelian randomization; SNP: single nucleotide polymorphism; OI: occupational income; PTSD: post-traumatic stress disorder.

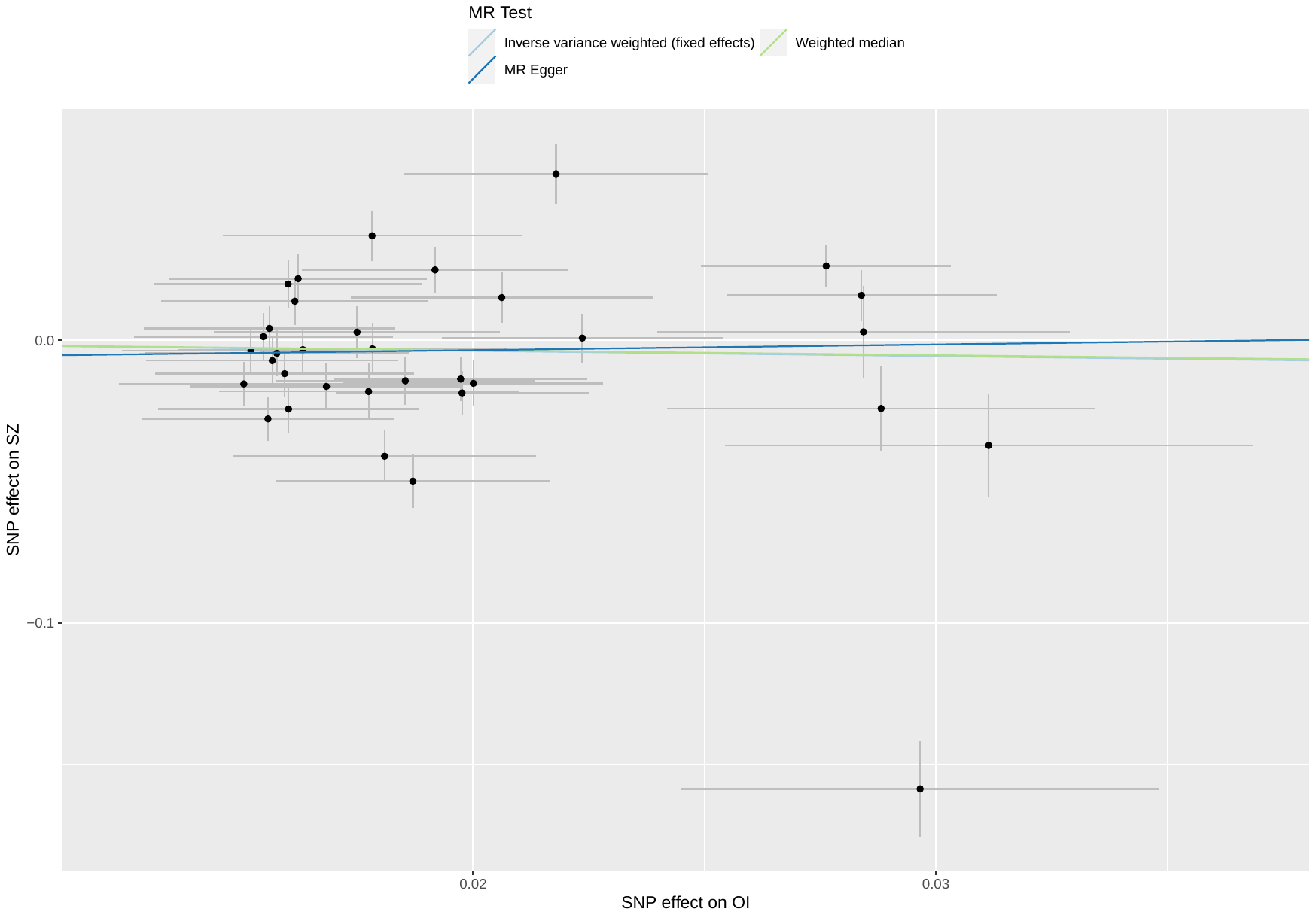

##### Supplementary Figure 64: scatterplot of occupational income against SZ

Abbreviations: MR: Mendelian randomization; SNP: single nucleotide polymorphism; OI: occupational income; SZ: schizophrenia.

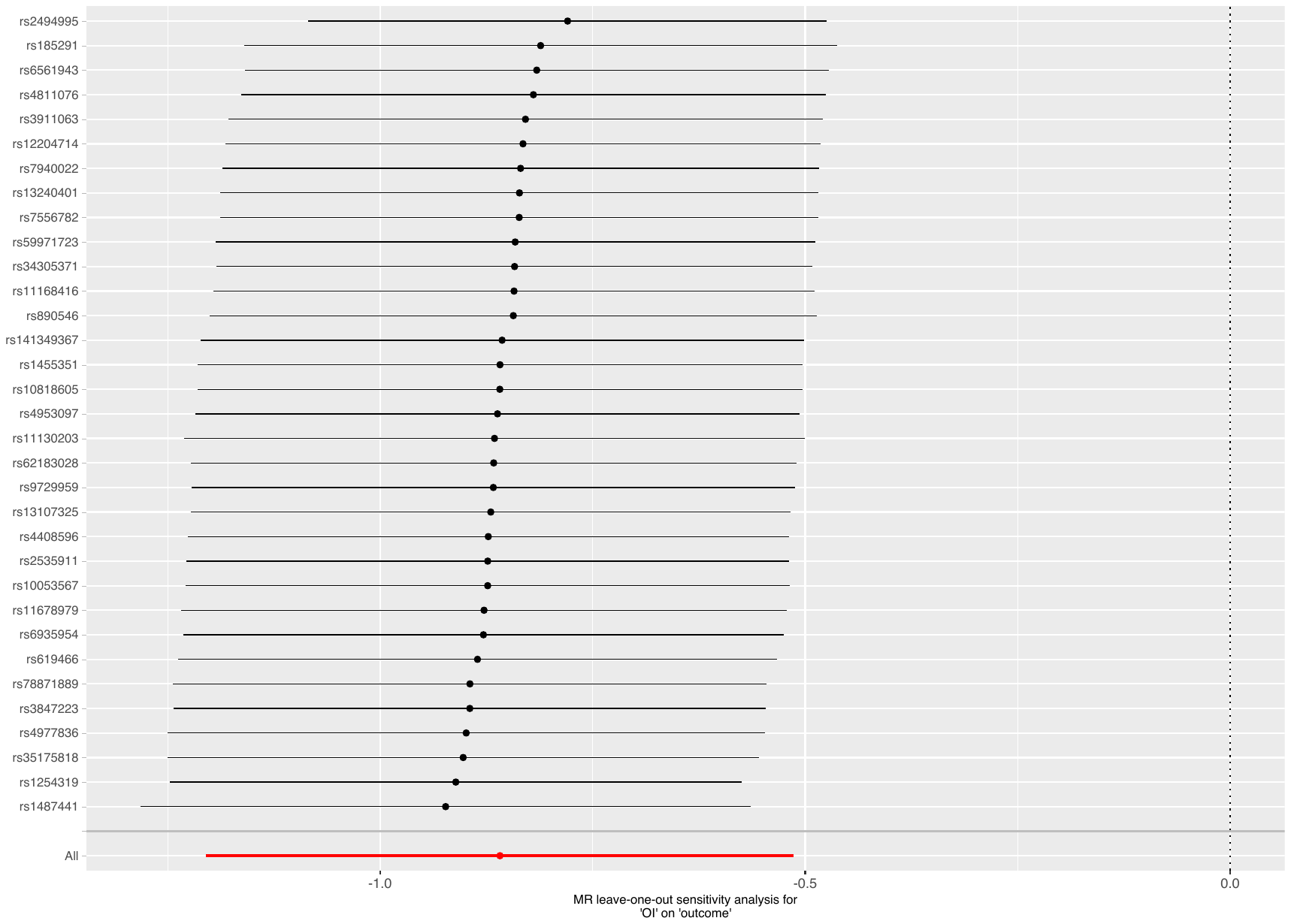

Supplementary Figure 65: leave-one-out analysis of occupational income against ADHD

Abbreviations: MR: Mendelian randomization; OI: occupational income; ADHD: attention deficit hyperactivity disorder.

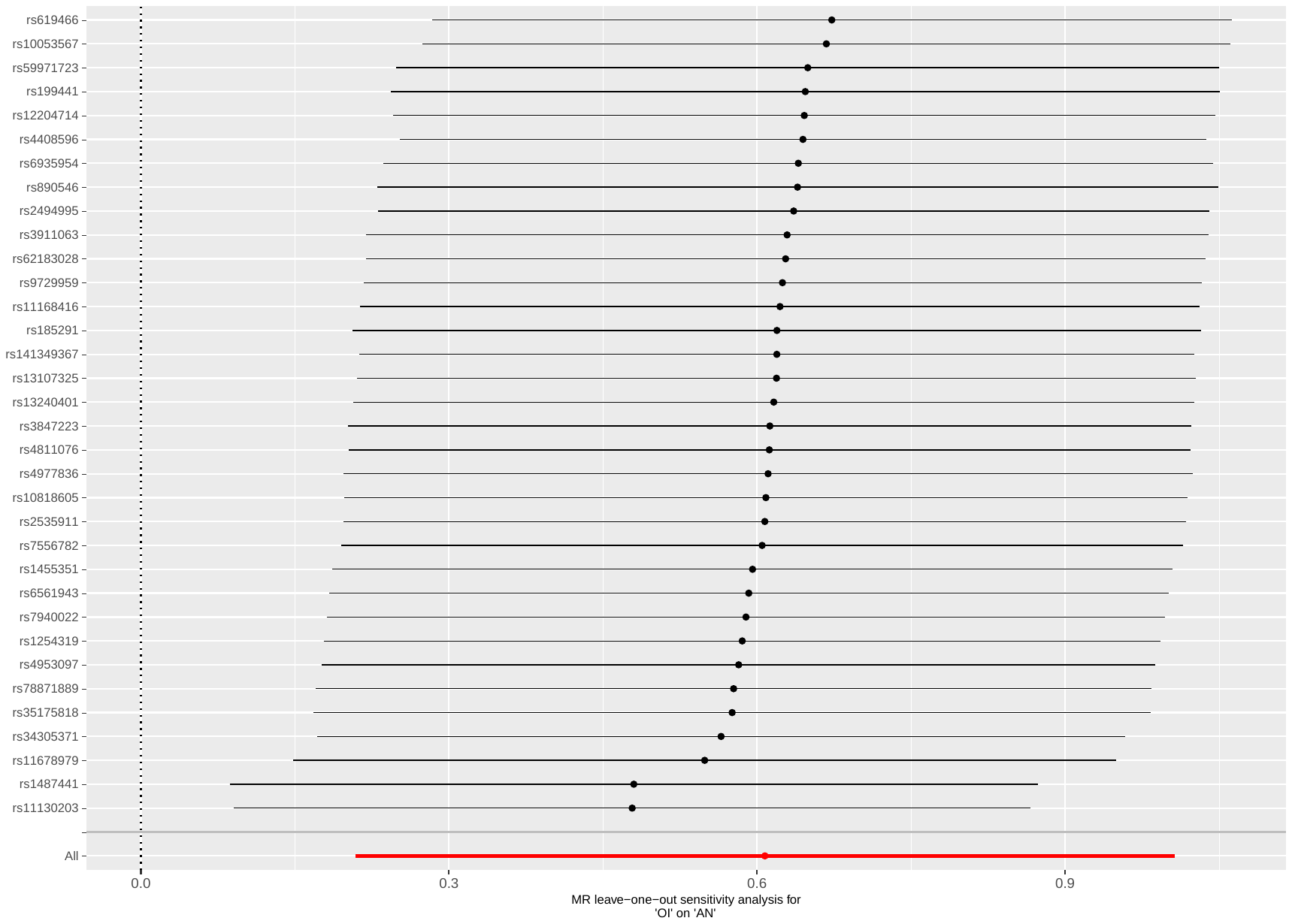

Supplementary Figure 66: leave-one-out analysis of occupational income against AN

Abbreviations: MR: Mendelian randomization; OI: occupational income; AN: anorexia nervosa.

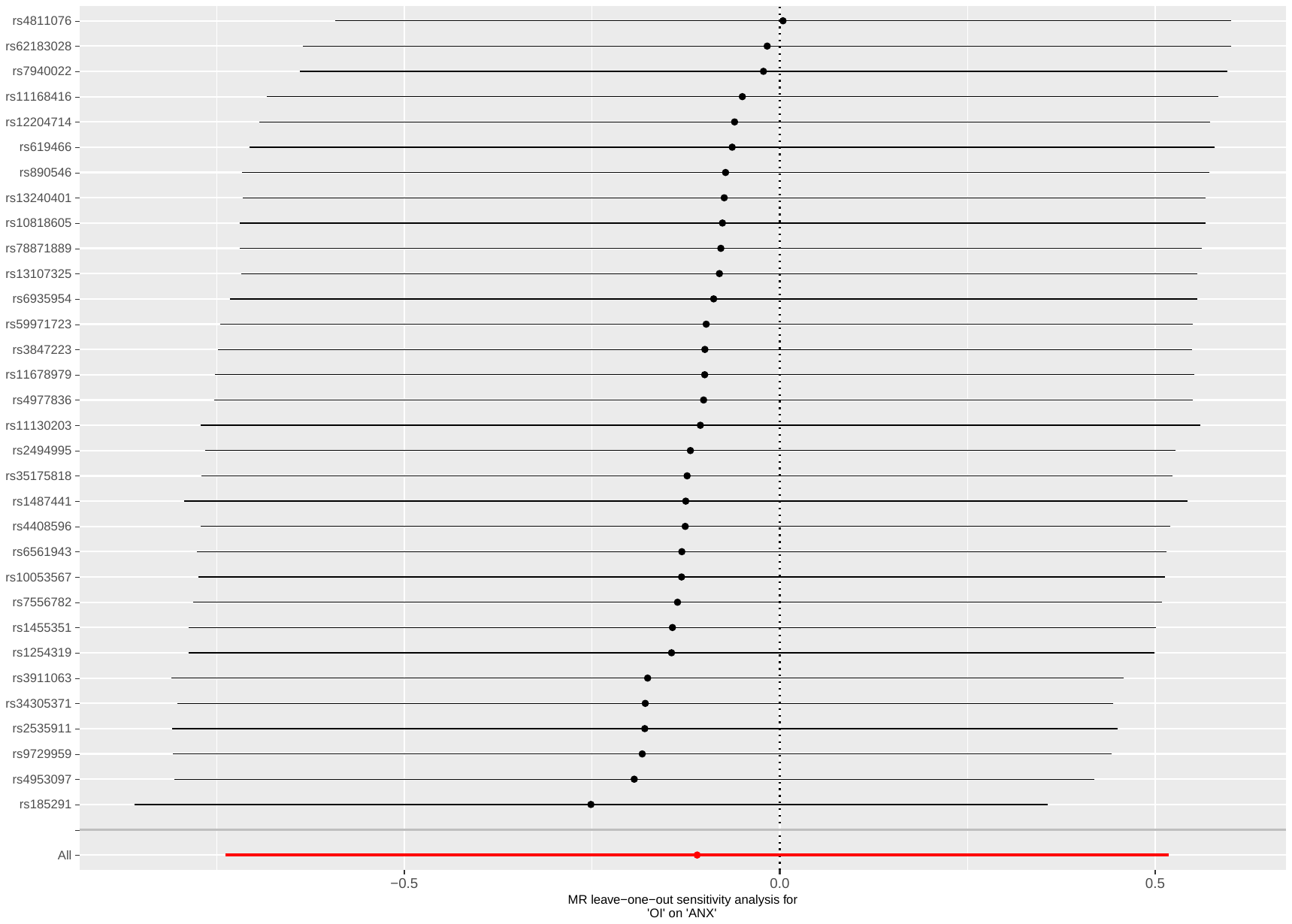

Supplementary Figure 67: leave-one-out analysis of occupational income against ANX

Abbreviations: MR: Mendelian randomization; OI: occupational income; ANX: anxiety disorders.

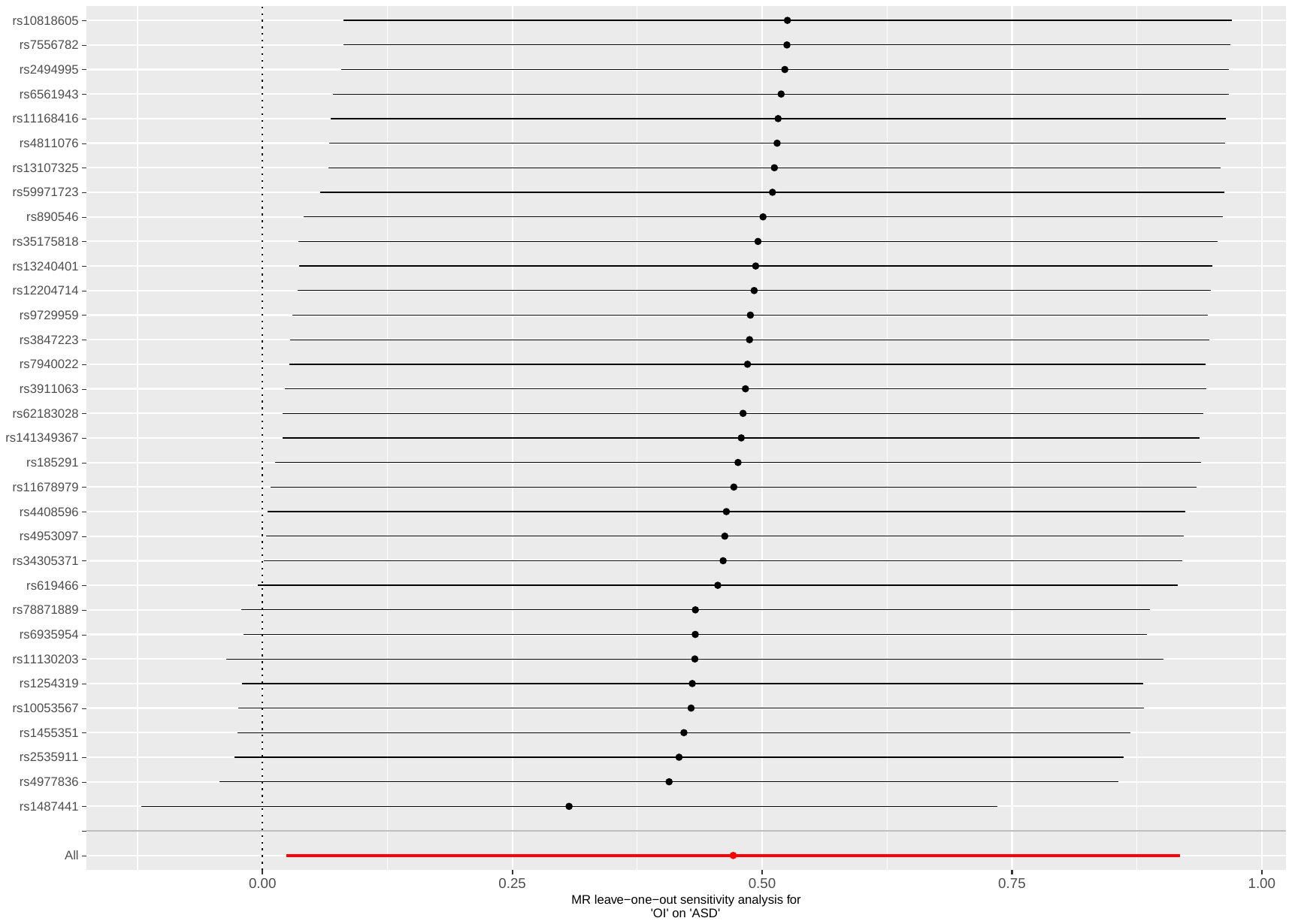

Supplementary Figure 68: leave-one-out analysis of occupational income against ASD

Abbreviations: MR: Mendelian randomization; OI: occupational income; ASD: autism spectrum disorders.

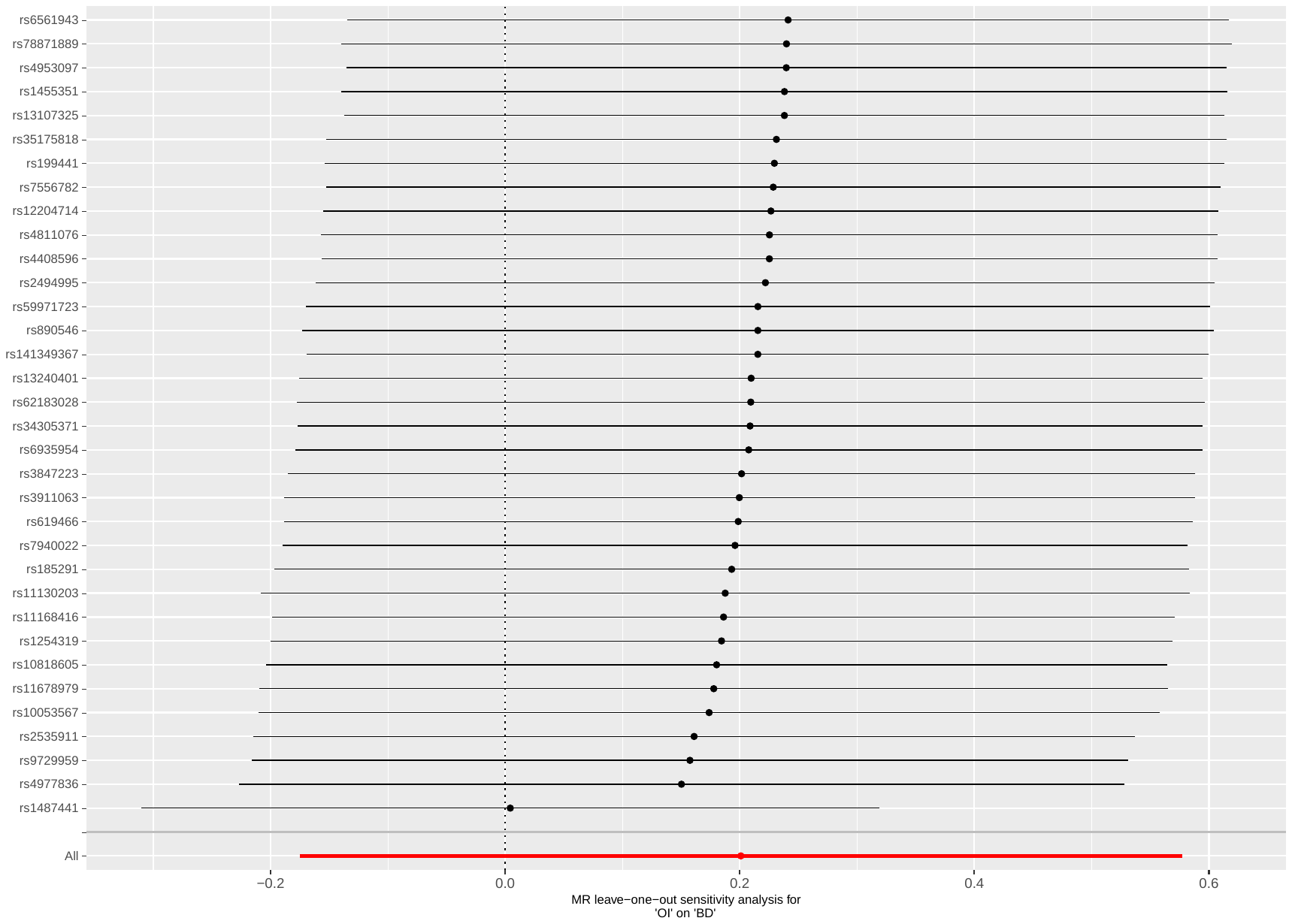

Supplementary Figure 69: leave-one-out analysis of occupational income against BD

Abbreviations: MR: Mendelian randomization; OI: occupational income; BD: bipolar disorder.

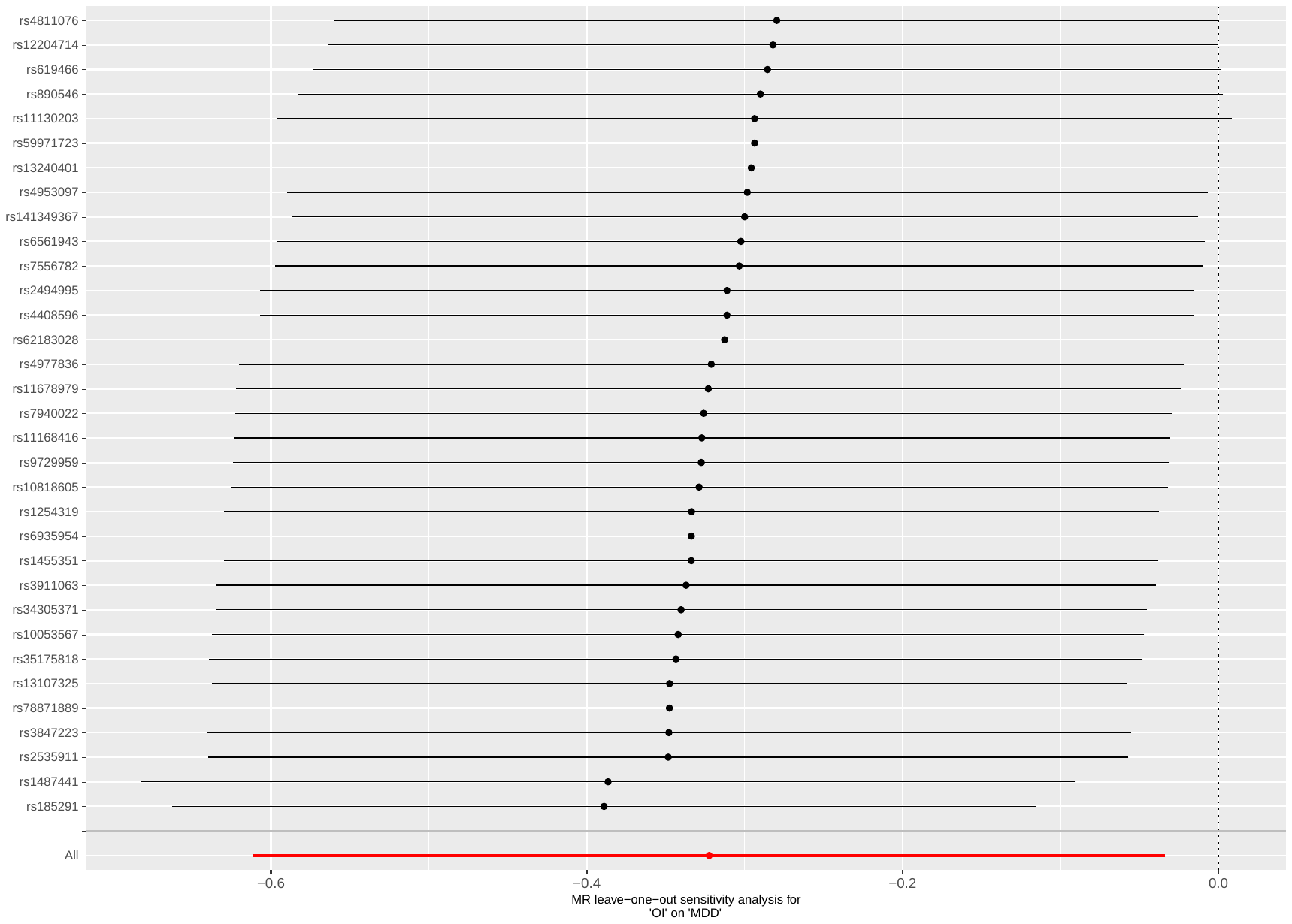

Supplementary Figure 70: leave-one-out analysis of occupational income against MDD

Abbreviations: MR: Mendelian randomization; OI: occupational income; MDD: major depressive disorder.

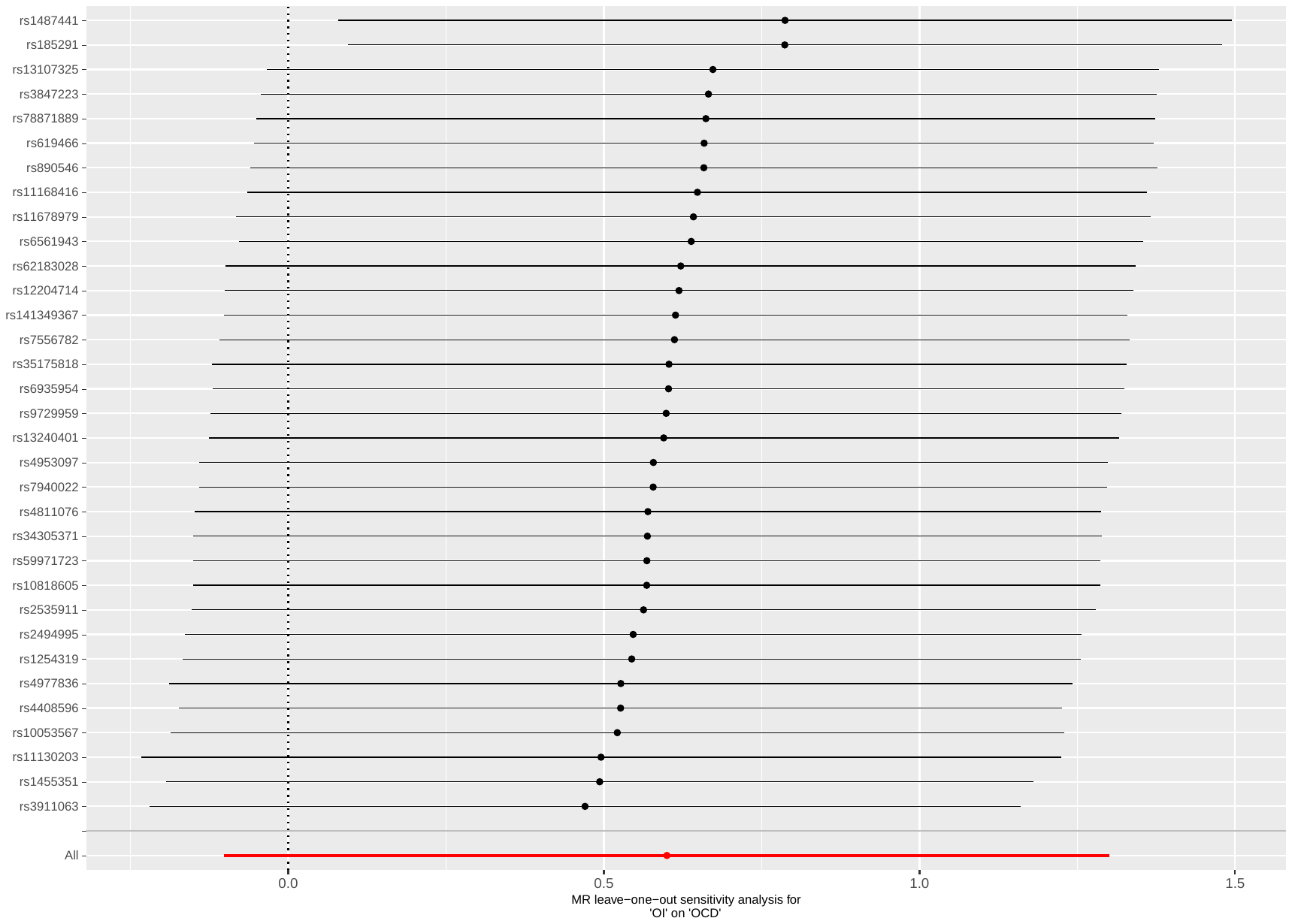

Supplementary Figure 71: leave-one-out analysis of occupational income against OCD

Abbreviations: MR: Mendelian randomization; OI: occupational income; OCD: obsessive-compulsive disorder.

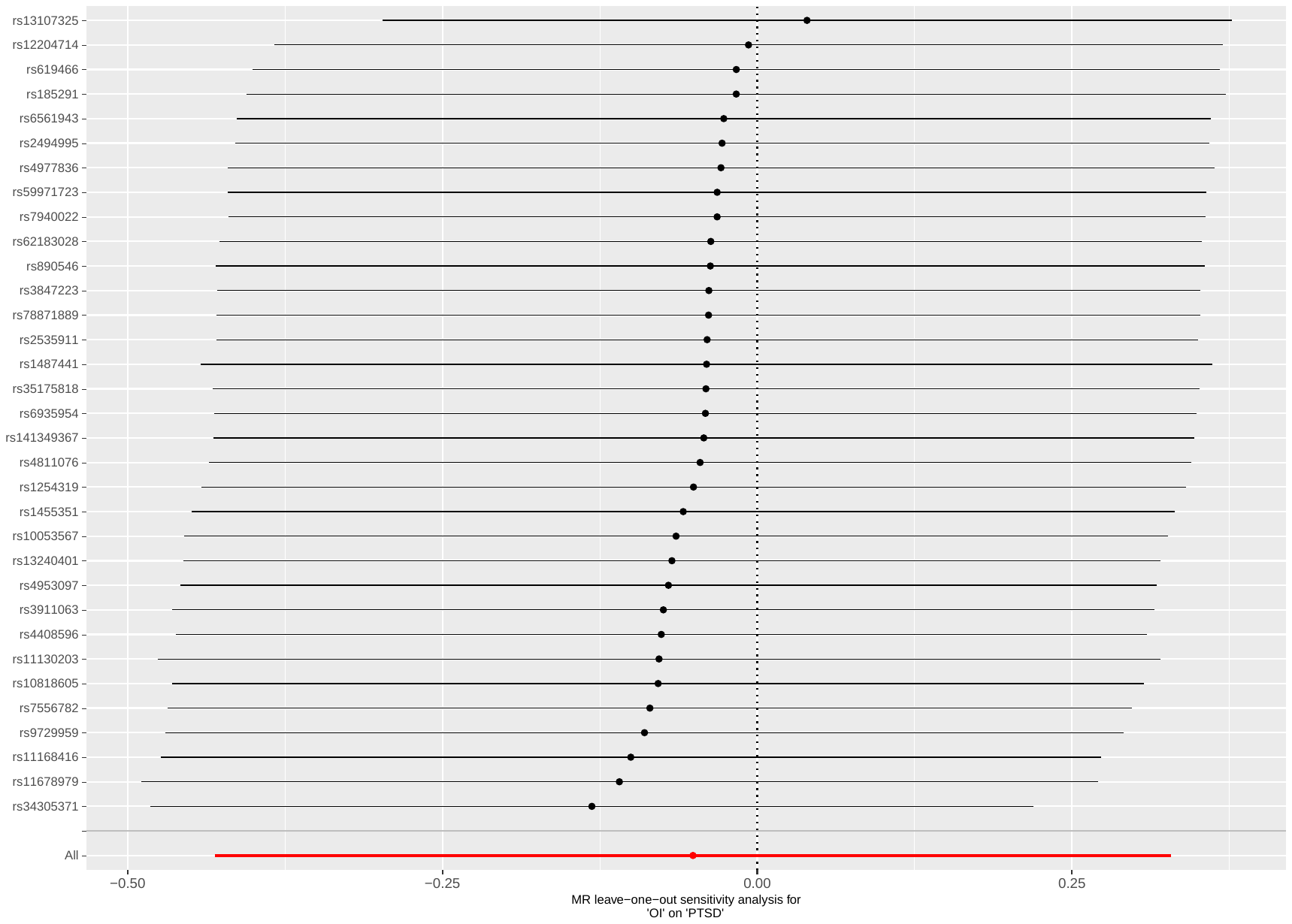

Supplementary Figure 72: leave-one-out analysis of occupational income against PTSD

Abbreviations: MR: Mendelian randomization; OI: occupational income; PTSD: post-traumatic stress disorder.

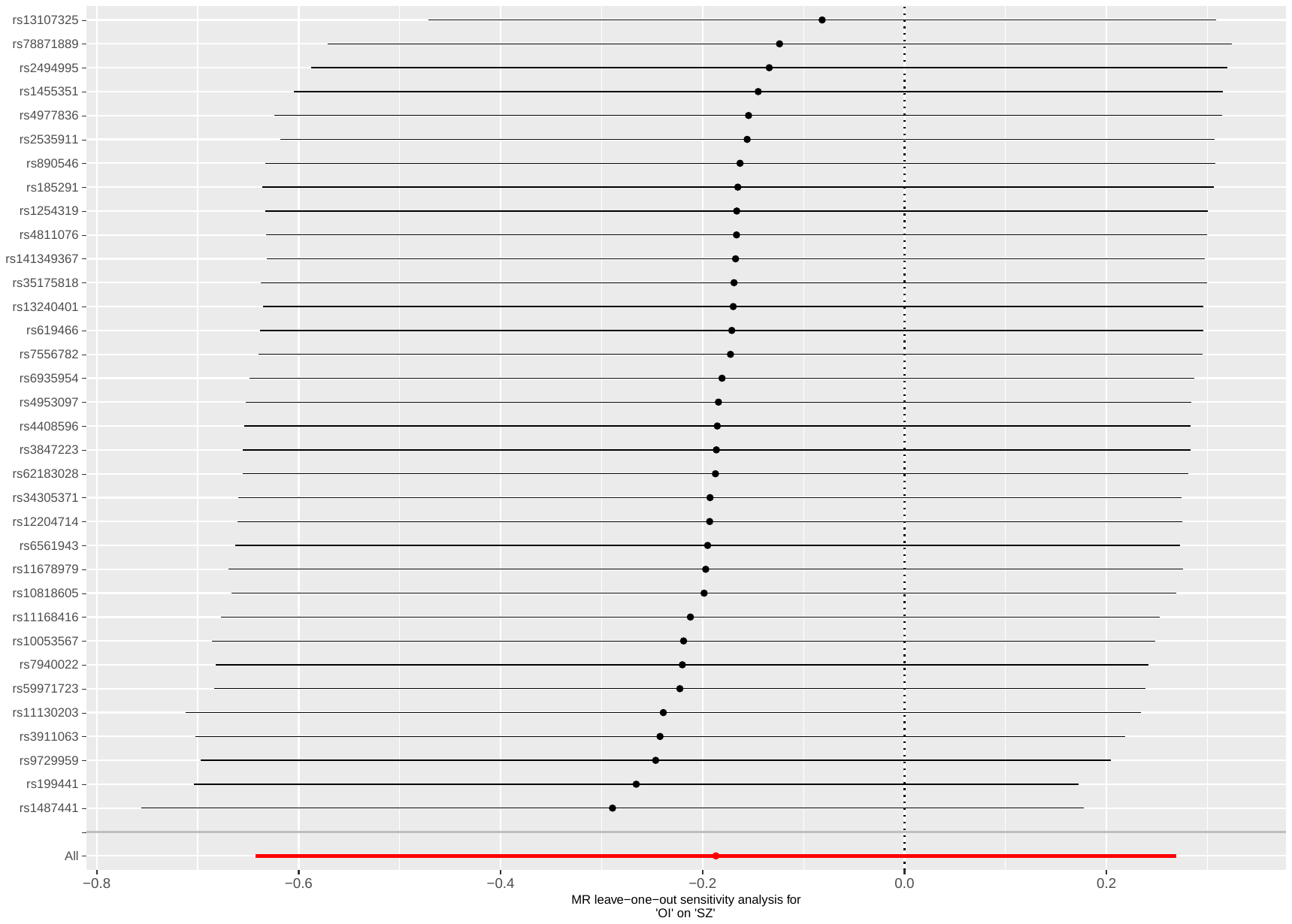

Supplementary Figure 73: leave-one-out analysis of occupational income against SZ

Abbreviations: MR: Mendelian randomization; OI: occupational income; SZ: schizophrenia.

#### Plots - Backward analyses

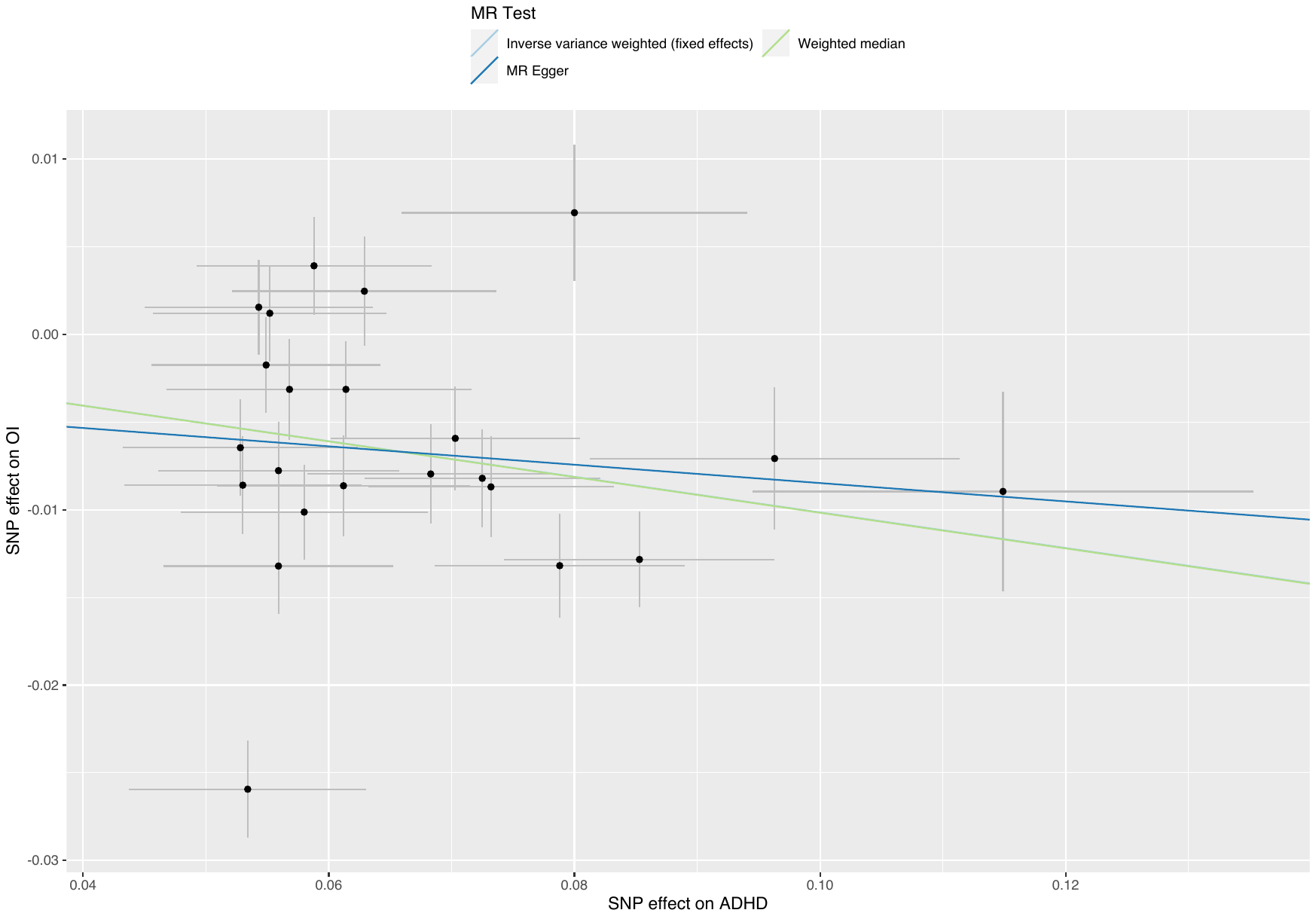

##### Supplementary Figure 74: scatterplot of ADHD against occupational income

Abbreviations: MR: Mendelian randomization; OI: occupational income; ADHD: attention deficit hyperactivity disorder.

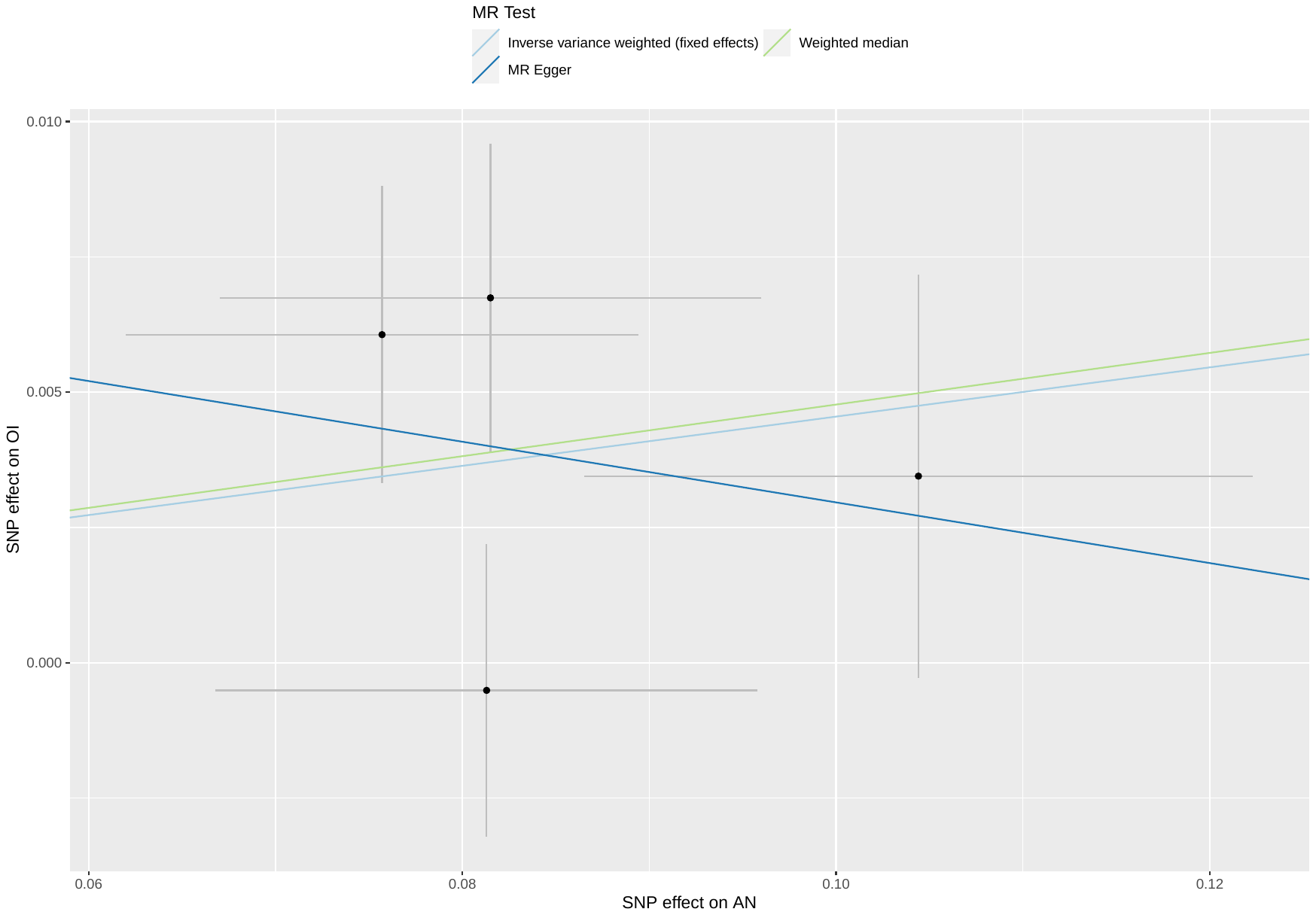

##### Supplementary Figure 75: scatterplot of AN against occupational income

Abbreviations: MR: Mendelian randomization; OI: occupational income; AN: anorexia nervosa.

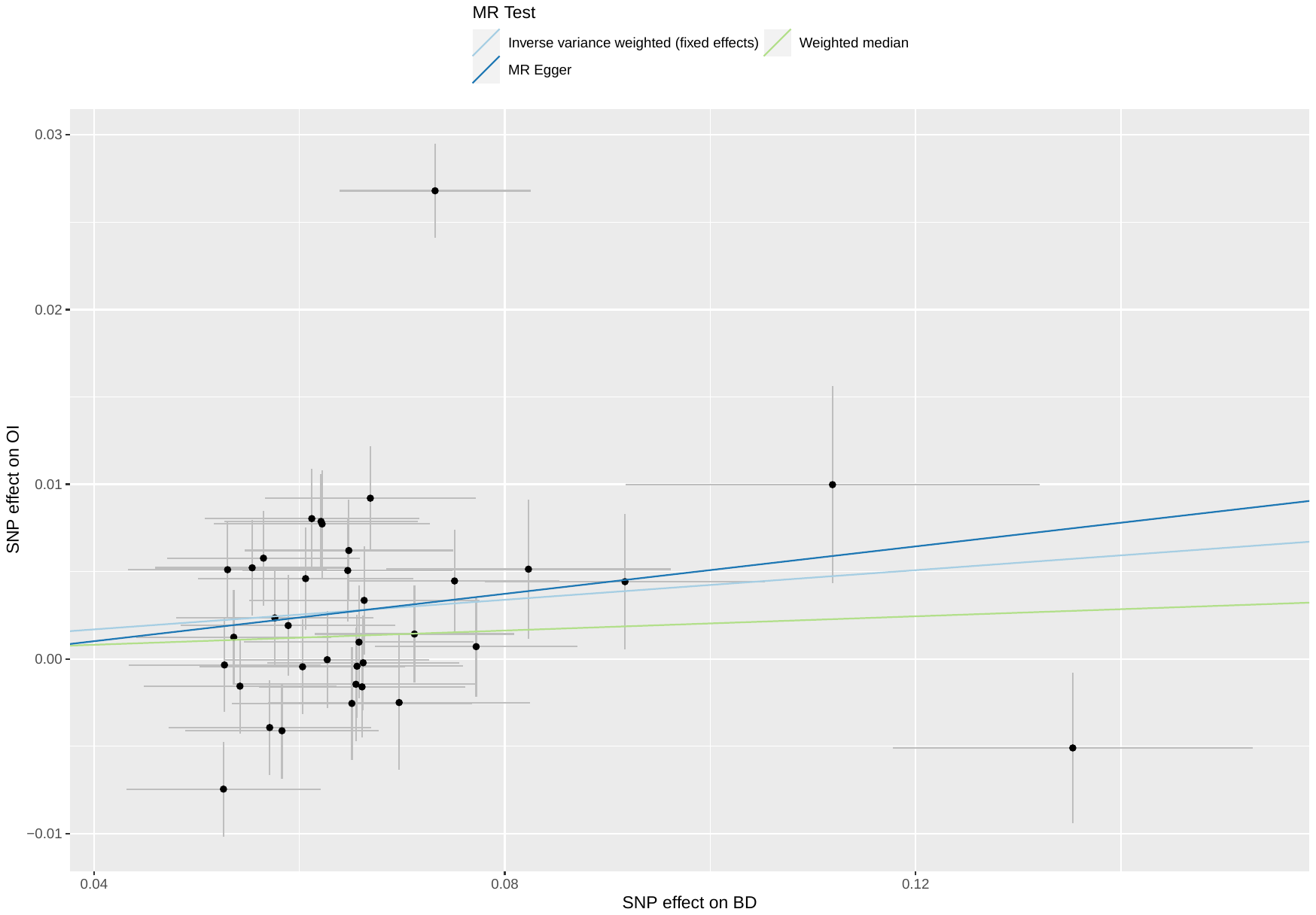

##### Supplementary Figure 76: scatterplot of BD against occupational income

Abbreviations: MR: Mendelian randomization; OI: occupational income; BD: bipolar disorder.

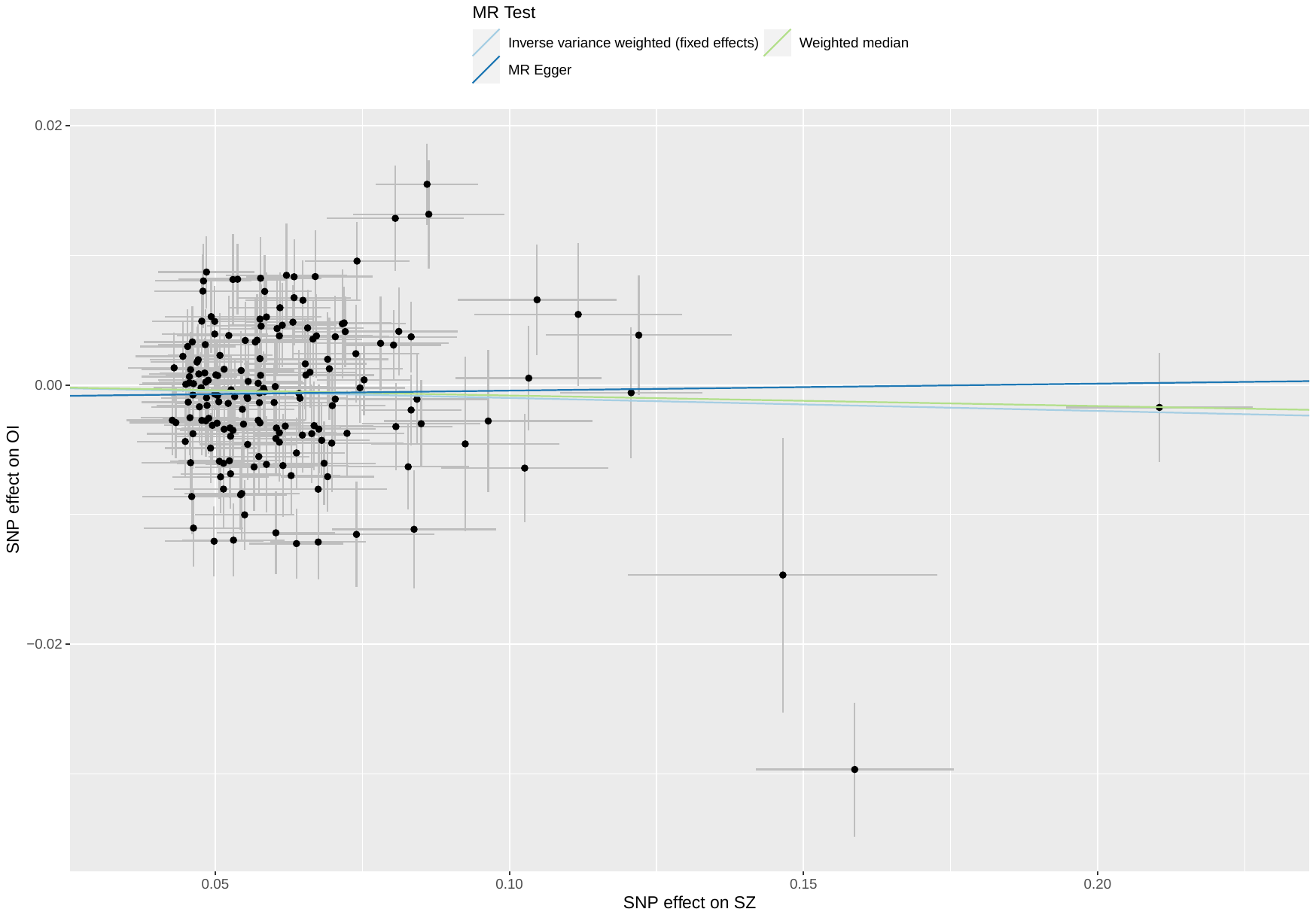

##### Supplementary Figure 77: scatterplot of SZ against occupational income

Abbreviations: MR: Mendelian randomization; OI: occupational income; SZ: schizophrenia.

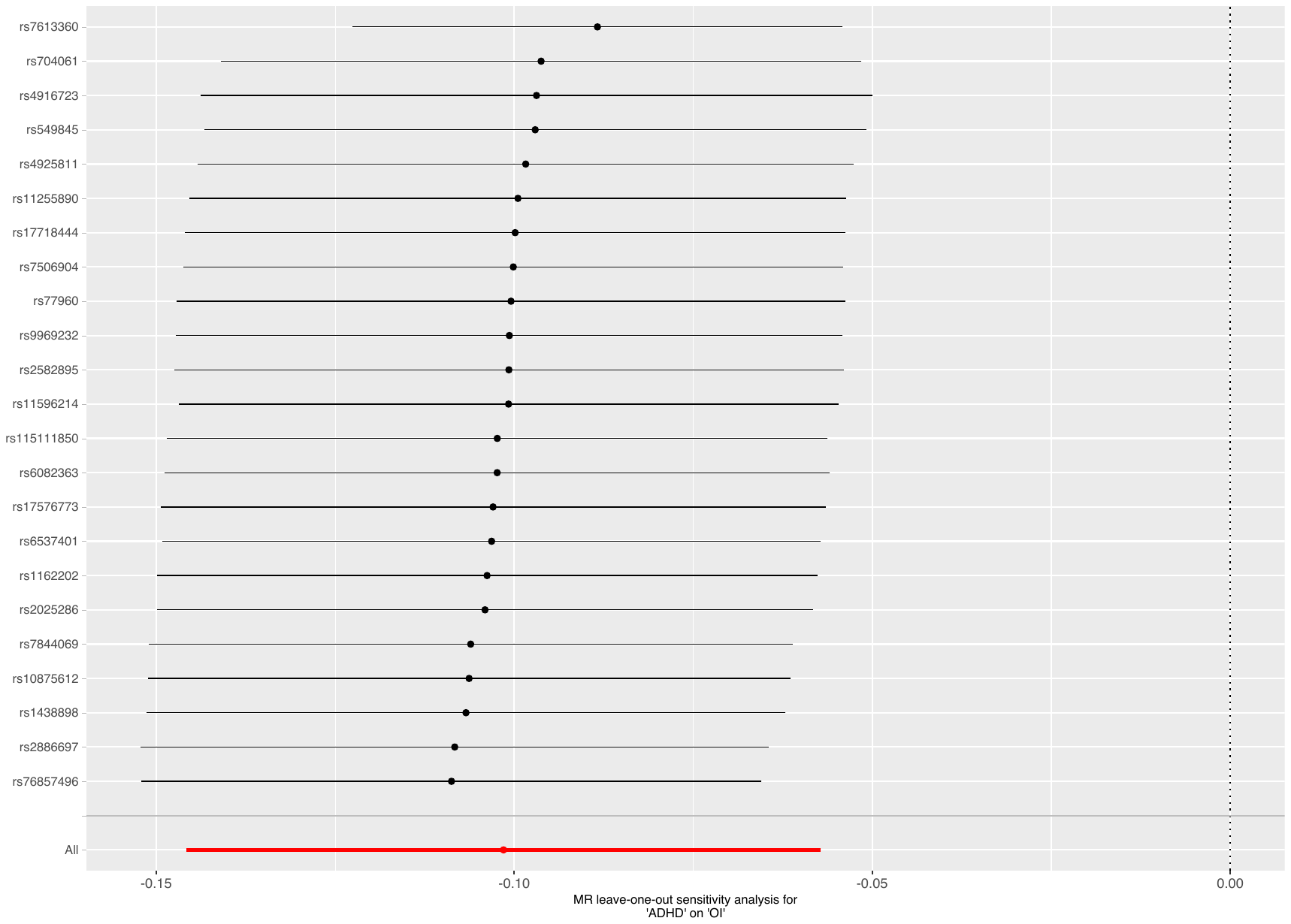

Supplementary Figure 78: leave-one-out analysis of ADHD against occupational income

Abbreviations: MR: Mendelian randomization; OI: occupational income; ADHD: attention deficit hyperactivity disorder.

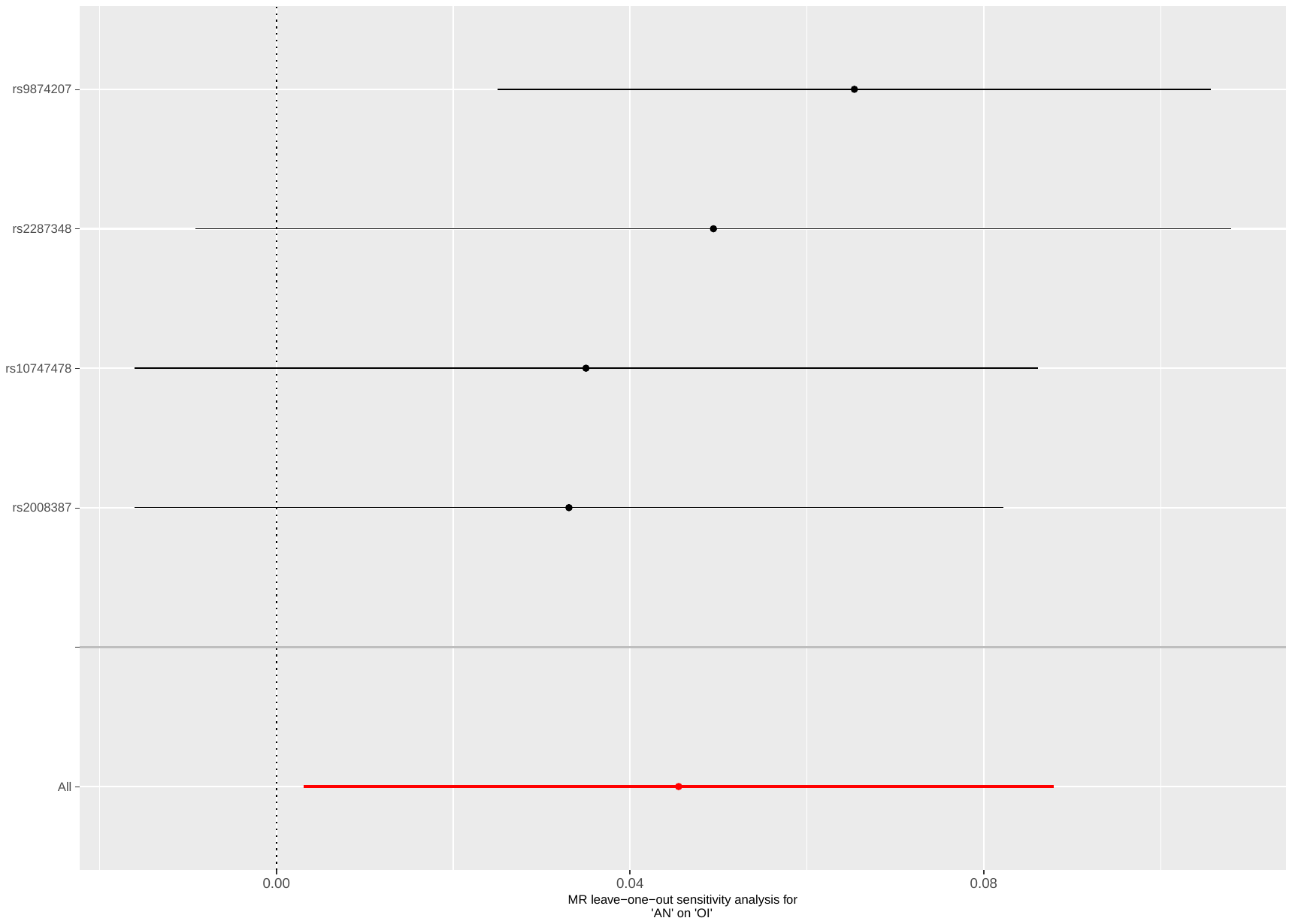

Supplementary Figure 79: leave-one-out analysis of AN against occupational income

Abbreviations: MR: Mendelian randomization; OI: occupational income; AN: anorexia nervosa.

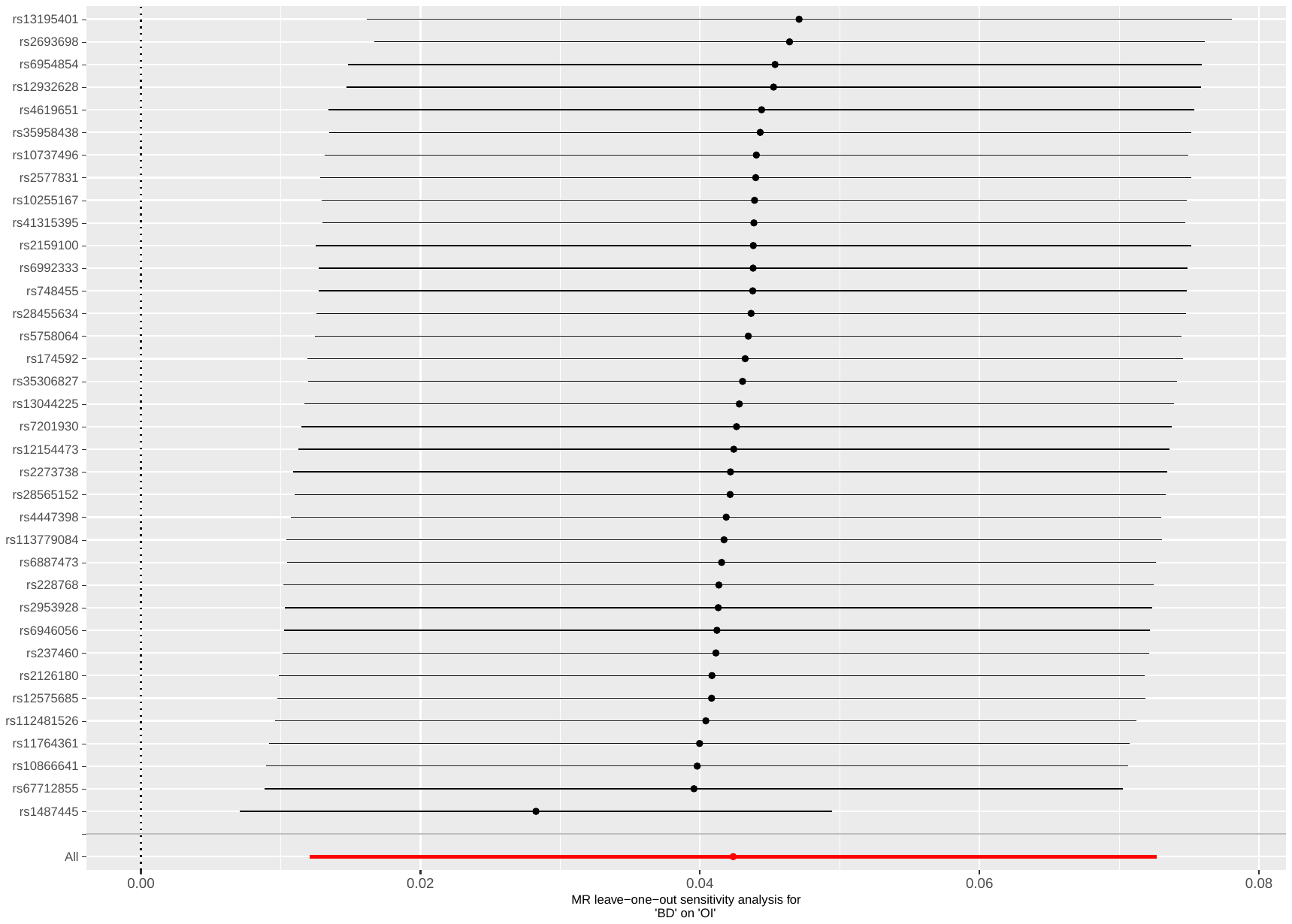

Supplementary Figure 80: leave-one-out analysis of BD against occupational income

Abbreviations: MR: Mendelian randomization; OI: occupational income; BD: bipolar disorder.

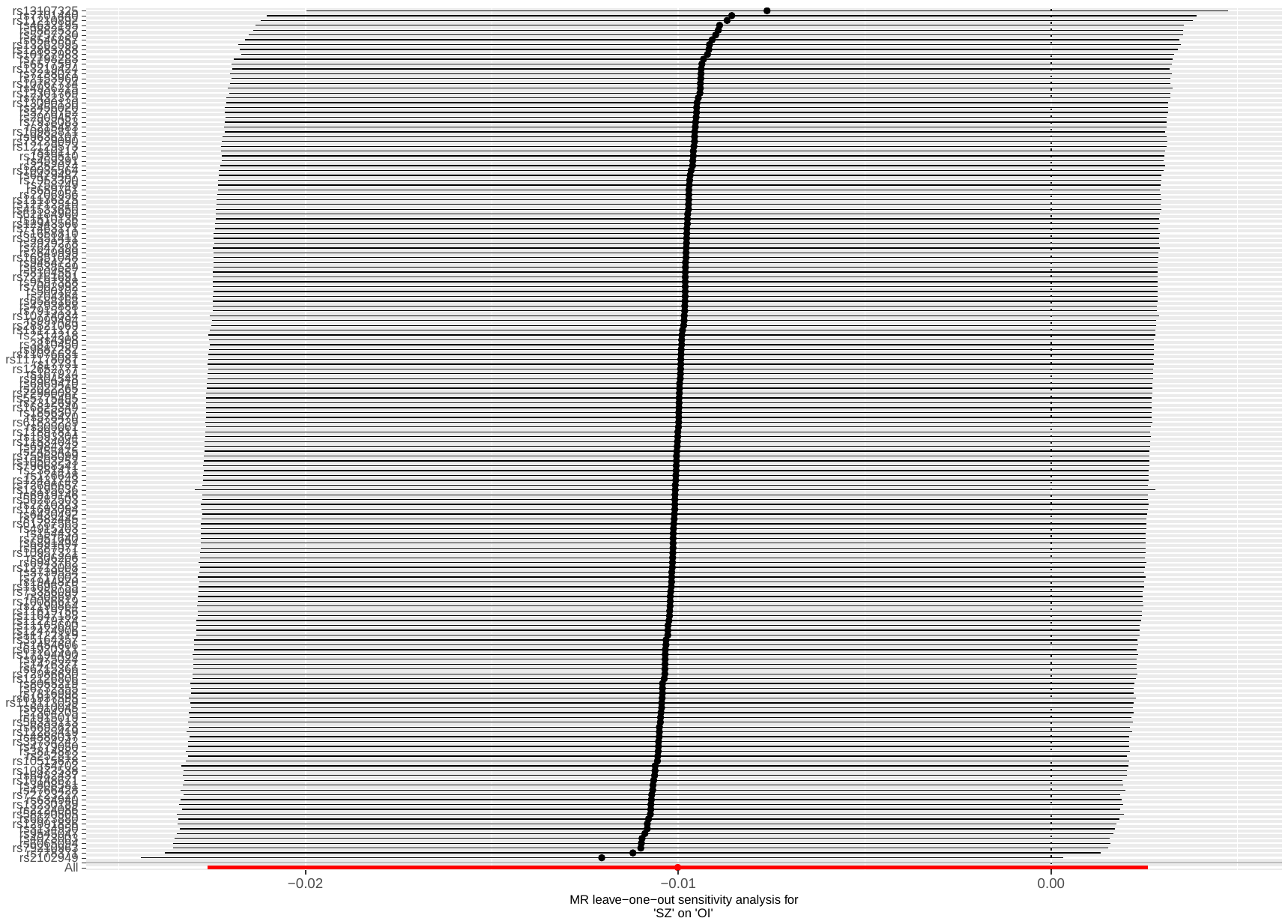

Supplementary Figure 81: leave-one-out analysis of SZ against occupational income

Abbreviations: MR: Mendelian randomization; OI: occupational income; SZ: schizophrenia.

#### Supplementary Table 15: CAUSE results of the relations between occupational income and mental illness

| **Model 1** | **Model 2** | **∆ ELPD** | **SE ∆ ELPD** | **z-score** | **p-value** |
| --- | --- | --- | --- | --- | --- |
| *Fw: OI on ADHD* | | | | | |
| Null  Null  Sharing | Sharing  Causal  Causal | -15.39  -21.30  -5.91 | 4.06  5.72  1.72 | -3.79  -3.72  -3.44 | <0.001  <0.001  <0.001 |
| *Bw: ADHD on OI* | | | | | |
| Null  Null  Sharing | Sharing  Causal  Causal | -11.42  -17.32  -5.91 | 3.31  5.04  1.76 | -3.45  -3.44  -3.36 | <0.001  <0.001  <0.001 |
| *Fw: OI on AN* | | | | | |
| Null  Null  Sharing | Sharing  Causal  Causal | -7.83  -10.22  -2.39 | 3.41  4.62  1.47 | -2.30  -2.21  -1.62 | 0.021  0.027  0.105 |
| *Bw: AN on OI* | | | | | |
| Null  Null  Sharing | Sharing  Causal  Causal | 0.29  0.17  -0.12 | 0.24  1.26  1.04 | 1.24  0.14  -0.12 | 0.215  0.889  0.904 |
| *Fw: OI on ANX* | | | | | |
| Null  Null  Sharing | Sharing  Causal  Causal | -2.19  -2.86  -0.66 | 2.13  2.97  1.26 | -1.03  -0.96  -0.53 | 0.303  0.337  0.596 |
| *Bw: ANX on OI* | | | | | |
| Null  Null  Sharing | Sharing  Causal  Causal | 0.09  0.82  0.73 | 0.10  0.42  0.34 | 0.86  1.96  2.16 | 0.390  0.050  0.031 |
| *Fw: OI on ASD* | | | | | |
| Null  Null  Sharing | Sharing  Causal  Causal | 0.38  0.63  0.24 | 0.21  1.07  0.87 | 1.83  0.59  0.28 | 0.067  0.555  0.779 |
| *Bw: ASD on OI* | | | | | |
| Null  Null  Sharing | Sharing  Causal  Causal | 0.34  0.58  0.24 | 0.14  0.90  0.77 | 2.39  0.65  0.32 | 0.017  0.516  0.749 |
| *Fw: OI on BD* | | | | | |
| Null  Null  Sharing | Sharing  Causal  Causal | 0.46  1.38  0.91 | 0.07  0.09  0.02 | 6.39  16.01  36.68 | <0.001  <0.001  <0.001 |
| *Bw: BD on OI* | | | | | |
| Null  Null  Sharing | Sharing  Causal  Causal | 0.08  -0.13  -0.21 | 0.59  1.69  1.13 | 0.13  -0.08  -0.19 | 0.447  0.469  0.427 |
| *Fw: OI on MDD* | | | | | |
| Null  Null  Sharing | Sharing  Causal  Causal | -0.69  -2.62  -1.93 | 0.95  2.63  1.71 | -0.73  -1.00  -1.13 | 0.465  0.317  0.258 |
| *Bw: MDD on OI* | | | | | |
| Null  Null  Sharing | Sharing  Causal  Causal | 0.24  0.06  -0.18 | 0.21  1.25  1.04 | 1.14  0.05  -0.17 | 0.254  0.960  0.865 |
| *Fw: OI on OCD* | | | | | |
| Null  Null  Sharing | Sharing  Causal  Causal | 0.39  0.72  0.33 | 0.24  1.07  0.84 | 1.61  0.67  0.39 | 0.107  0.503  0.697 |
| *Bw: OCD on OI* | | | | | |
| Null  Null  Sharing | Sharing  Causal  Causal | 0.24  0.80  0.56 | 0.05  0.41  0.37 | 4.39  1.92  1.52 | <0.001  0.055  0.129 |
| *Fw: OI on PTSD* | | | | | |
| Null  Null  Sharing | Sharing  Causal  Causal | -1.71  -3.60  -1.89 | 1.63  3.04  1.56 | -1.05  -1.18  -1.21 | 0.294  0.238  0.226 |
| *Bw: PTSD on OI* | | | | | |
| Null  Null  Sharing | Sharing  Causal  Causal | 0.27  1.05  0.78 | 0.03  0.13  0.12 | 8.54  7.92  6.75 | <0.001  <0.001  <0.001 |
| *Fw: OI on SZ* | | | | | |
| Null  Null  Sharing | Sharing  Causal  Causal | 0.467  1.32  0.849 | 0.03  0.05  0.04 | 16.1  25.5  19.4 | <0.001  <0.001  <0.001 |
| *Bw: SZ on OI* | | | | | |
| Null  Null  Sharing | Sharing  Causal  Causal | -0.88  0.03  0.92 | 2.61  2.38  0.55 | -0.34  0.01  1.68 | 0.734  0.992  0.093 |

Abbreviations: MR: mendelian randomization; ELPD: expected log pointwise posterior density; 95%CI: 95% confidence interval; SE: standard error; Fwd: forward MR; Bwd: backward MR; OI: occupational income; ADHD: attention deficit hyperactivity disorder; AN: anorexia nervosa; ANX: anxiety disorder; ASD: autism spectrum disorder; BD: bipolar disorder; MDD: major depressive disorder; OCD: obsessive-compulsive disorder; PTSD: post-traumatic stress disorder; SZ: schizophrenia.

#### Supplementary Table 16: Results of univariable bidirectional Mendelian Randomization of occupational income against mental illnesses, after Steiger filtering

| **MR** | **N SNP** | **IVW, B (95% CI)** | **p-value** | **WM, B (95% CI)** | **p-value** | **MR-Egger, B (95% CI)** | **p-value** | **Egger intercept p-value** | **Mean F** |
| --- | --- | --- | --- | --- | --- | --- | --- | --- | --- |
| OI on ADHD | 32 | -0.780 (-0.972; -0.587) | <0.001 | -0.556 (-0.890; 0.222) | 0.001 | 0.094 (-1.36; 1.55) | 0.900 | 0.033 | 40.6 |
| OI on AN | 34 | 0.608 (0.337; 0.879) | <0.001 | 0.437 (-0.007; 0.882) | 0.054 | 2.51 (0.635; 4.39) | 0.013 | 0.001 | 40.4 |
| OI on ANX | 31 | 0.004 (-0.566; 0.575) | 0.988 | -0.102 (-0.961; 0.757) | 0.816 | 0.164 (-2.83; 3.16) | 0.915 | 0.713 | 40.9 |
| OI on ASD | 29 | 0.125 (-0.179; 0.428) | 0.421 | 0.104 (-0.363; 0.571) | 0.664 | 2.04 (-0.054; 4.03) | 0.054 | 0.010 | 38.9 |
| OI on BD | 29 | -0.134 (-0.338; 0.071) | 0.201 | -0.091 (-0.397; 0.215) | 0.561 | 0.445 (-0.905; 1.79) | 0.524 | 0.024 | 38.7 |
| OI on MDD | 33 | -0.322 (-0.502; -0.143) | <0.001 | -0.125 (-0.420; 0.171) | 0.408 | 0.309 (-1.10; 1.72) | 0.671 | 0.376 | 40.3 |
| OI on OCD | 25 | 0.107 (-0.672; 0.885) | 0.788 | -0.075 (-1.18; 1.03) | 0.894 | -1.37 (-4.75; 2.02) | 0.437 | 0.590 | 40.6 |
| OI on PTSD | 33 | -0.051 (-0.347; 0.245) | 0.736 | -0.271 (-0.717; 0.176) | 0.235 | -0.407 (-2.26; 1.44) | 0.669 | 0.702 | 40.3 |
| OI on SZ | 33 | 0.082 (-0.240; 0.077) | 0.313 | -0.172 (-0.497; 0.153) | 0.299 | 1.24 (-0.700; 3.19) | 0.219 | 0.001 | 40.6 |

Abbreviations: OI: occupationall income; ADHD: attention deficit hyperactivity disorder; AN: anorexia nervosa; ANX: anxiety disorder; ASD: autism spectrum disorders; BD: bipolar disorder; MDD: major depressive disorder; OCD: obsessive-compulsive disorder; PTSD: post-traumatic stress disorder; SZ: schizophrenia; MR: mendelian randomization; SNP: single nucleotide polymorphism; IVW: inverse variance weighted (fixed effect); B: effect estimates are log-odds; 95% CI: 95% confidence interval; WM: weighted median; NR: not reported because not enough SNP to perform MR.

### **Univariable Mendelian randomization of social deprivation and mental illnesses**

#### Supplementary Table 17: Odds Ratio of univariable forward Mendelian randomization analysis of social deprivation against mental illnesses

| **MR: method** | **OR (95% CI)** | **p-value** |
| --- | --- | --- |
| SD → ADHD:  IVW  WM  MR-Egger | 2.04 (1.66; 2.51)  1.88 (1.35; 2.61)  0.855 (0.039; 18.9) | <0.001  <0.001  0.925 |
| SD→ AN:  IVW  WM  MR-Egger | 0.703 (0.539; 0.917)  0.746 (0.504; 1.11)  1.35 (0.055; 33.4) | 0.009  0.144  0.858 |
| SD → ANX:  IVW  WM  MR-Egger | 1.18 (0.671; 2.07)  1.30 (0.615; 2.74)  2.47 (0.027; 229.9) | 0.566  0.494  0.708 |
| SD → ASD:  IVW  WM  MR-Egger | 0.990 (0.756; 1.29)  1.24 (0.829; 1.86)  0.627 (0.013; 30.5) | 0.939  0.292  0.821 |
| SD → BD:  IVW  WM  MR-Egger | 0.949 (0.777; 1.16)  0.877 (0.656; 1.17)  2.78 (0.048; 162.5) | 0.607  0.375  0.639 |
| SD → MDD:  IVW  WM  MR-Egger | 1.16 (0.955; 1.39)  1.22 (0.939; 1.59)  1.45 (0.166; 12.6) | 0.138  0.137  0.749 |
| SD → OCD:  IVW  WM  MR-Egger | 1.12 (0.553; 2.25)  1.22 (0.451; 3.31)  0.306 (0.001; 549.4) | 0.761  0.692  0.767 |
| SD → PTSD:  IVW  WM  MR-Egger | 1.18 (0.881; 1.58)  1.21 (0.811; 1.80)  3.96 (0.255; 61.4) | 0.268  0.351  0.358 |
| SD → SZ:  IVW  WM  MR-Egger | 1.24 (1.05; 1.46)  1.37 (1.02; 1.88)  7.61 (0.371; 156.3) | 0.012  0.040  0.236 |

Abbreviations: MR: Mendelian randomization; OR: Odds Ratio; 95% CI: 95% confidence intervals; SD: social deprivation; ADHD: attention deficit hyperactivity disorder; AN: anorexia nervosa; ANX: anxiety disorder; ASD: autism spectrum disorders; BD: bipolar disorder; MDD: major depressive disorder; OCD: obsessive-compulsive disorder; PTSD: post-traumatic stress disorder; SZ: schizophrenia; IVW: inverse variance weighted (fixed effect); WM: weighted median; NR: not reported because not enough SNP to perform MR.

#### Supplementary Table 18: results of bidirectional MR of Social Deprivation (SD) against mental health traits

| **MR** | **N SNP** | **IVW, B (95% CI)** | **p-value** | **IVW Q(df)** | **Q p-value** | **WM, B (95% CI)** | **p-value** | **MR-Egger, B (95% CI)** | **p-value** | **Egger intercept p-value** | **Steiger Test p-value** | **MR-PRESSO** | **Mean F** |
| --- | --- | --- | --- | --- | --- | --- | --- | --- | --- | --- | --- | --- | --- |
| Fw: SD on ADHD | 7 | 0.713 (0.504; 0.922) | <0.001 | 17 (6) | 0.001 | 0.629 (0.298; 0.960) | <0.001 | -0.157 (-3.25; 2.94) | 0.925 | 0.603 | 0.079 | DT; p=0.083 | 32.1 |
| Bw: ADHD on SD | 23 | 0.222 (0.181; 0.262) | <0.001 | 41 (22) | 0.009 | 0.193 (0.129; 0.257) | <0.001 | 0.259 (-0.061; 0.579) | 0.128 | 0.819 | <0.001 | DT; p=0.464 | 39.2 |
| Fw: SD on AN | 10 | -0.352 (-0.618; -0.087) | 0.009 | 21 (9) | 0.014 | -0.293 (-0.658; 0.072) | 0.115 | 0.302 (-2.90; 3.51) | 0.858 | 0.697 | 0.170 | DT; p=0.100 | 32.8 |
| Bw: AN on SD | 4 | -0.038 (-0.113; 0.036) | 0.314 | 4 (3) | 0.303 | -0.052 (-0.146; 0.042) | 0.277 | -0.403 (-1.15; 0.342) | 0.400 | 0.436 | NR ^b^ | GT; p=0.352 | 31.9 |
| Fw: SD on ANX | 9 | 0.165 (-0.399; 0.730) | 0.566 | 5 (8) | 0.731 | 0.261 (-0.471; 0.992) | 0.485 | 0.904 (-3.63; 5.44) | 0.708 | 0.757 | NR ^b^ | GT; p=0.709 | 33.0 |
| Bw: ANX on SD | 0 | NR ^c^ | NR ^c^ | NR ^c^ | NR ^c^ | NR ^c^ | NR ^c^ | NR ^c^ | NR ^c^ | NR ^c^ | NR ^c^ | NR ^c^ | NR ^c^ |
| Fw: SD on ASD | 9 | -0.010 (-0.279; 0.258) | 0.939 | 21 (8) | 0.009 | 0.218 (-0.200; 0.635) | 0.308 | -0.466 (-4.35; 3.42) | 0.821 | 0.823 | NR ^b^ | DT; p=0.290 | 33.1 |
| Bw: ASD on SD | 0 | NR ^c^ | NR ^c^ | NR ^c^ | NR ^c^ | NR ^c^ | NR ^c^ | NR ^c^ | NR ^c^ | NR ^c^ | NR ^c^ | NR ^c^ | NR ^c^ |
| Fw: SD on BD | 8 | -0.052 (-0.252; 0.148) | 0.607 | 67 (7) | <0.001 | -0.131 (-0.427; 0.164) | 0.384 | 1.02 (-3.04; 5.09) | 0.639 | 0.618 | NR ^b^ | DT; p=0.756 | 31.8 |
| Bw: BD on SD | 36 | -0.019 (-0.051; 0.013) | 0.244 | 105 (35) | <0.001 | -0.004 (-0.060; 0.052) | 0.884 | 0.023 (-0.284; 0.330) | 0.885 | 0.787 | NR ^b^ | DT; p=0.096 | 39.2 |
| Fw: SD on MDD | 8 | 0.141 (-0.046; 0.328) | 0.138 | 12 (7) | 0.103 | 1.00 (-0.075; 0.475) | 0.155 | 0.370 (-1.80; 2.54) | 0.749 | 0.842 | NR ^b^ | GT; p=0.125 | 33.4 |
| Bw: MDD on SD | 0 | NR ^c^ | NR ^c^ | NR ^c^ | NR ^c^ | NR ^c^ | NR ^c^ | NR ^c^ | NR ^c^ | NR ^c^ | NR ^c^ | NR ^c^ | NR ^c^ |
| Fw: SD on OCD | 8 | 0.109 (-0.593; 0.811) | 0.761 | 10 (7) | 0.177 | 0.201 (-0.773; 1.18) | 0.685 | -1.19 (-8.68; 6.31) | 0.767 | 0.744 | NR ^b^ | GT; p=0.206 | 33.4 |
| Bw: OCD on SD | 0 | NR ^c^ | NR ^c^ | NR ^c^ | NR ^c^ | NR ^c^ | NR ^c^ | NR ^c^ | NR ^c^ | NR ^c^ | NR ^c^ | NR ^c^ | NR ^c^ |
| Fw: SD on PTSD | 9 | 0.165 (-0.127; 0.458) | 0.268 | 10 (8) | 0.273 | 0.190 (-0.228; 0.609) | 0.373 | 1.38 (-1.37; 4.12) | 0.358 | 0.412 | NR ^b^ | GT; p=0.287 | 33.1 |
| Bw: PTSD on SD | 0 | NR ^c^ | NR ^c^ | NR ^c^ | NR ^c^ | NR ^c^ | NR ^c^ | NR ^c^ | NR ^c^ | NR ^c^ | NR ^c^ | NR ^c^ | NR ^c^ |
| Fw: SD on SZ | 8 | 0.213 (0.046; 0.380) | 0.012 | 62 (7) | <0.001 | 0.314 (0.015; 0.614) | 0.040 | 2.03 (-0.993; 5.05) | 0.236 | 0.278 | 0.277 | DT; p=0.999 | 31.8 |
| Bw: SZ on SD | 176 | 0.042 (0.026; 0.058) | <0.001 | 402 (175) | <0.001 | 0.020 (-0.007; 0.048) | 0.147 | -0.043 (-0.137; 0.052) | 0.376 | 0.071 | <0.001 | DT; p=0.836 | 45.6 |

Abbreviations: Fw: forward analysis; Bw: backward analysis; SD: social deprivation; ADHD: attention deficit hyperactivity disorder; AN: anorexia nervosa; ANX: anxiety disorder; ASD: autism spectrum disorder; BD: bipolar disorder; MDD: major depressive disorder; OCD: obsessive-compulsive disorder; PTSD: post-traumatic stress disorder; SZ: schizophrenia; MR: mendelian randomization; SNP: single nucleotide polymorphism; IVW: inverse variance weighted (fixed effect); B: effect estimates are log-odds for binary traits (i.e., for mental illnesses) and unstandardized regression coefficient for continuous traits (i.e., for social deprivation); 95% CI: 95% confidence interval; Q: Cochran’s Q measure of heterogeneity; df: degree of freedom; WM: weighted median; DT: distortion test; GT: global test.

Legend:

P-value threshold <5e-8

^a^ The Mendelian randomization pleiotropy residual sum and outlier (MR-PRESSO) test identifies possible bias from horizontal pleiotropy. The test consists of three parts, (1) the MR-PRESSO global test which detects horizontal pleiotropy, (2) the outlier corrected causal estimate which corrects for the detected horizontal pleiotropy and (3) the MR-PRESSO distortion test which estimates if the causal estimate is significantly different (at p<0.05) after adjustment for outliers. We conduct all three stages (with the argument NbDistribution=1000, namely using1000 simulation form the null distribution to compute empirical p-values) and present the outlier adjusted causal estimates (OACE) when both global and distortion tests are significant.

^b^ We did not run Steiger Test if none of the MR analysis resulted significant (NR: not reported in the cell).

^c^ Not enough SNP to perform MR (NR: not reported in the cell).

#### Plots - Forward analyses

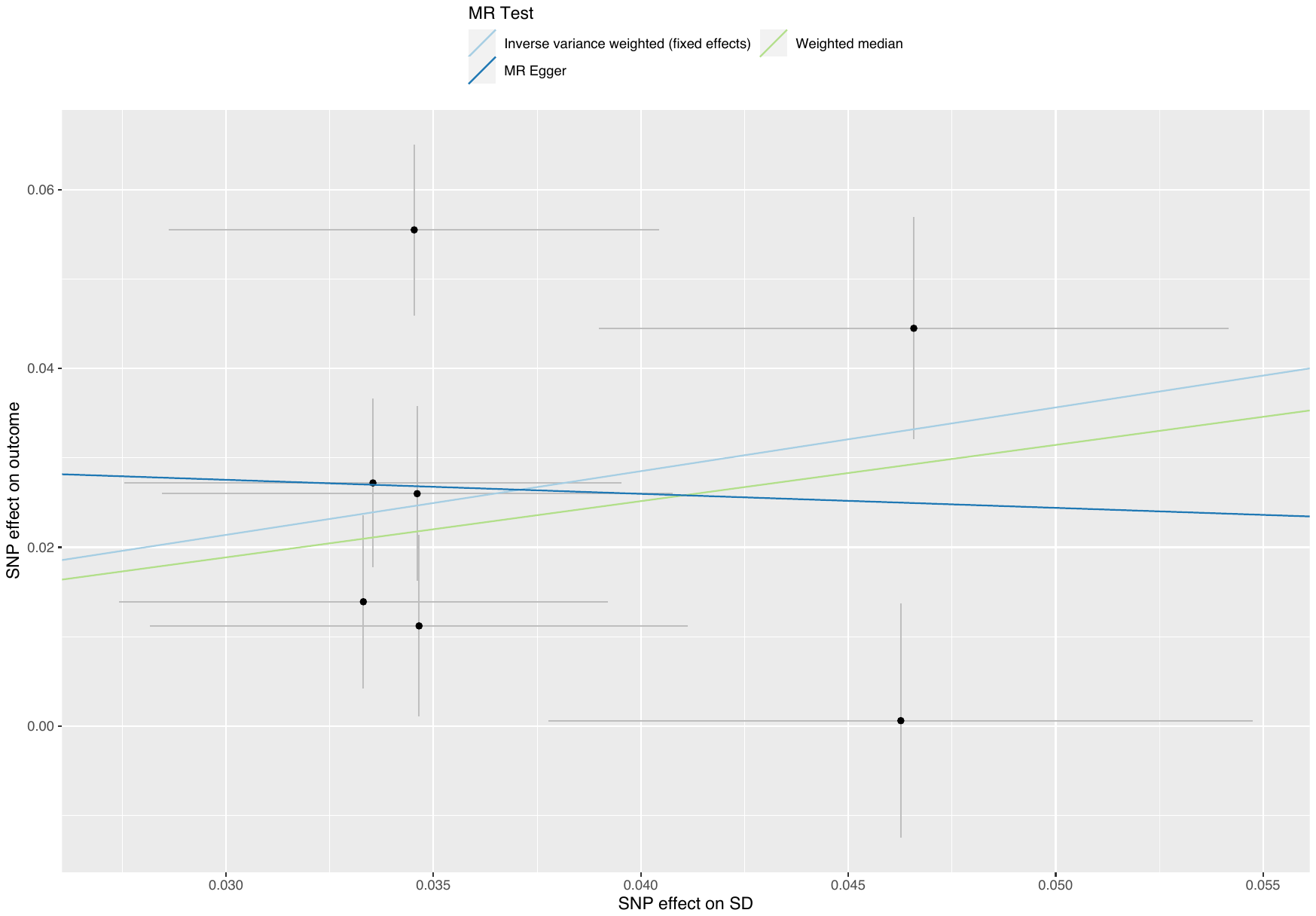

##### Supplementary Figure 82: scatterplot of social deprivation against ADHD

Abbreviations: MR: Mendelian randomization; SNP: single nucleotide polymorphism; SD: social deprivation; ADHD: attention deficit hyperactivity disorder.

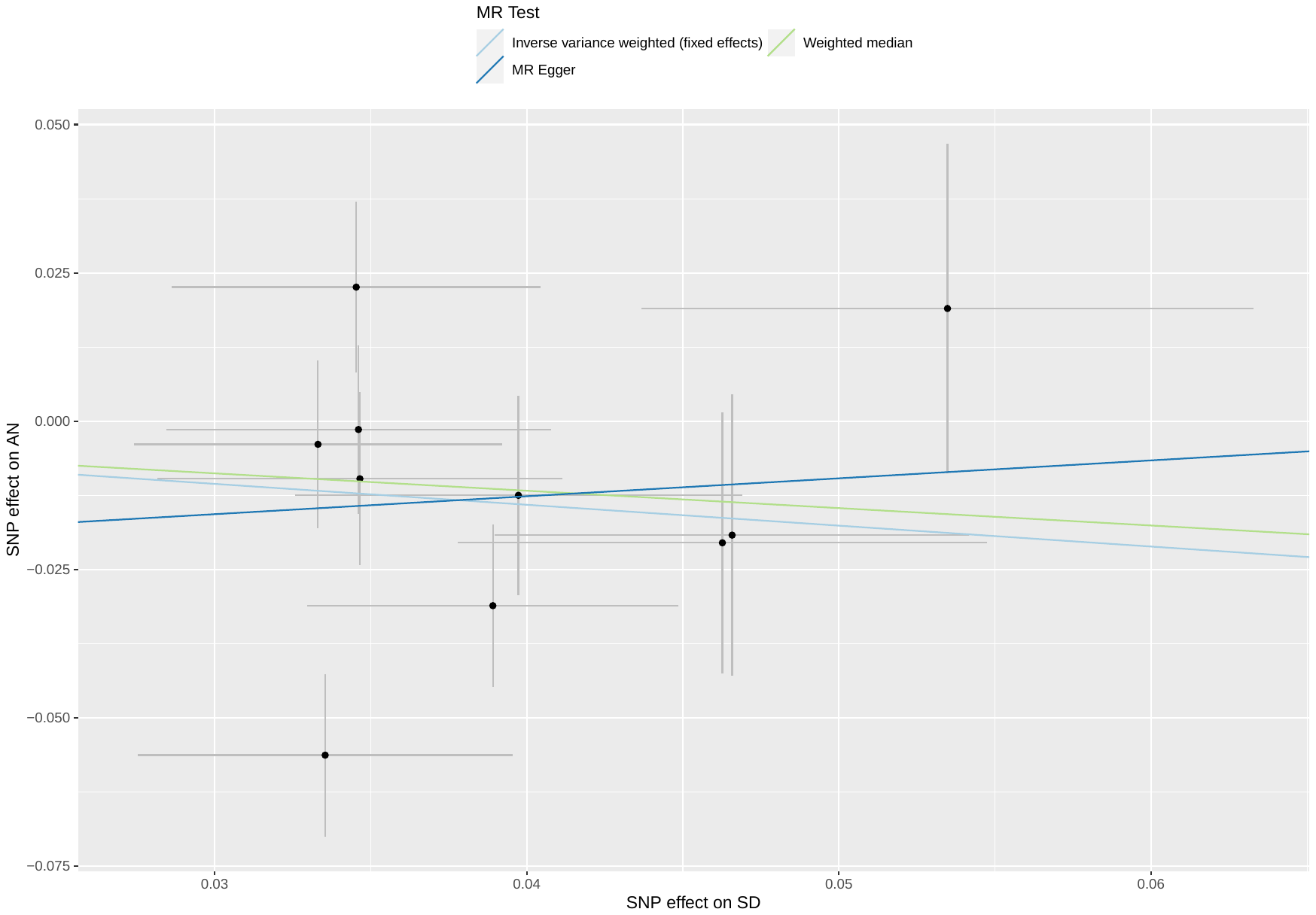

##### Supplementary Figure 83: scatterplot of social deprivation against AN

Abbreviations: MR: Mendelian randomization; SNP: single nucleotide polymorphism; SD: social deprivation; AN: anorexia nervosa.

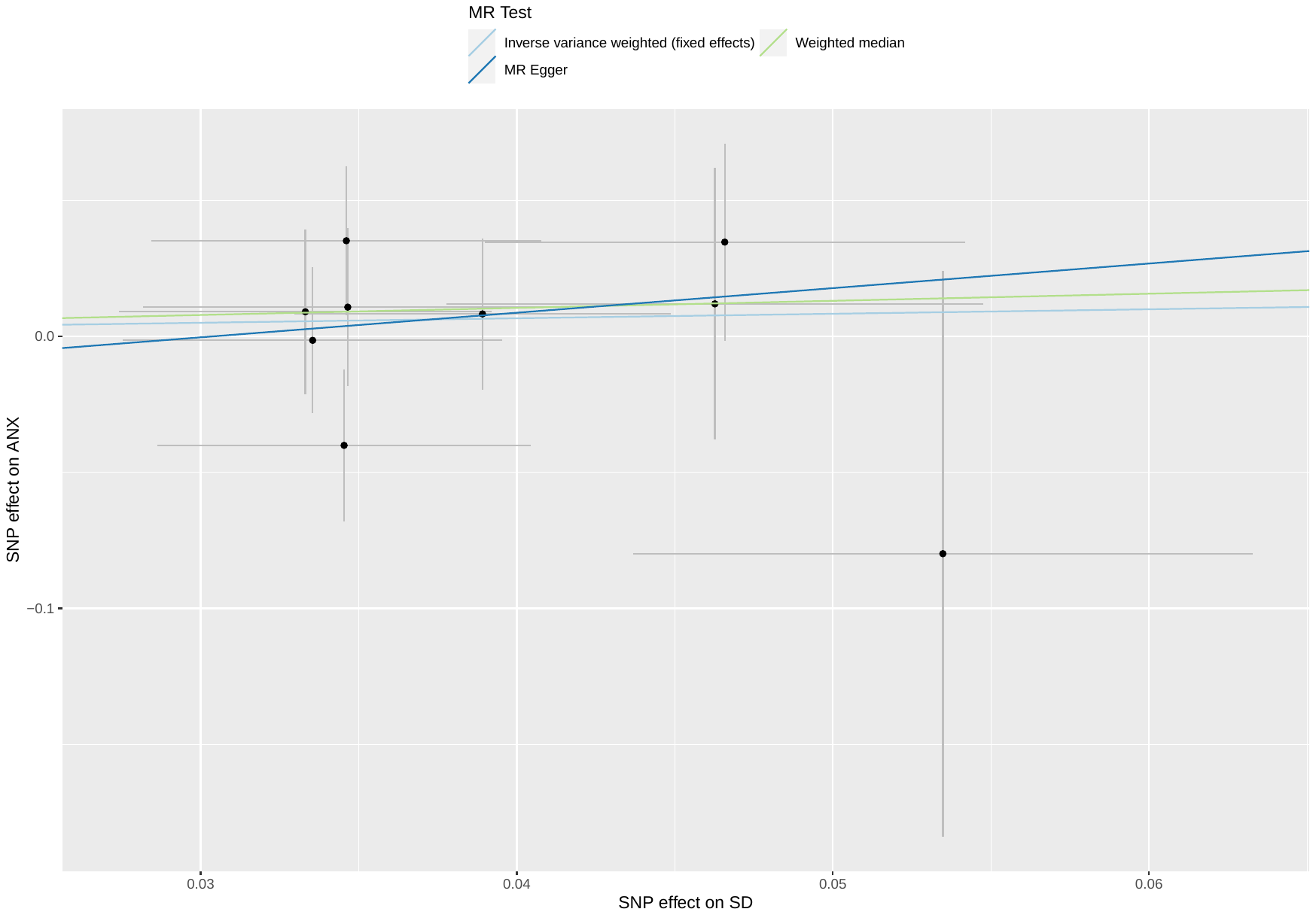

##### Supplementary Figure 84: scatterplot of social deprivation against ANX

Abbreviations: MR: Mendelian randomization; SNP: single nucleotide polymorphism; SD: social deprivation; ANX: anxiety disorders.

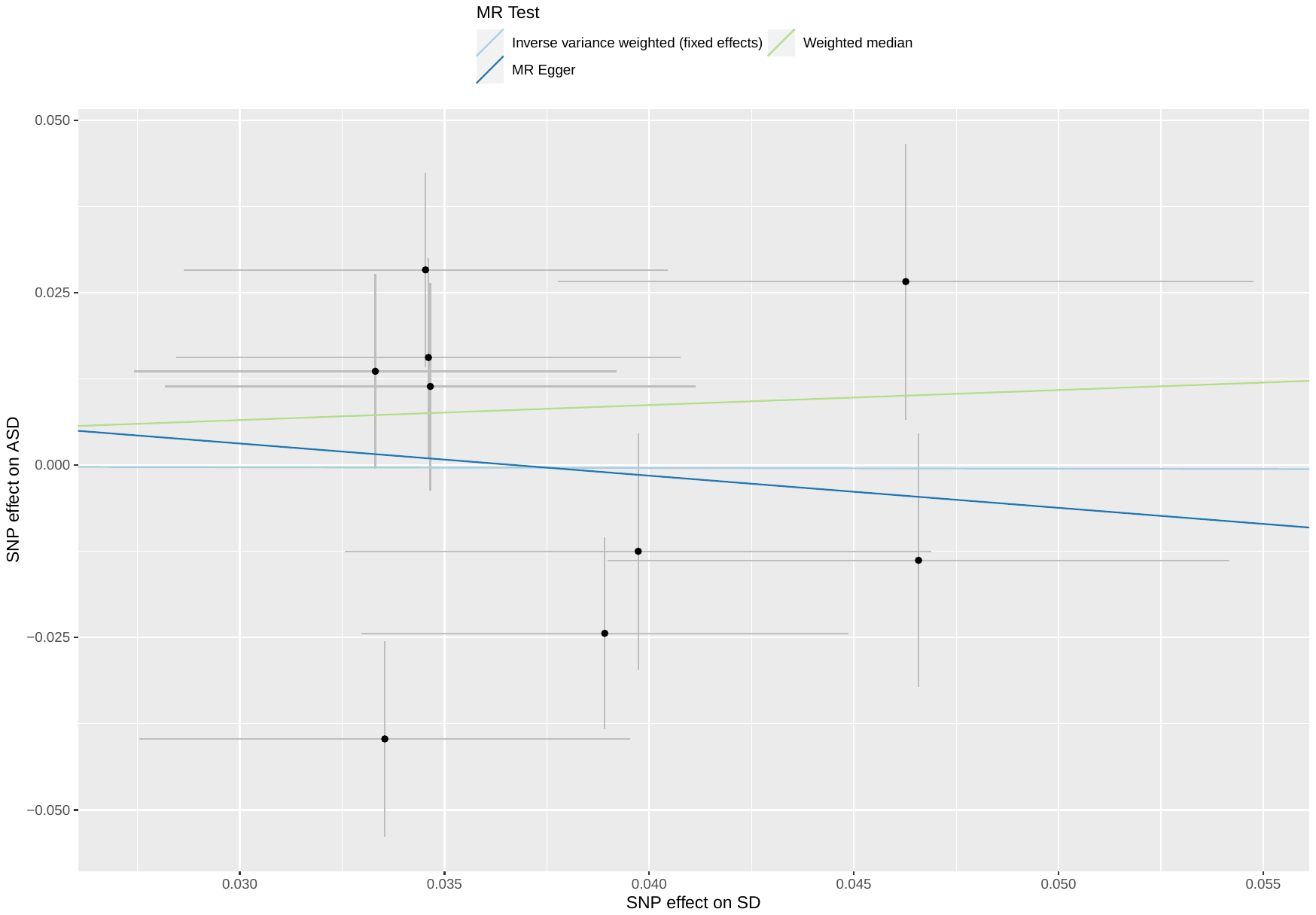

##### Supplementary Figure 85: scatterplot of social deprivation against ASD

Abbreviations: MR: Mendelian randomization; SNP: single nucleotide polymorphism; SD: social deprivation; ASD: autism spectrum disorders.

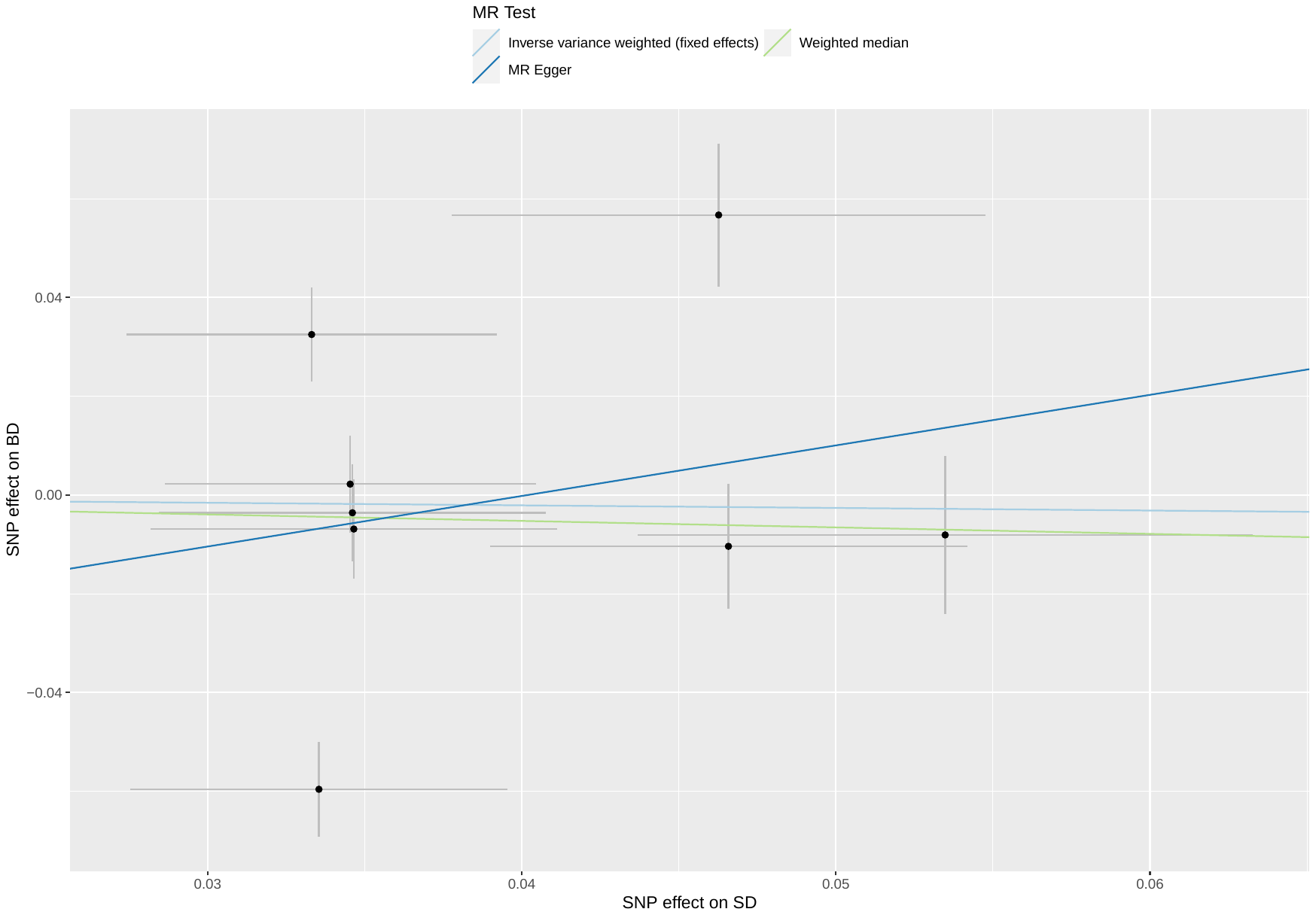

##### Supplementary Figure 86: scatterplot of social deprivation against BD

Abbreviations: MR: Mendelian randomization; SNP: single nucleotide polymorphism; SD: social deprivation; BD: bipolar disorder.

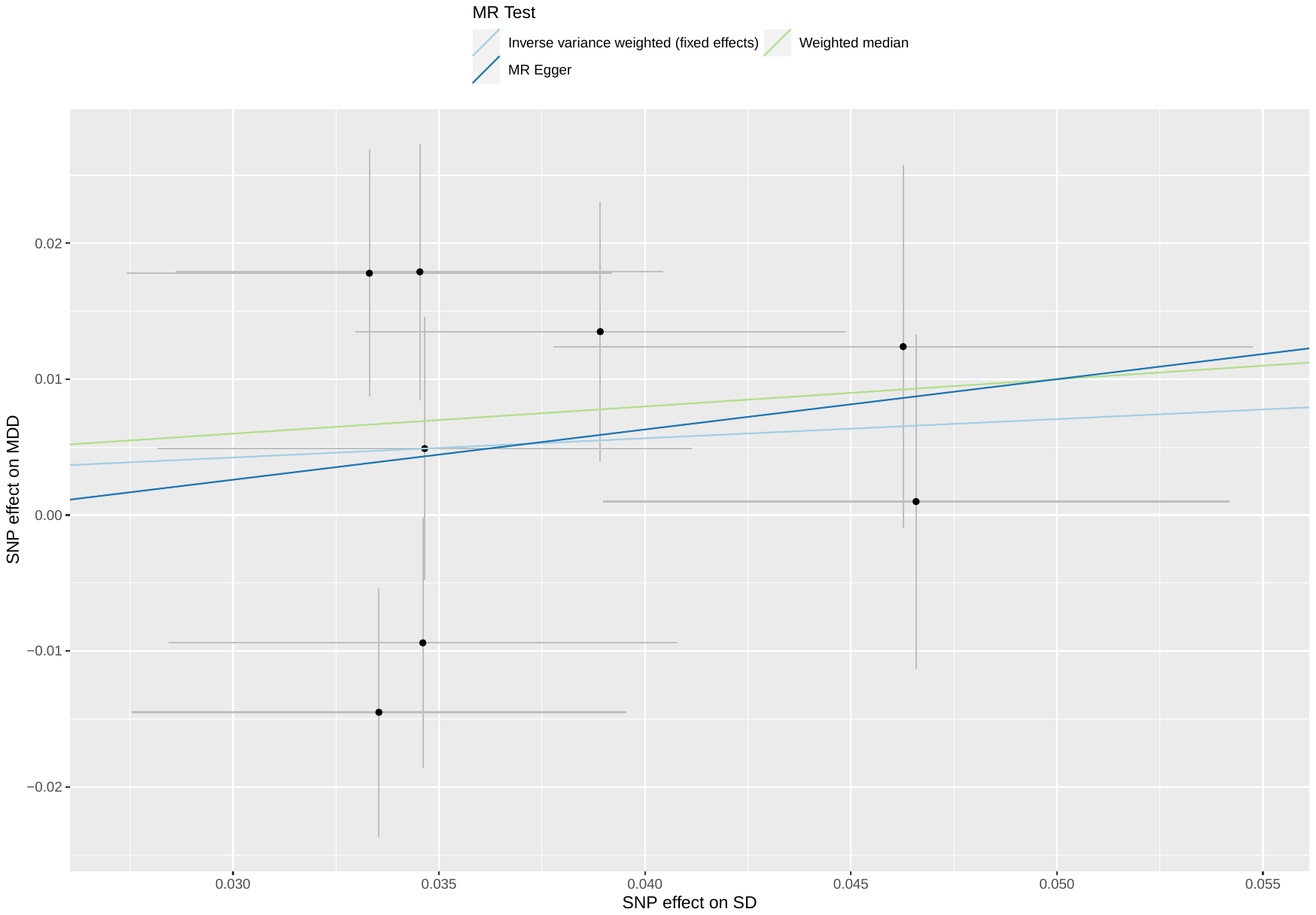

##### Supplementary Figure 87: scatterplot of social deprivation against MDD

Abbreviations: MR: Mendelian randomization; SNP: single nucleotide polymorphism; SD: social deprivation; MDD: major depressive disorder.

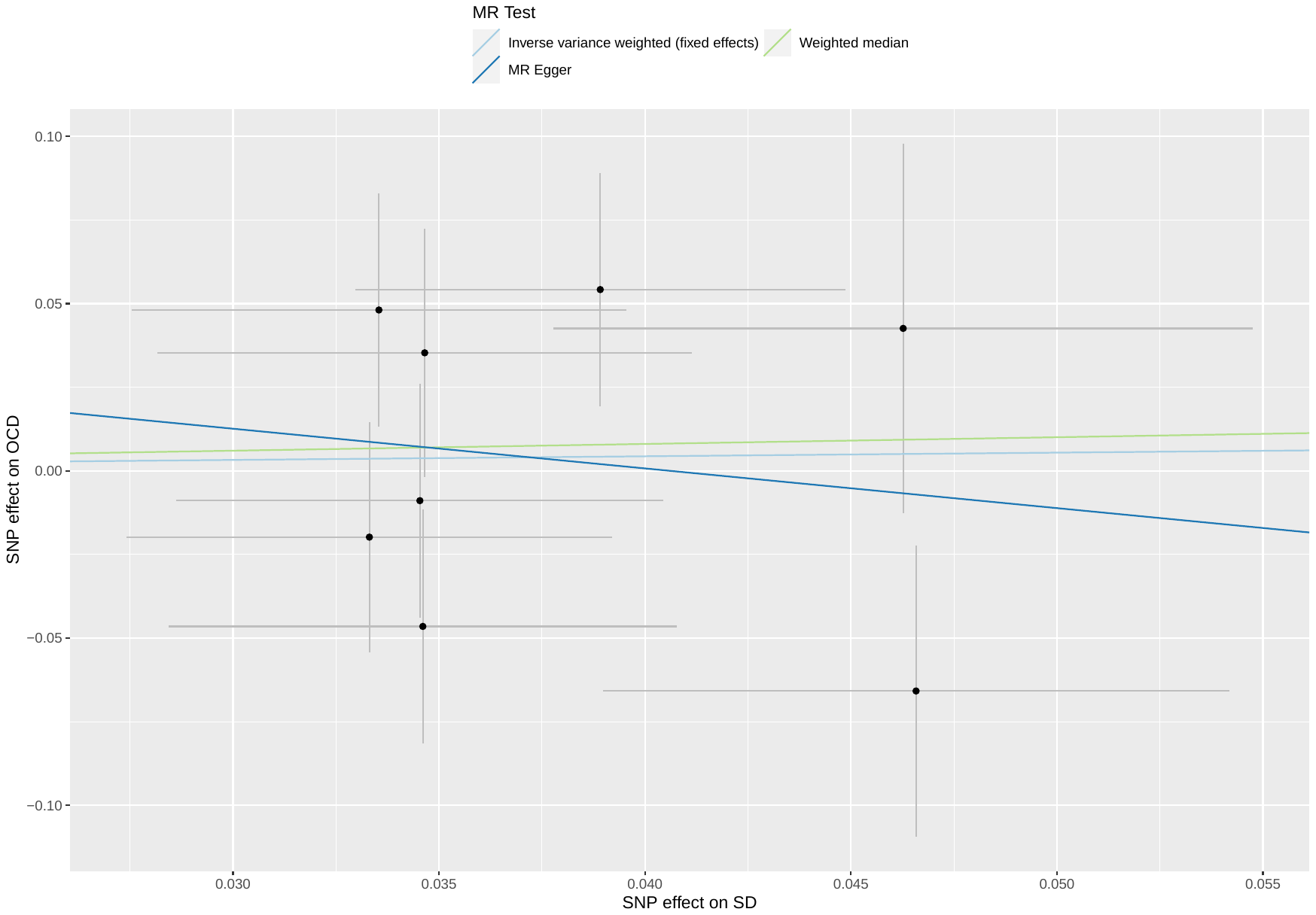

##### Supplementary Figure 88: scatterplot of social deprivation against OCD

Abbreviations: MR: Mendelian randomization; SNP: single nucleotide polymorphism; SD: social deprivation; OCD: obsessive-compulsive disorder.

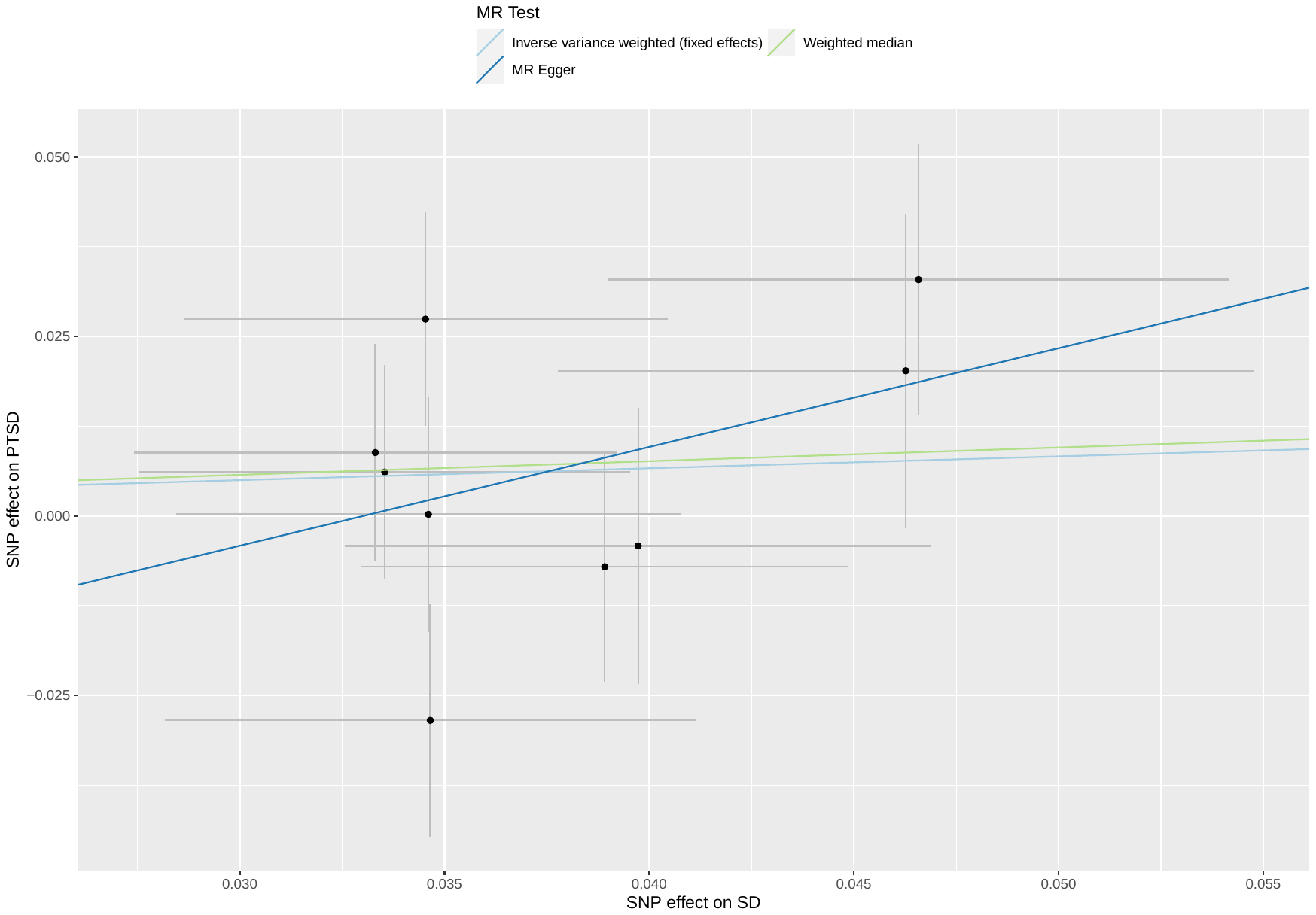

##### Supplementary Figure 89: scatterplot of social deprivation against PTSD

Abbreviations: MR: Mendelian randomization; SNP: single nucleotide polymorphism; SD: social deprivation; PTSD: post-traumatic stress disorder.

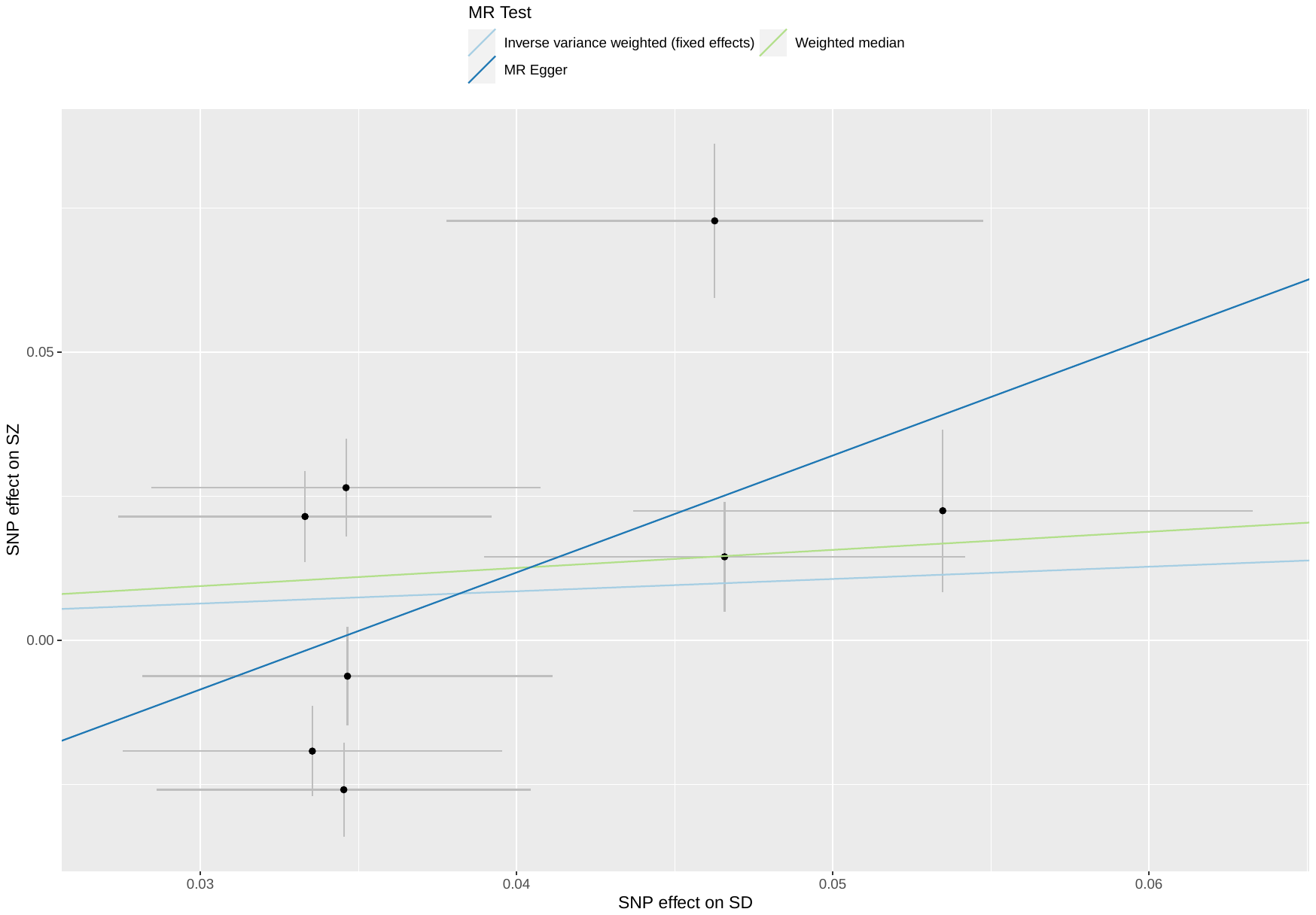

##### Supplementary Figure 90: scatterplot of social deprivation against SZ

Abbreviations: MR: Mendelian randomization; SNP: single nucleotide polymorphism; SD: social deprivation; SZ: schizophrenia.

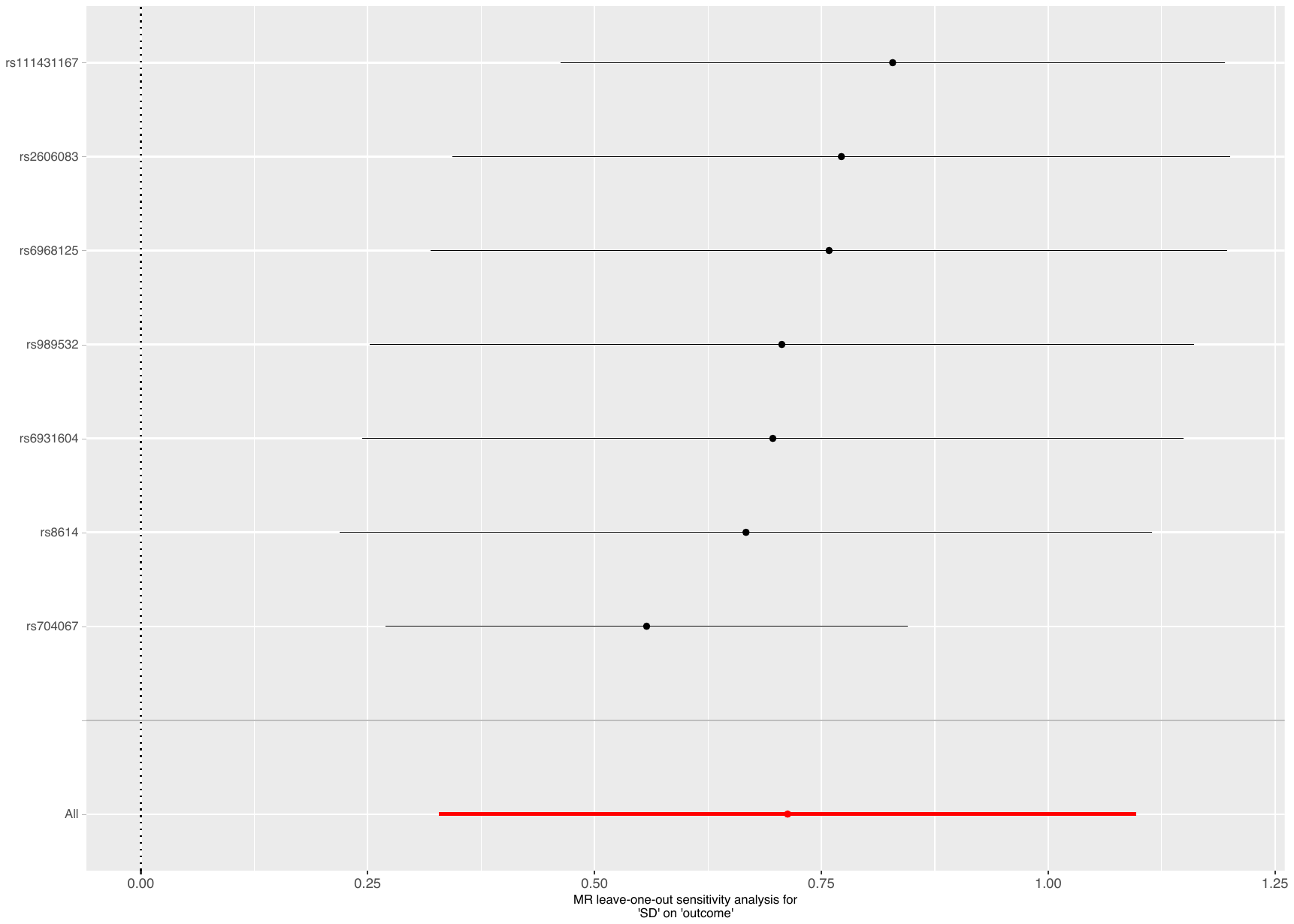

Supplementary Figure 91: leave-one-out analysis of social deprivation against ADHD

Abbreviations: MR: Mendelian randomization; SD: social deprivation; ADHD: attention deficit hyperactivity disorder.

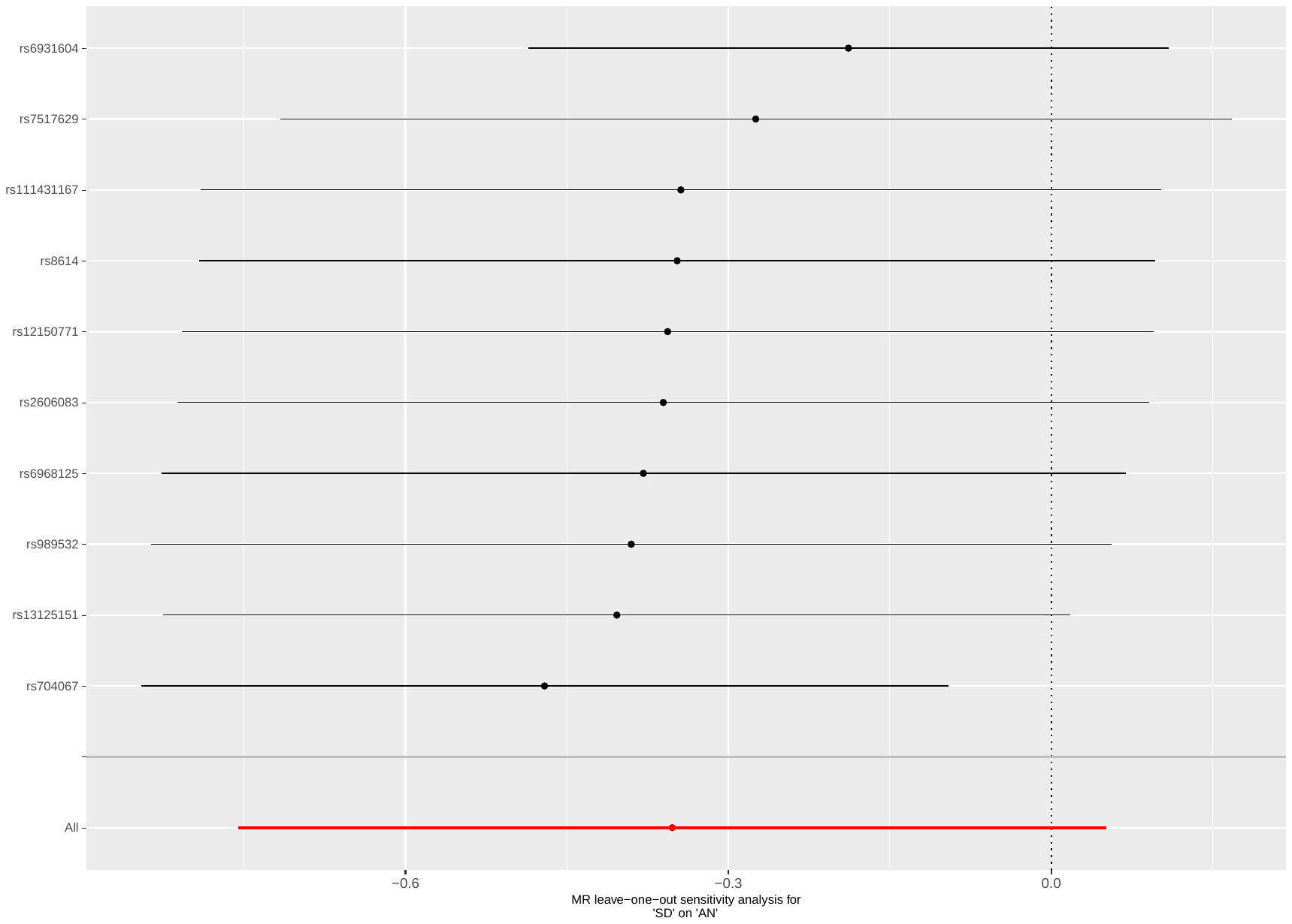

Supplementary Figure 92: leave-one-out analysis of social deprivation against AN

Abbreviations: MR: Mendelian randomization; SD: social deprivation; AN: anorexia nervosa.

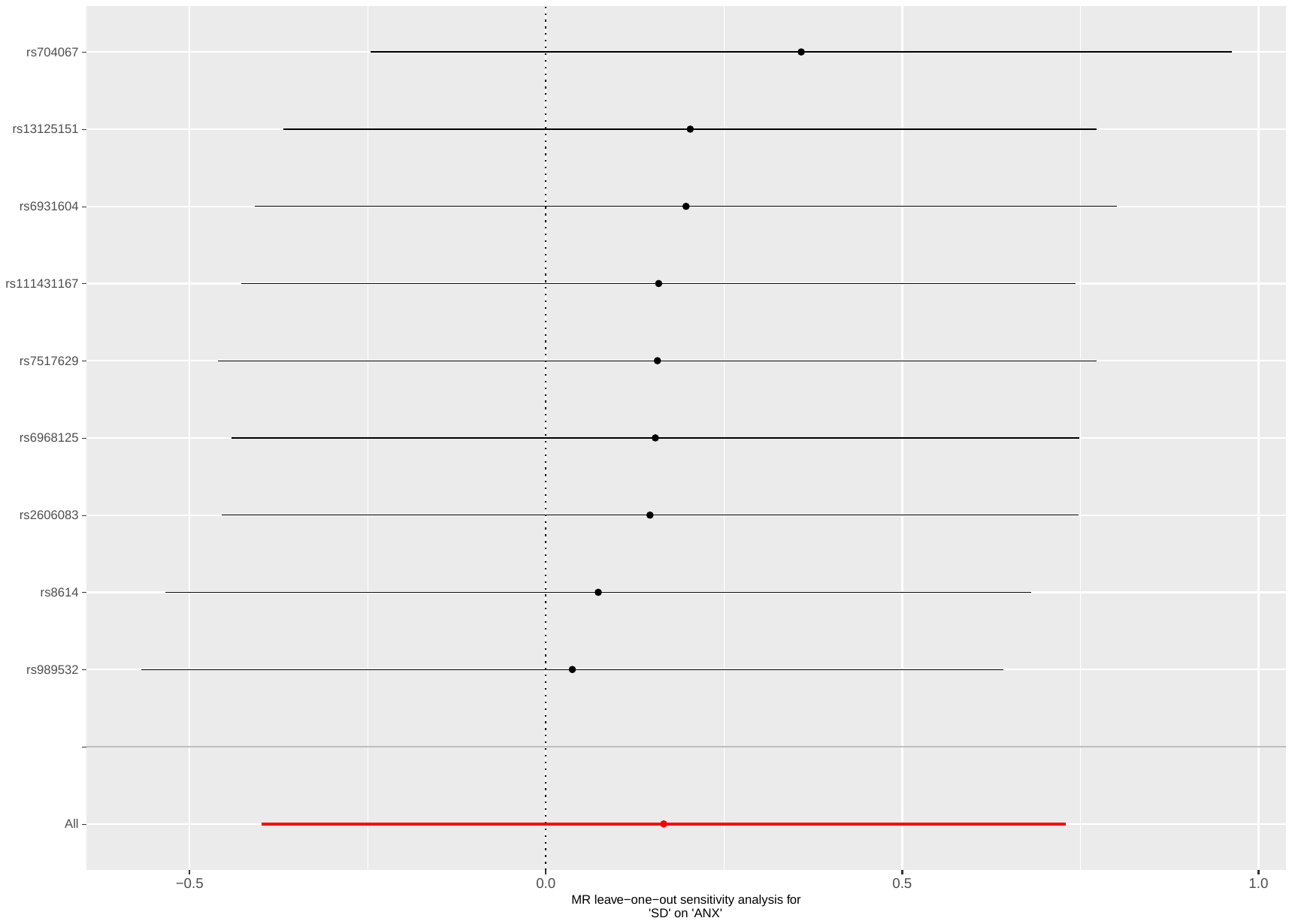

Supplementary Figure 93: leave-one-out analysis of social deprivation against ANX

Abbreviations: MR: Mendelian randomization; SD: social deprivation; ANX: anxiety disorders.

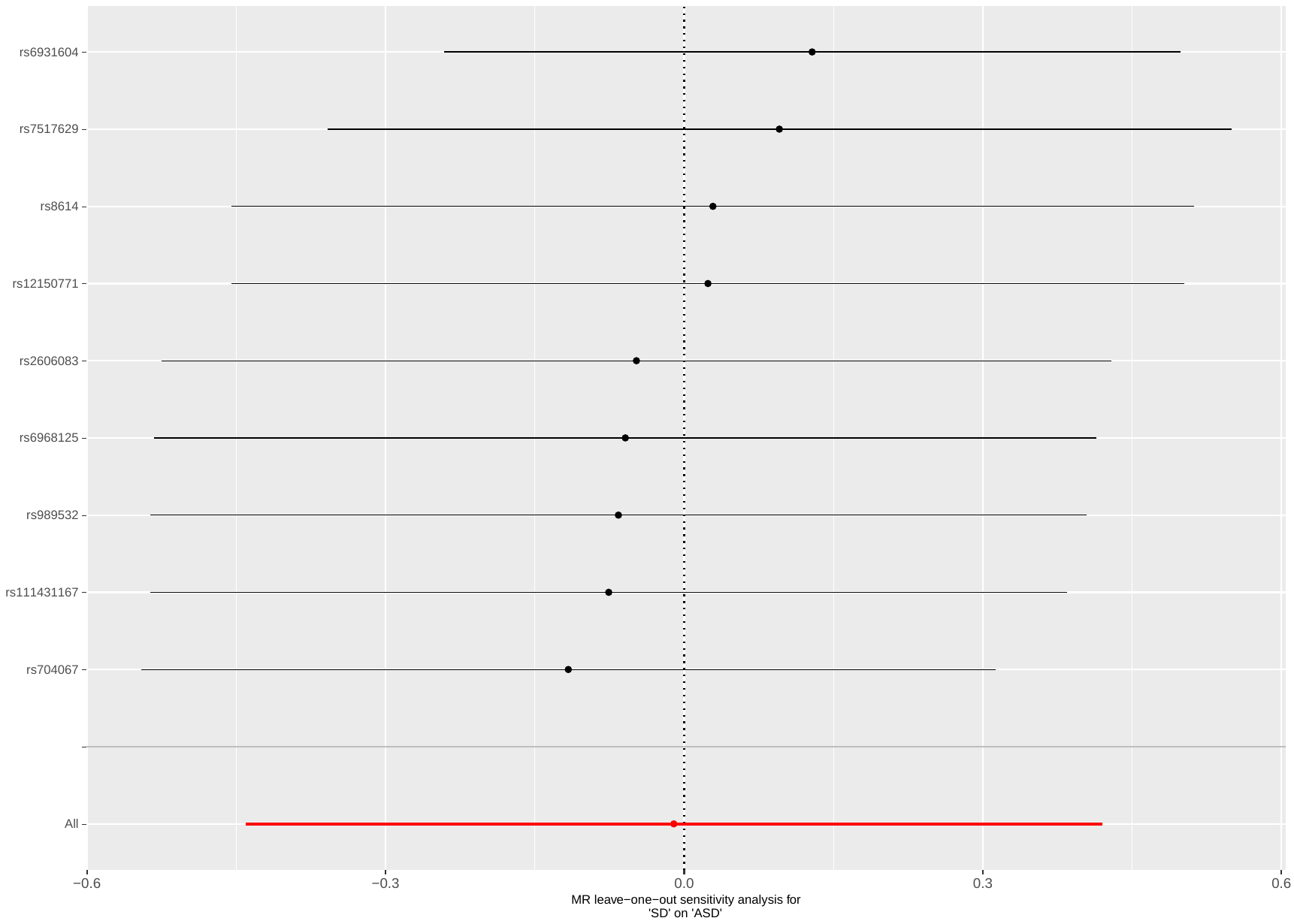

Supplementary Figure 94: leave-one-out analysis of social deprivation against ASD

Abbreviations: MR: Mendelian randomization; SD: social deprivation; ASD: autism spectrum disorders.

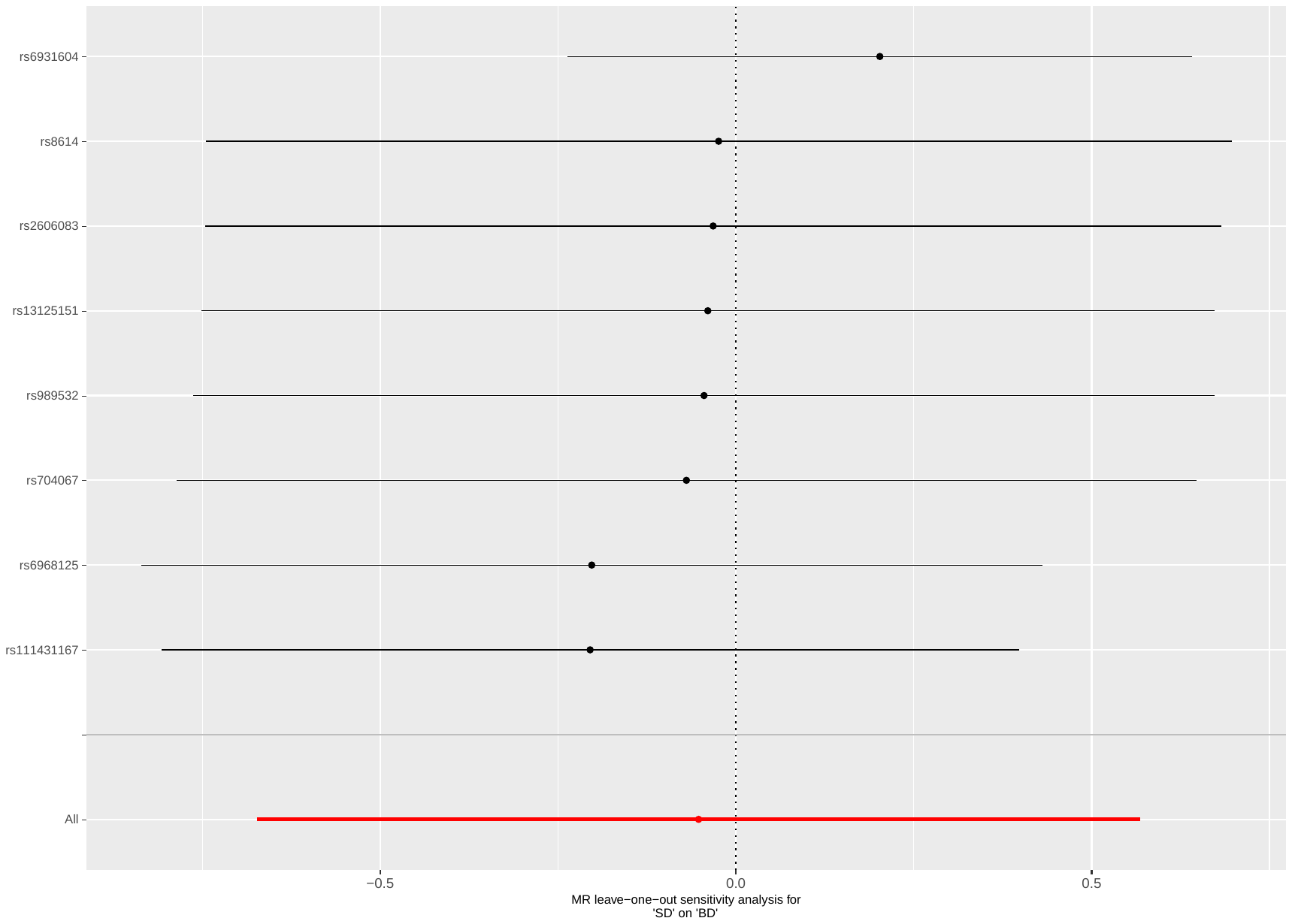

Supplementary Figure 95: leave-one-out analysis of social deprivation against BD

Abbreviations: MR: Mendelian randomization; SD: social deprivation; BD: bipolar disorder.

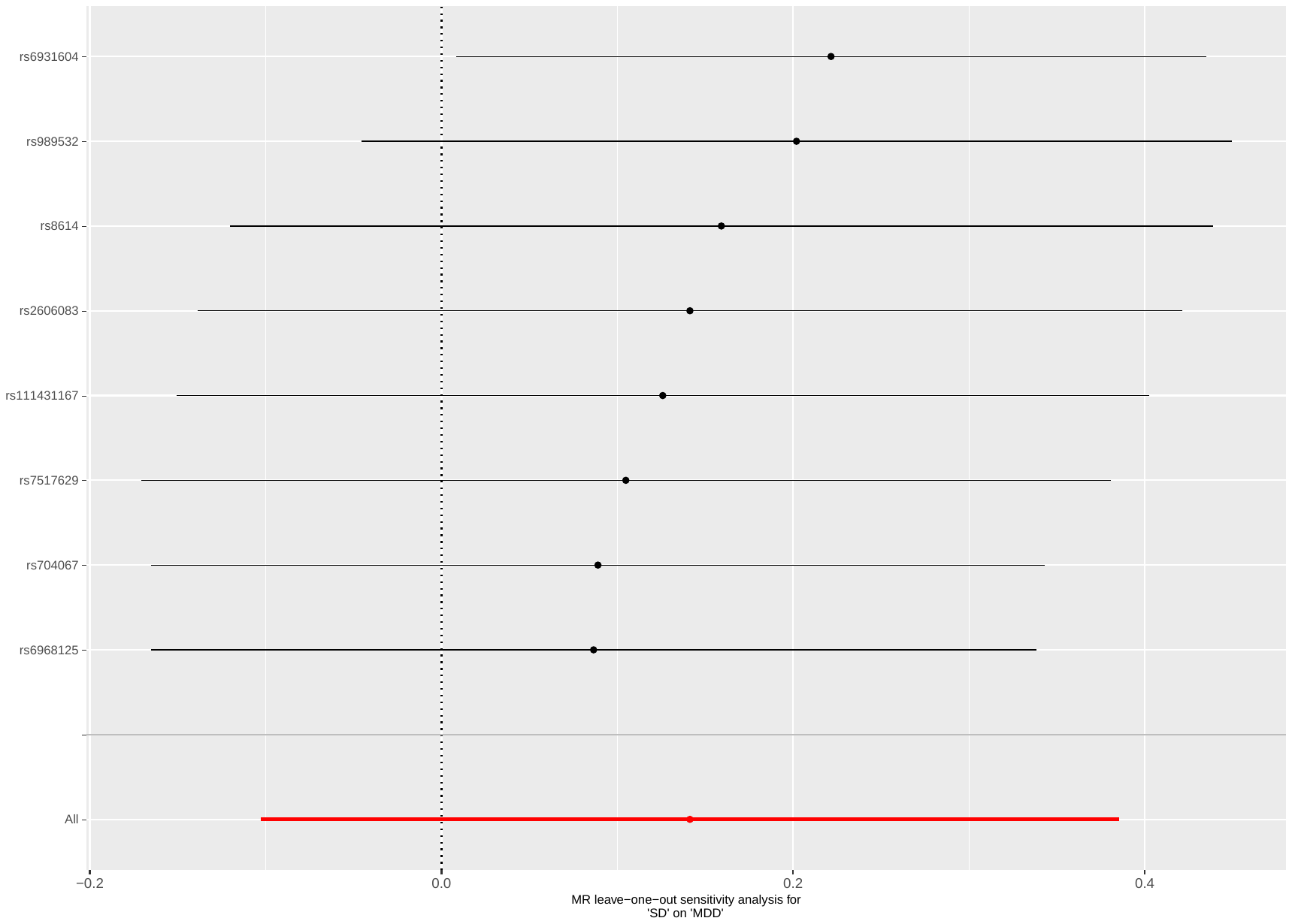

Supplementary Figure 96: leave-one-out analysis of social deprivation against MDD

Abbreviations: MR: Mendelian randomization; SD: social deprivation; MDD: major depressive disorder.

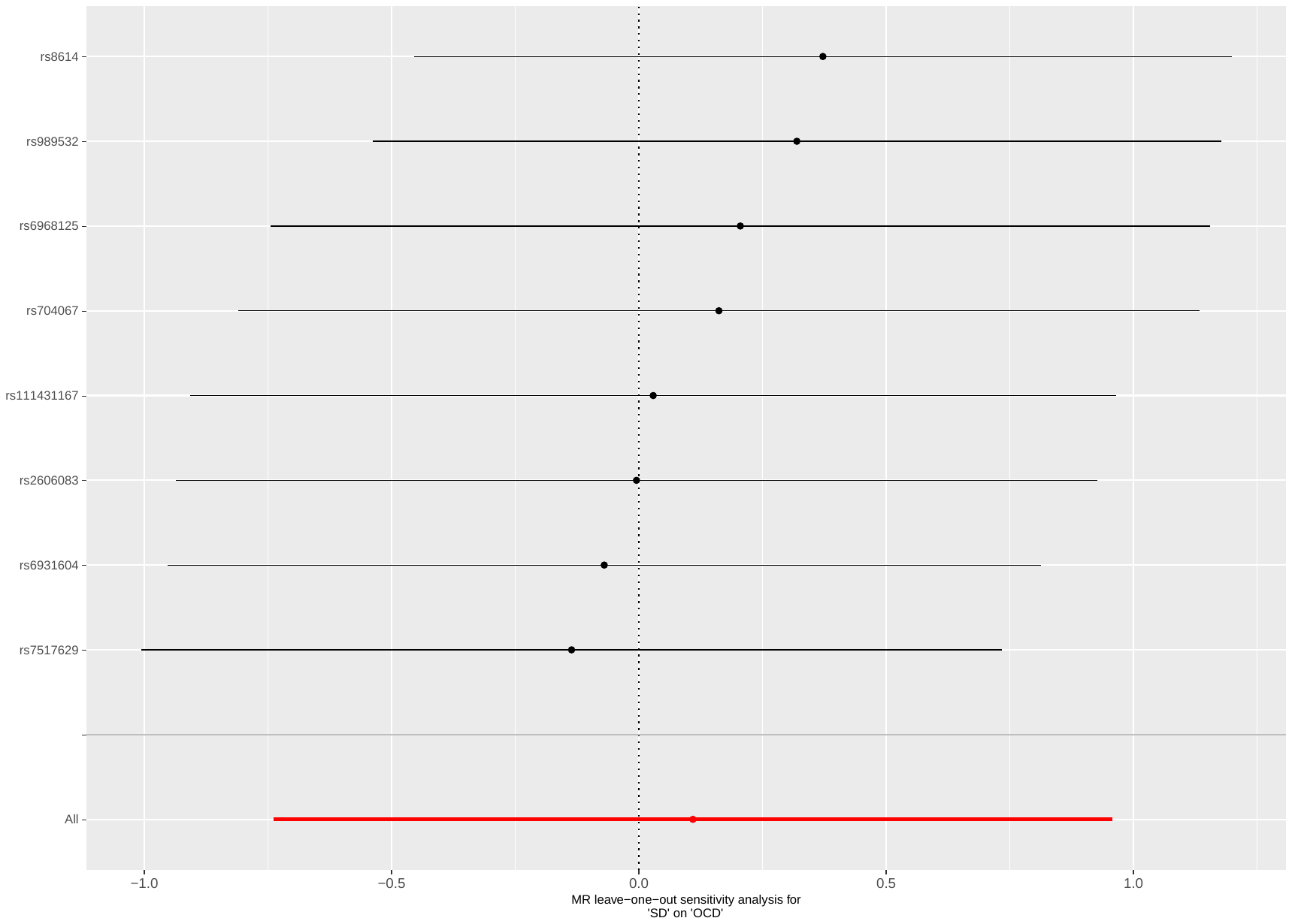

Supplementary Figure 97: leave-one-out analysis of social deprivation against OCD

Abbreviations: MR: Mendelian randomization; SD: social deprivation; OCD: obsessive-compulsive disorder.

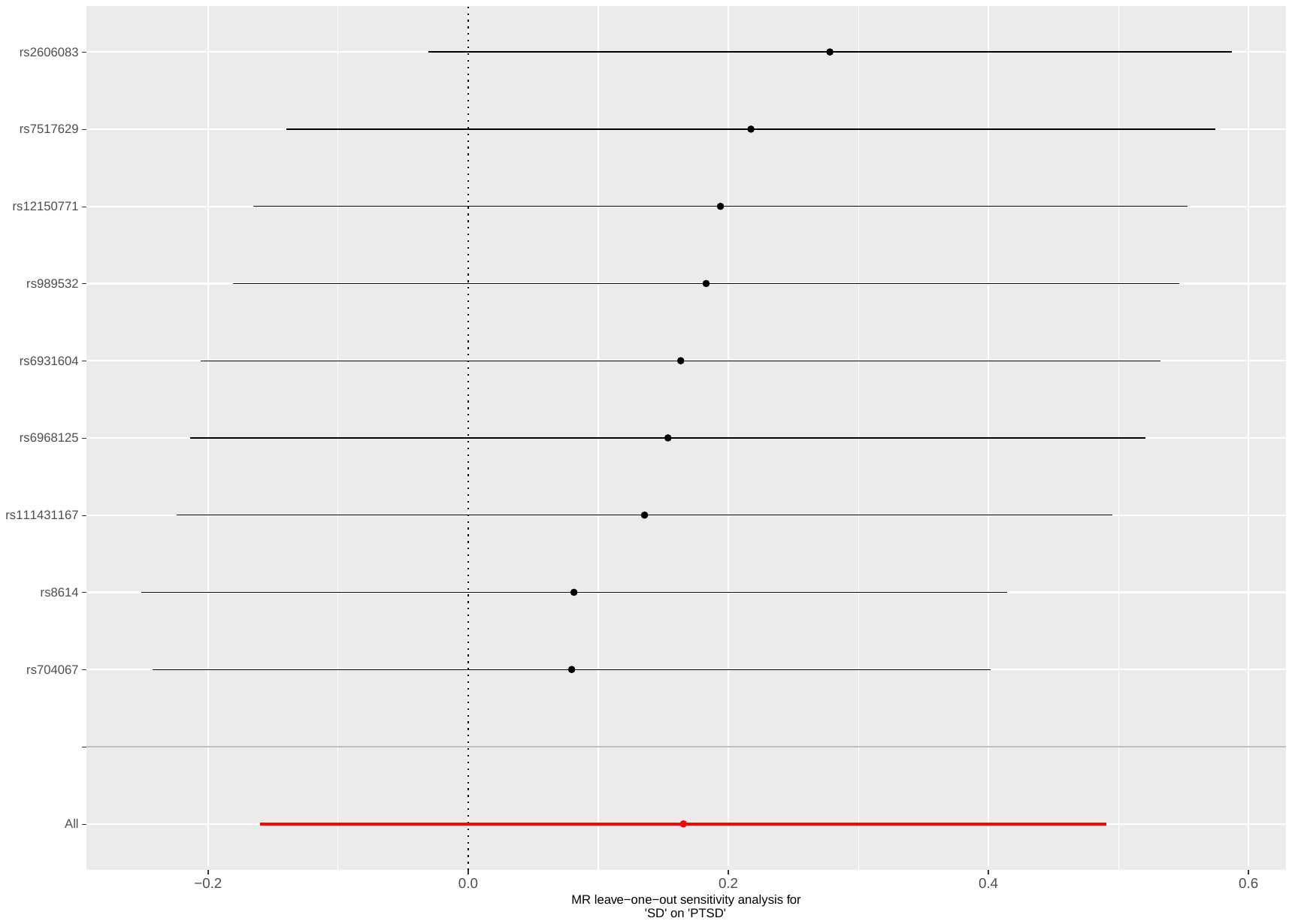

Supplementary Figure 98: leave-one-out analysis of social deprivation against PTSD

Abbreviations: MR: Mendelian randomization; SD: social deprivation; PTSD: post-traumatic stress disorder.

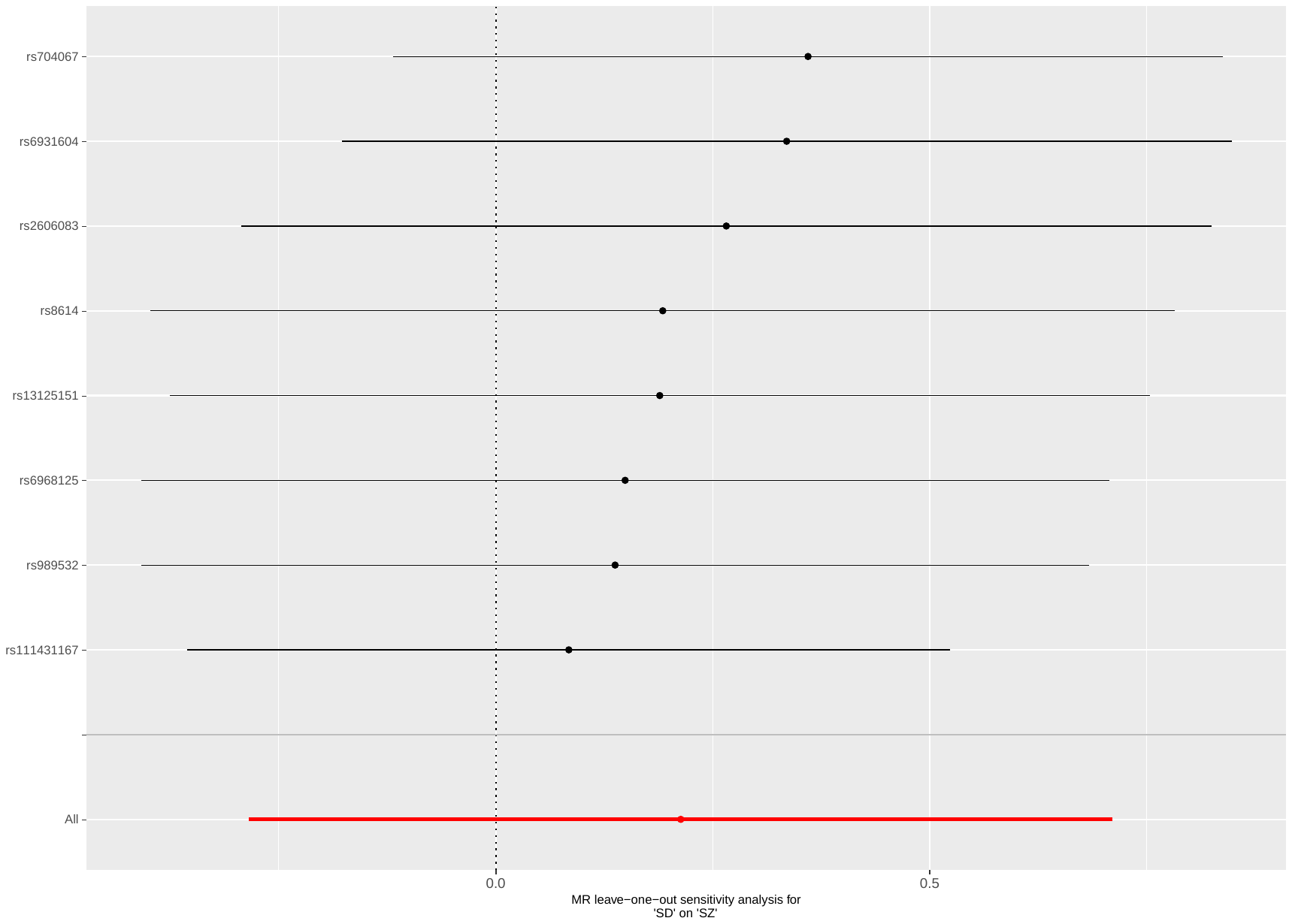

Supplementary Figure 99: leave-one-out analysis of social deprivation against SZ

Abbreviations: MR: Mendelian randomization; SD: social deprivation; SZ: schizophrenia.

#### Plots - Backward analyses

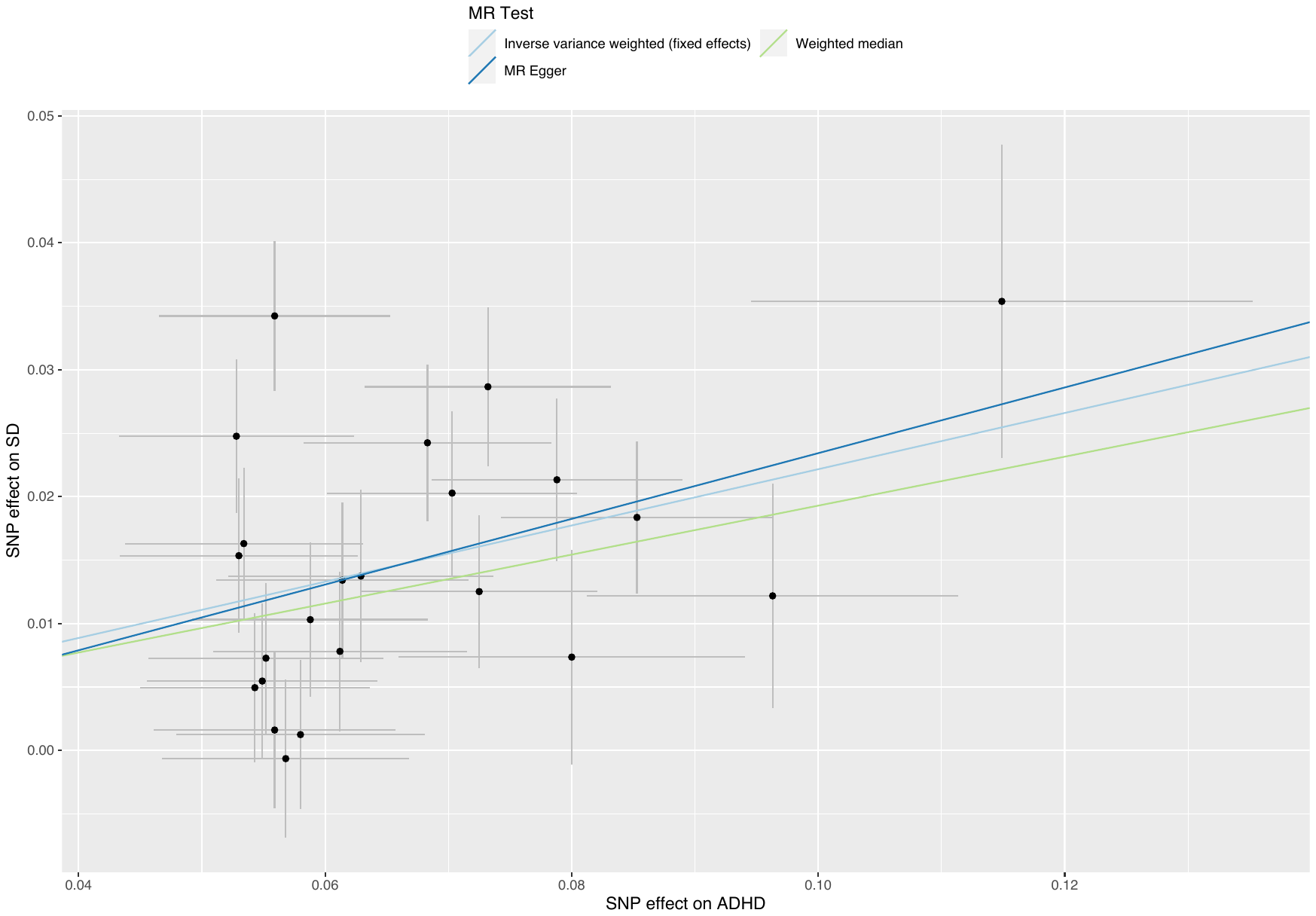

##### Supplementary Figure 100: scatterplot of ADHD against social deprivation

Abbreviations: MR: Mendelian randomization; SD: social deprivation; ADHD: attention deficit hyperactivity disorder.

##### Supplementary Figure 101: scatterplot analysis of AN against social deprivation

Abbreviations: MR: Mendelian randomization; SD: social deprivation; AN: anorexia nervosa.

##### Supplementary Figure 102: scatterplot analysis of BD against social deprivation

Abbreviations: MR: Mendelian randomization; SD: social deprivation; BD: bipolar disorder.

##### Supplementary Figure 103: scatterplot of SZ against social deprivation

Abbreviations: MR: Mendelian randomization; SD: social deprivation; SZ: schizophrenia.

Supplementary Figure 104: leave-one-out analysis of ADHD against social deprivation

Abbreviations: MR: Mendelian randomization; SD: social deprivation; ADHD: attention deficit hyperactivity disorder.

Supplementary Figure 105: leave-one-out analysis of AN against social deprivation

Abbreviations: MR: Mendelian randomization; SD: social deprivation; AN: anorexia nervosa.

Supplementary Figure 106: leave-one-out analysis of BD against social deprivation

Abbreviations: MR: Mendelian randomization; SD: social deprivation; BD: bipolar disorder.

Supplementary Figure 107: leave-one-out analysis of SZ against social deprivation

Abbreviations: MR: Mendelian randomization; SD: social deprivation; SZ: schizophrenia.

#### Supplementary Table 19: CAUSE results of the relations between social deprivation and mental illness

| **Model 1** | **Model 2** | **∆ ELPD** | **SE ∆ ELPD** | **z-score** | **p-value** |
| --- | --- | --- | --- | --- | --- |
| *Fw: SD on ADHD* | | | | | |
| Null  Null  Sharing | Sharing  Causal  Causal | -19.58  -24.85  -5.27 | 5.15  6.50  1.54 | -3.80  -3.82  -3.43 | <0.001  <0.001  <0.001 |
| *Bw: ADHD on SD* | | | | | |
| Null  Null  Sharing | Sharing  Causal  Causal | -40.14  -45.66  -5.52 | 7.96  9.11  1.56 | -5.05  -5.01  -3.54 | <0.001  <0.001  <0.001 |
| *Fw: SD on AN* | | | | | |
| Null  Null  Sharing | Sharing  Causal  Causal | 0.36  0.94  0.57 | 0.09  0.57  0.49 | 3.98  1.64  1.18 | <0.001  0.101  0.238 |
| *Bw: AN on SD* | | | | | |
| Null  Null  Sharing | Sharing  Causal  Causal | 0.53  1.34  0.82 | 0.07  0.15  0.11 | 7.61  9.09  7.40 | <0.001  <0.001  <0.001 |
| *Fw: SD on ANX* | | | | | |
| Null  Null  Sharing | Sharing  Causal  Causal | 0.35  1.00  0.65 | 0.33  0.82  0.51 | 1.09  1.23  1.27 | 0.276  0.219  0.204 |
| *Bw: ANX on SD* | | | | | |
| Null  Null  Sharing | Sharing  Causal  Causal | 0.14  0.61  0.46 | 0.07  0.76  0.70 | 2.12  0.80  0.67 | 0.034  0.424  0.503 |
| *Fw: SD on ASD* | | | | | |
| Null  Null  Sharing | Sharing  Causal  Causal | 0.45  1.26  0.81 | 0.08  0.22  0.19 | 5.84  5.67  4.32 | <0.001  <0.001  <0.001 |
| *Bw: ASD on SD* | | | | | |
| Null  Null  Sharing | Sharing  Causal  Causal | 0.38  1.09  0.72 | 0.09  0.51  0.46 | 4.41  2.14  1.56 | <0.001  0.032  0.119 |
| *Fw: SD on BD* | | | | | |
| Null  Null  Sharing | Sharing  Causal  Causal | -1.00  -4.61  -3.61 | 0.76  2.73  1.97 | -1.30  -1.69  -1.83 | 0.192  0.091  0.067 |
| *Bw: BD on SD* | | | | | |
| Null  Null  Sharing | Sharing  Causal  Causal | 0.01  -1.01  -1.01 | 0.50  2.03  1.54 | 0.01  -0.50  -0.66 | 0.499  0.309  0.256 |
| *Fw: SD on MDD* | | | | | |
| Null  Null  Sharing | Sharing  Causal  Causal | -2.55  5.44  -2.89 | 1.65  3.40  1.80 | -1.55  -1.60  -1.61 | 0.121  0.110  0.107 |
| *Bw: MDD on SD* | | | | | |
| Null  Null  Sharing | Sharing  Causal  Causal | -0.86  -3.39  -2.53 | 0.93  2.77  1.87 | -0.92  -1.23  -1.36 | 0.358  0.219  0.174 |
| *Fw: SD on OCD* | | | | | |
| Null  Null  Sharing | Sharing  Causal  Causal | 0.48  1.40  0.93 | 0.18  0.37  0.20 | 2.67  3.84  4.60 | 0.008  <0.001  <0.001 |
| *Bw: OCD on SD* | | | | | |
| Null  Null  Sharing | Sharing  Causal  Causal | 0.23  0.99  0.76 | 0.07  0.20  0.15 | 3.40  4.90  5.03 | 0.001  <0.001  <0.001 |
| *Fw: SD on PTSD* | | | | | |
| Null  Null  Sharing | Sharing  Causal  Causal | -3.57  -4.38  -0.81 | 2.40  3.39  1.17 | -1.49  -1.29  -0.69 | 0.136  0.197  0.490 |
| *Bw: PTSD on SD* | | | | | |
| Null  Null  Sharing | Sharing  Causal  Causal | 0.16  0.74  0.59 | 0.07  0.65  0.59 | 2.15  1.14  1.00 | 0.032  0.254  0.317 |
| *Fw: SD on SZ* | | | | | |
| Null  Null  Sharing | Sharing  Causal  Causal | -0.77  -4.27  -3.50 | 0.64  2.66  2.02 | -1.19  -1.61  -1.74 | 0.234  0.107  0.082 |
| *Bw: SZ on SD* | | | | | |
| Null  Null  Sharing | Sharing  Causal  Causal | -4.63  -7.90  -3.28 | 2.41  3.99  1.63 | -1.92  -1.98  -2.01 | 0.055  0.048  0.044 |

Abbreviations: MR: mendelian randomization; ELPD: expected log pointwise posterior density; 95%CI: 95% confidence interval; SE: standard error; Fwd: forward MR; Bwd: backward MR; SD: social deprivation; ADHD: attention deficit hyperactivity disorder; AN: anorexia nervosa; ANX: anxiety disorder; ASD: autism spectrum disorder; BD: bipolar disorder; MDD: major depressive disorder; OCD: obsessive-compulsive disorder; PTSD: post-traumatic stress disorder; SZ: schizophrenia.

#### Supplementary Table 20: Results of univariable bidirectional Mendelian Randomization of social deprivation against mental illnesses, after Steiger filtering

| **MR** | **N SNP** | **IVW, B (95% CI)** | **p-value** | **WM, B (95% CI)** | **p-value** | **MR-Egger, B (95% CI)** | **p-value** | **Egger intercept p-value** | **Mean F** |
| --- | --- | --- | --- | --- | --- | --- | --- | --- | --- |
| SD on ADHD | 6 | 0.557 (0.331; 0.784) | <0.001 | 0.526 (0.205; 0.847) | 0.001 | 0.326 (-1.95; 2.60) | 0.793 | 0.088 | 31.8 |
| SD on AN | 9 | -0.188 (-0.470; 0.093) | 0.189 | -0.281 (-0.664; 0.102) | 0.150 | -0.761 (-3.10; 1.57) | 0.543 | 0.299 | 33.0 |
| SD on ANX | 9 | 0.165 (-0.399; 0.730) | 0.566 | 0.261 (-0.486; 1.01) | 0.494 | 0.904 (-3.63; 5.44) | 0.708 | 0.355 | 33.0 |
| SD on ASD | 7 | 0.028 (-0.276; 0.332) | 0.855 | 0.169 (-0.240; 0.579) | 0.418 | -1.19 (-4.27; 1.89) | 0.483 | 0.027 | 33.3 |
| SD on BD | 5 | -0.124 (-0.374; 0.127) | 0.334 | -0.152 (-0.462; 0.158) | 0.337 | -0.418 (-1.90; 1.07) | 0.620 | 0.880 | 32.3 |
| SD on MDD | 8 | 0.141 (-0.046; 0.328) | 0.138 | 0.200 (-0.060; 0.460) | 0.132 | 0.370 (-1.80; 2.54) | 0.749 | 0.888 | 33.4 |
| SD on OCD | 2 | -0.422 (1.84; 0.995) | 0.559 | NR | NR | NR | NR | NR | 33.0 |
| SD on PTSD | 9 | 0.165 (-0.127; 0.458) | 0.268 | 0.190 (-0.232; 0.612) | 0.377 | 1.38 (-1.37; 4.12) | 0.358 | 0.424 | 33.1 |
| SD on SZ | 7 | 0.084 (-0.091; 0.259) | 0.345 | 0.247 (-0.059; 0.553) | 0.114 | 1.18 (-1.76; 4.12) | 0.468 | 0.686 | 32.1 |

Abbreviations: SD: social deprivation; ADHD: attention deficit hyperactivity disorder; AN: anorexia nervosa; ANX: anxiety disorder; ASD: autism spectrum disorders; BD: bipolar disorder; MDD: major depressive disorder; OCD: obsessive-compulsive disorder; PTSD: post-traumatic stress disorder; SZ: schizophrenia; MR: mendelian randomization; SNP: single nucleotide polymorphism; IVW: inverse variance weighted (fixed effect); B: effect estimates are log-odds; 95% CI: 95% confidence interval; WM: weighted median; NR: not reported because not enough SNP to perform MR.

### **Univariable Mendelian randomization of household income levels and mental illnesses**

#### Supplementary Table 21: results of univariable bidirectional Mendelian randomization of Low Household income (LHI) against mental illnesses

| **MR** | **N SNP** | **IVW, B (95% CI)** | **p-value** | **IVW Q(df)** | **Q p-value** | **WM, B (95% CI)** | **p-value** | **MR-Egger, B (95% CI)** | **p-value** | **Egger intercept p-value** | **Steiger Test p-value** | **MR-PRESSO** | **Mean F** |
| --- | --- | --- | --- | --- | --- | --- | --- | --- | --- | --- | --- | --- | --- |
| Fw: LHI on ADHD | 8 | 0.610 (0.415; 0.805) | <0.001 | 8 (7) | 0.338 | 0.569 (0.286; 0.853) | <0.001 | 0.695 (-0.288; 1.68) | 0.215 | 0.867 | <0.001 | GT; p=0.364 | 33.5 |
| Bw: ADHD on LHI | 23 | 0.210 (0.170; 0.250) | <0.001 | 44 (22) | 0.004 | 0.203 (0.136; 0.270) | <0.001 | -0.042 (-0.353; 0.268) | 0.792 | 0.120 | <0.001 | GT; p=0.444 | 39.2 |
| Fw: LHI on AN | 11 | -0.092 (-0.333; 0.148) | 0.452 | 16 (10) | 0.106 | 0.018 (-0.329; 0.365) | 0.919 | -0.094 (-1.58; 1.39) | 0.904 | 0.998 | <0.001 | GT; p=0.102 | 33.7 |
| Bw: AN on LHI | 4 | 0.026 (-0.049; 0.100) | 0.500 | 7 (3) | 0.067 | 0.016 (-0.073; 0.106) | 0.725 | -0.519 (-1.52; 0.484) | 0.417 | 0.396 | NR ^b^ | GT; p=0.133 | 31.9 |
| Fw: LHI on ANX | 10 | 0.628 (0.095; 1.16) | 0.021 | 6 (9) | 0.734 | 0.700 (0.016; 1.38) | 0.045 | 0.300 (-3.08; 3.68) | 0.866 | 0.852 | 0.012 | GT; p=0.767 | 33.6 |
| Bw: ANX on LHI | 0 | NR ^c^ | NR ^c^ | NR ^c^ | NR ^c^ | NR ^c^ | NR ^c^ | NR ^c^ | NR ^c^ | NR ^c^ | NR ^c^ | NR ^c^ | NR ^c^ |
| Fw: LHI on ASD | 12 | 0.177 (-0.053; 0.408) | 0.132 | 45 (11) | <0.001 | 0.228 (-0.145; 0.602) | 0.231 | 0.407 (-1.82; 2.63) | 0.727 | 0.840 | NR ^b^ | DT; p=0.575 | 33.5 |
| Bw: ASD on LHI | 0 | NR ^c^ | NR ^c^ | NR ^c^ | NR ^c^ | NR ^c^ | NR ^c^ | NR ^c^ | NR ^c^ | NR ^c^ | NR ^c^ | NR ^c^ | NR ^c^ |
| Fw: LHI on BD | 9 | 0.306 (0.125; 0.487) | 0.001 | 13 (8) | 0.106 | 0.159 (-0.108; 0.427) | 0.243 | -0.545 (-1.42; 0.332) | 0.263 | 0.092 | <0.001 | GT; p=0.141 | 33.7 |
| Bw: BD on LHI | 36 | -0.001 (-0.032; 0.032) | 0.983 | 91 (35) | <0.001 | 0.029 (-0.023; 0.081) | 0.275 | 0.125 (-0.158; 0.407) | 0.394 | 0.384 | NR ^b^ | DT; p=0.215 | 39.2 |
| Fw: LHI on MDD | 10 | 0.351 (0.189; 0.513) | <0.001 | 13 (9) | 0.175 | 0.398 (0.058; 0.558) | 0.016 | 0.880 (0.082; 1.68) | 0.006 | 0.218 | <0.001 | GT; p=0.207 | 33.5 |
| Bw: MDD on LHI | 0 | NR ^c^ | NR ^c^ | NR ^c^ | NR ^c^ | NR ^c^ | NR ^c^ | NR ^c^ | NR ^c^ | NR ^c^ | NR ^c^ | NR ^c^ | NR ^c^ |
| Fw: LHI on OCD | 10 | -0.108 (-0.738; 0.522) | 0.737 | 11 (9) | 0.306 | 0.293 (-0.536; 1.12) | 0.488 | 0.593 (-2.60; 3.78) | 0.725 | 0.670 | NR ^b^ | GT; p=0.309 | 33.5 |
| Bw: OCD on LHI | 0 | NR ^c^ | NR ^c^ | NR ^c^ | NR ^c^ | NR ^c^ | NR ^c^ | NR ^c^ | NR ^c^ | NR ^c^ | NR ^c^ | NR ^c^ | NR ^c^ |
| Fw: LHI on PTSD | 12 | 0.506 (0.253; 0.759) | <0.001 | 8 (11) | 0.672 | 0.511 (0.167; 0.855) | 0.004 | 0.412 (-0.769; 1.59) | 0.510 | 0.877 | <0.001 | GT; p=0.716 | 33.5 |
| Bw: PTSD on LHI | 0 | NR ^c^ | NR ^c^ | NR ^c^ | NR ^c^ | NR ^c^ | NR ^c^ | NR ^c^ | NR ^c^ | NR ^c^ | NR ^c^ | NR ^c^ | NR ^c^ |
| Fw: LHI on SZ | 9 | 0.648 (0.488; 0.808) | <0.001 | 58 (8) | <0.001 | 0.400 (0.102; 0.699) | 0.009 | -0.151 (-2.16; 1.86) | 0.887 | 0.452 | 0.421 | DT; p=0.783 | 33.7 |
| Bw: SZ on LHI | 176 | 0.082 (0.066; 0.098) | <0.001 | 439 (175) | <0.001 | 0.071 (0.045; 0.098) | <0.001 | 0.115 (0.016; 0.214) | 0.024 | 0.511 | <0.001 | DT; p=0.543 | 45.6 |

Abbreviations: Fw: forward analysis; Bw: backward analysis; LHI: low household income (<£18,000); ADHD: attention deficit hyperactivity disorder; AN: anorexia nervosa; ANX: anxiety disorder; ASD: autism spectrum disorder; BD: bipolar disorder; MDD: major depressive disorder; OCD: obsessive-compulsive disorder; PTSD: post-traumatic stress disorder; SZ: schizophrenia; MR: mendelian randomization; SNP: single nucleotide polymorphism; IVW: inverse variance weighted (fixed effect); B: effect estimates are log-odds; 95% CI: 95% confidence interval; Q: Cochran’s Q measure of heterogeneity; df: degree of freedom; WM: weighted median; DT: distortion test; GT: global test.

Legend:

P-value threshold <5e-8

The phenotypes are analyzed as binary cases (coded as 1) and controls (coded as 0). Do note that this leads to change in the direction of effect when comparing across some of the traits. LHI = class 1 (i.e., HI<£18,000, cases) vs classes 2,3,4,5 (controls)

^a^ The Mendelian randomization pleiotropy residual sum and outlier (MR-PRESSO) test identifies possible bias from horizontal pleiotropy. The test consists of three parts, (1) the MR-PRESSO global test which detects horizontal pleiotropy, (2) the outlier corrected causal estimate which corrects for the detected horizontal pleiotropy and (3) the MR-PRESSO distortion test which estimates if the causal estimate is significantly different (at p<0.05) after adjustment for outliers. We conduct all three stages (with the argument NbDistribution=1000, namely using1000 simulation form the null distribution to compute empirical p-values) and present the outlier adjusted causal estimates (OACE) when both global and distortion tests are significant.

^b^ We did not run Steiger Test if none of the MR analysis resulted significant (NR: not reported in the cell).

^c^ Not enough SNP to perform MR (NR: not reported in the cell).

#### Supplementary Table 22: results of univariable bidirectional Mendelian randomization of Low-Mid Household income (LMHI) against mental illnesses

| **MR** | **N SNP** | **IVW, B (95% CI)** | **p-value** | **IVW Q(df)** | **Q p-value** | **WM, B (95% CI)** | **p-value** | **MR-Egger, B (95% CI)** | **p-value** | **Egger intercept p-value** | **Steiger Test p-value** | **MR-PRESSO** | **Mean F** |
| --- | --- | --- | --- | --- | --- | --- | --- | --- | --- | --- | --- | --- | --- |
| Fw: LMHI on ADHD | 15 | 0.356 (0.198; 0.513) | <0.001 | 57 (14) | <0.001 | 0.161 (-0.075; 0.398) | 0.181 | -1.00 (-3.51; 1.51) | 0.448 | 0.304 | <0.001 | DT; p=0.115 | 35.8 |
| Bw: ADHD on LMHI | 23 | 0.174 (0.140; 0.208) | <0.001 | 74 (22) | <0.001 | 0.166 (0.107; 0.226) | <0.001 | -0.015 (-0.373; 0.343) | 0.936 | 0.304 | <0.001 | DT; p=0.852 | 39.2 |
| Fw: LMHI on AN | 17 | -0.309 (-0.526; -0.091) | 0.005 | 40 (16) | <0.001 | -0.222 (-0.542; 0.099) | 0.176 | -1.35 (-4.23; 1.53) | 0.373 | 0.486 | <0.001 | DT; p=0.160 | 35.3 |
| Bw: AN on LMHI | 4 | 0.026 (-0.038; 0.090) | 0.426 | 8 (3) | 0.047 | 0.023 (-0.056; 0.102) | 0.564 | -0.652 (-1.29; -0.014) | 0.183 | 0.171 | NR ^b^ | GT; p=0.103 | 31.9 |
| Fw: LMHI on ANX | 17 | 0.376 (-0.078; 0.831) | 0.105 | 9 (16) | 0.930 | 0.189 (-0.387; 0.765) | 0.520 | -0.884 (-4.80; 3.03) | 0.664 | 0.535 | NR ^b^ | GT; p=0.952 | 35.3 |
| Bw: ANX on LMHI | 0 | NR ^c^ | NR ^c^ | NR ^c^ | NR ^c^ | NR ^c^ | NR ^c^ | NR ^c^ | NR ^c^ | NR ^c^ | NR ^c^ | NR ^c^ | NR ^c^ |
| Fw: LMHI on ASD | 18 | -0.268 (-0.488; -0.047) | 0.017 | 55 (17) | <0.001 | -0.374 (-0.753; 0.005) | 0.053 | -2.15 (-5.04; 0.746) | 0.165 | 0.217 | 0.994 | DT; p=0.275 | 33.4 |
| Bw: ASD on LMHI | 0 | NR ^c^ | NR ^c^ | NR ^c^ | NR ^c^ | NR ^c^ | NR ^c^ | NR ^c^ | NR ^c^ | NR ^c^ | NR ^c^ | NR ^c^ | NR ^c^ |
| Fw: LMHI on BD | 16 | -0.099 (-0.251; 0.053) | 0.201 | 95 (15) | <0.001 | 0.067 (-0.352; 0.217) | 0.643 | -2.40 (-5.36; 0.553) | 0.133 | 0.146 | NR ^b^ | DT; p=0.255 | 35.6 |
| Bw: BD on LMHI | 36 | -0.030 (-0.058; -0.003) | 0.032 | 138 (35) | <0.001 | -0.030 (-0.076; 0.017) | 0.208 | -0.001 (-0.302; 0.301) | 0.999 | 0.844 | <0.001 | DT; p=0.226 | 39.2 |
| Fw: LMHI on MDD | 17 | 0.224 (0.077; 0.371) | 0.003 | 19 (16) | 0.281 | 0.204 (-0.004; 0.411) | 0.054 | -0.159 (-1.39; 1.07) | 0.803 | 0.547 | <0.001 | GT; p=0.278 | 33.5 |
| Bw: MDD on LMHI | 0 | NR ^c^ | NR ^c^ | NR ^c^ | NR ^c^ | NR ^c^ | NR ^c^ | NR ^c^ | NR ^c^ | NR ^c^ | NR ^c^ | NR ^c^ | NR ^c^ |
| Fw: LMHI on OCD | 17 | -0.774 (-1.34; -0.212) | 0.007 | 11 (16) | 0.795 | -0.833 (-1.57; -0.097) | 0.027 | -0.350 (-4.75; 4.05) | 0.878 | 0.852 | 0.249 | GT; p=0.781 | 33.5 |
| Bw: OCD on LMHI | 0 | NR ^c^ | NR ^c^ | NR ^c^ | NR ^c^ | NR ^c^ | NR ^c^ | NR ^c^ | NR ^c^ | NR ^c^ | NR ^c^ | NR ^c^ | NR ^c^ |
| Fw: LMHI on PTSD | 17 | 0.283 (0.045; 0.521) | 0.020 | 24 (16) | 0.087 | 0.285 (-0.071; 0.640) | 0.117 | -0.485 (-2.89; 1.92) | 0.698 | 0.537 | <0.001 | GT; p=0.096 | 35.3 |
| Bw: PTSD on LMHI | 0 | NR ^c^ | NR ^c^ | NR ^c^ | NR ^c^ | NR ^c^ | NR ^c^ | NR ^c^ | NR ^c^ | NR ^c^ | NR ^c^ | NR ^c^ | NR ^c^ |
| Fw: LMHI on SZ | 16 | 0.270 (0.138; 0.401) | <0.001 | 122 (15) | <0.001 | 0.117 (-0.132; 0.366) | 0.357 | -0.273 (-3.42; 2.87) | 0.867 | 0.738 | <0.001 | DT; p=0.361 | 35.6 |
| Bw: SZ on LMHI | 176 | 0.045 (0.031; 0.059) | <0.001 | 436 (175) | <0.001 | 0.041 (0.018; 0.063) | <0.001 | 0.032 (-0.052; 0.117) | 0.455 | 0.768 | <0.001 | DT; p=0.775 | 45.6 |

Abbreviations: Fw: forward analysis; Bw: backward analysis; LMHI: low-mid household income (≤£30,999); ADHD: attention deficit hyperactivity disorder; AN: anorexia nervosa; ANX: anxiety disorder; ASD: autism spectrum disorder; BD: bipolar disorder; MDD: major depressive disorder; OCD: obsessive-compulsive disorder; PTSD: post-traumatic stress disorder; SZ: schizophrenia; MR: mendelian randomization; SNP: single nucleotide polymorphism; IVW: inverse variance weighted (fixed effect); B: effect estimates are log-odds; 95% CI: 95% confidence interval; Q: Cochran’s Q measure of heterogeneity; df: degree of freedom; WM: weighted median; DT: distortion test; GT: global test.

Legend:

P-value threshold <5e-8

The phenotypes are analyzed as binary cases (coded as 1) and controls (coded as 0). Do note that this leads to change in the direction of effect when comparing across some of the traits. LMHI = cases: classes 1 (i.e., HI<£18,000) and 2 (i.e., £18,000≤HI≤£29,000) vs controls: classes 3,4,5.

^a^ The Mendelian randomization pleiotropy residual sum and outlier (MR-PRESSO) test identifies possible bias from horizontal pleiotropy. The test consists of three parts, (1) the MR-PRESSO global test which detects horizontal pleiotropy, (2) the outlier corrected causal estimate which corrects for the detected horizontal pleiotropy and (3) the MR-PRESSO distortion test which estimates if the causal estimate is significantly different (at p<0.05) after adjustment for outliers. We conduct all three stages (with the argument NbDistribution=1000, namely using1000 simulation form the null distribution to compute empirical p-values) and present the outlier adjusted causal estimates (OACE) when both global and distortion tests are significant.

^b^ We did not run Steiger Test if none of the MR analysis resulted significant (NR: not reported in the cell).

^c^ Not enough SNP to perform MR (NR: not reported in the cell).

#### Supplementary Table 23: results of univariable bidirectional Mendelian randomization of Mid-High Household income (MHHI) against mental illnesses

| **MR** | **N SNP** | **IVW, B (95% CI)** | **p-value** | **IVW Q(df)** | **Q p-value** | **WM, B (95% CI)** | **p-value** | **MR-Egger, B (95% CI)** | **p-value** | **Egger intercept p-value** | **Steiger Test p-value** | **MR-PRESSO** | **Mean F** |
| --- | --- | --- | --- | --- | --- | --- | --- | --- | --- | --- | --- | --- | --- |
| Fw: MHHI on ADHD | 20 | -0.298 (-0.422; -0.174) | <0.001 | 49 (19) | <0.001 | -0.112 (-0.312; 0.088) | 0.272 | -0.198 (-1.11; 0.709) | 0.674 | 0.827 | <0.001 | OACE: -0.182 (-0.342; -0.023); p=0.038 | 36.4 |
| Bw: ADHD on MHHI | 23 | -0.160 (-0.199; -0.122) | <0.001 | 45 (22) | 0.002 | -0.155 (-0.216; -0.093) | <0.001 | 0.027 (-0.280; 0.333) | 0.867 | 0.238 | <0.001 | DT; p=0.549 | 39.2 |
| Fw: MHHI on AN | 20 | 0.371 (0.193; 0.550) | <0.001 | 49 (19) | <0.001 | 0.281 (-0.018; 0.581) | 0.066 | 1.05 (-0.224; 2.33) | 0.123 | 0.297 | <0.001 | OACE: 0.181 (-0.058; 0.420); p=0.156 | 36.6 |
| Bw: AN on MHHI | 4 | -0.009 (-0.081; 0.062) | 0.803 | 7 (3) | 0.066 | 0.036 (-0.052; 0.124) | 0.425 | 0.314 (-0.811; 1.44) | 0.639 | 0.628 | NR ^b^ | GT; p=0.130 | 31.9 |
| Fw: MHHI on ANX | 20 | -0.320 (-0.694; 0.053) | 0.093 | 15 (19) | 0.691 | -0.098 (-0.583; 0.389) | 0.694 | -0.951 (-3.27; 1.36) | 0.431 | 0.595 | NR ^b^ | GT; p=0.722 | 36.4 |
| Bw: ANX on MHHI | 0 | NR ^c^ | NR ^c^ | NR ^c^ | NR ^c^ | NR ^c^ | NR ^c^ | NR ^c^ | NR ^c^ | NR ^c^ | NR ^c^ | NR ^c^ | NR ^c^ |
| Fw: MHHI on ASD | 21 | 0.480 (0.303; 0.658) | <0.001 | 36 (20) | 0.015 | 0.393 (0.093; 0.692) | 0.010 | 0.345 (-0.763; 1.45) | 0.549 | 0.809 | 0.057 | DT; p=0.208 | 36.2 |
| Bw: ASD on MHHI | 0 | NR ^c^ | NR ^c^ | NR ^c^ | NR ^c^ | NR ^c^ | NR ^c^ | NR ^c^ | NR ^c^ | NR ^c^ | NR ^c^ | NR ^c^ | NR ^c^ |
| Fw: MHHI on BD | 19 | 0.179 (0.053; 0.304) | 0.005 | 125 (18) | <0.001 | 0.028 (-0.208; 0.264) | 0.816 | 1.69 (0.376; 3.00) | 0.022 | 0.033 | <0.001 | DT; p=0.092 | 36.8 |
| Bw: BD on MHHI | 36 | 0.040 (0.009; 0.071) | 0.011 | 133 (35) | <0.001 | 0.030 (-0.020; 0.081) | 0.239 | 0.025 (-0.308; 0.357) | 0.885 | 0.927 | <0.001 | OACE: 0.002 (-0.039; 0.042); p=0.943 | 39.2 |
| Fw: MHHI on MDD | 21 | -0.123 (-0.238; -0.008) | 0.036 | 34 (20) | 0.025 | -0.070 (-0.245; 0.105) | 0.433 | 0.474 (-0.178; 1.13) | 0.171 | 0.082 | <0.001 | DT; p=0.256 | 36.2 |
| Bw: MDD on MHHI | 0 | NR ^c^ | NR ^c^ | NR ^c^ | NR ^c^ | NR ^c^ | NR ^c^ | NR ^c^ | NR ^c^ | NR ^c^ | NR ^c^ | NR ^c^ | NR ^c^ |
| Fw: MHHI on OCD | 21 | -0.079 (-0.511; 0.353) | 0.720 | 23 (20) | 0.265 | -0.065 (-0.679; 0.549) | 0.836 | -0.399 (-2.63; 1.83) | 0.729 | 0.776 | NR ^b^ | GT; p=0.271 | 36.2 |
| Bw: OCD on MHHI | 0 | NR ^c^ | NR ^c^ | NR ^c^ | NR ^c^ | NR ^c^ | NR ^c^ | NR ^c^ | NR ^c^ | NR ^c^ | NR ^c^ | NR ^c^ | NR ^c^ |
| Fw: MHHI on PTSD | 21 | -0.233 (-0.421; -0.044) | 0.016 | 25 (20) | 0.184 | -0.195 (-0.471; 0.080) | 0.165 | -0.080 (-1.06; 0.900) | 0.874 | 0.758 | <0.001 | GT; p=0.197 | 36.2 |
| Bw: PTSD on MHHI | 0 | NR ^c^ | NR ^c^ | NR ^c^ | NR ^c^ | NR ^c^ | NR ^c^ | NR ^c^ | NR ^c^ | NR ^c^ | NR ^c^ | NR ^c^ | NR ^c^ |
| Fw: MHHI on SZ | 20 | -0.098 (-0.201; 0.005) | 0.063 | 143 (19) | <0.001 | -0.171 (-0.376; 0.033) | 0.100 | -0.236 (-1.60; 1.12) | 0.738 | 0.841 | NR ^b^ | DT; p=0.574 | 36.4 |
| Bw: SZ on MHHI | 176 | -0.046 (-0.061; -0.030) | <0.001 | 432 (175) | <0.001 | -0.039 (-0.066; -0.012) | 0.005 | 0.018 (-0.077; 0.112) | 0.707 | 0.172 | <0.001 | DT; p=0.431 | 45.6 |

Abbreviations: Fw: forward analysis; Bw: backward analysis; MHHI: mid-high household income (≥ £52,000); ADHD: attention deficit hyperactivity disorder; AN: anorexia nervosa; ANX: anxiety disorder; ASD: autism spectrum disorder; BD: bipolar disorder; MDD: major depressive disorder; OCD: obsessive-compulsive disorder; PTSD: post-traumatic stress disorder; SZ: schizophrenia; MR: mendelian randomization; SNP: single nucleotide polymorphism; IVW: inverse variance weighted (fixed effect); B: effect estimates are log-odds; 95% CI: 95% confidence interval; Q: Cochran’s Q measure of heterogeneity; df: degree of freedom; WM: weighted median; DT: distortion test; GT: global test.

Legend:

P-value threshold <5e-8

The phenotypes are analyzed as binary cases (coded as 1) and controls (coded as 0). Do note that this leads to change in the direction of effect when comparing across some of the traits. MHHI = controls: classes 1, 2, and 3 vs cases: classes 4 (i.e., £52,000 ≤HI≤£100,000) and 5 (i.e., HI>£100,000)

^a^ The Mendelian randomization pleiotropy residual sum and outlier (MR-PRESSO) test identifies possible bias from horizontal pleiotropy. The test consists of three parts, (1) the MR-PRESSO global test which detects horizontal pleiotropy, (2) the outlier corrected causal estimate which corrects for the detected horizontal pleiotropy and (3) the MR-PRESSO distortion test which estimates if the causal estimate is significantly different (at p<0.05) after adjustment for outliers. We conduct all three stages (with the argument NbDistribution=1000, namely using1000 simulation form the null distribution to compute empirical p-values) and present the outlier adjusted causal estimates (OACE) when both global and distortion tests are significant.

^b^ We did not run Steiger Test if none of the MR analysis resulted significant (NR: not reported in the cell).

^c^ Not enough SNP to perform MR (NR: not reported in the cell).

#### Supplementary Table 24: results of univariable bidirectional Mendelian randomization of High Household income (HHI) against mental illness

| **MR** | **N SNP** | **IVW, B (95% CI)** | **p-value** | **IVW Q(df)** | **Q p-value** | **WM, B (95% CI)** | **p-value** | **MR-Egger, B (95% CI)** | **p-value** | **Egger intercept p-value** | **Steiger Test p-value** | **MR-PRESSO** | **Mean F** |
| --- | --- | --- | --- | --- | --- | --- | --- | --- | --- | --- | --- | --- | --- |
| Fw: HHI on ADHD | 2 | -0.033 (-0.216; 0.149) | 0.720 | 0.4 (2) | 0.513 | NR ^c^ | NR ^c^ | NR ^c^ | NR ^c^ | NR ^c^ | <0.001 | NR ^c^ | 45.1 |
| Bw: ADHD on HHI | 23 | -0.193 (-0.266; -0.120) | <0.001 | 42 (22) | 0.007 | -0.211 (-0.322; -0.100) | <0.001 | 0.085 (-0.464; 0.633) | 0.765 | 0.324 | <0.001 | DT; p=0.982 | 39.2 |
| Fw: HHI on AN | 2 | 0.333 (0.065; 0.600) | 0.015 | 2 (1) | 0.195 | NR ^c^ | NR ^c^ | NR ^c^ | NR ^c^ | NR ^c^ | 0.183 | NR ^c^ | 45.1 |
| Bw: AN on HHI | 4 | -0.043 (-0.177; 0.092) | 0.535 | 2 (3) | 0.544 | -0.036 (-0.196; 0.125) | 0.663 | -0.113 (-1.36; 1.13) | 0.875 | 0.921 | NR ^b^ | GT; p=0.581 | 31.9 |
| Fw: HHI on ANX | 2 | -0.101 (-0.641; 0.440) | 0.715 | 0.004 (1) | 0.949 | NR ^c^ | NR ^c^ | NR ^c^ | NR ^c^ | NR ^c^ | NR ^b^ | NR ^c^ | 45.1 |
| Bw: ANX on HHI | 0 | NR ^c^ | NR ^c^ | NR ^c^ | NR ^c^ | NR ^c^ | NR ^c^ | NR ^c^ | NR ^c^ | NR ^c^ | NR ^c^ | NR ^c^ | NR ^c^ |
| Fw: HHI on ASD | 3 | 0.094 (-0.121; 0.308) | 0.392 | 40 (2) | <0.001 | 0.509 (0.163; 0.855) | 0.004 | -4.07 (-6.29; 1.85) | 0.173 | 0.167 | 0.031 | NR ^c^ | 46.8 |
| Bw: ASD on HHI | 0 | NR ^c^ | NR ^c^ | NR ^c^ | NR ^c^ | NR ^c^ | NR ^c^ | NR ^c^ | NR ^c^ | NR ^c^ | NR ^c^ | NR ^c^ | NR ^c^ |
| Fw: HHI on BD | 2 | 0.608 (0.423; 0.792) | <0.001 | 1 (1) | 0.283 | NR ^c^ | NR ^c^ | NR ^c^ | NR ^c^ | NR ^c^ | 0.015 | NR ^c^ | 45.1 |
| Bw: BD on HHI | 36 | 0.076 (0.017; 0.134) | 0.011 | 87 (35) | <0.001 | 0.066 (-0.026; 0.159) | 0.158 | 0.065 (-0.443; 0.573) | 0.803 | 0.968 | <0.001 | DT; p=0.117 | 39.2 |
| Fw: HHI on MDD | 2 | 0.013 (-0.163; 0.188) | 0.889 | 2 (1) | 0.219 | NR ^c^ | NR ^c^ | NR ^c^ | NR ^c^ | NR ^c^ | NR ^c^ | NR ^c^ | 45.1 |
| Bw: MDD on HHI | 0 | NR ^c^ | NR ^c^ | NR ^c^ | NR ^c^ | NR ^c^ | NR ^c^ | NR ^c^ | NR ^c^ | NR ^c^ | NR ^c^ | NR ^c^ | NR ^c^ |
| Fw: HHI on OCD | 2 | 0.199 (-0.469; 0.866) | 0.560 | 2 (1) | 0.180 | NR ^c^ | NR ^c^ | NR ^c^ | NR ^c^ | NR ^c^ | NR ^c^ | NR ^c^ | 45.1 |
| Bw: OCD on HHI | 0 | NR ^c^ | NR ^c^ | NR ^c^ | NR ^c^ | NR ^c^ | NR ^c^ | NR ^c^ | NR ^c^ | NR ^c^ | NR ^c^ | NR ^c^ | NR ^c^ |
| Fw: HHI on PTSD | 3 | -0.190 (-0.425; 0.046) | 0.114 | 1 (2) | 0.569 | -0.157 (-0.453; 0.139) | 0.300 | -0.887 (-2.40; 0.626) | 0.456 | 0.528 | NR ^b^ | NR ^c^ | 46.8 |
| Bw: PTSD on HHI | 0 | NR ^c^ | NR ^c^ | NR ^c^ | NR ^c^ | NR ^c^ | NR ^c^ | NR ^c^ | NR ^c^ | NR ^c^ | NR ^c^ | NR ^c^ | NR ^c^ |
| Fw: HHI on SZ | 2 | 0.030 (-0.121; 0.182) | 0.693 | 14 (1) | <0.001 | NR ^c^ | NR ^c^ | NR ^c^ | NR ^c^ | NR ^c^ | NR ^c^ | NR ^c^ | 45.1 |
| Bw: SZ on HHI | 176 | -0.013 (-0.042; 0.016) | 0.395 | 295 (175) | <0.001 | 0.002 (-0.049; 0.052) | 0.950 | 0.095 (-0.051; 0.242) | 0.205 | 0.138 | NR ^c^ | DT; p=0.195 | 45.6 |

Abbreviations: Fw: forward analysis; Bw: backward analysis; HHI: high household income (> £100,000); ADHD: attention deficit hyperactivity disorder; AN: anorexia nervosa; ANX: anxiety disorder; ASD: autism spectrum disorder; BD: bipolar disorder; MDD: major depressive disorder; OCD: obsessive-compulsive disorder; PTSD: post-traumatic stress disorder; SZ: schizophrenia; MR: mendelian randomization; SNP: single nucleotide polymorphism; IVW: inverse variance weighted (fixed effect); B: effect estimates are log-odds; 95% CI: 95% confidence interval; Q: Cochran’s Q measure of heterogeneity; df: degree of freedom; WM: weighted median; DT: distortion test; GT: global test.

Legend:

P-value threshold <5e-8

The phenotypes are analyzed as binary cases (coded as 1) and controls (coded as 0). Do note that this leads to change in the direction of effect when comparing across some of the traits. HHI = cases: class 5 (i.e., HI>£100,000) vs controls: classes 1,2,3,4.

^a^ The Mendelian randomization pleiotropy residual sum and outlier (MR-PRESSO) test identifies possible bias from horizontal pleiotropy. The test consists of three parts, (1) the MR-PRESSO global test which detects horizontal pleiotropy, (2) the outlier corrected causal estimate which corrects for the detected horizontal pleiotropy and (3) the MR-PRESSO distortion test which estimates if the causal estimate is significantly different (at p<0.05) after adjustment for outliers. We conduct all three stages (with the argument NbDistribution=1000, namely using1000 simulation form the null distribution to compute empirical p-values) and present the outlier adjusted causal estimates (OACE) when both global and distortion tests are significant.

^b^ We did not run Steiger Test if none of the MR analysis resulted significant (NR: not reported in the cell).

^c^ Not enough SNP to perform MR (NR: not reported in the cell).

#### Supplementary Table 25: Odds Ratio of univariable bidirectional Mendelian randomization analysis of household income levels against mental illnesses

| *Forward MR analysis* | | | *Backward analysis* | | |
| --- | --- | --- | --- | --- | --- |
| **MR: method** | **OR (95% CI)** | **p-value** | **MR: method** | **OR (95% CI)** | **p-value** |
| LHI → ADHD:  IVW  WM  MR-Egger | 1.84 (1.51; 2.24)  1.77 (1.33; 2.35)  2.00 (0.750; 5.35) | <0.001  0.013  0.215 | ADHD → LHI:  IVW  WM  MR-Egger | 1.23 (1.19; 1.28)  1.23 (1.15; 1.31)  0.959 (0.703; 1.31) | <0.001  <0.001  0.792 |
| LMHI → ADHD:  IVW  WM  MR-Egger | 1.43 (1.22; 1.67)  1.18 (0.928; 1.49)  0.367 (0.030; 4.51) | <0.001  0.181  0.448 | ADHD → LMHI:  IVW  WM  MR-Egger | 1.19 (1.15; 1.23)  1.18 (1.11; 1.25)  0.985 (0.689; 1.41) | <0.001  <0.001  0.936 |
| MHHI → ADHD:  IVW  WM  MR-Egger | 0.742 (0.656; 0.841)  0.894 (0.732; 1.09)  0.820 (0.331; 2.03) | <0.001  0.272  0.674 | ADHD → MHHI:  IVW  WM  MR-Egger | 0.852 (0.820; 0.885)  0.856 (0.805; 0.911)  1.03 (0.756; 1.40) | <0.001  <0.001  0.867 |
| HHI → ADHD:  IVW  WM  MR-Egger | 0.967 (0.806; 1.16)  NR  NR | 0.720  NR  NR | ADHD → HHI:  IVW  WM  MR-Egger | 0.824 (0.767; 0.886)  0.810 (0.724; 0.905)  1.09 (0.629; 1.88) | <0.001  <0.001  0.765 |
| LHI → AN:  IVW  WM  MR-Egger | 0.912 (0.717; 1.16)  1.02 (0.716; 1.45)  0.910 (0.206; 4.03) | 0.452  0.919  0.904 | AN → LHI:  IVW  WM  MR-Egger | 1.03 (0.952; 1.10)  1.02 (0.925; 1.12)  0.595 (0.218; 1.62) | 0.500  0.725  0.417 |
| LMHI → AN:  IVW  WM  MR-Egger | 0.734 (0.591; 0.913)  0.801 (0.588; 1.09)  0.259 (0.015; 4.61) | 0.005  0.176  0.373 | AN → LMHI:  IVW  WM  MR-Egger | 1.03 (0.963; 1.09)  1.02 (0.944; 1.11)  0.521 (0.275; 0.986) | 0.426  0.564  0.183 |
| MHHI → AN:  IVW  WM  MR-Egger | 1.45 (1.21; 1.73)  1.32 (0.994; 1.77)  2.87 (0.800; 10.3) | <0.001  0.066  0.123 | AN → MHHI:  IVW  WM  MR-Egger | 0.991 (0.923; 1.06)  1.04 (0.946; 1.14)  1.37 (0.444; 4.22) | 0.803  0.425  0.639 |
| HHI → AN:  IVW  WM  MR-Egger | 1.39 (1.07; 1.82)  NR  NR | 0.015  NR  NR | AN → HHI:  IVW  WM  MR-Egger | 0.958 (0.837; 1.10)  0.965 (0.826; 1.13)  0.893 (0.258; 3.10) | 0.535  0.663  0.875 |
| LHI → ANX:  IVW  WM  MR-Egger | 1.87 (1.10; 3.20)  2.01 (1.01; 4.00)  1.35 (0.046; 39.8) | 0.021  0.045  0.866 | ANX → LHI:  IVW  WM  MR-Egger | NR  NR  NR | NR  NR  NR |
| LMHI → ANX:  IVW  WM  MR-Egger | 1.46 (0.925; 2.30)  1.21 (0.648; 2.25)  0.413 (0.008; 20.7) | 0.105  0.520  0.664 | ANX → LMHI:  IVW  WM  MR-Egger | NR  NR  NR | NR  NR  NR |
| MHHI → ANX:  IVW  WM  MR-Egger | 0.726 (0.499; 1.05)  0.907 (0.543; 1.52)  0.386 (0.038; 3.91) | 0.093  0.694  0.431 | ANX → MHHI:  IVW  WM  MR-Egger | NR  NR  NR | NR  NR  NR |
| HHI → ANX:  IVW  WM  MR-Egger | 0.904 (0.527; 1.55)  NR  NR | 0.715  NR  NR | ANX → HHI:  IVW  WM  MR-Egger | NR  NR  NR | NR  NR  NR |
| LHI → ASD:  IVW  WM  MR-Egger | 1.19 (0.948; 1.50)  1.26 (0.856; 1.85)  1.50 (0.162; 13.9) | 0.132  0.231  0.727 | ASD → LHI:  IVW  WM  MR-Egger | NR  NR  NR | NR  NR  NR |
| LMHI → ASD:  IVW  WM  MR-Egger | 0.765 (0.614; 0.954)  0.688 (0.463; 1.02)  0.117 (0.006; 2.11) | 0.017  0.053  0.165 | ASD → LMHI:  IVW  WM  MR-Egger | NR  NR  NR | NR  NR  NR |
| MHHI → ASD:  IVW  WM  MR-Egger | 1.62 (1.35; 1.93)  1.48 (1.14; 1.93)  1.41 (0.466; 4.28) | <0.001  0.010  0.549 | ASD → MHHI:  IVW  WM  MR-Egger | NR  NR  NR | NR  NR  NR |
| HHI → ASD:  IVW  WM  MR-Egger | 1.10 (0.886; 1.36)  1.66 (1.17; 2.36)  0.017 (0.002; 0.157) | 0.392  0.004  0.173 | ASD → HHI:  IVW  WM  MR-Egger | NR  NR  NR | NR  NR  NR |
| LHI → BD:  IVW  WM  MR-Egger | 1.36 (1.13; 1.63)  1.17 (0.877; 1.57)  0.580 (0.241; 1.39) | 0.001  0.243  0.263 | BD → LHI:  IVW  WM  MR-Egger | 1.00 (0.968; 1.03)  1.03 (0.975; 1.09)  1.13 (0.854; 1.50) | 0.983  0.275  0.394 |
| LMHI → BD:  IVW  WM  MR-Egger | 0.906 (0.778; 1.05)  0.935 (0.714; 1.22)  0.091 (0.005; 1.74) | 0.201  0.643  0.133 | BD → LMHI:  IVW  WM  MR-Egger | 0.970 (0.944; 0.997)  0.971 (0.927; 1.02)  1.00 (0.739; 1.35) | 0.032  0.208  0.999 |
| MHHI → BD:  IVW  WM  MR-Egger | 1.20 (1.05; 1.36)  1.03 (0.818; 1.29)  5.41 (1.46; 20.1) | 0.005  0.816  0.022 | BD → MHHI:  IVW  WM  MR-Egger | 1.04 (1.01; 1.07)  1.03 (0.983; 1.08)  1.02 (0.735; 1.43) | 0.011  0.239  0.885 |
| HHI → BD:  IVW  WM  MR-Egger | 1.84 (1.53; 2.21)  NR  NR | <0.001  NR  NR | BD → HHI:  IVW  WM  MR-Egger | 1.08 (1.02; 1.14)  1.07 (0.976; 1.17)  1.07 (0.642; 1.77) | 0.011  0.158  0.803 |
| LHI → MDD:  IVW  WM  MR-Egger | 1.42 (1.21; 1.67)  1.36 (1.07; 1.73)  2.41 (1.09; 5.35) | <0.001  0.016  0.006 | MDD → LHI:  IVW  WM  MR-Egger | NR  NR  NR | NR  NR  NR |
| LMHI → MDD:  IVW  WM  MR-Egger | 1.25 (1.08; 1.45)  1.23 (0.995; 1.51)  0.853 (0.250; 2.91) | 0.003  0.054  0.803 | MDD → LMHI:  IVW  WM  MR-Egger | NR  NR  NR | NR  NR  NR |
| MHHI → MDD:  IVW  WM  MR-Egger | 0.884 (0.788; 0.992)  0.932 (0.781; 1.11)  1.61 (0.837; 3.08) | 0.036  0.433  0.171 | MDD → MHHI:  IVW  WM  MR-Egger | NR  NR  NR | NR  NR  NR |
| HHI → MDD:  IVW  WM  MR-Egger | 1.01 (0.849; 1.21)  NR  NR | 0.889  NR  NR | MDD → HHI:  IVW  WM  MR-Egger | NR  NR  NR | NR  NR  NR |
| LHI → OCD:  IVW  WM  MR-Egger | 0.897 (0.478; 1.69)  1.34 (0.575; 3.13)  1.81 (0.075; 43.9) | 0.737  0.488  0.725 | OCD → LHI:  IVW  WM  MR-Egger | NR  NR  NR | NR  NR  NR |
| LMHI → OCD:  IVW  WM  MR-Egger | 0.461 (0.263; 0.809)  0.435 (0.201; 0.938)  0.705 (0.009; 57.5) | 0.007  0.027  0.878 | OCD → LMHI:  IVW  WM  MR-Egger | NR  NR  NR | NR  NR  NR |
| MHHI → OCD:  IVW  WM  MR-Egger | 0.924 (0.600; 1.42)  0.937 (0.497; 1.77)  0.671 (0.072; 6.23) | 0.720  0.836  0.729 | OCD → MHHI:  IVW  WM  MR-Egger | NR  NR  NR | NR  NR  NR |
| HHI → OCD:  IVW  WM  MR-Egger | 1.22 (0.626; 2.38)  NR  NR | 0.560  NR  NR | OCD → HHI:  IVW  WM  MR-Egger | NR  NR  NR | NR  NR  NR |
| LHI → PTSD:  IVW  WM  MR-Egger | 1.66 (1.29; 2.14)  1.67 (1.19; 2.34)  1.51 (0.463; 4.92) | <0.001  0.004  0.510 | PTSD → LHI:  IVW  WM  MR-Egger | NR  NR  NR | NR  NR  NR |
| LMHI → PTSD:  IVW  WM  MR-Egger | 1.33 (1.05; 1.68)  1.33 (0.934; 1.89)  0.616 (0.056; 6.79) | 0.020  0.117  0.537 | PTSD → LMHI:  IVW  WM  MR-Egger | NR  NR  NR | NR  NR  NR |
| MHHI → PTSD:  IVW  WM  MR-Egger | 0.793 (0.656; 0.957)  0.823 (0.619; 1.09)  0.923 (0.346; 2.46) | 0.016  0.165  0.874 | PTSD → MHHI:  IVW  WM  MR-Egger | NR  NR  NR | NR  NR  NR |
| HHI → PTSD:  IVW  WM  MR-Egger | 0.827 (0.654; 1.05)  0.855 (0.642; 1.14)  0.412 (0.091; 1.87) | 0.114  0.300  0.456 | PTSD → HHI:  IVW  WM  MR-Egger | NR  NR  NR | NR  NR  NR |
| LHI → SZ:  IVW  WM  MR-Egger | 1.91 (1.63; 2.24)  1.49 (1.11; 2.00)  0.860 (0.115; 6.44) | <0.001  0.009  0.887 | SZ → LHI:  IVW  WM  MR-Egger | 1.09 (1.07; 1.10)  1.07 (1.05; 1.10)  1.12 (1.02; 1.24) | <0.001  <0.001  0.024 |
| LMHI → SZ:  IVW  WM  MR-Egger | 1.31 (1.15; 1.49)  1.12 (0.877; 1.44)  0.761 (0.033; 17.6) | <0.001  0.357  0.867 | SZ → LMHI:  IVW  WM  MR-Egger | 1.05 (1.03; 1.06)  1.04 (1.02; 1.07)  1.03 (0.949; 1.12) | <0.001  <0.001  0.455 |
| MHHI → SZ:  IVW  WM  MR-Egger | 0.907 (0.818; 1.01)  0.843 (0.687; 1.03)  0.790 (0.202; 3.08) | 0.063  0.100  0.738 | SZ → MHHI:  IVW  WM  MR-Egger | 0.955 (0.941; 0.970)  0.962 (0.936; 0.989)  1.02 (0.927; 1.12) | <0.001  0.005  0.707 |
| HHI → SZ:  IVW  WM  MR-Egger | 1.03 (0.886; 1.20)  NR  NR | 0.693  NR  NR | SZ → HHI:  IVW  WM  MR-Egger | 0.987 (0.959; 1.02)  1.00 (0.956; 1.05)  1.10 (0.950; 1.27) | 0.395  0.950  0.205 |

Abbreviations: MR: Mendelian randomization; OR: Odds Ratio; 95% CI: 95% confidence intervals; HI: household income; ADHD: attention deficit hyperactivity disorder; AN: anorexia nervosa; ANX: anxiety disorder; ASD: autism spectrum disorders; BD: bipolar disorder; MDD: major depressive disorder; OCD: obsessive-compulsive disorder; PTSD: post-traumatic stress disorder; SZ: schizophrenia; IVW: inverse variance weighted (fixed effect); WM: weighted median; NR: not reported because not enough SNP to perform MR.

Legend: LHI: low household income, cases were those less than £18,000; LMHI: low-mid HI, cases were those less than £29,999; MHHI: mid-high HI, cases were those more than £52,000; HHI: high HI, cases were those more than £100,000.

#### Supplementary Table 26: Results of univariable bidirectional Mendelian randomization analysis of household income levels against mental illnesses, after Steiger filtering

| **MR** | **N SNP** | **IVW, B (95% CI)** | **p-value** | **WM, B (95% CI)** | **p-value** | **MR-Egger, B (95% CI)** | **p-value** | **Egger intercept p-value** | **Mean F** |
| --- | --- | --- | --- | --- | --- | --- | --- | --- | --- |
| LHI on ADHD | 8 | 0.610 (0.415; 0.805) | <0.001 | 0.569 (0.278; 0.861) | <0.001 | 0.695 (-0.288; 1.68) | 0.215 | 0.744 | 33.5 |
| LMHI on ADHD | 14 | 0.238 (0.075; 0.401) | 0.004 | 0.156 (-0.088; 0.399) | 0.209 | -1.62 (-3.18; 0.057) | 0.065 | 0.170 | 36.0 |
| MHHI on ADHD | 19 | -0.246 (-0.373; -0.118) | <0.001 | -0.111 (-0.311; 0.090) | 0.279 | 0.375 (-0.476; 1.23) | 0.400 | 0.428 | 36.5 |
| HHI on ADHD | 2 | -0.033 (-0.216; 0.149) | 0.720 | NR | NR | NR | NR | NR | 45.1 |
| LHI on AN | 10 | 0.005 (-0.247; 0.256) | 0.972 | 0.028 (-0.310; 0.365) | 0.871 | 0.217 (-0.970; 1.40) | 0.729 | 0.807 | 34.0 |
| LMHI on AN | 15 | -0.096 (-0.330; 0.138) | 0.421 | -0.078 (-0.401; 0.244) | 0.634 | -1.34 (-3.15; 0.468) | 0.170 | 0.130 | 34.9 |
| MHHI on AN | 17 | 0.137 (-0.062; 0.336) | 0.178 | 0.212 (-0.072; 0.496) | 0.143 | 0.429 (-0.624; 1.48) | 0.437 | 0.921 | 34.8 |
| HHI on AN | 2 | 0.333 (0.065; 0.600) | 0.015 | NR | NR | NR | NR | NR | 45.1 |
| LHI on ANX | 9 | 0.509 (-0.061; 1.08) | 0.080 | 0.677 (-0.070; 1.42) | 0.076 | 0.437 (-2.96; 3.83) | 0.808 | 0.377 | 33.5 |
| LMHI on ANX | 16 | 0.285 (-0.184; 0.754) | 0.233 | 0.174 (-0.421; 0.769) | 0.566 | -0.608 (-4.54; 3.32) | 0.766 | 0.022 | 35.5 |
| MHHI on ANX | 19 | -0.286 (-0.662; 0.089) | 0.135 | -0.096 (-0.604; 0.411) | 0.710 | -0.400 (-2.79; 1.99) | 0.747 | 0.097 | 36.8 |
| HHI on ANX | 2 | -0.101 (-0.641; 0.440) | 0.715 | NR | NR | NR | NR | NR | 45.1 |
| LHI on ASD | 8 | 0.213 (-0.066; 0.492) | 0.135 | 0.274 (-0.127; 0.675) | 0.180 | -0.739 (-1.90; 0.423) | 0.259 | 0.459 | 33.8 |
| LMHI on ASD | 9 | 0.003 (-0.311; 0.318) | 0.980 | -0.110 (-0.522; 0.302) | 0.602 | -1.10 (-3.49; 1.29) | 0.396 | 0.287 | 32.7 |
| MHHI on ASD | 16 | 0.240 (0.034; 0.445) | 0.023 | 0.136 (-0.160; 0.430) | 0.367 | 0.230 (-0.618; 1.08) | 0.603 | 0.004 | 35.6 |
| HHI on ASD | 0 | NR | NR | NR | NR | NR | NR | NR | NR |
| LHI on BD | 6 | 0.073 (-0.154; 0.300) | 0.528 | 0.068 (-0.210; 0.345) | 0.633 | -0.146 (-0.995; 0.702) | 0.752 | 0.554 | 32.5 |
| LMHI on BD | 11 | -0.001 (-0.190; 0.190) | 0.998 | -0.004 (-0.286; 0.279) | 0.980 | 0.876 (-1.17; 2.92) | 0.424 | 0.988 | 32.5 |
| MHHI on BD | 14 | -0.042 (-0.197; 0.114) | 0.598 | -0.140 (-0.375; 0.094) | 0.241 | 0.211 (-0.746; 1.17) | 0.673 | 0.247 | 33.0 |
| HHI on BD | 0 | NR | NR | NR | NR | NR | NR | NR | NR |
| LHI on MDD | 10 | 0.351 (0.189; 0.513) | <0.001 | 0.308 (0.072; 0.544) | 0.011 | 0.880 (0.082; 1.68) | 0.063 | 0.942 | 33.5 |
| LMHI on MDD | 16 | 0.224 (0.077; 0.371) | 0.003 | 0.204 (-0.005; 0.412) | 0.056 | -0.026 (-1.35; 1.30) | 0.970 | 0.650 | 35.6 |
| MHHI on MDD | 21 | -0.123 (-0.238; -0.008) | 0.036 | -0.070 (-0.240; 0.100) | 0.420 | 0.474 (-0.178; 1.13) | 0.171 | 0.526 | 36.2 |
| HHI on MDD | 2 | 0.013 (-0.163; 0.188) | 0.889 | NR | NR | NR | NR | NR | 45.1 |
| LHI on OCD | 8 | 0.119 (-0.592; 0.830) | 0.743 | 0.272 (-0.590; 1.13) | 0.536 | -0.417 (-3.55; 2.72) | 0.803 | 0.064 | 33.5 |
| LMHI on OCD | 9 | -0.036 (-0.793; 0.722) | 0.927 | 0.095 (-0.863; 1.05) | 0.846 | 1.60 (-3.87; 7.07) | 0.584 | 0.781 | 35.9 |
| MHHI on OCD | 12 | 0.072 (-0.478; 0.621) | 0.798 | 0.231 (-0.514; 0.976) | 0.544 | 0.753 (-1.82; 3.33) | 0.579 | 0.726 | 39.0 |
| HHI on OCD | 0 | NR | NR | NR | NR | NR | NR | NR | NR |
| LHI on PTSD | 12 | 0.506 (0.253; 0.759) | <0.001 | 0.511 (0.167; 0.854) | 0.004 | 0.412 (-0.769; 1.59) | 0.510 | 0.949 | 33.5 |
| LMHI on PTSD | 17 | 0.283 (0.045; 0.521) | 0.002 | 0.285 (-0.064; 0.633) | 0.109 | -0.485 (-2.89; 1.92) | 0.698 | 0.508 | 35.3 |
| MHHI on PTSD | 21 | -0.233 (-0.421; -0.044) | 0.016 | -0.195 (-0.475; 0.084) | 0.171 | -0.080 (-1.06; 0.900) | 0.874 | 0.506 | 36.2 |
| HHI on PTSD | 3 | -0.189 (-0.425; 0.046) | 0.114 | -0.157 (-0.452; 0.139) | 0.298 | -0.887 (-2.40; 0.626) | 0.456 | 0.695 | 46.8 |
| LHI on SZ | 8 | 0.491 (0.320; 0.662) | <0.001 | 0.382 (0.088; 0.675) | 0.011 | 0.300 (-1.45; 2.05) | 0.749 | 0.447 | 33.5 |
| LMHI on SZ | 15 | 0.145 (0.009; 0.282) | 0.037 | 0.110 (-0.141; 0.360) | 0.391 | -0.212 (-2.84; 2.42) | 0.877 | 0.325 | 35.2 |
| MHHI on SZ | 19 | -0.023 (-0.129; 0.083) | 0.665 | -0.170 (-0.380; 0.040) | 0.112 | -0.331 (-1.53; 0.868) | 0.596 | 0.454 | 36.3 |
| HHI on SZ | 2 | 0.030 (-0.121; 0.182) | 0.693 | NR | NR | NR | NR | NR | 45.1 |

Abbreviations: MR: Mendelian randomization; B: effect estimates are log-odds; 95% CI: 95% confidence intervals; HI: household income; ADHD: attention deficit hyperactivity disorder; AN: anorexia nervosa; ANX: anxiety disorder; ASD: autism spectrum disorders; BD: bipolar disorder; MDD: major depressive disorder; OCD: obsessive-compulsive disorder; PTSD: post-traumatic stress disorder; SZ: schizophrenia; IVW: inverse variance weighted (fixed effect); WM: weighted median; NR: not reported because not enough SNP to perform MR.

Legend: LHI: low household income, cases were those less than £18,000; LMHI: low-mid HI, cases were those less than £29,999; MHHI: mid-high HI, cases were those more than £52,000; HHI: high HI, cases were those more than £100,000.

### **Univariable Mendelian randomization of cognitive abilities and mental illnesses**

#### Supplementary Table 27: Odds Ratio of univariable forward Mendelian randomization analysis of cognitive abilities against mental illnesses

| **MR: method** | **OR (95% CI)** | **p-value** |
| --- | --- | --- |
| CA → ADHD:  IVW  WM  MR-Egger | 0.528 (0.487; 0.573)  0.611 (0.532; 0.702)  0.530 (0.282; 0.995) | <0.001  <0.001  0.050 |
| CA→ AN:  IVW  WM  MR-Egger | 1.36 (1.21; 1.53)  1.47 (1.22; 1.76)  2.65 (1.25; 5.62) | <0.001  <0.001  0.012 |
| CA → ANX:  IVW  WM  MR-Egger | 0.709 (0.561; 0.896)  0.853 (0.604; 1.20)  0.765 (0.246; 2.37) | 0.004  0.358  0.643 |
| CA → ASD:  IVW  WM  MR-Egger | 1.36 (1.21; 1.53)  1.32 (1.10; 1.60)  1.74 (0.760; 3.97) | <0.001  0.004  0.193 |
| CA → BD:  IVW  WM  MR-Egger | 1.00 (0.925; 1.09)  0.887 (0.767; 1.02)  1.16 (0.564; 2.38) | 0.934  0.102  0.689 |
| CA → MDD:  IVW  WM  MR-Egger | 0.870 (0.807; 0.937)  0.902 (0.799; 1.02)  0.838 (0.520; 1.35) | <0.001  0.099  0.470 |
| CA → OCD:  IVW  WM  MR-Egger | 1.31 (0.988; 1.75)  1.20 (0.775; 1.87)  0.903 (0.198; 4.12) | 0.061  0.414  0.896 |
| CA → PTSD:  IVW  WM  MR-Egger | 0.870 (0.768; 0.987)  0.813 (0.671; 0.984)  1.25 (0.633; 2.46) | 0.030  0.032  0.525 |
| CA → SZ:  IVW  WM  MR-Egger | 0.743 (0.694; 0.796)  0.862 (0.748; 0.993)  0.945 (0.393; 2.27) | <0.001  0.038  0.899 |

Abbreviations: MR: Mendelian randomization; OR: Odds Ratio; 95% CI: 95% confidence intervals; CA: cognitive abilities; ADHD: attention deficit hyperactivity disorder; AN: anorexia nervosa; ANX: anxiety disorder; ASD: autism spectrum disorders; BD: bipolar disorder; MDD: major depressive disorder; OCD: obsessive-compulsive disorder; PTSD: post-traumatic stress disorder; SZ: schizophrenia; IVW: inverse variance weighted (fixed effect); WM: weighted median; NR: not reported because not enough SNP to perform MR.

#### Supplementary Table 28: results of bidirectional MR of cognitive abilities against mental illness

| **MR** | **N SNP** | **IVW, B (95% CI)** | **p-value** | **IVW Q(df)** | **p-value** | **WM, B (95% CI)** | **p-value** | **MR-Egger, B (95% CI)** | **p-value** | **Egger intercept p-value** | **Steiger Test p-value** | **MR-PRESSO** | **Mean F** |
| --- | --- | --- | --- | --- | --- | --- | --- | --- | --- | --- | --- | --- | --- |
| Fw: CA on ADHD | 131 | -0.638 (-0.720; -0.556) | <0.001 | 341 (130) | <0.001 | -0.493 (-0.632; -0.354) | <0.001 | -0.635 (-1.27; -0.005) | 0.050 | 0.993 | <0.001 | DT; p=0.714 | 44.0 |
| Bw: ADHD on CA | 23 | -0.151 (-0.172; -0.131) | <0.001 | 91 (22) | <0.001 | -0.106 (-0.142; -0.071) | <0.001 | -0.010 (-0.236; 0.256) | 0.938 | 0.206 | <0.001 | OACE: -0.122 (-0.155; -0.090); p<0.001 | 39.2 |
| Fw: CA on AN | 137 | 0.306 (0.190; 0.422) | <0.001 | 291 (136) | <0.001 | 0.384 (0.197; 0.571) | <0.001 | 0.974 (0.223; 1.73) | 0.012 | 0.076 | <0.001 | DT; p=0.874 | 43.9 |
| Bw: AN on CA | 3 | -0.005 (-0.047; 0.038) | 0.826 | 6 (2) | 0.053 | -0.008 (-0.072; 0.055) | 0.795 | -0.007 (-0.931; 0.917) | 0.991 | 0.997 | NR ^b^ | NR ^c^ | 32.3 |
| Fw: CA on ANX | 137 | -0.344 (-0.578; -0.110) | 0.004 | 149 (136) | 0.204 | -0.159 (-0.499; 0.180) | 0.358 | -0.268 (-1.40; 0.865) | 0.643 | 0.894 | <0.001 | GT; p=0.210 | 43.9 |
| Bw: ANX on CA | 0 | NR ^c^ | NR ^c^ | NR ^c^ | NR ^c^ | NR ^c^ | NR ^c^ | NR ^c^ | NR ^c^ | NR ^c^ | NR ^c^ | NR ^c^ | NR ^c^ |
| Fw: CA on ASD | 138 | 0.310 (0.193; 0.427) | <0.001 | 338 (137) | <0.001 | 0.280 (0.087; 0.473) | 0.004 | 0.552 (-0.274; 1.38) | 0.193 | 0.556 | <0.001 | DT; p=0.553 | 43.8 |
| Bw: ASD on CA | 0 | NR ^c^ | NR ^c^ | NR ^c^ | NR ^c^ | NR ^c^ | NR ^c^ | NR ^c^ | NR ^c^ | NR ^c^ | NR ^c^ | NR ^c^ | NR ^c^ |
| Fw: CA on BD | 134 | 0.003 (-0.078; 0.085) | 0.934 | 491 (133) | <0.001 | -0.120 (-0.265; 0.024) | 0.102 | 0.147 (-0.573; 0.867) | 0.689 | 0.689 | NR ^b^ | DT; p=0.420 | 44.0 |
| Bw: BD on CA | 36 | 0.015 (-0.001; 0.032) | 0.069 | 370 (35) | <0.001 | 0.024 (-0.006; 0.054) | 0.119 | -0.149 (-0.445; 0.147) | 0.330 | 0.276 | NR ^b^ | OACE: -0.003 (-0.030; 0.024); p=0.805 | 39.2 |
| Fw: CA on MDD | 138 | -0.140 (-0.215; -0.065) | <0.001 | 279 (137) | <0.001 | -0.103 (-0.225; 0.019) | 0.099 | -0.177 (-0.654; 0.301) | 0.470 | 0.876 | <0.001 | DT; p=0.842 | 43.8 |
| Bw: MDD on CA | 0 | NR ^c^ | NR ^c^ | NR ^c^ | NR ^c^ | NR ^c^ | NR ^c^ | NR ^c^ | NR ^c^ | NR ^c^ | NR ^c^ | NR ^c^ | NR ^c^ |
| Fw: CA on OCD | 138 | 0.272 (-0.012; 0.557) | 0.061 | 192 (137) | 0.001 | 0.186 (-0.259; 0.630) | 0.414 | -0.102 (-1.62; 1.41) | 0.896 | 0.621 | NR ^b^ | DT; p=0.848 | 43.8 |
| Bw: OCD on CA | 0 | NR ^c^ | NR ^c^ | NR ^c^ | NR ^c^ | NR ^c^ | NR ^c^ | NR ^c^ | NR ^c^ | NR ^c^ | NR ^c^ | NR ^c^ | NR ^c^ |
| Fw: CA on PTSD | 136 | -0.139 (-0.264; -0.013) | 0.030 | 192 (135) | <0.001 | -0.207 (-0.396; -0.018) | 0.032 | 0.220 (-0.458; 0.898) | 0.525 | 0.289 | <0.001 | DT; p=0.172 | 43.9 |
| Bw: PTSD on CA | 0 | NR ^c^ | NR ^c^ | NR ^c^ | NR ^c^ | NR ^c^ | NR ^c^ | NR ^c^ | NR ^c^ | NR ^c^ | NR ^c^ | NR ^c^ | NR ^c^ |
| Fw: CA on SZ | 134 | -0.297 (-0.365; -0.228) | <0.001 | 986 (133) | <0.001 | -0.149 (-0.289; -0.008) | 0.038 | -0.057 (-0.933; 0.819) | 0.899 | 0.584 | <0.001 | DT; p=0.863 | 44.0 |
| Bw: SZ on CA | 175 | -0.055 (-0.063; -0.047) | <0.001 | 1010 (174) | <0.001 | -0.046 (-0.062; -0.030) | <0.001 | -0.092 (-0.170; 0.014) | 0.023 | 0.345 | <0.001 | OACE: -0.036 (-0.051; -0.022); p<0.001 | 45.6 |

Abbreviations: Fw: forward analysis; Bw: backward analysis; CA: cognitive abilities; ADHD: attention deficit hyperactivity disorder; AN: anorexia nervosa; ANX: anxiety disorder; ASD: autism spectrum disorder; BD: bipolar disorder; MDD: major depressive disorder; OCD: obsessive-compulsive disorder; PTSD: post-traumatic stress disorder; SZ: schizophrenia; MR: mendelian randomization; SNP: single nucleotide polymorphism; IVW: inverse variance weighted (fixed effect); B: effect estimates are log-odds for binary traits (i.e., for mental illnesses) and unstandardized regression coefficient for continuous traits (i.e., for cognitive abilities); 95% CI: 95% confidence interval; Q: Cochran’s Q measure of heterogeneity; df: degree of freedom; WM: weighted median; DT: distortion test; GT: global test.

Legend:

P-value threshold <5e-8

^a^ The Mendelian randomization pleiotropy residual sum and outlier (MR-PRESSO) test identifies possible bias from horizontal pleiotropy. The test consists of three parts, (1) the MR-PRESSO global test which detects horizontal pleiotropy, (2) the outlier corrected causal estimate which corrects for the detected horizontal pleiotropy and (3) the MR-PRESSO distortion test which estimates if the causal estimate is significantly different (at p<0.05) after adjustment for outliers. We conduct all three stages (with the argument NbDistribution=1000, namely using1000 simulation form the null distribution to compute empirical p-values) and present the outlier adjusted causal estimates (OACE) when both global and distortion tests are significant.

^b^ We did not run Steiger Test if none of the MR analysis resulted significant (NR: not reported in the cell).

^c^ Not enough SNP to perform MR (NR: not reported in the cell).

#### Plots - Forward analyses

##### Supplementary Figure 108: scatterplot of cognitive abilities against ADHD

Abbreviations: MR: Mendelian randomization; SNP: single nucleotide polymorphism; CA: cognitive abilities; ADHD: attention deficit hyperactivity disorder.

##### Supplementary Figure 109: scatterplot of cognitive abilities against AN

Abbreviations: MR: Mendelian randomization; SNP: single nucleotide polymorphism; CA: cognitive abilities; AN: anorexia nervosa.

##### Supplementary Figure 110: scatterplot of cognitive abilities against ANX

Abbreviations: MR: Mendelian randomization; SNP: single nucleotide polymorphism; CA: cognitive abilities; ANX: anxiety disorders.

##### Supplementary Figure 111: scatterplot of cognitive abilities against ASD

Abbreviations: MR: Mendelian randomization; SNP: single nucleotide polymorphism; CA: cognitive abilities; ASD: autism spectrum disorders.

##### Supplementary Figure 112: scatterplot of cognitive abilities against BD

Abbreviations: MR: Mendelian randomization; SNP: single nucleotide polymorphism; CA: cognitive abilities; BD: bipolar disorder.

##### Supplementary Figure 113: scatterplot of cognitive abilities against MDD

Abbreviations: MR: Mendelian randomization; SNP: single nucleotide polymorphism; CA: cognitive abilities; MDD: major depressive disorder.

##### Supplementary Figure 114: scatterplot of cognitive abilities against OCD

Abbreviations: MR: Mendelian randomization; SNP: single nucleotide polymorphism; CA: cognitive abilities; OCD: obsessive-compulsive disorder.

##### Supplementary Figure 115: scatterplot of cognitive abilities against PTSD

Abbreviations: MR: Mendelian randomization; SNP: single nucleotide polymorphism; CA: cognitive abilities; PTSD: post-traumatic stress disorder.

##### Supplementary Figure 116: scatterplot of cognitive abilities against SZ

Abbreviations: MR: Mendelian randomization; SNP: single nucleotide polymorphism; CA: cognitive abilities; SZ: schizophrenia.

##### Supplementary Figure 117: leave-one out analysis of cognitive abilities against ADHD

Abbreviations: MR: Mendelian randomization; CA: cognitive abilities; ADHD: attention deficit hyperactivity disorder.

##### Supplementary Figure 118: leave-one out analysis of cognitive abilities against AN

Abbreviations: MR: Mendelian randomization; CA: cognitive abilities; AN: anorexia nervosa.

##### Supplementary Figure 119: leave-one out analysis of cognitive abilities against ANX

Abbreviations: MR: Mendelian randomization; CA: cognitive abilities; ANX: anxiety disorders.

##### Supplementary Figure 120: leave-one out analysis of cognitive abilities against ASD

Abbreviations: MR: Mendelian randomization; CA: cognitive abilities; ASD: autism spectrum disorders.

##### Supplementary Figure 121: leave-one out analysis of cognitive abilities against BD

Abbreviations: MR: Mendelian randomization; CA: cognitive abilities; BD: bipolar disorder.

##### Supplementary Figure 122: leave-one-out analysis of cognitive abilities against MDD

Abbreviations: MR: Mendelian randomization; CA: cognitive abilities; MDD: major depressive disorder.

##### Supplementary Figure 123: leave-one out analysis of cognitive abilities against OCD

Abbreviations: MR: Mendelian randomization; CA: cognitive abilities; OCD: obsessive-compulsive disorder.

##### Supplementary Figure 124: leave-one out analysis of cognitive abilities against PTSD

Abbreviations: MR: Mendelian randomization; CA: cognitive abilities; PTSD: post-traumatic stress disorder.

##### Supplementary Figure 125: leave-one-out analysis of cognitive abilities against SZ

Abbreviations: MR: Mendelian randomization; CA: cognitive abilities; SZ: schizophrenia.

#### Plots - Backward analyses

##### Supplementary Figure 126: scatterplot of ADHD against cognitive abilities

Abbreviations: MR: Mendelian randomization; SNP: single nucleotide polymorphism; CA: cognitive abilities; ADHD: attention deficit hyperactivity disorder.

##### Supplementary Figure 127: scatterplot of AN against cognitive abilities

Abbreviations: MR: Mendelian randomization; SNP: single nucleotide polymorphism; CA: cognitive abilities; AN: anorexia nervosa.

##### Supplementary Figure 128: scatterplot of BD against cognitive abilities

Abbreviations: MR: Mendelian randomization; SNP: single nucleotide polymorphism; CA: cognitive abilities; BD: bipolar disorder.

##### Supplementary Figure 129: scatterplot of SZ against cognitive abilities

Abbreviations: MR: Mendelian randomization; SNP: single nucleotide polymorphism; CA: cognitive abilities; SZ: schizophrenia.

##### Supplementary Figure 130: leave-one-out analysis of ADHD against cognitive abilities

Abbreviations: MR: Mendelian randomization; CA: cognitive abilities; ADHD: attention deficit hyperactivity disorder.

##### Supplementary Figure 131: leave-one-out analysis of AN against cognitive abilities

Abbreviations: MR: Mendelian randomization; CA: cognitive abilities; AN: anorexia nervosa.

##### Supplementary Figure 132: leave-one-out analysis of BD against cognitive abilities

Abbreviations: MR: Mendelian randomization; CA: cognitive abilities; BD: bipolar disorder.

##### Supplementary Figure 133: leave-one-out analysis of SZ against cognitive abilities

Abbreviations: MR: Mendelian randomization; CA: cognitive abilities; SZ: schizophrenia.

#### Supplementary Table 29: CAUSE results of the relations between cognitive abilities and mental illnesses

| **Model 1** | **Model 2** | **∆ ELPD** | **SE ∆ ELPD** | **z-score** | **p-value** |
| --- | --- | --- | --- | --- | --- |
| *Fw: CA on ADHD* | | | | | |
| Null  Null  Sharing | Sharing  Causal  Causal | -69.24  -76.54  -7.30 | 9.79  10.84  1.21 | -7.07  -7.06  -6.04 | <0.001  <0.001  <0.001 |
| *Bw: ADHD on CA* | | | | | |
| Null  Null  Sharing | Sharing  Causal  Causal | -22.00  -28.89  -6.89 | 4.75  6.27  1.57 | -4.64  -4.61  -4.39 | <0.001  <0.001  <0.001 |
| *Fw: CA on AN* | | | | | |
| Null  Null  Sharing | Sharing  Causal  Causal | -0.01  -1.01  -1.00 | 0.53  2.02  1.50 | -0.02  -0.50  -0.66 | 0.984  0.617  0.509 |
| *Bw: AN on CA* | | | | | |
| Null  Null  Sharing | Sharing  Causal  Causal | 0.46  1.35  0.89 | 0.07  0.07  0.02 | 6.83  18.32  40.41 | <0.001  <0.001  <0.001 |
| *Fw: CA on ANX* | | | | | |
| Null  Null  Sharing | Sharing  Causal  Causal | -7.02  -7.17  -0.14 | 3.68  4.27  1.38 | -1.91  -1.68  -0.11 | 0.056  0.093  0.912 |
| *Bw: ANX on CA* | | | | | |
| Null  Null  Sharing | Sharing  Causal  Causal | 0.25  1.08  0.83 | 0.09  0.30  0.25 | 2.88  3.61  3.35 | 0.004  <0.001  0.001 |
| *Fw: CA on ASD* | | | | | |
| Null  Null  Sharing | Sharing  Causal  Causal | -0.40  -1.88  -1.48 | 0.88  2.43  1.57 | -0.46  -0.78  -0.94 | 0.646  0.435  0.347 |
| *Bw: ASD on CA* | | | | | |
| Null  Null  Sharing | Sharing  Causal  Causal | 0.31  0.39  0.08 | 0.17  1.01  0.85 | 1.88  0.39  0.09 | 0.060  0.697  0.928 |
| *Fw: CA on BD* | | | | | |
| Null  Null  Sharing | Sharing  Causal  Causal | 0.42  0.79  0.37 | 0.15  0.98  0.84 | 2.80  0.81  0.84 | 0.003  0.210  0.328 |
| *Bw: BD on CA* | | | | | |
| Null  Null  Sharing | Sharing  Causal  Causal | 0.45  1.09  0.64 | 0.07  0.52  0.45 | 5.98  2.08  1.42 | <0.001  0.038  0.156 |
| *Fw: CA on MDD* | | | | | |
| Null  Null  Sharing | Sharing  Causal  Causal | -7.83  -12.20  -4.38 | 2.96  4.61  1.68 | -2.64  -2.65  -2.60 | 0.008  0.008  0.009 |
| *Bw: MDD on CA* | | | | | |
| Null  Null  Sharing | Sharing  Causal  Causal | 0.36  0.78  0.42 | 0.08  0.73  0.66 | 4.74  1.07  0.64 | <0.001  0.285  0.522 |
| *Fw: CA on OCD* | | | | | |
| Null  Null  Sharing | Sharing  Causal  Causal | -1.11  -1.66  -0.54 | 1.55  2.54  1.10 | -0.72  -0.65  -0.50 | 0.471  0.516  0.617 |
| *Bw: OCD on CA* | | | | | |
| Null  Null  Sharing | Sharing  Causal  Causal | 0.28  1.05  0.77 | 0.03  0.11  0.10 | 8.40  9.63  8.08 | <0.001  <0.001  <0.001 |
| *Fw: CA on PTSD* | | | | | |
| Null  Null  Sharing | Sharing  Causal  Causal | -10.48  -13.58  -3.10 | 3.93  5.18  1.48 | -2.67  -2.62  -2.09 | 0.008  0.009  0.037 |
| *Bw: PTSD on CA* | | | | | |
| Null  Null  Sharing | Sharing  Causal  Causal | 0.27  0.77  0.50 | 0.09  0.63  0.55 | 2.86  1.23  0.92 | 0.004  0.219  0.358 |
| *Fw: CA on SZ* | | | | | |
| Null  Null  Sharing | Sharing  Causal  Causal | -6.54  -12.02  -5.48 | 2.36  4.25  1.90 | -2.77  -2.83  -2.88 | 0.006  0.005  0.004 |
| *Bw: SZ on CA* | | | | | |
| Null  Null  Sharing | Sharing  Causal  Causal | -5.95  -10.84  -4.89 | 2.35  4.20  1.86 | -2.53  -2.58  -2.62 | 0.011  0.010  0.009 |

Abbreviations: MR: mendelian randomization; ELPD: expected log pointwise posterior density; 95%CI: 95% confidence interval; SE: standard error; Fwd: forward MR; Bwd: backward MR; CA: cognitive abilities; ADHD: attention deficit hyperactivity disorder; AN: anorexia nervosa; ANX: anxiety disorder; ASD: autism spectrum disorder; BD: bipolar disorder; MDD: major depressive disorder; OCD: obsessive-compulsive disorder; PTSD: post-traumatic stress disorder; SZ: schizophrenia.

#### Supplementary Table 30: Results of univariable bidirectional Mendelian Randomization of cognitive abilities against mental illnesses, after Steiger filtering

| **MR** | **N SNP** | **IVW, B (95% CI)** | **p-value** | **WM, B (95% CI)** | **p-value** | **MR-Egger, B (95% CI)** | **p-value** | **Egger intercept p-value** | **Mean F** |
| --- | --- | --- | --- | --- | --- | --- | --- | --- | --- |
| CA on ADHD | 131 | -0.638 (-0.720; -0.556) | <0.001 | -0.493 (-0.632; -0.354) | <0.001 | -0.635 (-1.27; -0.005) | 0.050 | 0.993 | 44.0 |
| CA on AN | 132 | 0.298 (0.180; 0.416) | <0.001 | 0.366 (0.170; 0.561) | <0.001 | 0.854 (0.183; 1.52) | 0.014 | 0.147 | 44.0 |
| CA on ANX | 125 | -0.109 (-0.353; 0.135) | 0.380 | -0.068 (-0.425; 0.288) | 0.706 | 0.143 (-0.967; 1.25) | 0.801 | 0.138 | 44.1 |
| CA on ASD | 116 | 0.102 (-0.026; 0.229) | 0.117 | 0.172 (-0.021; 0.366) | 0.081 | 0.557 (-0.096; 1.21) | 0.097 | 0.076 | 44.3 |
| CA on BD | 112 | -0.083 (-0.173; 0.006) | 0.069 | -0.138 (-0.283; 0.006) | 0.061 | -0.082 (-0.623; 0.460) | 0.768 | 0.632 | 42.9 |
| CA on MDD | 137 | -0.153 (-0.228; -0.078) | <0.001 | -0.106 (-0.228; 0.016) | 0.089 | -0.098 (-0.557; 0.362) | 0.677 | 0.282 | 43.9 |
| CA on OCD | 94 | -0.013 (-0.354; 0.327) | 0.940 | -0.097 (-0.583; 0.389) | 0.695 | -0.125 (-1.57; 1.32) | 0.866 | 0.855 | 44.8 |
| CA on PTSD | 136 | -0.139 (-0.264; -0.013) | 0.030 | -0.207 (-0.406; -0.009) | 0.041 | 0.220 (-0.458; 0.898) | 0.525 | 0.498 | 43.9 |
| CA on SZ | 130 | -0.209 (-0.279; -0.139) | <0.001 | -0.140 (-0.276; -0.005) | 0.043 | 0.161 (-0.624; 0.945) | 0.689 | 0.001 | 44 |

Abbreviations: CA: cognitive abilities; ADHD: attention deficit hyperactivity disorder; AN: anorexia nervosa; ANX: anxiety disorder; ASD: autism spectrum disorders; BD: bipolar disorder; MDD: major depressive disorder; OCD: obsessive-compulsive disorder; PTSD: post-traumatic stress disorder; SZ: schizophrenia; MR: mendelian randomization; SNP: single nucleotide polymorphism; IVW: inverse variance weighted (fixed effect); B: effect estimates are log-odds; 95% CI: 95% confidence interval; WM: weighted median; NR: not reported because not enough SNP to perform MR.

### **Multivariable Mendelian Randomization of poverty indicators and cognitive abilities against mental illness**

#### Supplementary Table 31: Multivariable Mendelian Randomization results of household income and cognitive abilities on mental illness

| **Regression** | **N SNP** | **IVW, B (95% CI)** | **p-value** |
| --- | --- | --- | --- |
| *Outcome: ADHD*  Exposure 1: HI  Exposure 2: CA | 192 | -0.124 (-0.326; 0.078)  -0.508 (-0.639; -0.376) | 0.230  <0.001 |
| *Outcome: AN*  Exposure 1: HI  Exposure 2: CA | 201 | 0.010 (-0.229; 0.250)  0.350 (0.202; 0.498) | 0.933  <0.001 |
| *Outcome: ANX*  Exposure 1: HI  Exposure 2: CA | 199 | 0.010 (-0.350; 0.370)  -0.260 (-0.475; -0.045) | 0.956  0.018 |
| *Outcome: ASD*  Exposure 1: HI  Exposure 2: CA | 204 | 0.196 (-0.047; 0.440)  0.273 (0.115; 0.432) | 0.114  0.001 |
| *Outcome: BD*  Exposure 1: HI  Exposure 2: CA | 195 | 0.049 (-0.164; 0.261)  -0.021 (-0.159; 0.117) | 0.654  0.766 |
| *Outcome: MDD*  Exposure 1: HI  Exposure 2: CA | 204 | -0.029 (-0.177; 0.119)  -0.118 (-0.211; -0.025) | 0.701  0.013 |
| *Outcome: OCD*  Exposure 1: HI  Exposure 2: CA | 204 | 0.108 (-0.341; 0.558)  0.296 (0.013; 0.580) | 0.636  0.041 |
| *Outcome: PTSD*  Exposure 1: HI  Exposure 2: CA | 204 | 0.143 (-0.052; 0.338)  -0.077 (-0.206; 0.051) | 0.150  0.236 |
| *Outcome: SZ*  Exposure 1: HI  Exposure 2: CA | 195 | 0.085 (-0.167; 0.336)  -0.163 (-0.328; 0.002) | 0.510  0.052 |

Abbreviations: SNP: single nucleotide polymorphism; IVW: multivariable mendelian randomization via inverse variance weighted method (random effects); B: effect estimates are log-odds; 95% CI: 95% confidence intervals; ADHD: attention deficit hyperactivity disorder; HI: household income; CA: cognitive abilities; AN: anorexia nervosa; ANX: anxiety disorder; ASD: autism spectrum disorder; BD: bipolar disorder; MDD: major depressive disorder; OCD: obsessive-compulsive disorder; PTSD: post-traumatic stress disorder; SZ: schizophrenia

#### Supplementary Table 32: Multivariable Mendelian Randomization results of occupational income and cognitive abilities on mental illness

| **Regression** | **N SNP** | **IVW, B (95% CI)** | **p-value** |
| --- | --- | --- | --- |
| *Outcome: ADHD*  Exposure 1: OI  Exposure 2: CA | 163 | -0.189 (-0.427; 0.048)  -0.511 (-0.654; -0.368) | 0.118  <0.001 |
| *Outcome: AN*  Exposure 1: OI  Exposure 2: CA | 165 | 0.248 (-0.021; 0.517)  0.329 (0.166; 0.491) | 0.071  <0.001 |
| *Outcome: ANX*  Exposure 1: OI  Exposure 2: CA | 164 | -0.032 (-0.425; 0.360)  0.121 (-0.600; -0.126) | 0.872  0.003 |
| *Outcome: ASD*  Exposure 1: OI  Exposure 2: CA | 166 | 0.393 (0.103; 0.682)  0.267 (0.094; 0.441) | 0.008  0.003 |
| *Outcome: BD*  Exposure 1: OI  Exposure 2: CA | 166 | 0.204 (-0.039; 0.447)  0.050 (-0.097; 0.196) | 0.100  0.506 |
| *Outcome: MDD*  Exposure 1: OI  Exposure 2: CA | 166 | -0.079 (-0.258; 0.101)  -0.121 (-0.229; -0.014) | 0.389  0.027 |
| *Outcome: OCD*  Exposure 1: OI  Exposure 2: CA | 166 | -0.133 (-0.657; 0.391)  0.201 (-0.114; 0.516) | 0.618  0.210 |
| *Outcome: PTSD*  Exposure 1: OI  Exposure 2: CA | 166 | 0.143 (-0.092; 0.378)  -0.183 (-0.326; -0.041) | 0.234  0.011 |
| *Outcome: SZ*  Exposure 1: OI  Exposure 2: CA | 166 | -0.003 (-0.298; 0.291)  -0.229 (-0.406; -0.052) | 0.981  0.011 |

Abbreviations: SNP: single nucleotide polymorphism; IVW: multivariable mendelian randomization via inverse variance weighted method (random effects); B: effect estimates are log-odds; 95% CI: 95% confidence intervals; ADHD: attention deficit hyperactivity disorder; OI: occupational income; CA: cognitive abilities; AN: anorexia nervosa; ANX: anxiety disorder; ASD: autism spectrum disorder; BD: bipolar disorder; MDD: major depressive disorder; OCD: obsessive-compulsive disorder; PTSD: post-traumatic stress disorder; SZ: schizophrenia

#### Supplementary Table 33: Multivariable Mendelian Randomization results of social deprivation and cognitive abilities on mental illness

| **Regression** | **N SNP** | **IVW, B (95% CI)** | **p-value** |
| --- | --- | --- | --- |
| *Outcome: ADHD*  Exposure 1: SD  Exposure 2: CA | 152 | -0.135 (-0.317; 0.048)  -0.656 (-0.785; -0.528) | 0.147  <0.001 |
| *Outcome: AN*  Exposure 1: SD  Exposure 2: CA | 161 | -0.240 (-0.448; -0.033)  0.342 (0.185; 0.499) | 0.023  <0.001 |
| *Outcome: ANX*  Exposure 1: SD  Exposure 2: CA | 161 | 0.083 (-0.223; 0.389)  -0.267 (-0.490; -0.44) | 0.593  0.019 |
| *Outcome: ASD*  Exposure 1: SD  Exposure 2: CA | 162 | -0.354 (-0.587; -0.120)  0.235 (0.066; 0.404) | 0.003  0.007 |
| *Outcome: BD*  Exposure 1: SD  Exposure 2: CA | 155 | 0.088 (-0.124; 0.299)  -0.007 (-0.159; 0.145) | 0.416  0.925 |
| *Outcome: MDD*  Exposure 1: SD  Exposure 2: CA | 162 | 0.005 (-0.142; 0.152)  -0.129 (-0.229; -0.028) | 0.946  0.012 |
| *Outcome: OCD*  Exposure 1: SD  Exposure 2: CA | 162 | -0.055 (-0.519; 0.409)  0.335 (0.023; 0.647) | 0.817  0.035 |
| *Outcome: PTSD*  Exposure 1: SD  Exposure 2: CA | 162 | -0.053 (-0.244; 0.139)  -0.140 (-0.279; -0.002) | 0.588  0.047 |
| *Outcome: SZ*  Exposure 1: SD  Exposure 2: CA | 155 | 0.142 (-0.104; 0.387)  -0.218 (-0.395; -0.040) | 0.259  0.016 |

Abbreviations: SNP: single nucleotide polymorphism; IVW: multivariable mendelian randomization via inverse variance weighted method (random effects); B: effect estimates are log-odds; 95% CI: 95% confidence intervals; ADHD: attention deficit hyperactivity disorder; SD: social deprivation measured with Towsend deprivation index; CA: cognitive abilities; AN: anorexia nervosa; ANX: anxiety disorder; ASD: autism spectrum disorder; BD: bipolar disorder; MDD: major depressive disorder; OCD: obsessive-compulsive disorder; PTSD: post-traumatic stress disorder; SZ: schizophrenia.
