## Supplementary File 3 for "The impact of poverty on mental illness: Emerging evidence of a causal relationship"

**Frequently Asked Questions (FAQ)**

Contents:

1. What was your goal with this study?
2. What do you mean when you talk about ‘poverty’?
3. What did you do in this study?
4. What did you find?
5. Does your research imply that poverty and mental illnesses are determined at birth?
6. Why do you use genetics to investigate the relation between poverty and mental illness?
7. Isn’t research like this going to lead to discrimination of those with certain genes?
8. Isn’t research like this going to demotivate people with lower incomes to pursue a better socioeconomic status and mental health?
9. Does your research imply that mentally ill people are not able to earn the money they need to live?
10. Do your results suggest that governments should give more money to the poor to prevent mental illness?
11. What are the practical implications of this research?

#### What was your goal with this study?

Our goal was to investigate the evidence for causality of the relationship between poverty and mental illness. A causal role of poverty in mental illness would be of significant importance for mental illness prevention and our understanding of mental health.

Is the relation between poverty and mental health not already obvious?

Previous research has identified strong correlations between poverty and mental illness, but disentangling cause-and-effect is more difficult. A relation between poverty and mental illness may be due to the effects of mental illness on someone’s financial situation (less income or more spending on health care) or may be caused a third factor (which are called confounding factors). For poverty-mental health relationship for instance education may play a role.

How do you find evidence of causality?

This is why scientists usually rely on randomization to infer causality. For example, to determine whether a specific factor, such as a medical treatment, causes a particular outcome, such depression reduction, the gold standard is to perform Randomized Controlled Trials (RCTs). These studies involve a random assignment of participants into different groups. Randomization enables the casual distribution of potential confounders across the treatment groups, mitigating their effects, and allowing scientists to attribute observed changes to the effect of the intervention. However, conducting such experiments is not always ethical or feasible, such as when it comes to investigating poverty. Clearly, it would not be ethical to deliberately induce poverty in a subset of individuals to observe whether this leads to the development of mental illnesses.

What does genetics have to do with this?

Fortunately, we have all been recruited in an experiment without knowing it, at the point at which we were conceived. Our genes, which have passed on randomly from generation to generation, influence our behaviors but are also markers for our social circumstances. These genetic influences are not related to confounding factors, therefore can be used as instrumental variables. An instrumental variable is a variable that is used as a substitute or proxy for an exposure of interest, which may be difficult to directly observe or manipulate, such as poverty. We can use this knowledge to learn about cause-and-effect, by grouping people according to their genetic code. This method is called Mendelian randomization (MR). In this study we used MR to investigate if poverty is causally associated with mental illness, and vice-versa.

The Figure below represents the distinctions and similarities between MR and RCTs.

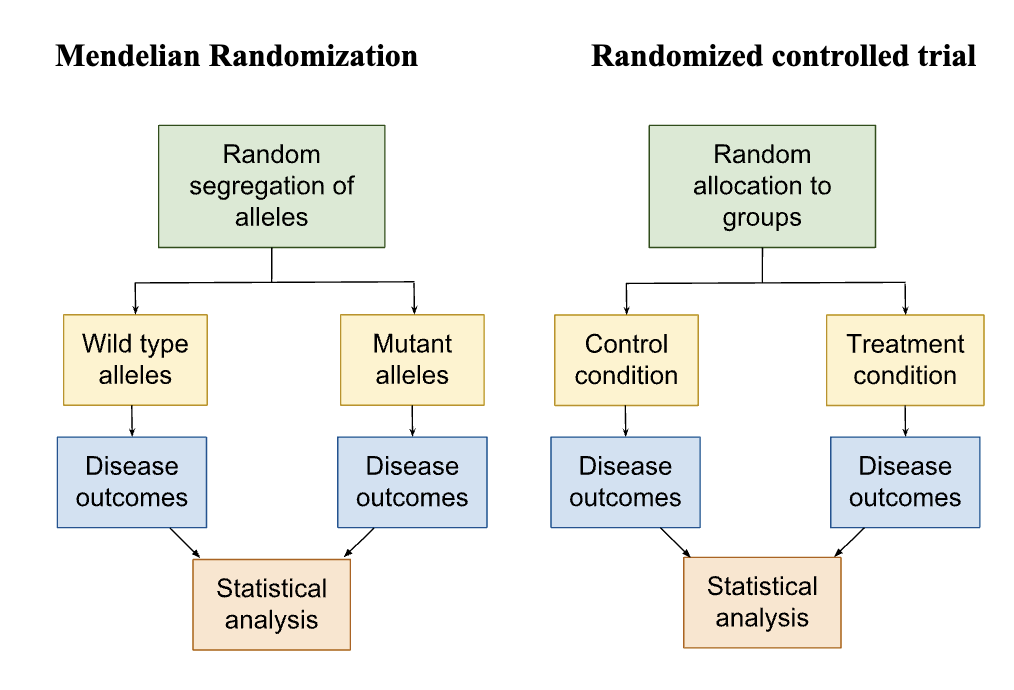

#### What do you mean when you talk about ‘poverty’?

Poverty can be generally described as a situation where people do not have enough money to meet their basic needs and face disadvantages in society. But there are several aspects of poverty that can be distinguished. We used the following definitions of poverty: household income (how much money the whole family earns in a year), occupational income (how much individuals earn from their jobs), and social deprivation (the lack of access to important resources and opportunities).

What do you mean with household income?

Household income refers to the total amount of money earned by everyone in a family across one year span. When household income is low, it means there is not enough money to cover essential things like food, housing, and healthcare. This makes it difficult for people to have a decent quality of life.

What do you mean with occupational income?

Occupational income is the money individuals earn from their jobs. If someone's occupational income is limited, it means they might have low-paying jobs or struggle to find stable employment. This makes it hard to make ends meet and can contribute to poverty.

What is social deprivation?

Social deprivation means not having access to things that are necessary for a good life. This includes things like education, healthcare, housing, transportation, and support from friends and community. When people experience high levels of social deprivation, it can make it even more challenging to escape poverty and improve their circumstances.

#### What did you do in this study?

Our study aimed to investigate the relationship between poverty and various mental illnesses using the method described above, called Mendelian randomization. We utilized genetic variations associated with both poverty and mental illnesses to explore causality. The mental illnesses we focused on included attention deficit and hyperactivity disorder (ADHD), anorexia nervosa, anxiety disorders, autism spectrum disorders, bipolar disorder, major depressive disorder, obsessive-compulsive disorder, post-traumatic stress disorder (PTSD), and schizophrenia. We also considered the impact of cognitive abilities on this relationship, recognizing that the level of education may influence both income and health-related choices. To conduct our statistical analyses, we incorporated cognitive ability as a potential confounding factor.

How did you find the genetic variation that is related to poverty?

Our study employed genome-wide association studies (GWAS) to identify genetic variations linked to poverty. We made a composite measure of poverty that was most strongly related to genetic background. Using genetic data to identify poverty risk may appear unconventional since poverty is not traditionally viewed as a biological condition or trait. However, GWAS has been instrumental in understanding genetic associations with various health conditions and traits. The genetic variations we examined, known as Single-Nucleotide Polymorphisms (SNPs), represent specific locations on DNA where individuals exhibit differences in nucleotide composition. A GWAS study lines up all the SNPs a person has and tests the extent to which each one is linked to the phenotype of interest. In the case of poverty, each SNP might contribute a tiny fraction towards the risk people grow up in poor conditions.

By utilizing these approaches, our research provides valuable insights into the complex interplay between poverty and mental health.

#### What did you find?

Our study identified household income as the most significant measure of poverty, comparing household income with occupational income and social deprivation. Through our investigations using Mendelian randomization, we found compelling evidence that mental illness can contribute to poverty, and in turn, poverty can play a causal role in the development of ADHD, major depressive disorder, and schizophrenia. Interestingly, we observed that poverty was inversely associated with the risk of anorexia nervosa. It is important to note that cognitive ability emerged as a significant factor that influenced many of these findings, affecting the relationship between poverty and mental illness.

Why is this relevant?

These findings underscore the potential benefits of income-based policies as may promote better mental health outcomes in the population. It suggests that targeted interventions aimed at addressing poverty as a cause of mental illness will advance health equity. Our research provides robust evidence supporting the need to address poverty as a significant contributing factor to the development of mental illness.

In summary, our study highlights the critical role poverty plays in mental illness risk and emphasizes the urgency of implementing effective strategies that address both poverty and mental health concerns. This is particularly relevant in an era where inequities and mental illness epidemiology are growing worldwide.

#### Do your methods imply that poverty and mental illnesses are determined at birth?

No, our results do not suggest that an individual's income or likelihood of developing mental illnesses is predetermined at birth. The associations we observed between genetic variations, poverty, and mental health were small and based on extensive sample sizes. These associations indicate that even individuals with similar genes vary with respect to poverty and mental health outcomes. However, there is a slightly elevated likelihood that individuals with specific combinations of genetic variants may have higher incomes or be more susceptible to mental illnesses.

Do genes determine whether you become poor?

Discovering genetic associations with poverty does not mean that other environmental and genetic factors cannot influence them. The concept of "genetic determinism" is entirely false. Another example is educational achievement. Although intelligence and cognitive abilities have a genetic component, environmental factors such as access to quality education, supportive learning environments, and parental involvement play crucial roles in determining educational outcomes. Individuals with the same genetic potential can achieve different educational levels based on their environment and opportunities.

Genetic factors alone do not determine outcomes like income or mental health. Environmental influences and interventions can significantly impact these traits. Therefore, interpreting the finding that "genetic factors are associated with “poverty" as "genes solely determine income, and nothing can be done to change it" reflects a misunderstanding of genetic research (including our study).

#### Why do you use genetics to investigate the relation between poverty and mental illness?

In this study, genetics is used as a measure of poverty risk. In technical terms: an instrumental variable. An instrumental variable is a statistical concept used in research to address potential problems with causality and measurement error. As mentioned, it is a variable that is used as a substitute or proxy for an exposure of interest, which may be difficult to directly observe or manipulate, such as poverty.

Therefore, we utilize genetics to investigate the relationship between poverty and mental illness because it offers a unique perspective and contributes to our understanding of the complex interplay between these factors. While our research does not imply that poverty or mental illness are determined at birth, studying genetic variations associated with these traits allows us to explore potential underlying mechanisms and identify patterns. By examining the genetic links, we can build on the tremendously large datasets that include hundreds of thousands of participants.

#### Isn’t research like this going to lead to discrimination of those with certain genes?

Unfortunately, a lot of scientific research has the potential to be misused. It is crucial to approach genetic research with caution and recognize that it is just one piece of the puzzle when exploring the relationship between poverty and mental illness. While genetic research provides valuable insights, it is important to acknowledge the limitations that include significant influence of many other environmental and genetic factors in the development of mental illness and socioeconomic disparities.

What is the influence of other environmental factors?

Genetic factors interact with many other environmental factors throughout a person's life, shaping their experiences, opportunities, and overall well-being. It is essential to consider that genetics alone does not determine one's fate or potential. Environmental factors such as access to quality education, healthcare services, supportive communities, and early-life experiences play substantial roles in shaping mental health and socioeconomic status.

Is the model used in the study not too simplistic?

It is crucial to adopt a comprehensive and holistic perspective that incorporates both genetic and environmental factors. Genetic research can help us understand the potential genetic vulnerabilities or predispositions that may interact with environmental factors. It is essential to avoid oversimplification or deterministic interpretations of genetic research.

Does this study give rise to discrimination?

To prevent discrimination, it is crucial to emphasize the importance of creating inclusive and supportive environments that foster equal opportunities and access to resources for all individuals, regardless of their genetic or environmental backgrounds. By addressing the broader social determinants of mental health and socioeconomic disparities, we can work towards a more equitable society that promotes mental well-being and socioeconomic mobility for all.

#### Isn’t research like this going to provide an ‘excuse’ to those with lower incomes, which makes them less motivated to pursue a better socioeconomic status and mental health?

Our research aims to provide a comprehensive understanding of the factors influencing mental health and socioeconomic outcomes, including the role of genetics. It is important to note that our findings do not justify or provide an excuse for inaction or a lack of motivation. Instead, they shed light on the complex interplay between genetic factors, poverty, and mental health. By understanding these relationships, we can develop targeted interventions and policies that address the root causes of mental illness and socioeconomic disparities, ultimately promoting better mental health and socioeconomic mobility for all individuals.

#### Does your research imply that mentally ill people are not able to earn the money they need to live?

While the research indicates that there may be causal relations between mental illness and poverty, it does not imply that mentally ill individuals are unable to earn the money they need to live. Mental illness can impact individuals in different ways. Factors such as access to resources, supportive environments, and individual strengths and abilities also play significant roles in determining socioeconomic outcomes.

It is crucial to avoid generalizations and stereotypes about the capabilities and potential of individuals with mental illness. Many individuals with mental health conditions lead fulfilling lives, hold jobs, and contribute to society. However, it is true that mental health challenges can present additional obstacles and may require appropriate support systems to ensure individuals can access equal opportunities. The research highlights the importance of addressing the systemic barriers and stigma that individuals with mental illness may face in employment and socioeconomic domains. By promoting inclusive workplaces, providing reasonable accommodations, and fostering supportive environments, we can create conditions that enable individuals with mental illness to thrive professionally and achieve financial stability.

#### Do your results suggest giving more money to the poor to prevent mental illness?

While our research highlights the evidence of causal relations between poverty and mental illness, it is crucial to recognize that the solution to preventing mental illness goes beyond financial assistance alone. Our findings emphasize the need for comprehensive approaches that address the multifaceted factors influencing mental health outcomes, including socioeconomic conditions, access to healthcare, social support, and environmental factors. Simply providing more money to individuals living in poverty may not be sufficient to prevent mental illness. Instead, our results support the importance of targeted interventions that address the broader social determinants of mental health, such as improving access to quality education, healthcare services, affordable housing, and supportive community programs.

#### What are the practical implications of this research?

At present, there are none. We did not test any practical applications of our genetic knowledge in this study, and nor do we advocate for any. This is basic (as opposed to applied) science, finding links and causes among interesting and important variables, and building a picture of how genes relate to different phenotypes. It is important to note that while our research provides valuable insights, any practical implications may vary depending on the specific context and population under study. Implementation of interventions and policies should be done in a thoughtful and context-specific manner, taking into account the unique needs and resources of different communities and individuals.
